# Prospective Observational Cohort Study Of Tenecteplase Versus Alteplase In Routine Clinical Practice

**DOI:** 10.1101/2022.07.12.22277564

**Authors:** Steven J Warach, Adrienne N Dula, Truman J Milling, Samantha Miller, Leigh Allen, Nathan D Zuck, Collin Miller, Christine A Jesser, Lotika R Misra, Jefferson T Miley, Manzure Mawla, Ming-Chieh Ding, John A Bertelson, Annie Y Tsui, John R Jefferson, Holly M Davison, Darshan N Shah, Matthew M Padrick, Alan S. Nova, Kent T Ellington, Vivek R Krishna, Lisa A Davis, David Paydarfar

## Abstract

**Background and Purpose:** A 10-hospital regional network transitioned to tenecteplase as the standard of care stroke thrombolytic in September 2019 because of its workflow advantages and reported non-inferior clinical outcomes relative to alteplase in meta-analyses of randomized trials. We assessed whether tenecteplase use in routine clinical practice reduces thrombolytic workflow times with non-inferior clinical outcomes.

**Methods:** We designed a prospective registry-based observational, sequential cohort comparison tenecteplase (n=234) to alteplase (n=354) treated stroke patients. We hypothesized: (1) an increase in the proportion of patients meeting target times for target door to needle (DTN) and transfer door-in-door-out (DIDO), and (2) non-inferior favorable (discharge to home with independent ambulation) and unfavorable (symptomatic intracranial hemorrhage, in-hospital mortality or discharge to hospice) in the tenecteplase group. Total hospital cost associated with each treatment was also compared.

**Results:** Target DTN within 45 minutes was superior for tenecteplase, 41% versus 29%; aOR 1.76 (95% CI 1.24, 2.52), *P* = 0.002. Target DIDO within 90 minutes was superior for tenecteplase 37% (15/43) versus 14% (9/65); OR 3.69 (95% CI 1.47, 9.7), *P* =0.006, overall, and 67% (12/18) versus 14% (2/14) for those transferred for thrombectomy after thrombolytic treatment (*P* =0.009). Favorable outcome for tenecteplase fell within the 6.5% non-inferiority margin; aOR 1.28 (95% CI 0.92, 1.77). Unfavorable outcome was less for tenecteplase 7.7% versus 11.9%, aOR 0.79 (95% CI 0.46, 1.32), but did not fall within the pre-specified 1% non-inferior boundary. Net benefit (%favorable – %unfavorable) was greater for the tenecteplase sample: 36% v 27%. *P* =0.022. Median cost per hospital encounter was less for tenecteplase cases ($13,382 vs $15,841; *P* <0.001).

**Conclusions:** Switching to tenecteplase in routine clinical practice in a 10-hospital network was associated with shorter DTN and DIDO times, non-inferior favorable clinical outcomes at discharge, and reduced hospital costs. Evaluation in larger, multicenter cohorts is recommended to determine if these observations generalize.

## Introduction

Alteplase is currently the only thrombolytic approved by US Food and Drug Administration (FDA) for the treatment of ischemic stroke. Tenecteplase, FDA approved only for use in ST-elevation myocardial infarction, is given as a 5-10 second single bolus, as opposed to bolus plus 60-minute infusion for alteplase, owing to greater fibrin specificity and a longer half-life than alteplase. Single bolus thrombolytic has several potential advantages over alteplase in addition to quicker times to prepare and administer the drug. Gaps between the alteplase bolus administration and start of the one-hour infusion is a common occurrence that may result in underdosing the drug^1, 2^. Alteplase infusion requires a second, dedicated intravenous catheter insertion that may delay treatment initiation if access is difficult to obtain. Tenecteplase also has a cost advantage: the average wholesale price in the United States for tenecteplase treatment is approximately $3000 less than alteplase ^3^.

Emerging evidence from randomized ischemic stroke trials and meta-analyses thereof suggests that tenecteplase leads to a greater degree of early reperfusion of large vessel occlusions and is at least noninferior to alteplase in terms of efficacy and safety^4-12^. Recently published experience with tenecteplase in clinical practice from New Zealand concluded that routine use of tenecteplase for stroke thrombolysis was feasible and had comparable safety profile and outcomes to alteplase.^13, 14^ Our 10-hospital network in the United States adopted tenecteplase at 0.25mg/kg as the local standard of care thrombolytic for all ischemic stroke types in September 2019. We designed a prospective cohort study to test for improvements in timing factors and non-inferiority in clinical outcomes relative to our prior experience with alteplase.

## Methods

### Study overview

The switch from alteplase to tenecteplase was approved by local clinical and administrative oversight committees and conditionally approved by the Ascension national pharmacy committee, which required a report on key workflow and clinical metrics to these oversight committees on a regular basis. We therefore designed an observational, open-label, sequential cohort registry study comparing key workflow and clinical metrics between a planned two-year prospective data collection of our tenecteplase treated patients and a retrospective two-year alteplase cohort. Our main hypotheses were that tenecteplase would be associated with reduction in door to needle times and interfacility transfer times while demonstrating non-inferiority in early indices of clinical outcomes compared to our previous alteplase use. During required ongoing quarterly analyses of unadjusted descriptive statistics between the two treatment cohorts, we observed an indication of improvements in door to needle and door to transfer times. We therefore decided to perform and report the full, covariate adjusted statistical comparison of the two cohorts after 15 months of the planned two-year prospective tenecteplase data collection. The corresponding author had full access to all the data in the study and takes responsibility for its integrity and the data analysis. The data that support the findings of this study are available from the corresponding author upon reasonable request.

### Patients

#### Primary Cohort

The primary cohorts for comparisons were comprised of *all* patients treated with an intravenous thrombolytic for ischemic stroke at the 10 Ascension Seton Hospitals in central Texas from September 1, 2017 through December 16, 2020. All Ascension Seton adult hospitals, including two Comprehensive and two Primary Stroke Centers, switched from alteplase to tenecteplase 0.25 mg/kg (maximum dose 25 mg) as the standard of care stroke thrombolytic on September 17, 2019.

#### Secondary Cohort

A secondary cohort was comprised of the subgroup of patients aligned with Class I eligibility recommendations for intravenous alteplase treatment specified in the 2019 American Heart Association/American Stroke Association Acute Ischemic Stroke treatment guidelines.^15^ This included only patients with confirmed ischemic stroke, treated in emergency departments at certified stroke centers within 270 minutes of time last known well and functionally independent prior to their stroke (pre-stroke modified Rankin score [mRS] 0-2 or documentation that the patient had independent ambulation prior to the stroke). See Supplemental Figure S1 for patients selected for Class I subgroup

### Data Source

The Ascension Seton Stroke Program REDCap Registry contains data abstracted from the electronic health record required for reporting data to Get With the Guidelines – Stroke (GWTG), for reporting to stroke center certifying bodies (The Joint Commission, DNV-GL), and for other local quality assessment monitoring purposes. Unless otherwise specified, the data fields reported here were defined in accordance with GWTG definitions and abstraction rules specified in the Ascension Seton Stroke Program Registry Data Dictionary (Supplemental Material), abstracted in proximity to each patient’s admission by staff not directly involved in the patient’s care, and not for the purpose of this cohort analysis. Use of stroke registry data was compliant with local IRB requirements and was exempt from written informed consent.

### Outcomes

Hypothesized differences on four outcomes were tested: two workflow timing and two clinical outcome metrics.

#### Workflow Outcomes

The primary workflow timing outcomes were the percent of patients treated within a target door to needle time (DTN) of 45 minutes and the percent of patients target door-in-door-out transfer time (DIDO) within 90 minutes, target times independently set by GWTG and stroke center certifying bodies. DTN for the main analysis included all treated cases, not excluding cases that GWTG excludes in its consideration of DTN (inpatient strokes, thrombolytic initiation > 4.5 hours from time last known well, or documented delay for medical or eligibility reasons). Secondary analyses of GWTG defined DTN were also included for comparison. The DIDO was defined as the time from emergency department arrival at the treating hospital to discharge from that emergency department to transfer to another Ascension Seton hospital for a higher level of care post thrombolysis, including but not limited to assessment for mechanical thrombectomy. To control for the possibility of overall improvements in workflow times during the years of the study we performed these additional analyses: (1) correlation of DTN with date of treatment (ordinal) within each of the alteplase periods and the tenecteplase periods (2) comparison of DIDO changes between the alteplase and tenecteplase periods for interfacility transfers that were not preceded by thrombolytic administration.

#### Clinical Outcomes

The lead *favorable* clinical outcome was the composite of walking independently at discharge and discharge to home. The lead *unfavorable* clinical outcome included any of the following events: symptomatic intracranial hemorrhage, in-hospital all-cause mortality or discharge to hospice. The mRS at day 90 was not used for the main outcome comparisons, because these values were not documented for more than 40% of alteplase-treated patients, but an exploratory analysis of mRS on available patient data was performed.

#### Other Analyses and Outcomes

1. The relationship between time since last known well and favorable outcome for tenecteplase treatment.
2. A comparison of hospital cost categories and total encounter costs between the two groups.

### Statistical Analysis

Descriptive statistics were compared between the treatment groups using the Wilcoxon rank sum, chi squared, and Fisher exact tests, as appropriate. For testing the relationship of thrombolytic agent to the four main outcomes of interest, general linear models were used. Covariates for adjustment were chosen if the patient characteristics differed between the lytic groups at *P* ≤0.1000 and were also univariable logistic regression predictors of outcome of interest at *P* ≤0.1000. Where no variables met these criteria the unadjusted odds ratio (OR) are reported. Hypothesis testing for the lead analyses used an alpha of 5%. For the hypotheses of improved DTN and DIDO, two-way comparisons were used. For the non-inferiority hypotheses, a one-way comparison was used based on the 90% CI. For the non-inferior clinical outcomes, we chose the minimal clinically important differences (MCID) that were used in previous non-inferiority analyses of stroke thrombolysis: 6.5% for the favorable outcome and 1% for the unfavorable outcomes.^5, 16^ Analyses were performed for both the entire sample and for the Class I subgroup with additional analyses exploring DTN as defined by GWTG.

We evaluated hospital costs associated with each type of thrombolytic treatment by linking the registry data to the hospital cost accounting system to compare overall hospital cost and cost categories between the two groups.

An expanded methods section is provided in the Supplemental Material.

## Results

The 15-month tenecteplase cohort consisted of 234 patients. A total of 356 patients were treated with alteplase; one was excluded for receiving lytic as part of a clinical trial, another treatment was excluded because it was the patient’s second thrombolytic treatment during the study period. The final two-year alteplase cohort was comprised of 354 patients. The Class I subgroup contained 143 and 219 patients, respectively.

### Sample Characteristics (Table 1)

The samples were well matched on age and pre-treatment National Institutes of Health Stroke Scale (NIHSS). A higher proportion of men than women were treated with tenecteplase. The proportion of pre-treatment imaging type differed between the groups, reflecting a change in our practice beginning in 2018 to increasingly use a CT-Stroke protocol (including vascular and perfusion imaging) or MRI as the pretreatment imaging, particularly at the comprehensive stroke centers.

**Table 1.**
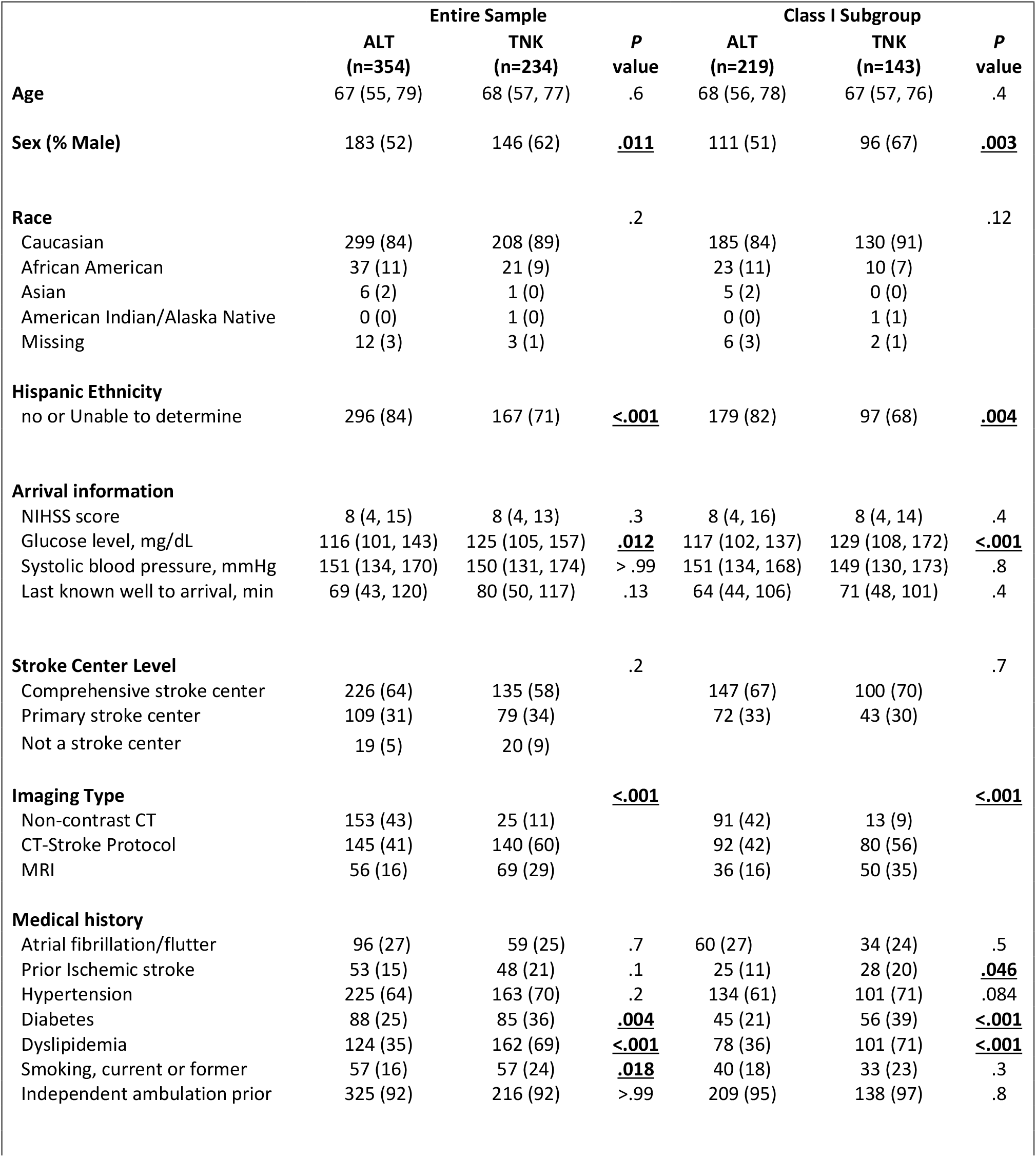
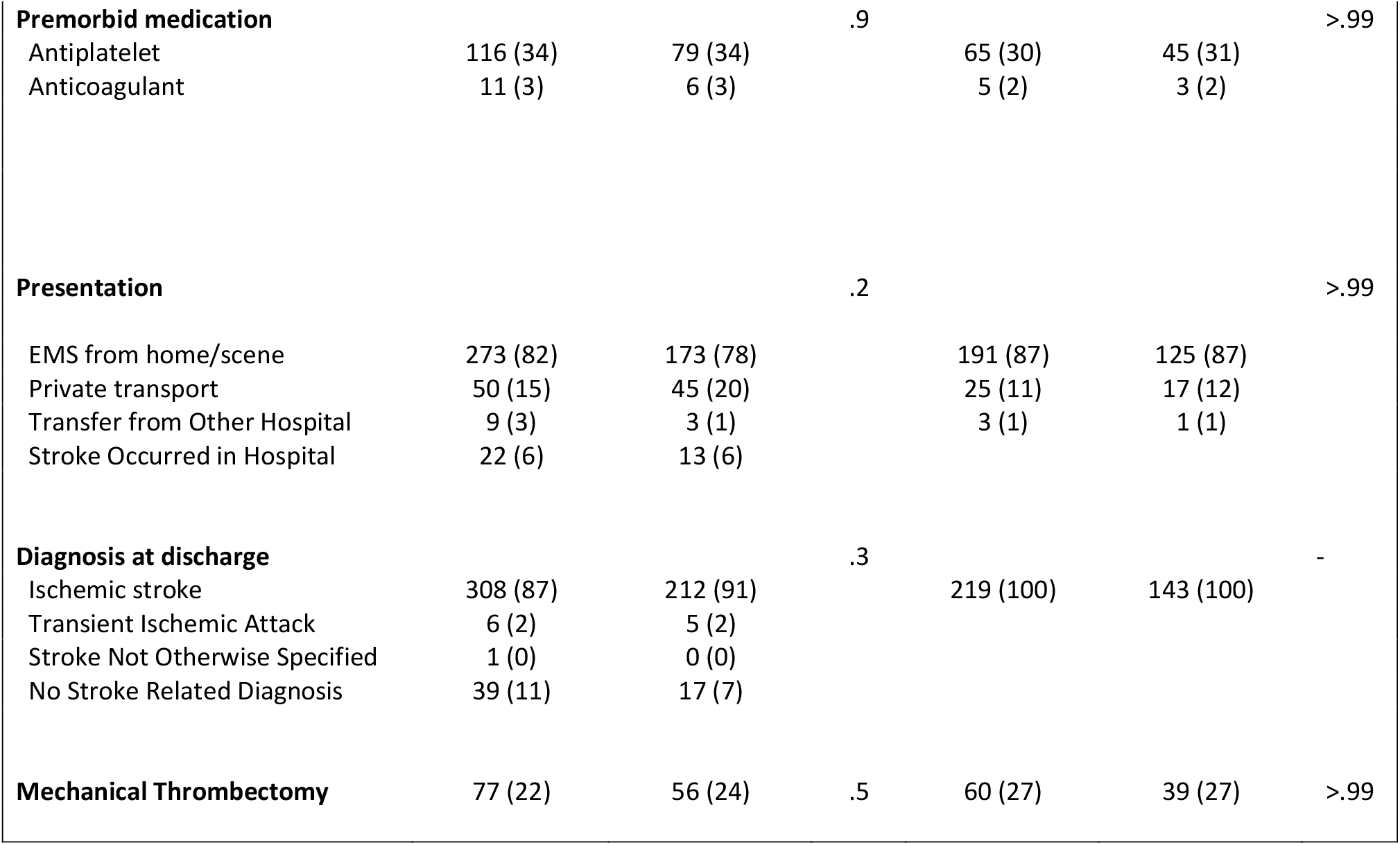
Sample Characteristics [Median (IQR) or No./total No. (%)]

|  | Entire Sample |  |  | Class I Subgroup |  |  |
| --- | --- | --- | --- | --- | --- | --- |
|  | ALT<br>(n=354) | TNK<br>(n=234) | P<br>value | ALT<br>(n=219) | TNK<br>(n=143) | P<br>value |
| **Age** | 67 (55, 79) | 68 (57, 77) | .6 | 68 (56, 78) | 67 (57, 76) | .4 |
| **Sex (% Male)** | 183 (52) | 146 (62) | **.011** | 111 (51) | 96 (67) | **.003** |
| **Race** |  |  | .2 |  |  | .12 |
| Caucasian | 299 (84) | 208 (89) |  | 185 (84) | 130 (91) |  |
| African American | 37 (11) | 21 (9) |  | 23 (11) | 10 (7) |  |
| Asian | 6 (2) | 1 (0) |  | 5 (2) | 0 (0) |  |
| American Indian/Alaska Native | 0 (0) | 1 (0) |  | 0 (0) | 1 (1) |  |
| Missing | 12 (3) | 3 (1) |  | 6 (3) | 2 (1) |  |
| **Hispanic Ethnicity** |  |  |  |  |  |  |
| no or Unable to determine | 296 (84) | 167 (71) | **<.001** | 179 (82) | 97 (68) | **.004** |
| **Arrival information** |  |  |  |  |  |  |
| NIHSS score | 8 (4, 15) | 8 (4, 13) | .3 | 8 (4, 16) | 8 (4, 14) | .4 |
| Glucose level, mg/dL | 116 (101, 143) | 125 (105, 157) | **.012** | 117 (102, 137) | 129 (108, 172) | **<.001** |
| Systolic blood pressure, mmHg | 151 (134, 170) | 150 (131, 174) | > .99 | 151 (134, 168) | 149 (130, 173) | .8 |
| Last known well to arrival, min | 69 (43, 120) | 80 (50, 117) | .13 | 64 (44, 106) | 71 (48, 101) | .4 |
| **Stroke Center Level** |  |  | .2 |  |  | .7 |
| Comprehensive stroke center | 226 (64) | 135 (58) |  | 147 (67) | 100 (70) |  |
| Primary stroke center | 109 (31) | 79 (34) |  | 72 (33) | 43 (30) |  |
| Not a stroke center | 19 (5) | 20 (9) |  |  |  |  |
| **Imaging Type** |  |  | **<.001** |  |  | **<.001** |
| Non-contrast CT | 153 (43) | 25 (11) |  | 91 (42) | 13 (9) |  |
| CT-Stroke Protocol | 145 (41) | 140 (60) |  | 92 (42) | 80 (56) |  |
| MRI | 56 (16) | 69 (29) |  | 36 (16) | 50 (35) |  |
| **Medical history** |  |  |  |  |  |  |
| Atrial fibrillation/flutter | 96 (27) | 59 (25) | .7 | 60 (27) | 34 (24) | .5 |
| Prior Ischemic stroke | 53 (15) | 48 (21) | .1 | 25 (11) | 28 (20) | **.046** |
| Hypertension | 225 (64) | 163 (70) | .2 | 134 (61) | 101 (71) | .084 |
| Diabetes | 88 (25) | 85 (36) | **.004** | 45 (21) | 56 (39) | **<.001** |
| Dyslipidemia | 124 (35) | 162 (69) | **<.001** | 78 (36) | 101 (71) | **<.001** |
| Smoking, current or former | 57 (16) | 57 (24) | **.018** | 40 (18) | 33 (23) | .3 |
| Independent ambulation prior | 325 (92) | 216 (92) | >.99 | 209 (95) | 138 (97) | .8 |
| **Premorbid medication** |  |  | .9 |  |  | >.99 |
| Antiplatelet | 116 (34) | 79 (34) |  | 65 (30) | 45 (31) |  |
| Anticoagulant | 11 (3) | 6 (3) |  | 5 (2) | 3 (2) |  |
| **Presentation** |  |  | .2 |  |  | >.99 |
| EMS from home/scene | 273 (82) | 173 (78) |  | 191 (87) | 125 (87) |  |
| Private transport | 50 (15) | 45 (20) |  | 25 (11) | 17 (12) |  |
| Transfer from Other Hospital | 9 (3) | 3 (1) |  | 3 (1) | 1 (1) |  |
| Stroke Occurred in Hospital | 22 (6) | 13 (6) |  |  |  |  |
| **Diagnosis at discharge** |  |  | .3 |  |  | - |
| Ischemic stroke | 308 (87) | 212 (91) |  | 219 (100) | 143 (100) |  |
| Transient Ischemic Attack | 6 (2) | 5 (2) |  |  |  |  |
| Stroke Not Otherwise Specified | 1 (0) | 0 (0) |  |  |  |  |
| No Stroke Related Diagnosis | 39 (11) | 17 (7) |  |  |  |  |
| **Mechanical Thrombectomy** | 77 (22) | 56 (24) | .5 | 60 (27) | 39 (27) | >.99 |

### Workflow Outcomes (Tables 2 and 4)

In the primary sample, DTN ≤ 45 minutes was observed in 41% of tenecteplase patients versus 29% for alteplase, with an aOR 1.76 (95% CI: 1.24, 2.52); *P* = 0.002. When limited to GWTG defined DTN, 56% versus 41% achieved DTN <u>≤</u> 45 minutes (*P* =0.011). The median DTN was 6 minutes less for tenecteplase, not significantly different for the entire sample, although for the GWTG defined DTN, the median difference of 43 compared to 48 minutes was significant (*P* = 0.012), favoring tenecteplase. Similar differences were observed for the Class I subgroup.

**Table 2.** Treatment Related Timings [Median (IQR) or Number (%)]

|  | Entire Sample |  |  | Class 1 Subgroup |  |  |
| --- | --- | --- | --- | --- | --- | --- |
|  | ALT<br>(n=354) | TNK<br>(n=234) | P<br>value | ALT<br>(n=219) | TNK<br>(n=143) | P<br>value |
| **Door to Needle Time**<br>**within 45 minutes (%)** |  |  |  |  |  |  |
| Entire cohort | 104 (29%) | 95 (41%) | **.006** | 75 (34%) | 69 (48%) | **.011** |
| GWTG-defined DTN | 83 (41%) | 72 (56%) | **.011** | 73 (43%) | 62 (58%) | **.015** |
| **Door to Needle (DTN),**<br>**minutes** |  |  |  |  |  |  |
| Entire cohort | 57 (43, 75) | 51 (38, 80) | .14 | 52 (41, 64) | 47 (37, 66) | .065 |
| GWTG-defined DTN | 48 (39, 59)<br>n=203 | 43 (35, 53)<br>n=129 | **.012** | 47 (39, 58)<br>n=171 | 43 (35, 52)<br>n=106 | **.017** |
| **Onset to Treatment**<br>**Time, minutes** | 139 (103, 197) | 148 (107, 202) | .2 | 130 (96, 172) | 128 (100, 167) | .9 |
| 0-3 hr (0-180 min) | 247 (70%) | 157 (67%) | .7 | 171 (78%) | 117 (82%) | .5 |
| 3-4.5 hr (181-270 min) | 86 (24%) | 59 (25%) |  | 48 (22%) | 26 (18%) |  |
| >4.5 hr (>270 mins) | 21 (6%) | 18 (8%) |  |  |  |  |
| **Length of stay, days** | 4.0 (2.6, 7.1) | 4.4 (2.9, 6.9) | .4 | 4.4 (2.8, 7.3) | 4.8 (3.0, 6.9) | .6 |
| ***Transferred Patients*** |  |  |  |  |  |  |
| **Door-in-door-out (DIDO)** |  |  |  |  |  |  |
| Minutes | 135 (100, 177)<br>n = 65 | 113 (83, 153)<br>n=43 | .054 | 126 (98, 167)<br>n=32 | 88 (83, 182)<br>n=9 | .3 |
| within 90 min | 9 (14%) | 16 (37%) | **.010** | 4 (12%) | 5 (56%) | **.014** |
| **Needle to door out time** |  |  |  |  |  |  |
| Minutes | 58 (32,108)<br>n= 67 | 42 (28, 74)<br>n=43 | .13 | 61 (37, 112)<br>n=33 | 42 (36, 121)<br>n=9 | .5 |
| within 60 minutes | 34/65 (52%) | 29 (67%) | .2 | 16/32 (50%) | 6 (67%) | .5 |
| ***Mechanical Thrombectomy after transfer*** |  |  |  |  |  |  |
| **Door-in-door-out (DIDO)** |  |  |  |  |  |  |
| Minutes | 108 (93, 136)<br>n=14 | 83 (68, 110)<br>n=18 | **.029** | 99 (93, 109)<br>n=9 | 85 (83, 131)<br>n=6 | .6 |
| within 90 min | 2 (14%) | 12 (67%) | **.009** | 2 (22%) | 4 (67%) | .14 |
| **Needle to door out time** |  |  |  |  |  |  |
| Minutes | 44 (36, 77)<br>n=14 | 36 (28, 42)<br>n=18 | .087 | 39 (37, 61)<br>n=9 | 36 (30, 45)<br>n=6 | .3 |
| within 60 minutes | 9 (64%) | 16 (89%) | .2 | 6 (67%) | 5 (83%) | .6 |
| **Needle to arterial**<br>**puncture, minutes** | 124 (85, 173)<br>n=14 | 91 (72, 110)<br>n=18 | **.049** | 149 (86, 201)<br>n=9 | 99 (91, 110)<br>n=6 | .6 |
| **Arrival to arterial**<br>**puncture, minutes** | 179 (155, 225)<br>n=14 | 134 (114,161)<br>n=18 | **.015** | 184 (147, 255)<br>n=9 | 143 (127, 206)<br>n=6 | .4 |

There was no correlation between the order of treatment date and DTN overall (r=0.016) or within either the alteplase (r=0.096) or tenecteplase (r=-0.092) groups.

The percentage DIDO <u>≤</u> 90 minutes for transferred patients was 37% (16/43) for tenecteplase versus 14% (9/65), with OR 3.69 (95% CI: 1.47, 9.7); *P* = 0.006. The difference in median DIDO was 22 minutes. For the subset that received mechanical thrombectomy after transfer from the treating emergency department, 67% (12/18) tenecteplase versus 14% (2/14) achieved the goal of DIDO <u>≤</u> 90 minutes (*P* =0.009). The difference in median DIDO for the thrombectomy transfers was 25 minutes (*P* =0.029). Of note, in this small subgroup, the median delay between arrival at the thrombolysis hospital to arterial puncture at the thrombectomy center was 45 minutes less for the tenecteplase group (*P* =0.015). During the tenecteplase and alteplase time periods, respectively, there was no difference in percentage DIDO <u>≤</u> 90 minutes for patients who did *not* receive lytic prior to transfer: 37/205 (18%) vs 62/278 (22%) overall, and 11/18 (61%) vs 8/20 (40%) for thrombectomy after transfer.

### Clinical Outcomes (Tables 3 and 4)

The lead favorable clinical outcome occurred in 44% in the tenecteplase cohort versus 39% in alteplase cohort. The lower bound of the 90% CI for the aOR 1.28 (0.92, 1.77) was used to obtain to the proportion of favorable events for tenecteplase relative to alteplase, with the difference in proportions of -1.96%, within the non-inferiority margin of -6.5%. For the Class I subgroup, the adjusted difference in proportions exceeded the non-inferiority margin.

**Table 3.** Clinical Outcomes [No./total No. (%)]

|  | Entire Sample |  |  | Class I Subgroup |  |  |
| --- | --- | --- | --- | --- | --- | --- |
|  | ALT<br>(n=354) | TNK<br>(n=234) | *P* value | ALT<br>(n=219) | TNK<br>(n=143) | *P* value |
| **Favorable Outcomes at Hospital discharge** |  |  |  |  |  |  |
| Independent ambulation at discharge | 42.9% | 48.3% | .2332 | 45.2% | 49.6% | .4711 |
| Discharge to home | 51.7% | 52.1% | .9835 | 52.1% | 49.7% | .7340 |
| **Discharge to Home AND Independent Ambulatory** | **39.3%** | **44.0%** | **.2890** | **41.1%** | **44.1%** | **.6538** |
| **Unfavorable Outcomes during Hospitalization** |  |  |  |  |  |  |
| PH, IVH, or SAH on scan by 36 hours | 7.9% | 7.7% | 1.000 | 8.2% | 9.1% | 1.000 |
| Symptomatic ICH | 2.8% | 1.7% | .5538 | 2.3% | 0.7% | .4637 |
| All Cause Death during hospitalization | 6.2% | 3.8% | .2849 | 5.9% | 2.8% | .2602 |
| All Cause Death or discharge to hospice | 10.5% | 6.4% | .1233 | 9.1% | 9.6% | .1523 |
| **All Cause Death, Hospice, or sICH** | **11.9%** | **7.3%** | **.0936** | **9.6%** | **4.2%** | **.0882** |
| **Net Favorable Outcome** | **27.4%** | **36.8%** | **.0181** | **31.5%** | **39.9%** | **.1146** |

**TABLE 4.**
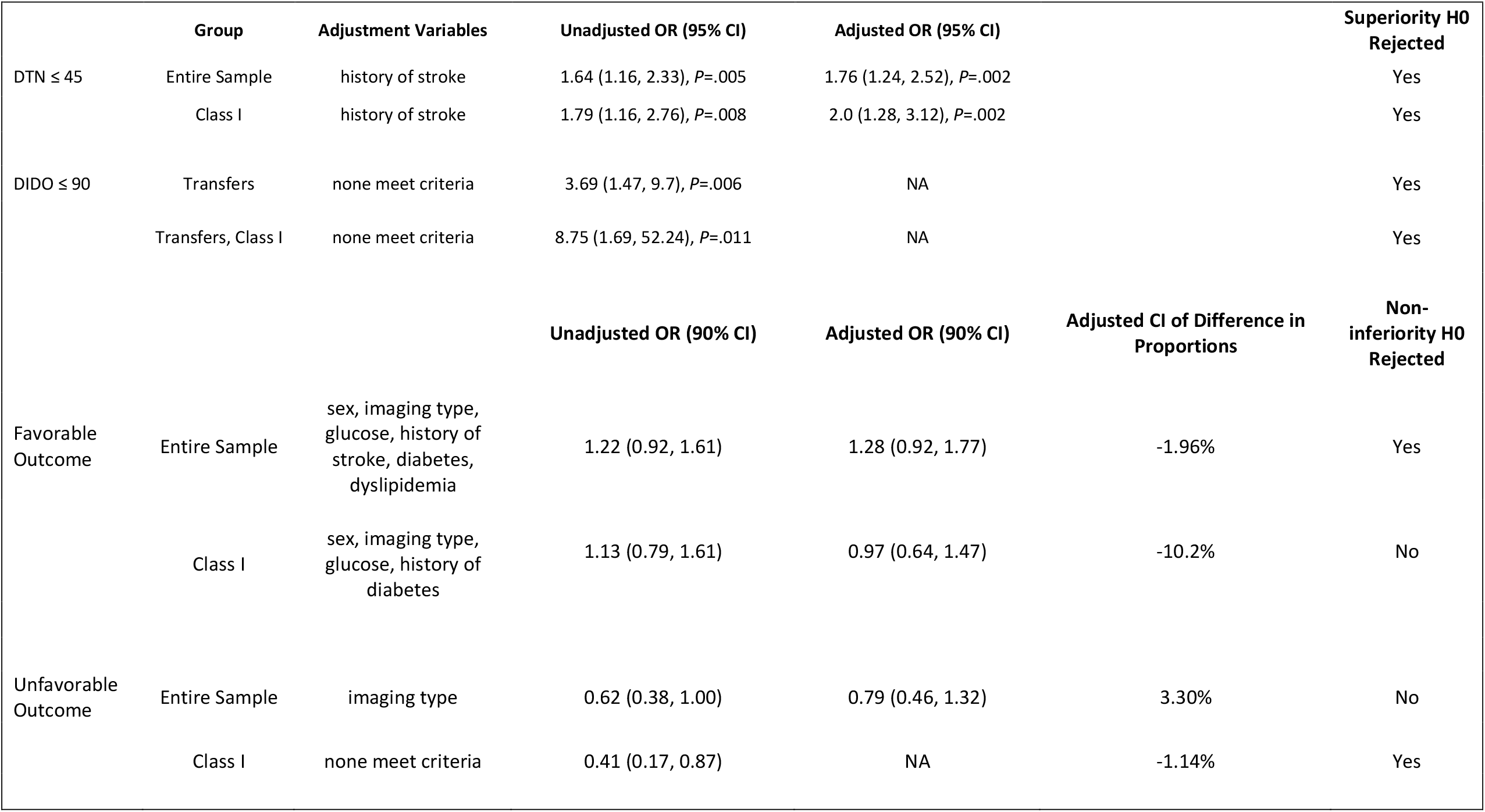
Logistic regression models for superiority and non-inferiority analyses.

|  | Group | Adjustment Variables | Unadjusted OR (95% CI) | Adjusted OR (95% CI) |  | Superiority H0 Rejected |
| --- | --- | --- | --- | --- | --- | --- |
| DTN ≤ 45 | Entire Sample | history of stroke | 1.64 (1.16, 2.33), *P*=.005 | 1.76 (1.24, 2.52), *P*=.002 |  | Yes |
|  | Class I | history of stroke | 1.79 (1.16, 2.76), *P*=.008 | 2.0 (1.28, 3.12), *P*=.002 |  | Yes |
| DIDO ≤ 90 | Transfers | none meet criteria | 3.69 (1.47, 9.7), *P*=.006 | NA |  | Yes |
|  | Transfers, Class I | none meet criteria | 8.75 (1.69, 52.24), *P*=.011 | NA |  | Yes |
|  |  |  | Unadjusted OR (90% CI) | Adjusted OR (90% CI) | Adjusted CI of Difference in Proportions | Non-inferiority H0 Rejected |
| Favorable Outcome | Entire Sample | sex, imaging type, glucose, history of stroke, diabetes, dyslipidemia | 1.22 (0.92, 1.61) | 1.28 (0.92, 1.77) | -1.96% | Yes |
|  | Class I | sex, imaging type, glucose, history of diabetes | 1.13 (0.79, 1.61) | 0.97 (0.64, 1.47) | -10.2% | No |
| Unfavorable Outcome | Entire Sample | imaging type | 0.62 (0.38, 1.00) | 0.79 (0.46, 1.32) | 3.30% | No |
|  | Class I | none meet criteria | 0.41 (0.17, 0.87) | NA | -1.14% | Yes |

The composite lead unfavorable endpoint at discharge of sICH, death, or hospice was 7.7% for tenecteplase and 11.9% for alteplase. Non-inferiority of unfavorable outcomes was not demonstrated for the entire sample, as the lower bound of the 90% CI for the aOR 0.79 (0.46, 1.32) corresponded to a difference in proportions of 3.3%, which exceeded the 1% non-inferiority margin. For the Class I subgroup, the lower bound of the 90% CI for the aOR 0.41 (0.17, 0.87) corresponded to a difference in the proportion of unfavorable outcomes, -1.14% (fewer events for tenecteplase), which was within the non-inferiority margin of 1%.

One life threatening systemic hemorrhagic was noted within each group. Orolingual angioedema was documented in one alteplase and three tenecteplase patients. Positive brain scan for hemorrhage and clinical worsening was recorded for 2.1% of tenecteplase and 4.5% of alteplase cases; these were adjudicated as symptomatic intracranial hemorrhage (sICH) in 1.7% and 2.8%, respectively.

A net favorable outcome (% favorable outcome – % unfavorable outcome) was greater for tenecteplase: 36% v 27%. p=0.022.

Results on available mRS at day 90 and last observation carried forward to day 90 are presented in Supplemental Tables S1 and S2, Supplemental Figure S2. The 90 day mRS analyzed by adjusted ordinal regression for the entire sample favored tenecteplase: the odds of a 1-point increase in score on the mRS scale are decreased for patients in the entire cohort receiving TNK, proportional OR=0.66 (0.45, 0.97), *P* =0.034.

Results for mild (median NIHSS 4), thrombectomy and non-thrombectomy subgroups are presented in Supplemental Table S3. These subgroups show similar trends to differences as in the entire sample.

### Other Analyses and Outcomes

1. There was an inverse relationship between onset to treatment time (time last known well to lytic bolus) and the favorable clinical outcome at discharge, with aOR 0.993 (95% CI: 0.987, 0.998) reaching significance at *P* =0.01, indicating a decreased likelihood of favorable outcome as time to treatment increased adjusted for age, NIHSS, blood glucose, history of hypertension, dyslipidemia, and diabetes (Figure 1).
2. Table 5 shows the hospital costs overall and by hospital category. Total costs were greater for the alteplase group: median (IQR) $15,841 (12,300, 22,323) for alteplase vs $13,382 (10,686, 20,636) for tenecteplase (*P* <0.001). The greatest difference was in total Pharmacy costs $9,288 (8,657, 11,751) vs $6,997 (6,460, 7,972), *P* <0.001.

**Figure 1.**
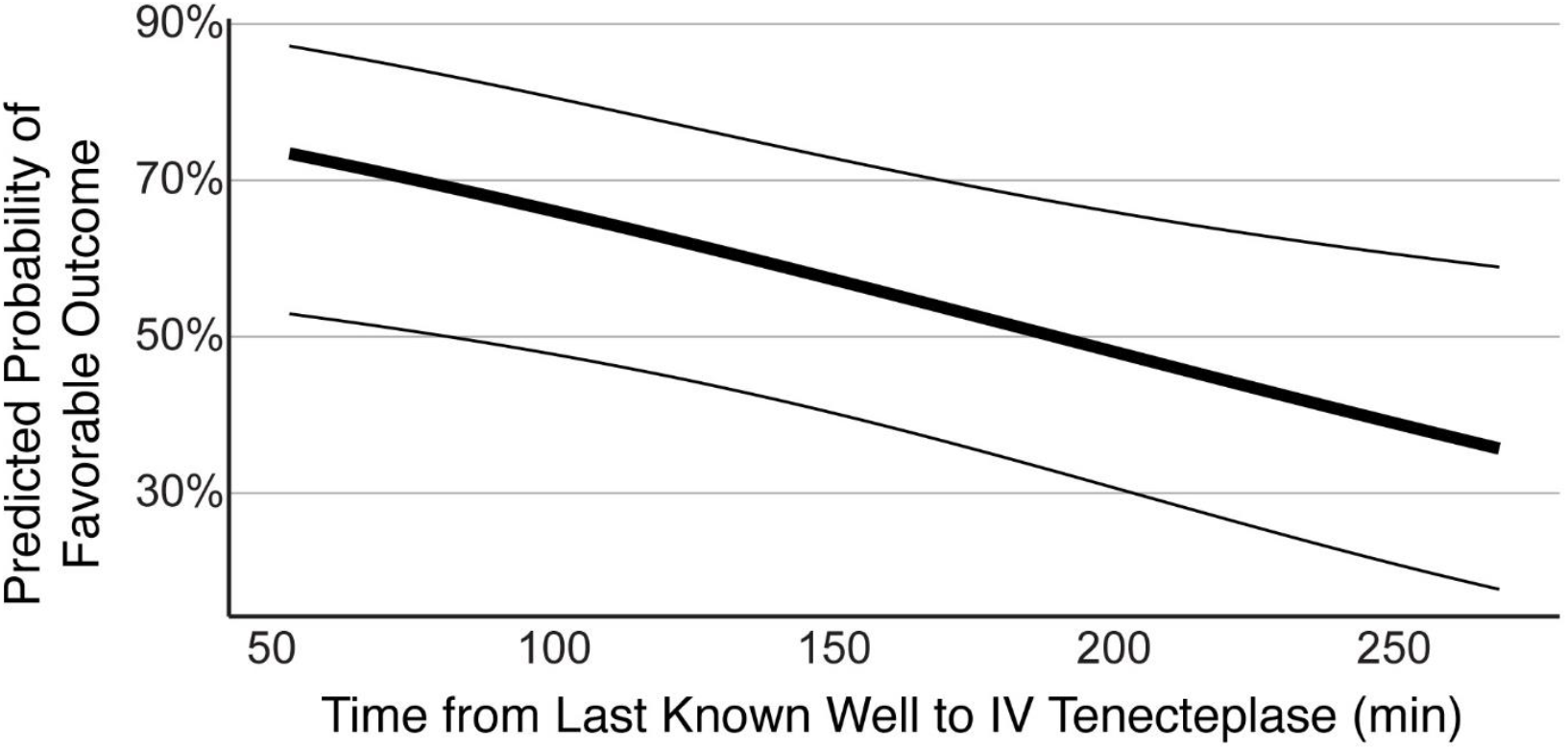
Relationship of time to treatment to probability of favorable outcome at discharge for tenecteplase treated patients. An inverse relationship was observed in the time from stroke onset to the favorable discharge outcome of discharge to home walking independently, with aOR 0.993 (0.9897, 0.998), p=0.01. The figure plots this relationship with the 95% CI, holding the covariates constant at their average values: age (66.09), NIHSS (9.27), blood glucose on arrival (152 mg/dl), history of hypertension (no), dyslipidemia (no), and diabetes (no).

**Table 5.** Hospital Cost Breakdown.

|  | **ALT** | **TNK** | **p-value** |
| --- | --- | --- | --- |
| Diagnostic | 16 (16, 127) | 32 (16, 127) | 0.4 |
| Imaging | 723 (507, 1,031) | 834 (533, 1,143) | 0.044 |
| Lab | 224 (142, 487) | 259 (164, 512) | 0.035 |
| Pharmacy | 9,288 (8,657, 11,751) | 6,997 (6,460, 7,972) | <0.001 |
| Service Line | 117 (105, 236) | 120 (105, 242) | 0.7 |
| Supplies and Devices | 6 (0, 702) | 12 (0, 2,758) | 0.2 |
| Ancillary Services | 446 (235, 773) | 514 (295, 882) | 0.036 |
| Room and Board | 3,110 (2,107, 5,009) | 3,110 (2,107, 4,665) | 0.5 |
| Operative Services | 0 (0, 983) | 0 (0, 1,247) | 0.3 |
| **OVERALL ENCOUNTER** | **15,841 (12,300, 22,323)** | **13,382 (10,686, 20,636)** | **<0.001** |

Median (IQR)
Wilcoxon rank-sum test

## Discussion

Our study is the first to test prospectively defined hypotheses about tenecteplase versus alteplase differences in real world clinical practice in the United States. We examined tenecteplase use as the standard stroke thrombolytic at a single dose in a broad sample from routine clinical practice at hospitals across all levels of stroke certification. We found tenecteplase use was associated with reduced DTN and DIDO, non-inferiority for early indices of efficacy and reduced hospital costs compared to alteplase use in the preceding two years. Although the lower rate of unfavorable outcomes did not meet our stringent non-inferiority margin, the unfavorable outcome rates were lower for tenecteplase and the net benefit (% favorable - % unfavorable) in the samples were significantly greater for tenecteplase: 36% to 27%. We also demonstrated the inverse temporal relationship between onset to treatment time and clinical benefit with tenecteplase, similar to this well described relationship for alteplase. ^17^ Overall, these results from routine clinical practice are consistent with the results of randomized trials and meta-analysis and are supportive of tenecteplase as an alternative to alteplase for stroke thrombolysis.^4-12^ Our results for tenecteplase are also similar in magnitude and difference to the experience reported from New Zealand.^13, 14^

Currently American Heart Association clinical practice guidelines give a Class IIb recommendation of tenecteplase as an alternative to alteplase^15^ but recommend different doses for large vessel occlusions for whom thrombectomy is planned (0.25 mg/kg; EXTEND IA TNK Part1^11^) and for milder strokes (0.4mg/kg NORTEST^12^). We treated all patients with 0.25 mg/kg and our results were similar for the entire sample, for mild stroke, for thrombectomy and for non-thrombectomy patients (Supplemental Table 1). There was no evidence in our data to suggest that stroke severity or subsequent thrombectomy interacted with the difference between tenecteplase and alteplase at this dose, an observation in line with meta-analyses and randomized comparisons in large vessel occlusions that show no advantage of the higher dose for any type of stroke^4, 18^.

### Workflow Outcomes

Our finding of 11% greater proportion of tenecteplase treated patients meeting the goal of treatment within 45 minutes of hospital arrival (14% greater for the Class I subgroup, and 15% for the GWTG defined DTN time) may be clinically meaningful. The clinical benefit of earlier time to initiate intravenous thrombolytic treatment of ischemic stroke with alteplase is well established from meta-analyses of placebo-controlled trials for 3-month mRS.^17^ This benefit was further confirmed in GWTG-Stroke registry data for discharge to home and walking independently^19-21^. Additionally, shorter time from stroke onset (time last known well) to tenecteplase bolus in our data was associated with a higher probability of the favorable outcome of combined discharge to home and walking independently (Figure 1). Time savings associated with tenecteplase treatment, most notable prior to transfer for mechanical thrombectomy, might therefore lead to greater likelihood of favorable clinical outcomes, especially considering data that tenecteplase is associated with higher rates of early recanalization than alteplase.^4, 5, 11^

The practical advantages of tenecteplase -- single bolus dosing eliminating the potential for a delay between bolus and start of infusion, quicker preparation and administration times, without the need for a second dedicated intravenous catheter or a one-hour infusion – may not only contribute to shorter door-to-needle times but may also facilitate more rapid transfer of patients following thrombolytic initiation, especially where ambulance crews, such as in our region, may not transport patients during the alteplase infusion. These interfacility transfer delays would also impact the times from arrival at the treating hospital to the start of thrombectomy at the receiving hospital, further risking a negative impact on stroke outcomes. In our sample, a greater proportion of transferred tenecteplase patients met the DIDO time goal of 90 minutes compared to the alteplase cohort. For the subset of patients transferred for thrombectomy median DIDO time interval was 25 minutes less for the tenecteplase group, and the median time from emergency department arrival at the transferring hospital to arterial puncture at the thrombectomy center was 45 minutes shorter. These time savings are likely to equate to significant clinical impact on functional independence and quality adjusted life years.^22, 23^ Meta-analysis of thrombectomy trials demonstrated that the odds of reduced disability at 90 days in thrombectomy patients decline significantly with longer times from emergency department arrival to arterial puncture or to reperfusion.^22^

### Other Analyses and Outcomes

We constructed the plot, often seen for alteplase treatment, of time from stroke onset to treatment versus favorable clinical outcome (Figure 1), finding a similar inverse relationship of time to treatment and favorable outcome, thus supporting similar biological properties to alteplase.

The analysis of overall hospital cost data across the study period indicated about $2,500 per dose lower cost with tenecteplase treatment, primarily attributed to differences in pharmacy costs. Hospitals that purchase thrombolytics under the Unites States government’s 340B Drug Pricing Program would be less likely to realize these cost savings with tenecteplase use. Reduced medication costs play an important role in cost effectiveness analyses and may amplify savings and quality adjusted life years across healthcare systems.^24^ The magnitude of savings that we observed could equate to an excess of $150 million if applied to the annual US stroke thrombolysis cases.

There are several limitations to the interpretation of these results. 1. Thrombolytic treatment assignment was neither randomized nor blinded and therefore subject to biases in management decisions and outcome assessments. We attempted to attenuate this limitation by choosing variables less prone to bias and that were already being abstracted from the medical record into the registry for other purposes prior to the design of this study and abstracted by personnel not directly involved in the patients’ care. 2. Our analysis was based on observations from a single regional network of hospitals and its stroke clinical program. It is unknown if similar results and conclusions would be obtained from other stroke centers or multicenter registries of tenecteplase use in practice. 3. The sequential nature of the alteplase and tenecteplase cohorts could potentially introduce bias due to changes in practice patterns or workflow efficiencies over time. Other than incorporation of imaging-based thrombolysis in the later (>4.5 hours) time windows, and increased use of MRI and CT perfusion as part of the pre-treatment assessment, there were no systemic changes in our thrombolysis or thrombectomy criteria or practice patterns over the years of observation. We found no improvement in DIDO between the two periods for patients not receiving thrombolytic prior to transfer and no improvement in DTN attributable to a general improvement in workflow efficiency over time. 4. Registry data may be highly variable, prone to variability and biases in data abstraction. Our rules for data abstraction were set independently and prior to the planning of this study, based on stroke center certification and quality assessment requirements, which included increased requirement of some data fields in later years of this analysis not required in earlier years, e.g., Day 90 mRS.

Data abstractors were prohibited from interpreting rather than only transcribing from the electronic health record, using standardized case definitions and coding instructions, predefined logic and range checks on data fields at data entry, audit trails, and regular data quality checks. Some of the differences in baseline medical history and demographic variables may have been due to changes in data abstraction requirements and practices. 5. Uncollected variables may have influenced our results, however, we controlled for available confounders in our multivariable analyses. 6. Finally, the lead clinical outcomes reported in this study are short-term (at hospital discharge), as we did not reliably collect day 90 mRS until the tenecteplase era. However, supporting their use, our lead favorable and unfavorable clinical outcomes components are routinely reported in publications using GWTG data,^19, 21, 25, 26^ and these early indices of clinical outcome have been shown to predict 90-day mRS.^27^ Although an unknown degree of bias may be reflected in subset of our patients for whom day 90 mRS was collected, the more favorable day 90 mRS in the tenecteplase cohort is consistent with our clinical outcomes at hospital discharge.

## Conclusions

This is a first prospective test of tenecteplase as a standard stroke thrombolytic in a broad clinical population at hospitals with varying levels of stroke certification. Attainment of door-to-needle and transfer time goals were significantly improved, and favorable clinical outcomes of discharge to home and walking independently were non-inferior to the previous time period of alteplase use. Unfavorable clinical outcomes, including sICH, in-hospital mortality and discharge to hospice, were fewer but did not satisfy the stringent non-inferiority margin. Although confirmation in larger multicenter registries and ongoing randomized trials is needed, these observations support the practice of tenecteplase use as a reasonable alternative to alteplase for ischemic stroke thrombolysis

## Supporting information

Ascension Seton Stroke Registry Data Dictionary

## Data Availability

All data produced in the present study are available upon reasonable request to the authors

## List of Supplemental material (in order of citation in the manuscript)

- Supplemental Figure S1. Patients selected for Class I subgroup
- Ascension Seton Stroke Program Registry Data Dictionary (separate PDF)
- Expanded Methods Section
- Supplemental Table S1. Modified Rankin Score (mRS) at 90 days comparing alteplase and tenecteplase. Descriptive statistics
- Supplemental Table S2. Modified Rankin Score (mRS) at 90 days comparing alteplase and tenecteplase. Regression analyses.
- Supplemental Figure S2. Modified Rankin Score (mRS) at 90 days comparing alteplase and tenecteplase. Horizontal stacked bar chart.
- Supplemental Table 3. Mild, Thrombectomy, and Non-Thrombectomy Subgroup Descriptive statistics

