## Supplemental Methods Figures and Tables for "Prospective Observational Cohort Study Of Tenecteplase Versus Alteplase In Routine Clinical Practice"

**List of Supplemental material (in order of citation in the manuscript)**

- Supplemental Figure S1. Patients selected for Class I subgroup
- Ascension Seton Stroke Program Registry Data Dictionary (separate PDF)
- Expanded Methods Section
- Supplemental Table S1. Modified Rankin Score (mRS) at 90 days comparing alteplase and tenecteplase. Descriptive statistics
- Supplemental Table S2. Modified Rankin Score (mRS) at 90 days comparing alteplase and tenecteplase. Regression analyses.
- Supplemental Figure S2. Modified Rankin Score (mRS) at 90 days comparing alteplase and tenecteplase. Horizontal stacked bar chart.
- Supplemental Table 3. Subgroup Descriptive statistics

### Figure S1. Patients selected for Class I subgroup

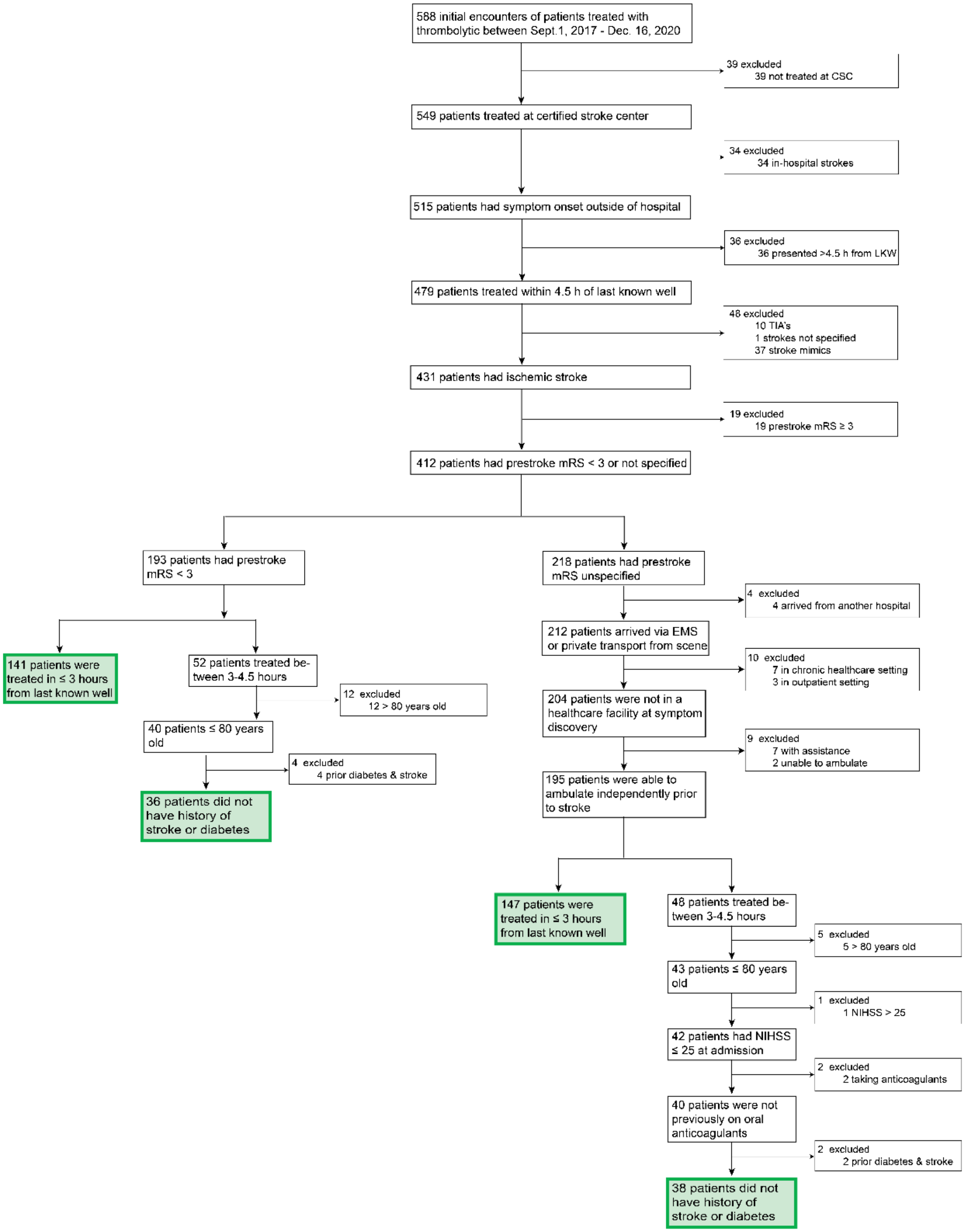

**Expanded Methods Section**

Study overview:

The switch from alteplase to tenecteplase was approved by local clinical and administrative oversight committees and legal office, as well as by an Ascension Healthcare national pharmacy committee, which conditionally approved and authorized the change in our region as a pilot project for Ascension Healthcare. We were required to report on key workflow and clinical metrics to these oversight committees on a regular basis. Descriptive statistics were updated on a roughly quarterly basis and compared these with summary data with alteplase from the approximately 2-year period prior to the switch. We therefore designed an observational, open-label, sequential cohort registry study comparing key workflow and clinical metrics between a planned two-year prospective data collection of our tenecteplase treated patients and a retrospective two-year alteplase cohort. Our main hypotheses were that tenecteplase would be associated with reduction in door to needle times and a reduction interfacility transfer times while demonstrating non-inferiority in early indices of clinical outcomes compared to previous alteplase use. During required ongoing quarterly comparative analyses of unadjusted descriptive statistics between the two treatment cohorts, we observed an early indication of improvements in door to needle and door to transfer times. We therefore decided to perform and report the full, covariate adjusted statistical comparison of the two cohorts after 15 months of the planned two-year prospective tenecteplase data collection.

Patients:

*Primary Cohort:* The primary cohorts for comparisons were comprised of *all* patients treated with an intravenous thrombolytic for ischemic stroke at one of the 10 Ascension Seton Hospitals from September 1, 2017 through December 16, 2020. All stroke thrombolytic treatment decisions and cases were staffed by members of the Ascension Seton neurology practice group at all of the hospitals, which used standardized stroke order sets and treatment protocols. After a brief run-in period at one of the comprehensive stroke centers, all Ascension Seton adult hospitals, which include two Comprehensive and two Primary Stroke Centers, switched from alteplase to tenecteplase 0.25 mg/kg (maximum dose 25 mg) as the standard of care stroke thrombolytic on September 17, 2019. If patients received thrombolytic for stroke more than once during this period, only the first admission was included. Patients enrolled in acute stroke treatment clinical trials were excluded. To reduce times to initiate treatment with alteplase, emergency departments were encouraged to mix and prepare alteplase for stroke thrombolytic candidates prior to the final determination of eligibility, since wasted drug was replaced by the manufacturer. Early mixing was not permitted for tenecteplase, since there was no replacement program for that drug. There were no changes in determining thrombolysis eligibility or post treatment monitoring that occurred with the change in thrombolytics.

*Secondary Cohort:* A secondary cohort was comprised of the subgroup of patients aligned with Class I eligibility recommendations for intravenous alteplase treatment specified the 2019 American Heart Association/American Stroke Association Acute Ischemic Stroke treatment guidelines.[14] This included only patients with confirmed ischemic stroke, treated in emergency departments or documentation that the patient had independent ambulation prior to the stroke at certified stroke centers within 270 minutes of time last known well and functionally independent prior to their stroke (pre-stroke modified Rankin score [mRS] 0-2). For patients with pre-stroke mRS unavailable, the stroke had to have occurred outside of a health care setting with documentation that the patient had independent ambulation prior to the stroke. Patients who upon presentation were intended to be treated within 270 minutes, but whose actual time to treat exceeded that were excluded from this Class I subgroup. (See Supplemental Figure S1).

Data Source:

The Ascension Seton Stroke Program REDCap Registry contains data abstracted from the electronic health record required for reporting data to Get With the Guidelines – Stroke (GWTG), for reporting to stroke center certifying bodies (The Joint Commission, DNV-GL), and for other local quality assessment monitoring purposes. Because the rules for data abstraction limited abstraction from patients at non-stroke centers and from patients whose discharge diagnosis was other than stroke, required data fields for this analysis on those patients, if missing, were abstracted from the electronic health record during the data cleaning process for this study. Patients transferred between Ascension Seton hospitals had separate records in the registry for each hospital component, and these records were merged in the creation of the data file for statistical analysis. Data cleaning also resolved out-of-range or extreme values (e.g., negative time intervals) if the correct values were present in the electronic health record. Data values were not permitted to be inferred, estimated, or interpreted from the electronic health record. Unless otherwise specified, the data fields abstracted into the registry and reported here were defined in accordance with GWTG requirements and definitions and abstraction rules were specified in the data dictionary (see Ascension Seton Stroke Program Registry Data Dictionary in Supplemental material). Since March 2019, the majority of data abstraction was outsourced to Navion Healthcare Solutions, prior to which data abstraction was performed by local stroke site coordinators. Data dictionary including abstraction rules are further detailed in the Supplemental Material. This study was conducted in compliance with institutional ethical requirements at University of Texas Austin and Ascension Seton. Informed consent was not required, and a de-identified data set derived from the local registry was prepared for data analysis.

Outcomes:

Hypothesized differences on four outcomes were tested: two workflow timing and two clinical outcome metrics.

*Workflow Outcomes:* The primary workflow timing outcomes were the percent of patients treated within a target door to needle time (DTN) of 45 minutes and the percent of patients target door-in-door-out transfer time (DIDO) within 90 minutes, target times independently set by GWTG and stroke center certifying bodies. DTN for the main analysis included all treated cases, not excluding cases that GWTG excludes in its consideration of DTN (inpatient strokes, thrombolytic initiation > 4.5 hours from time last known well, or documented delay for medical or eligibility reasons). Secondary analyses of GWTG defined DTN were also included for comparison. The DIDO was defined as the time from emergency department arrival at the treating hospital to discharge from that emergency department to transfer to another Ascension Seton hospital for a higher level of care post thrombolysis, including but not limited to assessment for mechanical thrombectomy. Patients admitted to the hospital prior to transfer were not included in the DIDO analysis.

Secondary workflow timing metrics included the following. Onset to treatment time was defined as the time last known well to the time of thrombolytic bolus. For transferred patients, Needle to door out time was defined as the time of thrombolytic bolus to initiation of transfer; Needle to arterial puncture time was defined as the time of thrombolytic bolus at sending hospital to initiation of angiography at receiving hospital; Arrival to arterial puncture time was defined as the time of arrival at sending hospital to initiation of angiography at receiving hospital. To control for the possibility of overall improvements in workflow times during the years of the study we performed these additional analyses: (1) correlation of DTN with date of treatment (ordinal) within each of the alteplase periods and the tenecteplase periods (2) comparison of DIDO changes between the alteplase and tenecteplase periods for interfacility transfers that were not preceded by thrombolytic administration.

*Clinical Outcomes:* The lead *favorable* clinical outcome was the composite of walking independently at discharge and discharge to home. The lead *unfavorable* clinical outcome included any of the following events: symptomatic intracranial hemorrhage, in-hospital all-cause mortality or discharge to hospice. Symptomatic intracranial hemorrhage was defined as parenchymal hematoma, subarachnoid hemorrhage, and/or intraventricular hemorrhage as documented on the radiology report of brain imaging accompanied by a documented worsening of National Institutes of Health Stroke Scale (NIHSS) score that was higher by at least 4 points within 36 hours of thrombolytic treatment. The hemorrhage must have been identified as the predominant cause of the neurologic deterioration, as adjudicated by a vascular neurologist. The mRS at discharge or at day 90 were not used as outcome comparisons, since these values were not documented for more than 40% of alteplase-treated patients, but an exploratoryanalysis of mRS on available patient data.

*Other Analyses and Outcomes:*

1. The relationship between time since last known well and favorable outcome for tenecteplase treatment was plotted and analyzed.
2. A comparison of hospital cost categories and total encounter costs between the two groups.

<u>Statistical Analysis:</u>

Descriptive statistics were compared between the treatment groups using the Wilcoxon Rank Sum, Chi Square, and Fisher Exact tests, as appropriate. For testing the relationship of thrombolytic agent to the four main outcomes of interest, general linear models were used. Covariates for adjustment were chosen if the patient characteristics differed between the lytic groups at p<=0.1 and were also univariate logistic regression predictors of the outcome of interest at p<=0.1. Where no variables met these criteria the unadjusted odds ratio (OR) are reported. Hypothesis testing for the lead analyses used an alpha of 5%. For the hypotheses of improved DTN and DIDO a two-way comparison was used. For the non-inferiority hypotheses, a one-way comparison was used based on the 90% CI. For the non-inferior clinical outcomes we chose the minimal clinically important differences (MCID) that were used in previous non-inferiority analyses of stroke thrombolysis: 6.5% for the favorable outcome and 1% for the unfavorable outcomes.[5,15]

Analyses were performed for both the entire sample and for the Class I subgroup. Additional analyses explored DTN as defined by GWTG. The subgroup of patients transferred after receiving thrombolytic were further subdivided into those for whom mechanical thrombectomy was planned. Patients were categorized as mechanical thrombectomy if the procedure was planned and angiography was performed, regardless of whether thrombus was identified at the initial angiogram or whether the thrombus could not be accessed because of anatomical reasons. A full analysis of the mechanical thrombectomy treated subgroup is underway and beyond the scope of this report. An additional analysis explored the relationship between favorable clinical outcome and time to treatment for patients treated within 4.5 hours.

We evaluated hospital costs associated with each type of thrombolytic treatment by linking the registry data to the hospital cost accounting system to compare overall hospital cost and cost categories between the two groups.

Supplemental Table S1. Modified Rankin Score (mRS) at 90 days comparing alteplase and tenecteplase. Descriptive statistics.

**Entire Cohort**

| | mRS at 90 Days Last Observation Carried Forward (N = 459) | | | mRS at 90 Days Actual (N = 344) | | |
|---|---|---|---|---|---|---|
| Characteristic | ALT, N = 231[1] | TNK, N = 228[1] | *P*-value[2] | ALT, N = 184[1] | TNK, N = 160[1] | *P*-value[2] |
| Age, y | 67 (54, 79) | 68 (57, 77) | 0.7138 | 70 (56, 79) | 69 (58, 79) | 0.9631 |
| Male Sex | 117 (51%) | 142 (62%) | 0.012 | 92 (50%) | 96 (60%) | 0.0631 |
| NIHSS prior to treatment | 8 (4, 15) | 8 (4, 13) | 0.1695 | 8 (4, 15) | 8 (4, 14) | 0.7016 |
| Glucose level, mg/dL | 118 (106, 144) | 124 (105, 157) | 0.233 | 117 (106, 144) | 126 (107, 154) | 0.1333 |
| Imaging modality | | | <0.0001 | | | <0.0001 |
| CT imaging | 88 (38%) | 25 (11%) | | 79 (43%) | 18 (11%) | |
| CT with perfusion imaging | 98 (42%) | 136 (60%) | | 67 (36%) | 93 (58%) | |
| Diffusion and perfusion MRI | 45 (19%) | 67 (29%) | | 38 (21%) | 49 (31%) | |
| Dyslipidemia | 77 (33%) | 157 (69%) | <0.0001 | 60 (33%) | 117 (73%) | <0.0001 |
| Diabetes | 55 (24%) | 83 (36%) | 0.0033 | 44 (24%) | 55 (34%) | 0.0325 |
| Ischemic Stroke | 34 (15%) | 46 (20%) | 0.1234 | 27 (15%) | 38 (24%) | 0.032 |
| mRS 90 days | | | <0.0001 | | | <0.0001 |
| 0 | 40 (17%) | 75 (33%) | | 28 (15%) | 56 (35%) | |
| 1 | 58 (25%) | 29 (13%) | | 50 (27%) | 20 (12%) | |
| 2 | 19 (8.2%) | 13 (5.7%) | | 17 (9.2%) | 7 (4.4%) | |
| 3 | 36 (16%) | 30 (13%) | | 35 (19%) | 21 (13%) | |
| 4 | 23 (10.0%) | 39 (17%) | | 9 (4.9%) | 24 (15%) | |
| 5 | 18 (7.8%) | 22 (9.6%) | | 13 (7.1%) | 14 (8.8%) | |
| 6 | 37 (16%) | 20 (8.8%) | | 32 (17%) | 18 (11%) | |
| Good outcome (90 day mRS ≤ 1) | 98 (42%) | 104 (46%) | 0.4912 | 78 (42%) | 76 (48%) | 0.3419 |

[1] Median (IQR); n (%)
[2] Wilcoxon rank sum test; Pearson's Chi-squared test

Class-I Group

| | mRS at 90 Days Last Observation Carried Forward (N = 290) | | | mRS at 90 Days Actual (N = 224) | | |
|---|---|---|---|---|---|---|
| Characteristic | ALT, N = 149[1] | TNK, N = 141[1] | *P*-value[2] | ALT, N = 119[1] | TNK, N = 105[1] | *P*-value[2] |
| Age, y | 67 (56, 78) | 67 (57, 76) | 0.7825 | 69 (56, 78) | 68 (57, 76) | 0.8654 |
| Male Sex | 73 (49%) | 94 (67%) | 0.0023 | 59 (50%) | 66 (63%) | 0.0458 |
| NIHSS prior to treatment | 8 (4, 16) | 8 (4, 13) | 0.4162 | 8 (4, 16) | 9 (4, 15) | 0.9571 |
| Glucose level, mg/dL | 119 (106, 143) | 129 (107, 175) | 0.0155 | 118 (108, 143) | 128 (106, 175) | 0.0724 |
| Imaging modality | | | <0.0001 | | | <0.0001 |
| CT imaging | 53 (36%) | 13 (9.2%) | | 48 (40%) | 11 (10%) | |
| CT with perfusion imaging | 67 (45%) | 80 (57%) | | 45 (38%) | 55 (52%) | |
| Diffusion and perfusion MRI | 29 (19%) | 48 (34%) | | 26 (22%) | 39 (37%) | |
| Dyslipidemia | 54 (36%) | 99 (70%) | <0.0001 | 42 (35%) | 75 (71%) | <0.0001 |
| Diabetes | 29 (19%) | 56 (40%) | 0.0002 | 24 (20%) | 40 (38%) | 0.003 |
| Ischemic Stroke | 19 (13%) | 28 (20%) | 0.1007 | 15 (13%) | 22 (21%) | 0.0932 |
| mRS 90 days | | | 0.0253 | | | 0.0016 |
| 0 | 29 (19%) | 43 (30%) | | 21 (18%) | 37 (35%) | |
| 1 | 38 (26%) | 21 (15%) | | 34 (29%) | 15 (14%) | |
| 2 | 15 (10%) | 9 (6.4%) | | 14 (12%) | 5 (4.8%) | |
| 3 | 20 (13%) | 20 (14%) | | 19 (16%) | 14 (13%) | |
| 4 | 16 (11%) | 24 (17%) | | 6 (5.0%) | 14 (13%) | |
| 5 | 12 (8.1%) | 15 (11%) | | 9 (7.6%) | 11 (10%) | |
| 6 | 19 (13%) | 9 (6.4%) | | 16 (13%) | 9 (8.6%) | |
| Good outcome (90 day mRS ≤ 1) | 67 (45%) | 64 (45%) | 0.9422 | 55 (46%) | 52 (50%) | 0.6212 |

[1] Median (IQR); n (%)

[2] Wilcoxon rank sum test; Pearson's Chi-squared test

Supplemental Table S2. Modified Rankin Score (mRS) at 90 days comparing alteplase and tenecteplase. Regression analyses.

Entire Cohort

| | mRS Last Observation Carried Forward | | | | mRS at 90 Days | | |
|---|---|---|---|---|---|---|---|
| | Proportional OR | 95% CI | *P*-value | | Proportional OR | 95% CI | *P*-value |
| Ordinal for 90 day mRS | | | | Ordinal for 90 day mRS | | | |
| Unadjusted | 0.73 | (0.53, 1.01) | 0.0576 | Unadjusted | 0.71 | (0.48, 1.03) | 0.0709 |
| Adjusted | 0.66 | (0.47, 0.92) | 0.0157 | Adjusted | 0.66 | (0.45, 0.97) | 0.0343 |
| Logistic for Good Outcome (90 day mRS ≤ 1) | | | | Logistic for Good Outcome (90 day mRS ≤ 1) | | | |
| Unadjusted | 1.14 | (0.79, 1.65) | 0.4910 | Unadjusted | 1.23 | (0.80, 1.89) | 0.3421 |
| Adjusted | 1.08 | (0.72 1.61) | 0.7240 | Adjusted | 1.26 | (0.79 2.02) | 0.3405 |

Adjustment variables: age, sex, NIHSS prior to treatment, last known well to IV bolus, mRS date

Class-I Cohort

| | mRS Last Observation Carried Forward | | | | mRS at 90 Days | | |
|---|---|---|---|---|---|---|---|
| | Proportional OR | 95% CI | *P*-value | | Proportional OR | 95% CI | *P*-value |
| Ordinal for 90 day mRS | | | | Ordinal for 90 day mRS | | | |
| Unadjusted | 0.84 | (0.56, 1.25) | 0.3858 | Unadjusted | 0.77 | (0.48, 1.22) | 0.2659 |
| Adjusted | 0.81 | (0.53, 1.24) | 0.3311 | Adjusted | 0.75 | (0.46, 1.20) | 0.2285 |
| Logistic for Good Outcome (90 day mRS ≤ 1) | | | | Logistic for Good Outcome (90 day mRS ≤ 1) | | | |
| Unadjusted | 1.01 | (0.64, 1.62) | 0.9420 | Unadjusted | 1.14 | (0.68, 1.93) | 0.6210 |
| Adjusted | 0.94 | (0.57, 1.54) | 0.7930 | Adjusted | 1.17 | (0.66, 2.09) | 0.5983 |

Adjustment variables: age, sex, NIHSS prior to treatment, last known well to IV bolus, mRS date

Interpretation:

- For the Ordinal regression, lower OR favors tenecteplase (TNK). The odds of a 1-point *increase* in score on the mRS scale are *decreased* for patients in the entire cohort receiving TNK, OR=.66, when adjusted for the noted variables.
- For the Logistic regression, higher OR favors tenecteplase (TNK).

Supplemental Figure S2. Modified Rankin Score (mRS) at 90 days comparing alteplase and tenecteplase.

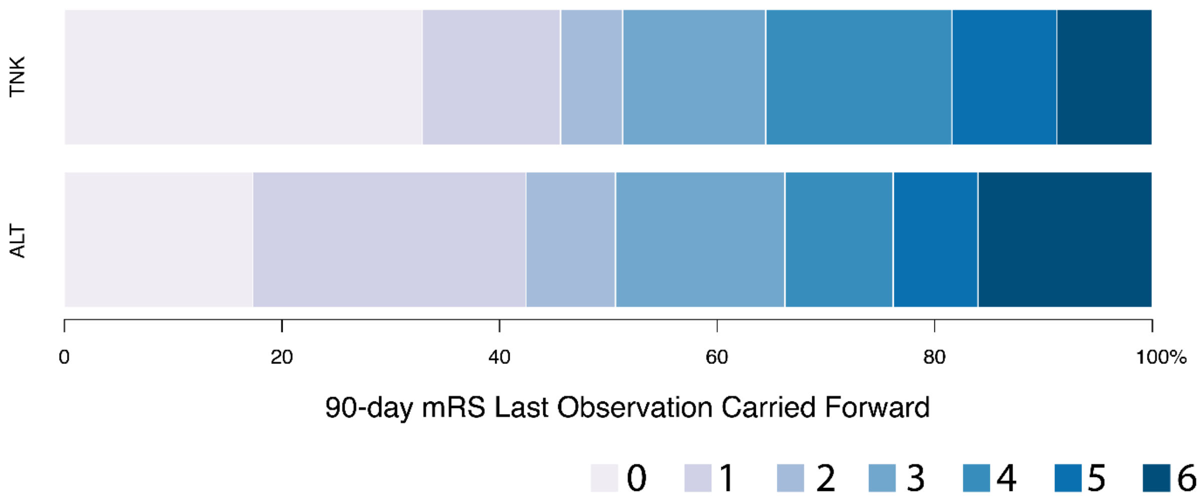


| | mRS Last Observation Carried Forward | | |
|---|---|---|---|
| | Proportional OR | 95% CI | *P*-value |
| **Ordinal for 90 day mRS** | | | |
| Unadjusted | 0.73 | (0.53, 1.01) | 0.0576 |
| Adjusted | 0.66 | (0.47, 0.92) | 0.0157 |
