## Supplementary material for "Prospective Observational Cohort Study Of Tenecteplase Versus Alteplase In Routine Clinical Practice": Ascension Seton Stroke Registry Data Dictionary

### Seton Stroke Program `PID 1816`

#### Data Dictionary Codebook

02/02/2021 2:00pm

Collapse all instruments

<table>
<tr><th>#</th><th>Variable / Field Name</th><th>Field Label<br>Field Note</th><th>Field Attributes (Field Type, Validation, Choices, Calculations, etc.)</th></tr>
<tr><td colspan="4">Instrument: Initial Stroke Information (initial_stroke_information) Collapse</td></tr>
<tr><td>1</td><td>fin</td><td>✪ FIN</td><td>text (integer, Min: 400000, Max: 9999999999), Identifier</td></tr>
<tr><td>2</td><td>mrn</td><td>✪ MRN</td><td>text (integer), Required, Identifier</td></tr>
<tr><td>3</td><td>site</td><td>✪ Site</td><td>dropdown, Required<table>
<tr><td>UMCB</td><td>DSMCUT</td></tr>
<tr><td>SMCA</td><td>SMCA</td></tr>
<tr><td>SMCH</td><td>SMCH</td></tr>
<tr><td>SMCW</td><td>SMCW</td></tr>
<tr><td>ASB</td><td>ASB</td></tr>
<tr><td>SEBD</td><td>SEBD</td></tr>
<tr><td>SHL</td><td>SHL</td></tr>
<tr><td>SNW</td><td>SNW</td></tr>
<tr><td>SSRH</td><td>SSRH</td></tr>
<tr><td>SSW</td><td>SSW</td></tr>
</table></td></tr>
<tr><td>4</td><td>jc_firstname</td><td>✪ First Name</td><td>text, Identifier<br>Field Annotation: @CHARLIMIT=25</td></tr>
<tr><td>5</td><td>jc_lastname</td><td>✪ Last Name</td><td>text, Identifier<br>Field Annotation: @CHARLIMIT=25</td></tr>
<tr><td>6</td><td>desc_jc_arrdatetime</td><td></td><td>descriptive</td></tr>
</table>

| | | | |
|---|---|---|---|
| 7 | jc_arrdatetime_precision | ✪ Arrival Date/Time Precision | dropdown<br>5 \| Date & Time (MM/DD/YYYY HH24:MM)<br>3 \| Date Only (MM/DD/YYYY)<br>0 \| Unknown |
| 8 | jc_arrdatetime<br><br>Show the field ONLY if:<br>[jc_arrdatetime_precision] <> '0' | ✪ Arrival Date/Time<br>*If Date Only, enter 00:00 for time.* | text (datetime_mdy, Min: 2010-01-01 00:00, Max: 2099-12-31 23:59), Identifier |
| 9 | gs_notadmit | ✪ Was the patient admitted? | radio<br>2 \| Yes, patient admitted as inpatient<br>1 \| No, patient was not admitted |
| 10 | gs_notadmitwhy<br><br>Show the field ONLY if:<br>[gs_notadmit] = '1' | ✪ Reason Not Admitted | dropdown<br>1 \| Transferred from your ED to another acute care hospital<br>2 \| Discharged directly from ED to home or other location that is not an acute care hospital<br>3 \| Left from ED AMA<br>4 \| Died in ED<br>5 \| Discharged from observation status without an inpatient admission<br>6 \| Other |
| 11 | unit<br><br>Show the field ONLY if:<br>[site]='SMCW' | ✪ Care Unit(s) | checkbox<br>1 \| unit___1 \| ICU<br>2 \| unit___2 \| Observation<br>3 \| unit___3 \| 3rd Floor<br>4 \| unit___4 \| 4th Floor<br>5 \| unit___5 \| 5th Floor |
| 12 | transfer_to_hosp<br><br>Show the field ONLY if:<br>[gs_notadmitwhy] = '1' and [gs_ahaidnd] <> '1' | ✪ If patient transferred from your ED to another hospital, specify hospital name: | dropdown<br>1 \| DSMCUT<br>2 \| SMCA<br>3 \| SMCH<br>4 \| SMCW |
| 13 | gs_ahaidnd<br><br>Show the field ONLY if:<br>[gs_notadmitwhy] = '1' | ✪ Hospital not documented | radio<br>1 \| Not documented |

| | | | |
|---|---|---|---|
| 14 | gs_ptarrtranrsn<br><br>Show the field ONLY if:<br>[gs_notadmitwhy] = '1' | ✪ Select reason(s) for why patient transferred | checkbox, Required<br>1 \| gs_ptarrtranrsn___1 \| Evaluation for IV tPA up to 4.5 hours<br>2 \| gs_ptarrtranrsn___2 \| Post Management of IV tPA (e.g. Drip and Ship)<br>3 \| gs_ptarrtranrsn___3 \| Evaluation for Endovascular thrombectomy<br>4 \| gs_ptarrtranrsn___4 \| Advanced stroke care (e.g., Neurocritical care, surgical or other time critical therapy)<br>5 \| gs_ptarrtranrsn___5 \| Patient/family request<br>6 \| gs_ptarrtranrsn___6 \| Other advanced care (not stroke related)<br>7 \| gs_ptarrtranrsn___7 \| Not documented |
| 15 | treatment_candidate | Section Header: *Acute Stroke Treatment*<br>✪ Was this patient an Acute Stroke Treatment candidate? | yesno<br>1 \| Yes<br>0 \| No<br><br>Custom alignment: RH |
| 16 | treatment_recvd<br><br>Show the field ONLY if:<br>[treatment_candidate]='1' | ✪ Did this patient receive an Acute Stroke Treatment? | yesno<br>1 \| Yes<br>0 \| No<br><br>Custom alignment: RH |
| 17 | candidate_exclusion<br><br>Show the field ONLY if:<br>[treatment_candidate] = '0' or [treatment_recvd] = '0' | ✪ If not an acute stroke treatment candidate, or not treated, documented reason for exclusion: | notes<br>Custom alignment: RH |
| 18 | treatment_type<br><br>Show the field ONLY if:<br>[treatment_recvd]='1' | ✪ Type of Acute Stroke Treatment received by patient:<br>*Select all that apply.* | checkbox<br>IAtPA \| treatment_type___iatpa \| IA thrombolytic<br>IVtPA \| treatment_type___ivtpa \| IV thrombolytic<br>MER \| treatment_type___mer \| MER (Thrombectomy) |
| 19 | iv_thrombolytic_type<br><br>Show the field ONLY if:<br>[treatment_type(IVtPA)]='1' | ✪ IV Thrombolytic Type | radio<br>1 \| Alteplase (Activase)<br>2 \| Tenecteplase (TNKase) |

| | | | |
|---|---|---|---|
| 20 | desc_treat_transfer_gwtg<br><br>Show the field ONLY if:<br>[gs_notadmitwhy]='1' and [treatment_recvd]='1' | This patient received an Acute Stroke Treatment and was transferred to another hospital.This record will need to be uploaded to Get With The Guidelines.To hide all fields not required for GWTG, check the box below. | descriptive |
| 21 | hide_nonessential_fields<br><br>Show the field ONLY if:<br>[gs_notadmitwhy]='1' and [treatment_recvd]='1' | To hide fields not required for GWTG submission: | checkbox<br>1 \| hide_nonessential_fields___1 \| Hide non-required fields |
| 22 | gs_symptomlocation<br><br>Show the field ONLY if:<br>([treatment_recvd]='1' or [gs_notadmit]<>'1') | Section Header: *Symptom Onset*<br><br>✪ Patient location when stroke symptoms discovered | dropdown<br>1 \| Not in a healthcare setting<br>2 \| Another acute care facility<br>3 \| Chronic health care facility<br>5 \| Outpatient healthcare setting<br>4 \| Stroke occurred after hospital arrival (in ED/Obs/inpatient)<br>9 \| ND or Cannot be determined |
| 23 | desc_gs_lastknownwell | | descriptive |
| 24 | gs_lastknownwell_precision | ✪ Last Known Well Precision<br>*For OP-STK ONLY, if the Time Last Known Well is clearly greater than 2 hours prior to hospital arrival and no specific time is documented, select Unknown.* | dropdown<br>5 \| Date & Time (MM/DD/YYYY HH24:MM)<br>3 \| Date Only (MM/DD/YYYY)<br>0 \| Unknown |
| 25 | gs_lastknownwell<br><br>Show the field ONLY if:<br>[gs_lastknownwell_precision] <> '0' | ✪ Date/Time patient last known to be well<br>*If Date Only, enter 00:00 for time.* | text (datetime_mdy, Min: 2010-01-01 00:00, Max: 2099-12-31 23:59), Identifier |
| 26 | gs_symptomdatetime_precision<br><br>Show the field ONLY if:<br>([treatment_recvd]='1' or [gs_notadmit]<>'1') | ✪ Symptom Date/Time Precision | dropdown<br>5 \| Date & Time (MM/DD/YYYY HH24:MM)<br>3 \| Date Only (MM/DD/YYYY)<br>0 \| Unknown |
| 27 | gs_symptomdatetime<br><br>Show the field ONLY if:<br>[gs_symptomdatetime_precision] <> '0' and ([treatment_recvd]='1' or [gs_notadmit]<>'1') | ✪ Date/Time of discovery of stroke or stroke-like symptoms<br>*If Date Only, enter 00:00 for time.* | text (datetime_mdy, Min: 2010-01-01 00:00, Max: 2099-12-31 23:59), Identifier |

| | | | |
|---|---|---|---|
| 28 | doortime_lkw<br><br>Show the field ONLY if:<br>[gs_symptomlocation] = '4' | ✪ Door Time for Last Known Well to Arrival<br>*Arrival Time used by Default* | radio<br>2 Use Symptom Discovery Time |
| 29 | calc_lkw_to_arrival | Last Known Well to Arrival<br>*Time in MinutesHH:MM - [calc_lkw_to_arrival_hrs]:[calc_lkw_to_arrival_min]* | calc<br>Calculation: if([doortime_lkw]='2',datediff([gs_lastknownwell], [gs_symptomdatetime], "m", "mdy", true), datediff([gs_lastknownwell], [jc_arrdatetime], "m", "mdy", true)) |
| 30 | calc_lkw_to_arrival_min | Last Known Well to Arrival (Minutes Only)<br>*Time in Hours* | calc<br>Calculation: if([doortime_lkw]='2', datediff([gs_lastknownwell], [gs_symptomdatetime], "m", "mdy", true) - (60* [calc_lkw_to_arrival_hrs]), datediff([gs_lastknownwell], [jc_arrdatetime], "m", "mdy", true) - (60* [calc_lkw_to_arrival_hrs]))<br>Field Annotation: @HIDDEN |
| 31 | calc_lkw_to_arrival_hrs | Last Known Well to Arrival (Hours)<br>*Time in Hours* | calc<br>Calculation: if([doortime_lkw]='2',rounddown(datediff([gs_lastknownwell], [gs_symptomdatetime], "h", "mdy", true),0), rounddown(datediff([gs_lastknownwell], [jc_arrdatetime], "h", "mdy", true),0))<br>Field Annotation: @HIDDEN |
| 32 | gs_pc_onsetcom<br><br>Show the field ONLY if:<br>([treatment_recvd]='1' or [gs_notadmit]<>'1') | ✪ Symptom Presentation History<br>*Symptom Onset Comments* | notes, Identifier<br>Field Annotation: @CHARLIMIT=200 |
| 33 | desc_patient_arrival | Section Header: *Patient Arrival* | descriptive |
| 34 | gs_patientarrival<br><br>Show the field ONLY if:<br>[gs_symptomlocation] <> '4' and ([treatment_recvd]='1' or [gs_notadmit]<>'1') | ✪ How patient arrived at your hospital | dropdown<br>1 EMS from home/scene<br>10 Mobile Stroke Unit<br>2 Private transport/taxi/other from home/scene<br>3 Transfer from other hospital<br>9 ND or unknown |
| 35 | transport_method<br><br>Show the field ONLY if:<br>[gs_patientarrival]='1' or [gs_patientarrival]='10' or [gs_patientarrival]='3' | ✪ Please select method of transport. | radio<br>1 Ground<br>2 Air<br><br>Custom alignment: RH |
| 36 | transfer_from_hosp | ✪ If transfered from another hospital, specify hospital name: | dropdown |

| | |
|---|---|
| | Show the field ONLY if:<br>[gs_patientarrival] = '3' and [gs_ahaid2nd] <> '1' |

| | |
|---|---|
| 27 | Ascension Seton Bastrop |
| 9 | Ascension Seton Edgar B. Davis |
| 32 | Ascension Seton Harker Heights |
| 10 | Ascension Seton Highland Lakes |
| 31 | Ascension Seton Highland Lakes Marble Falls Healthcare Center |
| 2 | Ascension Seton Medical Center Austin |
| 3 | Ascension Seton Hays |
| 4 | Ascension Seton Williamson |
| 12 | Ascension Seton Northwest |
| 13 | Ascension Seton Smithville |
| 14 | Ascension Seton Southwest |
| 24 | Baylor Scott & White Emergency Medical Center - Cedar Park |
| 28 | Baylor Scott & White Medical Center - Buda |
| 25 | Baylor Scott & White Medical Center - Lakeway |
| 16 | Baylor Scott & White Medical Center - Marble Falls |
| 23 | Baylor Scott & White Medical Center - Pflugerville |
| 5 | Baylor Scott & White Medical Center - Round Rock |
| 26 | Baylor Scott & White Medical Center - Taylor |
| 6 | Cedar Park Regional Medical Center |
| 7 | Central Texas Medical Center |
| 30 | Central Texas Rehabilitation Hospital |
| 29 | Dell Children's Medical Center of Central Texas |
| 1 | Dell Seton Medical Center at the University of Texas |
| 19 | Free-Standing Emergency Room |
| 8 | Gonzales Memorial Hospital |
| 18 | Non-Regional Hospital |
| 11 | Seton Medical Center Harker Heights |
| 20 | St. David's Medical Center |
| 21 | St. David's North Austin Medical Center |
| 17 | St. David's Round Rock Medical Center |
| 22 | St. David's South Austin Medical Center |

| | | | |
|---|---|---|---|
| | | | 15 Westlake Medical Center |
| 37 | gs_ahaid2nd<br>Show the field ONLY if:<br>[gs_patientarrival] = '3' | ✪ Hospital not documented | radio<br>1 Not documented |
| 38 | transferred_fin<br>Show the field ONLY if:<br>[transfer_from_hosp]=1 or [transfer_from_hosp]=2 or [transfer_from_hosp]=3 or [transfer_from_hosp]=4 or [transfer_from_hosp]=9 or [transfer_from_hosp]=10 or [transfer_from_hosp]=11 or [transfer_from_hosp]=12 or [transfer_from_hosp]=13 or [transfer_from_hosp]=14 or [transfer_from_hosp]=27 or [transfer_from_hosp]=29 or [transfer_from_hosp]=31 or [transfer_from_hosp]=32 or [transfer_to_hosp]<>'' | ✪ FIN from encounter at other Ascension facility (transferred patients only) | text (integer, Min: 400000, Max: 9999999999) |
| 39 | ems_recordavail<br>Show the field ONLY if:<br>([site]='SMCA' or [site]='SMCH' or [site]='SMCW' or [site]='UMCB') and ([gs_patientarrival]='1' or [gs_patientarrival]='3' or [gs_patientarrival]='10') | ✪ EMS run sheet available at time of patient arrival? | radio<br>1 Yes<br>2 No/ND<br><br>Custom alignment: RH |
| 40 | transfer_firstcall<br>Show the field ONLY if:<br>[gs_patientarrival] = '3' | ✪ What was the Date and Time of the first call from the transferring hospital?<br>*No longer Departure Time.* | text (datetime_mdy, Min: 2010-01-01 00:00, Max: 2099-12-31 23:59), Identifier<br>Field Annotation: @HIDDEN |
| 41 | stroke_alert | ✪ Was 324-TIME activated? | radio<br>1 Yes<br>0 No<br><br>Custom alignment: RH |
| 42 | gs_symptresolve<br>Show the field ONLY if:<br>([treatment_recvd]='1' or [gs_notadmit]<>'1') | ✪ Had stroke symptoms resolved at time of presentation? | radio<br>1 Yes<br>2 No<br>3 ND<br><br>Custom alignment: RH |

| | | | |
|---|---|---|---|
| 43 | gs_nihssperformed<br><br>Show the field ONLY if:<br>([treatment_recvd]='1' or [gs_notadmit]<>'1') | ✪ Initial NIH Stroke Scale | radio<br>1 Yes<br>2 No/ND<br><br>Custom alignment: RH |
| 44 | gs_nihssproretro<br><br>Show the field ONLY if:<br>[gs_nihssperformed] = '1' | ✪ if Yes, Initial NIHSS: | radio<br>1 Actual<br>2 Estimated from record<br>3 ND |
| 45 | gs_nihssadmit<br><br>Show the field ONLY if:<br>[gs_nihssperformed] = '1' | ✪ Total Score | text (integer, Min: 0, Max: 42) |
| 46 | c_nihssivtutd<br><br>Show the field ONLY if:<br>([site]='SMCA' or [site]='UMCB' or [site]='SMCH' or [site]='SMCW') and [treatment_type(IVtPA)]='1' | ✪ Unable to determine the last NIHSS score documented prior to initiation of IV thrombolytic therapy at this hospital. | radio<br>1 UTD<br><br>Custom alignment: RH |
| 47 | mer_ivscoreobt<br><br>Show the field ONLY if:<br>([site]='SMCA' or [site]='UMCB' or [site]='SMCH' or [site]='SMCW') and [treatment_type(IVtPA)]='1' and [c_nihssivtutd]<>'1' | ✪ This score obtained from:<br>*If Baseline NIHSS, use [gs_nihssadmit].* | radio<br>1 Baseline NIHSS<br>2 Subsequent NIHSS<br><br>Custom alignment: RH |
| 48 | c_nihssivt<br><br>Show the field ONLY if:<br>([site]='SMCA' or [site]='UMCB' or [site]='SMCH' or [site]='SMCW') and [treatment_type(IVtPA)]='1' and [c_nihssivtutd]<>'1' | ✪ What is the last NIHSS score documented prior to initiation of IV thrombolytic therapy at this hospital? | text (integer, Min: 0, Max: 42)<br>Custom alignment: RH |
| 49 | informed_consent<br><br>Show the field ONLY if:<br>[treatment_type(IVtPA)] = '1' | ✪ Informed Consent (risks, benefits, and alternative therapies regarding tPA) found in medical record? | yesno, Required<br>1 Yes<br>0 No<br><br>Custom alignment: RH |

| | | | |
|---|---|---|---|
| 50 | gs_telestrokeconsult<br><br>Show the field ONLY if:<br>([site] = 'SMCW' or [site] = 'SMCH' or [site] = 'SMCA') and [treatment_candidate] =1 | Section Header: *Telestroke*<br><br>✪ Was telestroke consultation performed? | radio<br>1 Yes, the patient received telestroke consultation from my hospital staff when the patient was located at another hospital<br>2 Yes, the patient received telestroke consultation from someone other than my staff when the patient was located at another hospital<br>3 Yes, the patient received telestroke consultation from a remotely located expert when the patient was located at my hospital<br>4 No telestroke consult performed<br>5 Not Documented |
| 51 | gs_teleconmtd<br><br>Show the field ONLY if:<br>[gs_telestrokeconsult] = '1' or [gs_telestrokeconsult] = '2' or [gs_telestrokeconsult] = '3' | ✪ If Yes, telestroke consult performed, select all applicable delivery methods. | checkbox<br>1 gs_teleconmtd___1 Interactive Video<br>2 gs_teleconmtd___2 Teleradiology<br>3 gs_teleconmtd___3 Telephone Call<br>4 gs_teleconmtd___4 ND<br><br>Field Annotation: @NONEOFTHEABOVE='4' |
| 52 | desc_gs_teleconsultdt<br><br>Show the field ONLY if:<br>([gs_telestrokeconsult] = '1' or [gs_telestrokeconsult] = '2' or [gs_telestrokeconsult] = '3') | | descriptive |
| 53 | gs_teleconsultdt_precision<br><br>Show the field ONLY if:<br>([gs_telestrokeconsult] = '1' or [gs_telestrokeconsult] = '2' or [gs_telestrokeconsult] = '3') | ✪ Date/Time of Telestroke Consultation Precision | dropdown<br>5 Date & Time (MM/DD/YYYY HH24:MM)<br>0 Unknown |
| 54 | gs_teleconsultdt<br><br>Show the field ONLY if:<br>([gs_telestrokeconsult] = '1' or [gs_telestrokeconsult] = '2' or [gs_telestrokeconsult] = '3') and [gs_teleconsultdt_precision] <> '0' | ✪ If Yes, What was the date/time of the telestroke consultation?<br>*Use computer link time if the telestroke cart was used.* | text (datetime_mdy, Min: 2010-01-01 00:00, Max: 2099-12-31 23:59) |

| | | | |
|---|---|---|---|
| 55 | doortime_telestroke<br><br>Show the field ONLY if:<br>([gs_telestrokeconsult] = '1' or [gs_telestrokeconsult] = '2' or [gs_telestrokeconsult] = '3') and [gs_teleconsultdt_precision] <> '0' and [gs_symptomlocation] = '4' | ✪ Door Time for Door to Telestroke<br>*Arrival Time used by Default* | radio<br>2 \| Use Symptom Discovery Time |
| 56 | calc_door_to_telestroke<br><br>Show the field ONLY if:<br>([gs_telestrokeconsult] = '1' or [gs_telestrokeconsult] = '2' or [gs_telestrokeconsult] = '3') and [gs_teleconsultdt_precision] <> '0' | Door to Telestroke<br>*Time in Minutes - Goal: ≤ 20 Minutes* | calc<br>Calculation:<br>if([doortime_telestroke]='2',datediff([gs_symptomdatetime],[gs_teleconsultdt], "m", "mdy", true), datediff([jc_arrdatetime],[gs_teleconsultdt], "m", "mdy", true)) |
| 57 | att_fields_override | Section Header: *Time Tracker Fields*<br><br>✪ Show/Hide Additional Time Tracker Fields | radio<br>1 \| Show<br>0 \| Hide<br><br>Custom alignment: RH |
| 58 | gs_jp_teamna<br><br>Show the field ONLY if:<br>([treatment_recvd]='1' or ([gs_lastknownwell_precision]<>'0' and [calc_lkw_to_arrival]<=1440 and [gs_patientarrival]<>'3') or [att_fields_override]='1' or [gs_jp_teamna]<>'') and [att_fields_override]<>'0' | ✪ Stroke Team | radio<br>1 \| N/A |
| 59 | desc_teamdatetime<br><br>Show the field ONLY if:<br>[gs_jp_teamna] <> '1' and ([treatment_recvd]='1' or ([gs_lastknownwell_precision]<>'0' and [calc_lkw_to_arrival]<=1440 and [gs_patientarrival]<>'3') or [att_fields_override]='1' or [gs_jp_teamdatetime_precision]<>'') and [att_fields_override]<>'0' | | descriptive |

| | | | |
|---|---|---|---|
| 60 | gs_jp_teamdatetime_precision<br><br>Show the field ONLY if:<br>[gs_jp_teamna] <> '1' and ([treatment_recvd]='1' or ([gs_lastknownwell_precision]<>'0' and [calc_lkw_to_arrival]<=1440 and [gs_patientarrival]<>'3') or [att_fields_override]='1' or [gs_jp_teamdatetime_precision]<>'') and [att_fields_override]<>'0' | ✪ Stroke Team Activated Precision | dropdown<br>5 Date & Time (MM/DD/YYYY HH24:MM)<br>3 Date Only (MM/DD/YYYY)<br>0 Unknown |
| 61 | desc_cs_notification_dt<br><br>Show the field ONLY if:<br>[gs_jp_teamna]<>'1' and [gs_jp_teamdatetime_precision]<>'0' and [cs_notification_dt]<>'' and ([treatment_recvd]='1' or ([gs_lastknownwell_precision]<>'0' and [calc_lkw_to_arrival]<=1440 and [gs_patientarrival]<>'3') or [att_fields_override]='1' or [stroke_alert]='1' or [desc_cs_notification_dt]<>'') and [att_fields_override]<>'0' | Date/Time Code Stroke Notification sent to Stroke Team: (Neurology contacted or paged)[cs_notification_dt]From: Code Stroke▲ ▼ copy the date/time into the field below | descriptive, Identifier<br>Field Annotation: @HIDDEN |
| 62 | gs_jp_teamdatetime<br><br>Show the field ONLY if:<br>[gs_jp_teamna]<>'1' and [gs_jp_teamdatetime_precision]<>'0' and ([treatment_recvd]='1' or ([gs_lastknownwell_precision]<>'0' and [calc_lkw_to_arrival]<=1440 and [gs_patientarrival]<>'3') or [att_fields_override]='1' or [gs_jp_teamdatetime]<>'') and [att_fields_override]<>'0' | ✪ Date/Time Stroke Team Activated<br>*If Date Only, enter 00:00 for time.* | text (datetime_mdy, Min: 2010-01-01 00:00, Max: 2099-12-31 23:59), Identifier |
| 63 | gs_jp_arrivdatetime_precision<br><br>Show the field ONLY if:<br>[gs_jp_teamna]<>'1' and ([treatment_recvd]='1' or ([gs_lastknownwell_precision]<>'0' and [calc_lkw_to_arrival]<=1440 and [gs_patientarrival]<>'3') or [att_fields_override]='1' or [gs_jp_arrivdatetime_precision]<>'') and [att_fields_override]<>'0' | ✪ Date/Time Stroke Team Arrived Precision | dropdown<br>5 Date & Time (MM/DD/YYYY HH24:MM)<br>3 Date Only (MM/DD/YYYY)<br>0 Unknown |

| | | | |
|---|---|---|---|
| 64 | gs_jp_arrivdatetime<br><br>Show the field ONLY if:<br>[gs_jp_teamna]<>'1' and [gs_jp_arrivdatetime_precision]<>'0' and ([treatment_recvd]='1' or ([gs_lastknownwell_precision]<>'0' and [calc_lkw_to_arrival]<=1440 and [gs_patientarrival]<>'3') or [att_fields_override]='1' or [gs_jp_arrivdatetime]<>'') and [att_fields_override]<>'0' | ✪ Date/Time Stroke Team Arrived (Neurology at bedside or on tele-stroke)<br>*If Date Only, enter 00:00 for time.* | text (datetime_mdy, Min: 2010-01-01 00:00, Max: 2099-12-31 23:59), Identifier |
| 65 | cs_neuro_notified_dt<br><br>Show the field ONLY if:<br>([treatment_recvd]='1' or ([gs_lastknownwell_precision]<>'0' and [calc_lkw_to_arrival]<=1440) or [att_fields_override]='1' or [cs_neuro_notified_dt]<>'') and [att_fields_override]<>'0' | ✪ Time Neurologist Notified | text (datetime_mdy, Min: 2010-01-01 00:00, Max: 2099-12-31 23:59), Identifier |
| 66 | resident_md_name<br><br>Show the field ONLY if:<br>[site] = 'UMCB' and ([treatment_recvd]='1' or [gs_notadmit]<>'1') | ✪ Name of Resident | text<br>Custom alignment: RH |
| 67 | neuro_md_name<br><br>Show the field ONLY if:<br>([treatment_recvd]='1' or [gs_notadmit]<>'1') | ✪ Name of Neuro MD | text |
| 68 | gs_jp_edphysna<br><br>Show the field ONLY if:<br>([treatment_recvd]='1' or ([gs_lastknownwell_precision]<>'0' and [calc_lkw_to_arrival]<=1440 and [gs_patientarrival]<>'3') or [att_fields_override]='1' or [gs_jp_edphysna]<>'') and [att_fields_override]<>'0' | ✪ ED Physician Assessment | radio<br>1 \| N/A |

| | | | |
|---|---|---|---|
| 69 | ed_phys_dt_source<br><br>Show the field ONLY if:<br>[gs_jp_edphysna]<>'1' and [gs_jp_eddatetime_precision]<>'0' and ([treatment_recvd]='1' or ([gs_lastknownwell_precision]<>'0' and [calc_lkw_to_arrival]<=1440 and [gs_patientarrival]<>'3') or [att_fields_override]='1' or [ed_phys_dt_source]<>'') and [att_fields_override]<>'0' | ✪ ED Physician Assessment Time Source | radio, Required<br>1 Physician's Note<br>2 Compass Timestamp |
| 70 | desc_eddatetime<br><br>Show the field ONLY if:<br>[gs_jp_edphysna]<>'1' and ([treatment_recvd]='1' or ([gs_lastknownwell_precision]<>'0' and [calc_lkw_to_arrival]<=1440 and [gs_patientarrival]<>'3') or [att_fields_override]='1' or [gs_jp_eddatetime_precision]<>'') and [att_fields_override]<>'0' | | descriptive |
| 71 | gs_jp_eddatetime_precision<br><br>Show the field ONLY if:<br>[gs_jp_edphysna]<>'1' and ([treatment_recvd]='1' or ([gs_lastknownwell_precision]<>'0' and [calc_lkw_to_arrival]<=1440 and [gs_patientarrival]<>'3') or [att_fields_override]='1' or [gs_jp_eddatetime_precision]<>'') and [att_fields_override]<>'0' | ✪ ED Physician Assessment Precision | dropdown<br>5 Date & Time (MM/DD/YYYY HH24:MM)<br>3 Date Only (MM/DD/YYYY)<br>0 Unknown |
| 72 | gs_jp_eddatetime<br><br>Show the field ONLY if:<br>[gs_jp_edphysna]<>'1' and [gs_jp_eddatetime_precision]<>'0' and ([treatment_recvd]='1' or ([gs_lastknownwell_precision]<>'0' and [calc_lkw_to_arrival]<=1440 and [gs_patientarrival]<>'3') or [att_fields_override]='1' or [gs_jp_eddatetime]<>'') and [att_fields_override]<>'0' | ✪ Date/Time of ED Physician Assessment<br>*If Date Only, enter 00:00 for time.* | text (datetime_mdy, Min: 2010-01-01 00:00, Max: 2099-12-31 23:59), Required, Identifier |

| | | | |
|---|---|---|---|
| 73 | doortime_md<br><br>Show the field ONLY if: [gs_jp_edphysna]<>'1' and [gs_jp_eddatetime_precision]<>'0' and ([treatment_recvd]='1' or ([gs_lastknownwell_precision]<>'0' and [calc_lkw_to_arrival]<=1440 and [gs_patientarrival]<>'3') or [att_fields_override]='1' or [doortime_md]<>'') and [att_fields_override]<>'0' and [gs_symptomlocation]='4' | ✪ Door Time for Door to MD<br>*Arrival Time used by Default* | radio<br>2 Use Symptom Discovery Time |
| 74 | calc_door_to_md<br><br>Show the field ONLY if: [gs_jp_edphysna]<>'1' and [gs_jp_eddatetime_precision]<>'0' and ([treatment_recvd]='1' or ([gs_lastknownwell_precision]<>'0' and [calc_lkw_to_arrival]<=1440 and [gs_patientarrival]<>'3') or [att_fields_override]='1' or [calc_door_to_md]<>'') and [att_fields_override]<>'0' | Door to ED Physician Assessment<br>*Time in Minutes - Goal: ≤ 10 Minutes* | calc<br>Calculation: if([gs_jp_eddatetime_precision]<>'3', if([doortime_md]='2', datediff([gs_symptomdatetime], [gs_jp_eddatetime], "m", "mdy", true), datediff([jc_arrdatetime], [gs_jp_eddatetime], "m", "mdy", true)), '') |
| 75 | ed_md_name<br><br>Show the field ONLY if: [gs_jp_edphysna] <> '1' and ([treatment_recvd]='1' or [gs_notadmit]<>'1') | ✪ Name of ED Physician | text<br>Custom alignment: RH |
| 76 | gs_jp_neurosurgna<br><br>Show the field ONLY if: ([treatment_recvd]='1' or ([gs_lastknownwell_precision]<>'0' and [calc_lkw_to_arrival]<=1440 and [gs_patientarrival]<>'3') or [att_fields_override]='1' or [gs_jp_neurosurgna]<>'') and [att_fields_override]<>'0' | ✪ Neurosurgical Services Consulted | radio<br>1 N/A |

| | | | |
|---|---|---|---|
| 77 | desc_nsurgdatetime<br><br>Show the field ONLY if:<br>[gs_jp_neurosurgna]<>'1' and ([treatment_recvd]='1' or ([gs_lastknownwell_precision]<>'0' and [calc_lkw_to_arrival]<=1440 and [gs_patientarrival]<>'3') or [att_fields_override]='1' or [gs_jp_nsurgdatetime_precision]<>'') and [att_fields_override]<>'0' | | descriptive |
| 78 | gs_jp_nsurgdatetime_precision<br><br>Show the field ONLY if:<br>[gs_jp_neurosurgna]<>'1' and ([treatment_recvd]='1' or ([gs_lastknownwell_precision]<>'0' and [calc_lkw_to_arrival]<=1440 and [gs_patientarrival]<>'3') or [att_fields_override]='1' or [gs_jp_nsurgdatetime_precision]<>'') and [att_fields_override]<>'0' | ✪ Neurosurgical Services Consulted Precision | dropdown<br>5 \| Date & Time (MM/DD/YYYY HH24:MM)<br>3 \| Date Only (MM/DD/YYYY)<br>0 \| Unknown |
| 79 | gs_jp_nsurgdatetime<br><br>Show the field ONLY if:<br>[gs_jp_neurosurgna]<>'1' and [gs_jp_nsurgdatetime_precision]<>'0' and ([treatment_recvd]='1' or ([gs_lastknownwell_precision]<>'0' and [calc_lkw_to_arrival]<=1440 and [gs_patientarrival]<>'3') or [att_fields_override]='1' or [gs_jp_nsurgdatetime]<>'') and [att_fields_override]<>'0' | ✪ Date/Time Neurosurgical Services Consulted<br>*If Date Only, enter 00:00 for time.* | text (datetime_mdy, Min: 2010-01-01 00:00, Max: 2099-12-31 23:59), Identifier |
| 80 | mer_imagep<br><br>Show the field ONLY if:<br>[gs_patientarrival]='3' and ([site]='UMCB' or [site]='SMCA' or [site]='SMCW') | Section Header: *Brain Imaging*<br><br>✪ Was brain or vascular imaging performed prior to transfer to your facility? | radio<br>1 \| Yes<br>2 \| No/ND<br><br>Custom alignment: RH |

| | | | |
|---|---|---|---|
| 81 | mer_imagepcomp<br><br>Show the field ONLY if:<br>[mer_imagep]='1' | ✪ If yes, which imaging tests were performed? (select all that apply) | checkbox<br>1 \| mer_imagepcomp___1 \| CT<br>2 \| mer_imagepcomp___2 \| CTA<br>3 \| mer_imagepcomp___3 \| CT Perfusion<br>4 \| mer_imagepcomp___4 \| MRI<br>5 \| mer_imagepcomp___5 \| MRA<br>6 \| mer_imagepcomp___6 \| MR Perfusion<br>0 \| mer_imagepcomp___0 \| Image type not documented<br><br>Field Annotation: @NONEOFTHEABOVE='0' |
| 82 | desc_mer_imagepdt<br><br>Show the field ONLY if:<br>[mer_imagepcomp(1)]='1' or [mer_imagepcomp(2)]='1' or [mer_imagepcomp(3)]='1' or [mer_imagepcomp(4)]='1' or [mer_imagepcomp(5)]='1' or [mer_imagepcomp(6)]='1' | | descriptive |
| 83 | mer_imagepdt_precision<br><br>Show the field ONLY if:<br>[mer_imagepcomp(1)]='1' or [mer_imagepcomp(2)]='1' or [mer_imagepcomp(3)]='1' or [mer_imagepcomp(4)]='1' or [mer_imagepcomp(5)]='1' or [mer_imagepcomp(6)]='1' | ✪ Date/Time 1st vessel or perfusion imaging initiated at prior hospital Precision | dropdown<br>5 \| Date & Time (MM/DD/YYYY HH24:MM)<br>3 \| Date Only (MM/DD/YYYY)<br>0 \| Unknown |
| 84 | mer_imagepdt<br><br>Show the field ONLY if:<br>([mer_imagepcomp(1)]='1' or [mer_imagepcomp(2)]='1' or [mer_imagepcomp(3)]='1' or [mer_imagepcomp(4)]='1' or [mer_imagepcomp(5)]='1' or [mer_imagepcomp(6)]='1') and [mer_imagepdt_precision]<>'0' | ✪ Date/Time 1st vessel or perfusion imaging initiated at prior hospital<br>*If Date Only, enter 00:00 for time.* | text (datetime_mdy, Min: 2010-01-01 00:00, Max: 2099-12-31 23:59), Identifier |

| | | | |
|---|---|---|---|
| 85 | gs_jp_ctna<br><br>Show the field ONLY if:<br>([treatment_recvd]='1' or ([gs_lastknownwell_precision]<>'0' and [calc_lkw_to_arrival]<=1440 and [gs_patientarrival]<>'3') or [att_fields_override]='1' or [gs_jp_ctna]<>'') and [att_fields_override]<>'0' | ✪ Brain Imaging | radio<br>1 \| N/A |
| 86 | gs_ctcomp | ✪ Brain imaging completed at your hospital for this episode of care? | radio<br>1 \| Yes<br>2 \| No/ND<br>3 \| NC<br><br>Custom alignment: RH |
| 87 | imaging_type<br><br>Show the field ONLY if:<br>[gs_ctcomp]='1' | ✪ Initial brain imaging type completed? | radio<br>1 \| CT<br>2 \| CT-Stroke<br>3 \| MRI<br><br>Custom alignment: RH |
| 88 | desc_ctorddatetime<br><br>Show the field ONLY if:<br>[gs_jp_ctna]<>'1' and ([treatment_recvd]='1' or ([gs_lastknownwell_precision]<>'0' and [calc_lkw_to_arrival]<=1440 and [gs_patientarrival]<>'3') or [att_fields_override]='1' or [gs_jp_ctorddatetime_precision]<>'') and [att_fields_override]<>'0' | | descriptive |
| 89 | gs_jp_ctorddatetime_precision<br><br>Show the field ONLY if:<br>[gs_jp_ctna]<>'1' and ([treatment_recvd]='1' or ([gs_lastknownwell_precision]<>'0' and [calc_lkw_to_arrival]<=1440 and [gs_patientarrival]<>'3') or [att_fields_override]='1' or [gs_jp_ctorddatetime_precision]<>'') and [att_fields_override]<>'0' | ✪ Brain Imaging Ordered Precision | dropdown<br>5 \| Date & Time (MM/DD/YYYY HH24:MM)<br>3 \| Date Only (MM/DD/YYYY)<br>0 \| Unknown |

| | | | |
|---|---|---|---|
| 90 | gs_jp_ctorddatetime<br><br>Show the field ONLY if:<br>[gs_jp_ctna]<>'1' and [gs_jp_ctorddatetime_precision]<>'0' and ([treatment_recvd]='1' or ([gs_lastknownwell_precision]<>'0' and [calc_lkw_to_arrival]<=1440 and [gs_patientarrival]<>'3') or [att_fields_override]='1' or [gs_jp_ctorddatetime]<>'') and [att_fields_override]<>'0' | ✪ Date/Time Brain Imaging Ordered<br>*If Date Only, enter 00:00 for time.* | text (datetime_mdy, Min: 2010-01-01 00:00, Max: 2099-12-31 23:59), Identifier |
| 91 | doortime_ct<br><br>Show the field ONLY if:<br>[gs_ctcomp]='1' and ([treatment_recvd]='1' or [gs_notadmit]<>'1') and [gs_symptomlocation]='4' | ✪ Door Time for Brain Imaging<br>*Arrival Time used by Default* | radio<br>2 Use Symptom Discovery Time |
| 92 | calc_door_to_ctord<br><br>Show the field ONLY if:<br>[gs_jp_ctna]<>'1' and [gs_jp_ctorddatetime_precision]<>'0' and ([treatment_recvd]='1' or ([gs_lastknownwell_precision]<>'0' and [calc_lkw_to_arrival]<=1440 and [gs_patientarrival]<>'3') or [att_fields_override]='1' or [calc_door_to_ctord]<>'') and [att_fields_override]<>'0' | Door to CT Ordered<br>*Time in Minutes* | calc<br>Calculation:<br>if([doortime_ct]='2',datediff([gs_symptomdatetime], [gs_jp_ctorddatetime], "m", "mdy", true), datediff([jc_arrdatetime], [gs_jp_ctorddatetime], "m", "mdy", true)) |
| 93 | desc_ctcompdatetime<br><br>Show the field ONLY if:<br>[gs_ctcomp]='1' and ([treatment_recvd]='1' or [gs_notadmit]<>'1') | | descriptive |
| 94 | gs_ctcompdatetime_precision<br><br>Show the field ONLY if:<br>[gs_ctcomp]='1' and ([treatment_recvd]='1' or [gs_notadmit]<>'1') | ✪ Brain Imaging Initiated Precision | dropdown<br>5 Date & Time (MM/DD/YYYY HH24:MM)<br>3 Date Only (MM/DD/YYYY)<br>0 Unknown |
| 95 | gs_ctcompdatetime<br><br>Show the field ONLY if:<br>[gs_ctcomp]='1' and [gs_ctcompdatetime_precision]<>'0' and ([treatment_recvd]='1' or [gs_notadmit]<>'1') | ✪ Date/Time Brain Imaging First Initiated at your hospital<br>*If Date Only, enter 00:00 for time.* | text (datetime_mdy, Min: 2010-01-01 00:00, Max: 2099-12-31 23:59), Identifier |

| | | | |
|---|---|---|---|
| 96 | calc_door_to_ctcomp<br><br>Show the field ONLY if:<br>[gs_ctcomp]='1' and [gs_ctcompdatetime_precision]<>'0' and ([treatment_recvd]='1' or [gs_notadmit]<>'1') | Door to CT Initiated<br>*Time in Minutes - Goal: ≤ 25 Minutes* | calc<br>Calculation:<br>if([doortime_ct]='2',datediff([gs_symptomdatetime], [gs_ctcompdatetime], "m", "mdy", true), datediff([jc_arrdatetime], [gs_ctcompdatetime], "m", "mdy", true)) |
| 97 | desc_ctrepdatetime<br><br>Show the field ONLY if:<br>[gs_jp_ctna]<>'1' and ([gs_notadmit]<>'1' or ([gs_ctcomp]<>'2' and [gs_ctcomp]<>'3')) | | descriptive |
| 98 | gs_jp_ctrepdatetime_precision<br><br>Show the field ONLY if:<br>[gs_jp_ctna]<>'1' and ([gs_notadmit]<>'1' or ([gs_ctcomp]<>'2' and [gs_ctcomp]<>'3')) | ✪ Brain Imaging Interpreted Precision | dropdown<br>5 \| Date & Time (MM/DD/YYYY HH24:MM)<br>3 \| Date Only (MM/DD/YYYY)<br>0 \| Unknown |
| 99 | gs_jp_ctrepdatetime<br><br>Show the field ONLY if:<br>[gs_jp_ctna]<>'1' and [gs_jp_ctrepdatetime_precision]<>'0' and ([gs_notadmit]<>'1' or ([gs_ctcomp]<>'2' and [gs_ctcomp]<>'3')) | ✪ Date/Time Brain Imaging Interpreted<br>*If Date Only, enter 00:00 for time.* | text (datetime_mdy, Min: 2010-01-01 00:00, Max: 2099-12-31 23:59), Identifier |
| 100 | br_img_dt_source<br><br>Show the field ONLY if:<br>[gs_jp_ctna]<>'1' and [gs_jp_ctrepdatetime_precision]<>'0' and ([gs_notadmit]<>'1' or ([gs_ctcomp]<>'2' and [gs_ctcomp]<>'3')) | ✪ Interpretation Time Source | radio<br>1 \| Neurologist Note<br>2 \| Radiology Report |
| 101 | calc_door_to_ctrep<br><br>Show the field ONLY if:<br>[gs_jp_ctna]<>'1' and [gs_jp_ctrepdatetime_precision]<>'0' and ([gs_notadmit]<>'1' or ([gs_ctcomp]<>'2' and [gs_ctcomp]<>'3')) | Door to CT Read<br>*Time in Minutes - Goal: ≤ 45 Minutes* | calc<br>Calculation:<br>if([doortime_ct]='2',datediff([gs_symptomdatetime], [gs_jp_ctrepdatetime], "m", "mdy", true),datediff([jc_arrdatetime], [gs_jp_ctrepdatetime], "m", "mdy", true)) |

| | | | |
|---|---|---|---|
| 102 | radiologist_name<br><br>Show the field ONLY if:<br>[gs_jp_ctna]<>'1' and ([treatment_recvd]='1' or ([gs_lastknownwell_precision]<>'0' and [calc_lkw_to_arrival]<=1440 and [gs_patientarrival]<>'3') or [att_fields_override]='1' or [radiologist_name]<>'') and [att_fields_override]<>'0' | ✪ Name of reading Radiologist | text<br>Custom alignment: RH |
| 103 | gs_ivtpaordered_na<br><br>Show the field ONLY if:<br>[treatment_recvd]='1' and (([gs_lastknownwell_precision]<>'0' and [calc_lkw_to_arrival]<=1440 and [gs_patientarrival]<>'3') or [att_fields_override]='1' or [gs_ivtpaordered_na]<>'') and [att_fields_override]<>'0' | Section Header: *Thrombolytic Therapy*<br><br>✪ IV Thrombolytic Orders | radio<br>1 \| N/A |
| 104 | desc_ivtpaordered<br><br>Show the field ONLY if:<br>[gs_ivtpaordered_na]<>'1' and [treatment_recvd]='1' and (([gs_lastknownwell_precision]<>'0' and [calc_lkw_to_arrival]<=1440 and [gs_patientarrival]<>'3') or [att_fields_override]='1' or [gs_ivtpaordered_precision]<>'') and [att_fields_override]<>'0' | | descriptive |
| 105 | gs_ivtpaordered_precision<br><br>Show the field ONLY if:<br>[gs_ivtpaordered_na]<>'1' and [treatment_recvd]='1' and (([gs_lastknownwell_precision]<>'0' and [calc_lkw_to_arrival]<=1440 and [gs_patientarrival]<>'3') or [att_fields_override]='1' or [gs_ivtpaordered_precision]<>'') and [att_fields_override]<>'0' | ✪ IV Thrombolytic Ordered Precision | dropdown<br>5 \| Date & Time (MM/DD/YYYY HH24:MM)<br>3 \| Date Only (MM/DD/YYYY)<br>0 \| Unknown |

| | | | |
|---|---|---|---|
| 106 | gs_ivtpaordered<br><br>Show the field ONLY if: [gs_ivtpaordered_na]<>'1' and [gs_ivtpaordered_precision]<>'0' and [treatment_recvd]='1' and (([gs_lastknownwell_precision]<>'0' and [calc_lkw_to_arrival]<=1440 and [gs_patientarrival]<>'3') or [att_fields_override]='1' or [gs_ivtpaordered]<>'') and [att_fields_override]<>'0' | ✪ Date/Time IV Thrombolytic Ordered<br>*If Date Only, enter 00:00 for time.* | text (datetime_mdy, Min: 2010-01-01 00:00, Max: 2099-12-31 23:59), Identifier |
| 107 | tnk_dose_ordered<br><br>Show the field ONLY if: [iv_thrombolytic_type]='2' | ✪ What dose of Tenecteplase was ordered?<br>*1 Decimal Place Required* | text (number_1dp) |
| 108 | tnk_dose_given<br><br>Show the field ONLY if: [iv_thrombolytic_type]='2' | ✪ What dose of Tenecteplase was given?<br>*1 Decimal Place Required (GWTG)* | text (number_1dp)<br>Custom alignment: RH |
| 109 | flush_before_tnk<br><br>Show the field ONLY if: [iv_thrombolytic_type]='2' | ✪ Was a Normal Saline Flush documented as given immediately before the IV Thrombolytic given? | yesno<br>1 \| Yes<br>0 \| No<br><br>Custom alignment: RH |
| 110 | flush_after_tnk<br><br>Show the field ONLY if: [iv_thrombolytic_type]='2' | ✪ Was a Normal Saline Flush documented as given immediately after the IV Thrombolytic given? | yesno<br>1 \| Yes<br>0 \| No<br><br>Custom alignment: RH |
| 111 | tpa_decision<br><br>Show the field ONLY if: [gs_ivtpaordered_na]<>'1' and [treatment_recvd]='1' and (([gs_lastknownwell_precision]<>'0' and [calc_lkw_to_arrival]<=1440 and [gs_patientarrival]<>'3') or [att_fields_override]='1' or [tpa_decision]<>'') and [att_fields_override]<>'0' | ✪ Thrombolytic Decision to Treat Date/Time | text (datetime_mdy, Min: 2010-01-01 00:00, Max: 2099-12-31 23:59) |
| 112 | prebolus_nvs_dt<br><br>Show the field ONLY if: ([site]='SMCA' or [site]='SMCH' or [site]='SMCW' or [site]='UMCB') and [treatment_type(IVtPA)]='1' | ✪ Date/Time of Pre-Bolus Neuro Assessment | text (datetime_mdy, Min: 2010-01-01 00:00, Max: 2099-12-31 23:59) |

| | | | |
|---|---|---|---|
| 113 | prebolus_vs_dt<br><br>Show the field ONLY if:<br>([site]='SMCA' or [site]='SMCH' or [site]='SMCW' or [site]='UMCB') and [treatment_type(IVtPA)]='1' | ✪ Date/Time of Pre-Bolus Vital Signs | text (datetime_mdy, Min: 2010-01-01 00:00, Max: 2099-12-31 23:59) |
| 114 | prebolus_bp<br><br>Show the field ONLY if:<br>[treatment_type(IVtPA)]='1' | ✪ Was pre-bolus BP below 185/110? | radio<br>1, Yes<br>0, No<br>ND, Not Documented<br><br>Custom alignment: RH |
| 115 | tpa_idc<br><br>Show the field ONLY if:<br>([site]='SMCA' or [site]='SMCH' or [site]='SMCW' or [site]='UMCB') and ([treatment_type(IAtPA)]='1' or [treatment_type(IVtPA)]='1') | ✪ Was IV thrombolytic independent double check performed? | yesno<br>1, Yes<br>0, No<br><br>Custom alignment: RH |
| 116 | desc_ivthrodatetime<br><br>Show the field ONLY if:<br>[treatment_type(IVtPA)]='1' | | descriptive |
| 117 | ivthrodttm_precision<br><br>Show the field ONLY if:<br>[treatment_type(IVtPA)]='1' | ✪ IV Thrombolytic Bolus Initiated Precision | dropdown<br>5, Date & Time (MM/DD/YYYY HH24:MM)<br>3, Date Only (MM/DD/YYYY)<br>0, Unknown |
| 118 | ivthrodttm<br><br>Show the field ONLY if:<br>[treatment_type(IVtPA)]='1' and [ivthrodttm_precision]<>'0' | ✪ Date/Time IV Thrombolytic Bolus Initiated<br>*If Date Only, enter 00:00 for time.* | text (datetime_mdy, Min: 2010-01-01 00:00, Max: 2099-12-31 23:59), Identifier |
| 119 | doortime_tpa<br><br>Show the field ONLY if:<br>[treatment_type(IVtPA)]='1' and [ivthrodttm_precision]<>'0' and [gs_symptomlocation]='4' | ✪ Door Time for Thrombolytic Bolus Administration<br>*Arrival Time used by Default* | radio<br>2, Use Symptom Discovery Time |
| 120 | calc_door_to_ivtpa<br><br>Show the field ONLY if:<br>[treatment_type(IVtPA)]='1' and [ivthrodttm_precision]<>'0' | Door to Needle<br>*Time in Minutes - Goal: ≤ 30 Minutes* | calc<br>Calculation:<br>if([doortime_tpa]='2',datediff([gs_symptomdatetime], [ivthrodttm], "m", "mdy", true), datediff([jc_arrdatetime], [ivthrodttm], "m", "mdy", true)) |

| | | | |
|---|---|---|---|
| 121 | calc_lkw_to_ivtpa<br>Show the field ONLY if:<br>[treatment_type(IVtPA)]='1' and [ivthrodttm_precision]<>'0' | Last Known Well to IV Thrombolytic<br>*Time in Minutes* | calc<br>Calculation: datediff([gs_lastknownwell], [ivthrodttm], "m", "mdy", true) |
| 122 | calc_ordered_to_bolus<br>Show the field ONLY if:<br>[treatment_type(IVtPA)]='1' and [gs_ivtpaordered_na]<>'1' and [ivthrodttm_precision]<>'0' | IV Thrombolytic Ordered to Bolus<br>*Time in Minutes* | calc<br>Calculation: datediff([gs_ivtpaordered], [ivthrodttm], "m", "mdy", true) |
| 123 | calc_decision_to_bolus<br>Show the field ONLY if:<br>[treatment_type(IVtPA)]='1' and [gs_ivtpaordered_na]<>'1' and [ivthrodttm_precision]<>'0' | Thrombolytic Decision to Bolus<br>*Time in Minutes* | calc<br>Calculation: datediff([tpa_decision], [ivthrodttm], "m", "mdy", true) |
| 124 | desc_pre_bolus_warning<br>Show the field ONLY if:<br>datediff([prebolus_nvs_dt], [ivthrodttm], "m", "mdy", true)>15 or datediff([prebolus_vs_dt], [ivthrodttm], "m", "mdy", true)>15 | Warning! Pre-Bolus Neuro Assessment or Vital Signs not within 15 minutes of Bolus Time! | descriptive |
| 125 | calc_nvs_to_bolus<br>Show the field ONLY if:<br>([site]='SMCA' or [site]='SMCH' or [site]='SMCW' or [site]='UMCB') and [treatment_type(IVtPA)]='1' and [ivthrodttm_precision]<>'0' | Pre-Bolus Neuro Assessment to IV Thrombolytic<br>*Time in Minutes* | calc<br>Calculation: datediff([prebolus_nvs_dt], [ivthrodttm], "m", "mdy", true) |
| 126 | calc_vs_to_bolus<br>Show the field ONLY if:<br>([site]='SMCA' or [site]='SMCH' or [site]='SMCW' or [site]='UMCB') and [treatment_type(IVtPA)]='1' and [ivthrodttm_precision]<>'0' | Pre-Bolus Vital Signs to IV Thrombolytic<br>*Time in Minutes* | calc<br>Calculation: datediff([prebolus_vs_dt], [ivthrodttm], "m", "mdy", true) |
| 127 | infusionstartdt<br>Show the field ONLY if:<br>[iv_thrombolytic_type]='1' and [ivthrodttm_precision]<>'0' | ✪ IV Thrombolytic Infusion Start Date/Time<br>*If Date Only, enter 00:00 for time.* | text (datetime_mdy, Min: 2010-01-01 00:00, Max: 2099-12-31 23:59), Identifier |
| 128 | calc_tpa_bolus_to_infusion<br>Show the field ONLY if:<br>[iv_thrombolytic_type]='1' and [ivthrodttm_precision]<>'0' | IV Thrombolytic Bolus to Infusion<br>*Time in Minutes* | calc<br>Calculation: datediff([ivthrodttm], [infusionstartdt], "m", "mdy", true) |

| | | | |
|---|---|---|---|
| 129 | flush_after_tpa<br><br>Show the field ONLY if:<br>([site]='SMCA' or [site]='SMCH' or [site]='SMCW' or [site]='UMCB') and ([iv_thrombolytic_type]='1' or [gs_ptrecvdreason(2)]='1' or [gs_ivtpaoutside]='1') | ✪ Was 50 cc Normal Saline Flush documented as given after infusion of Alteplase?(If infusion was completed at this hospital.) | radio<br>1 — Yes<br>0 — No<br>NA — NA / tPA Stopped<br><br>Custom alignment: RH<br>Field Annotation: @HIDDEN |
| 130 | gs_jp_labna<br><br>Show the field ONLY if:<br>([treatment_recvd]='1' or ([gs_lastknownwell_precision]<>'0' and [calc_lkw_to_arrival]<=1440 and [gs_patientarrival]<>'3') or [att_fields_override]='1' or [gs_jp_labna]<>'') and [att_fields_override]<>'0' | Section Header: *Labs*<br><br>✪ Labs | radio<br>1 — N/A |
| 131 | desc_laborddatetime<br><br>Show the field ONLY if:<br>[gs_jp_labna]<>'1' and ([treatment_recvd]='1' or ([gs_lastknownwell_precision]<>'0' and [calc_lkw_to_arrival]<=1440 and [gs_patientarrival]<>'3') or [att_fields_override]='1' or [gs_jp_laborddatetime_precision]<>'') and [att_fields_override]<>'0' | | descriptive |
| 132 | gs_jp_laborddatetime_precision<br><br>Show the field ONLY if:<br>[gs_jp_labna]<>'1' and ([treatment_recvd]='1' or ([gs_lastknownwell_precision]<>'0' and [calc_lkw_to_arrival]<=1440 and [gs_patientarrival]<>'3') or [att_fields_override]='1' or [gs_jp_laborddatetime_precision]<>'') and [att_fields_override]<>'0' | ✪ Lab Tests Ordered Precision | dropdown<br>5 — Date & Time (MM/DD/YYYY HH24:MM)<br>3 — Date Only (MM/DD/YYYY)<br>0 — Unknown |

| | | | |
|---|---|---|---|
| 133 | gs_jp_laborddatetime<br><br>Show the field ONLY if:<br>[gs_jp_labna]<>'1' and [gs_jp_laborddatetime_precision]<>'0' and ([treatment_recvd]='1' or ([gs_lastknownwell_precision]<>'0' and [calc_lkw_to_arrival]<=1440 and [gs_patientarrival]<>'3') or [att_fields_override]='1' or [gs_jp_laborddatetime]<>'') and [att_fields_override]<>'0' | ✪ Date/Time Lab Tests Ordered<br>*If Date Only, enter 00:00 for time.* | text (datetime_mdy, Min: 2010-01-01 00:00, Max: 2099-12-31 23:59), Identifier |
| 134 | gs_jp_labcpdatetime_precision<br><br>Show the field ONLY if:<br>[gs_jp_labna]<>'1' and ([treatment_recvd]='1' or ([gs_lastknownwell_precision]<>'0' and [calc_lkw_to_arrival]<=1440 and [gs_patientarrival]<>'3') or [att_fields_override]='1' or [gs_jp_labcpdatetime_precision]<>'') and [att_fields_override]<>'0' | ✪ Lab Tests Completed Precision | dropdown<br>5 \| Date & Time (MM/DD/YYYY HH24:MM)<br>3 \| Date Only (MM/DD/YYYY)<br>0 \| Unknown |
| 135 | gs_jp_labcpdatetime<br><br>Show the field ONLY if:<br>[gs_jp_labna]<>'1' and [gs_jp_labcpdatetime_precision]<>'0' and ([treatment_recvd]='1' or ([gs_lastknownwell_precision]<>'0' and [calc_lkw_to_arrival]<=1440 and [gs_patientarrival]<>'3') or [att_fields_override]='1' or [gs_jp_labcpdatetime]<>'') and [att_fields_override]<>'0' | ✪ Date/Time Lab Tests Completed<br>*If Date Only, enter 00:00 for time.Use PT/INR* | text (datetime_mdy, Min: 2010-01-01 00:00, Max: 2099-12-31 23:59), Identifier |
| 136 | doortime_labs<br><br>Show the field ONLY if:<br>[gs_jp_labna]<>'1' and [gs_jp_labcpdatetime_precision]<>'0' and ([treatment_recvd]='1' or ([gs_lastknownwell_precision]<>'0' and [calc_lkw_to_arrival]<=1440 and [gs_patientarrival]<>'3') or [att_fields_override]='1' or [doortime_labs]<>'') and [att_fields_override]<>'0' and [gs_symptomlocation]='4' | ✪ Door Time for Labs<br>*Arrival Time used by Default* | radio<br>2 \| Use Symptom Discovery Time |

| | | | |
|---|---|---|---|
| 137 | calc_order_to_labcp<br><br>Show the field ONLY if: [gs_jp_labna]<>'1' and [gs_jp_labcpdatetime_precision]<>'0' and ([treatment_recvd]='1' or ([gs_lastknownwell_precision]<>'0' and [calc_lkw_to_arrival]<=1440 and [gs_patientarrival]<>'3') or [att_fields_override]='1' or [calc_order_to_labcp]<>'') and [att_fields_override]<>'0' | Ordered to Labs Completed<br>*Time in Minutes - Goal: ≤ 45 Minutes* | calc<br>Calculation: datediff([gs_jp_laborddatetime], [gs_jp_labcpdatetime], "m", "mdy", true) |
| 138 | calc_door_to_labcp<br><br>Show the field ONLY if: [gs_jp_labna]<>'1' and [gs_jp_labcpdatetime_precision]<>'0' and ([treatment_recvd]='1' or ([gs_lastknownwell_precision]<>'0' and [calc_lkw_to_arrival]<=1440 and [gs_patientarrival]<>'3') or [att_fields_override]='1' or [calc_door_to_labcp]<>'') and [att_fields_override]<>'0' | Door to Labs Completed<br>*Time in Minutes - Goal: ≤ 45 Minutes* | calc<br>Calculation: if([doortime_labs]='2',datediff([gs_symptomdatetime], [gs_jp_labcpdatetime], "m", "mdy", true), datediff([jc_arrdatetime], [gs_jp_labcpdatetime], "m", "mdy", true)) |
| 139 | gs_jp_ecgna<br><br>Show the field ONLY if: ([treatment_recvd]='1' or ([gs_lastknownwell_precision]<>'0' and [calc_lkw_to_arrival]<=1440 and [gs_patientarrival]<>'3') or [att_fields_override]='1' or [gs_jp_ecgna]<>'') and [att_fields_override]<>'0' | Section Header: *ECG*<br><br>✪ ECG | radio<br>1 N/A |
| 140 | desc_ecgorddatetime<br><br>Show the field ONLY if: [gs_jp_ecgna]<>'1' and ([treatment_recvd]='1' or ([gs_lastknownwell_precision]<>'0' and [calc_lkw_to_arrival]<=1440 and [gs_patientarrival]<>'3') or [att_fields_override]='1' or [gs_jp_ecgorddatetime_precision]<>'') and [att_fields_override]<>'0' | | descriptive |

| | | | |
|---|---|---|---|
| 141 | gs_jp_ecgorddatetime_precision<br><br>Show the field ONLY if:<br>[gs_jp_ecgna]<>'1' and ([treatment_recvd]='1' or ([gs_lastknownwell_precision]<>'0' and [calc_lkw_to_arrival]<=1440 and [gs_patientarrival]<>'3') or [att_fields_override]='1' or [gs_jp_ecgorddatetime_precision]<>'') and [att_fields_override]<>'0' | ✪ ECG Ordered Precision | dropdown<br>5 \| Date & Time (MM/DD/YYYY HH24:MM)<br>3 \| Date Only (MM/DD/YYYY)<br>0 \| Unknown |
| 142 | gs_jp_ecgorddatetime<br><br>Show the field ONLY if:<br>[gs_jp_ecgna]<>'1' and [gs_jp_ecgorddatetime_precision]<>'0' and ([treatment_recvd]='1' or ([gs_lastknownwell_precision]<>'0' and [calc_lkw_to_arrival]<=1440 and [gs_patientarrival]<>'3') or [att_fields_override]='1' or [gs_jp_ecgorddatetime]<>'') and [att_fields_override]<>'0' | ✪ Date/Time ECG Ordered<br>*If Date Only, enter 00:00 for time.* | text (datetime_mdy, Min: 2010-01-01 00:00, Max: 2099-12-31 23:59), Identifier |
| 143 | gs_jp_ecgcpdatetime_precision<br><br>Show the field ONLY if:<br>[gs_jp_ecgna]<>'1' and ([treatment_recvd]='1' or ([gs_lastknownwell_precision]<>'0' and [calc_lkw_to_arrival]<=1440 and [gs_patientarrival]<>'3') or [att_fields_override]='1' or [gs_jp_ecgcpdatetime_precision]<>'') and [att_fields_override]<>'0' | ✪ ECG Completed Precision | dropdown<br>5 \| Date & Time (MM/DD/YYYY HH24:MM)<br>3 \| Date Only (MM/DD/YYYY)<br>0 \| Unknown |
| 144 | gs_jp_ecgcpdatetime<br><br>Show the field ONLY if:<br>[gs_jp_ecgna]<>'1' and [gs_jp_ecgcpdatetime_precision]<>'0' and ([treatment_recvd]='1' or ([gs_lastknownwell_precision]<>'0' and [calc_lkw_to_arrival]<=1440 and [gs_patientarrival]<>'3') or [att_fields_override]='1' or [gs_jp_ecgcpdatetime]<>'') and [att_fields_override]<>'0' | ✪ Date/Time ECG Completed<br>*If Date Only, enter 00:00 for time.* | text (datetime_mdy, Min: 2010-01-01 00:00, Max: 2099-12-31 23:59), Identifier |

| | | | |
|---|---|---|---|
| 145 | doortime_ecg<br><br>Show the field ONLY if:<br>[gs_jp_ecgna]<>'1' and [gs_jp_ecgcpdatetime_precision]<>'0' and ([treatment_recvd]='1' or ([gs_lastknownwell_precision]<>'0' and [calc_lkw_to_arrival]<=1440 and [gs_patientarrival]<>'3') or [att_fields_override]='1' or [doortime_ecg]<>'') and [att_fields_override]<>'0' and [gs_symptomlocation]='4' | ✪ Door Time for ECG<br>*Arrival Time used by Default* | radio<br>2 Use Symptom Discovery Time |
| 146 | calc_door_to_ecgcp<br><br>Show the field ONLY if:<br>[gs_jp_ecgna]<>'1' and [gs_jp_ecgcpdatetime_precision]<>'0' and ([treatment_recvd]='1' or ([gs_lastknownwell_precision]<>'0' and [calc_lkw_to_arrival]<=1440 and [gs_patientarrival]<>'3') or [att_fields_override]='1' or [calc_door_to_ecgcp]<>'') and [att_fields_override]<>'0' | Door to ECG Completed<br>*Time in Minutes - Goal: ≤ 45 Minutes* | calc<br>Calculation:<br>if([doortime_ecg]='2',datediff([gs_symptomdatetime], [gs_jp_ecgcpdatetime], "m", "mdy", true), datediff([jc_arrdatetime], [gs_jp_ecgcpdatetime], "m", "mdy", true)) |
| 147 | gs_jp_xrayna<br><br>Show the field ONLY if:<br>([treatment_recvd]='1' or ([gs_lastknownwell_precision]<>'0' and [calc_lkw_to_arrival]<=1440 and [gs_patientarrival]<>'3') or [att_fields_override]='1' or [gs_jp_xrayna]<>'') and [att_fields_override]<>'0' | Section Header: *Chest X-ray*<br><br>✪ Chest X-ray | radio<br>1 N/A |
| 148 | desc_xorddatetime<br><br>Show the field ONLY if:<br>[gs_jp_xrayna]<>'1' and ([treatment_recvd]='1' or ([gs_lastknownwell_precision]<>'0' and [calc_lkw_to_arrival]<=1440 and [gs_patientarrival]<>'3') or [att_fields_override]='1' or [gs_jp_xorderdatetime_precision]<>'') and [att_fields_override]<>'0' | | descriptive |

| | | | |
|---|---|---|---|
| 149 | gs_jp_xorderdatetime_precision<br><br>Show the field ONLY if:<br>[gs_jp_xrayna]<>'1' and ([treatment_recvd]='1' or ([gs_lastknownwell_precision]<>'0' and [calc_lkw_to_arrival]<=1440 and [gs_patientarrival]<>'3') or [att_fields_override]='1' or [gs_jp_xorderdatetime_precision]<>'') and [att_fields_override]<>'0' | ✪ Chest X-ray Ordered Precision | dropdown<br>5 \| Date & Time (MM/DD/YYYY HH24:MM)<br>3 \| Date Only (MM/DD/YYYY)<br>0 \| Unknown |
| 150 | gs_jp_xorderdatetime<br><br>Show the field ONLY if:<br>[gs_jp_xrayna]<>'1' and [gs_jp_xorderdatetime_precision]<>'0' and ([treatment_recvd]='1' or ([gs_lastknownwell_precision]<>'0' and [calc_lkw_to_arrival]<=1440 and [gs_patientarrival]<>'3') or [att_fields_override]='1' or [gs_jp_xorderdatetime]<>'') and [att_fields_override]<>'0' | ✪ Date/Time Chest X-ray Ordered<br>*If Date Only, enter 00:00 for time.* | text (datetime_mdy, Min: 2010-01-01 00:00, Max: 2099-12-31 23:59), Identifier |
| 151 | gs_jp_xcompdatetime_precision<br><br>Show the field ONLY if:<br>[gs_jp_xrayna]<>'1' and ([treatment_recvd]='1' or ([gs_lastknownwell_precision]<>'0' and [calc_lkw_to_arrival]<=1440 and [gs_patientarrival]<>'3') or [att_fields_override]='1' or [gs_jp_xcompdatetime_precision]<>'') and [att_fields_override]<>'0' | ✪ Chest X-ray Completed Precision | dropdown<br>5 \| Date & Time (MM/DD/YYYY HH24:MM)<br>3 \| Date Only (MM/DD/YYYY)<br>0 \| Unknown |
| 152 | gs_jp_xcompdatetime<br><br>Show the field ONLY if:<br>[gs_jp_xrayna]<>'1' and [gs_jp_xcompdatetime_precision]<>'0' and ([treatment_recvd]='1' or ([gs_lastknownwell_precision]<>'0' and [calc_lkw_to_arrival]<=1440 and [gs_patientarrival]<>'3') or [att_fields_override]='1' or [gs_jp_xcompdatetime]<>'') and [att_fields_override]<>'0' | ✪ Date/Time Chest X-ray Completed<br>*If Date Only, enter 00:00 for time.* | text (datetime_mdy, Min: 2010-01-01 00:00, Max: 2099-12-31 23:59), Identifier |

| | | | |
|---|---|---|---|
| 153 | nir_notified_dt<br><br>Show the field ONLY if:<br>[treatment_type(MER)]='1' | Section Header: *Skin Puncture (Endovascular Access)*<br>✪ Time Neuro Interventionalist Notified | text (datetime_mdy, Min: 2010-01-01 00:00, Max: 2099-12-31 23:59), Identifier |
| 154 | mer_decision<br><br>Show the field ONLY if:<br>([treatment_type(MER)]='1' or [treatment_type(IAtPA)]='1') and ([att_fields_override]<>'0' or [mer_decision]<>'') | ✪ Endovascular Decision to Treat Date/Time | text (datetime_mdy, Min: 2010-01-01 00:00, Max: 2099-12-31 23:59) |
| 155 | desc_artpuncdt<br><br>Show the field ONLY if:<br>[treatment_type(MER)]='1' or [treatment_type(IAtPA)]='1' | | descriptive |
| 156 | artpunc_dt_precision<br><br>Show the field ONLY if:<br>[treatment_type(MER)]='1' or [treatment_type(IAtPA)]='1' | ✪ Skin Puncture Date/Time Precision<br>Formerly: Arterial Puncture Date/Time Precision | dropdown<br>5 \| Date & Time (MM/DD/YYYY HH24:MM)<br>3 \| Date Only (MM/DD/YYYY)<br>0 \| Unknown |
| 157 | artpunc_dt<br><br>Show the field ONLY if:<br>([treatment_type(MER)]='1' or [treatment_type(IAtPA)]='1') and [artpunc_dt_precision]<>'0' | ✪ Skin Puncture Date/Time<br>Formerly: Arterial Puncture Date/Time<br>*If Date Only, enter 00:00 for time.* | text (datetime_mdy, Min: 2010-01-01 00:00, Max: 2099-12-31 23:59), Identifier |
| 158 | decision_evt<br><br>Show the field ONLY if:<br>([treatment_type(MER)]='1' or [treatment_type(IAtPA)]='1') and [artpunc_dt_precision]<>'0' and [gs_symptomlocation]<>'4' | ✪ Decision for Endovascular Treatment based on recurring or worsening symptoms? | yesno<br>1 \| Yes<br>0 \| No<br><br>Custom alignment: RH<br>Field Annotation: @DEFAULT=0 |
| 159 | worse_sympt_dt<br><br>Show the field ONLY if:<br>[decision_evt]=1 | ✪ Discovery of worsening or recurring symptoms | text (datetime_mdy, Min: 2010-01-01 00:00, Max: 2099-12-31 23:59)<br>Custom alignment: RH |
| 160 | doortime_evt<br><br>Show the field ONLY if:<br>([treatment_type(MER)]='1' or [treatment_type(IAtPA)]='1') and [artpunc_dt_precision]<>'0' and [gs_symptomlocation]='4' | ✪ Door Time for Endovascular Treatment<br>*Arrival Time used by Default* | radio<br>2 \| Use Symptom Discovery Time |

| | | | |
|---|---|---|---|
| 161 | calc_door_to_artpunc<br><br>Show the field ONLY if:<br>([treatment_type(MER)]='1' or [treatment_type(IAtPA)]='1') and [artpunc_dt_precision]<>'0' | Door to Groin<br>*Time in Minutes - Goal: ≤ 90 Minutes* | calc<br>Calculation: if([doortime_evt]='2', datediff([gs_symptomdatetime], [artpunc_dt], "m", "mdy", true), if([decision_evt]='1', datediff([worse_sympt_dt], [artpunc_dt], "m", "mdy", true), datediff([jc_arrdatetime], [artpunc_dt], "m", "mdy", true))) |
| 162 | calc_lkw_to_artpunc<br><br>Show the field ONLY if:<br>([treatment_type(MER)]='1' or [treatment_type(IAtPA)]='1') and [artpunc_dt_precision]<>'0' | LKW to Groin<br>*Time in Minutes* | calc<br>Calculation: datediff([gs_lastknownwell], [artpunc_dt], "m", "mdy", true) |
| 163 | desc_firstpassdatetime<br><br>Show the field ONLY if:<br>[treatment_type(MER)]='1' | | descriptive |
| 164 | firstpassdt_precision<br><br>Show the field ONLY if:<br>[treatment_type(MER)]='1' | ✪ First Pass Date/Time Precision | dropdown<br>5 Date & Time (MM/DD/YYYY HH24:MM)<br>3 Date Only (MM/DD/YYYY)<br>0 Unknown<br><br>Custom alignment: RH |
| 165 | firstpassdt<br><br>Show the field ONLY if:<br>[treatment_type(MER)]='1' and [firstpassdt_precision]<>'0' | ✪ What is the date and time of the first pass of a clot retrieval device at this hospital? | text (datetime_mdy, Min: 2010-01-01 00:00, Max: 2099-12-31 23:59), Identifier<br>Custom alignment: RH |
| 166 | calc_door_to_device<br><br>Show the field ONLY if:<br>[treatment_type(MER)]='1' and [firstpassdt_precision]<>'0' | Arrival to Device<br>*Time in Minutes* | calc<br>Calculation: if([doortime_evt]='2', datediff([gs_symptomdatetime], [firstpassdt], "m", "mdy", true), if([decision_evt]='1', datediff([worse_sympt_dt], [firstpassdt], "m", "mdy", true), datediff([jc_arrdatetime], [firstpassdt], "m", "mdy", true))) |
| 167 | calc_puncture_to_device<br><br>Show the field ONLY if:<br>[treatment_type(MER)]='1' and [firstpassdt_precision]<>'0' | Puncture to Device<br>*Time in Minutes* | calc<br>Calculation: datediff([artpunc_dt], [firstpassdt], "m", "mdy", true) |
| 168 | desc_ticidatetime<br><br>Show the field ONLY if:<br>([treatment_type(MER)]='1' or [treatment_type(IAtPA)]='1') | | descriptive |

| | | | |
|---|---|---|---|
| 169 | ticidt_precision<br><br>Show the field ONLY if:<br>([treatment_type(MER)]='1' or [treatment_type(IAtPA)]='1') | ✪ Date/time TICI grade 2B or 3 Precision | dropdown<br>5 – Date & Time (MM/DD/YYYY HH24:MM)<br>3 – Date Only (MM/DD/YYYY)<br>0 – Unknown<br><br>Custom alignment: RH |
| 170 | ticidt<br><br>Show the field ONLY if:<br>([treatment_type(MER)]='1' or [treatment_type(IAtPA)]='1') and [ticidt_precision]<>'0' | ✪ What was the date and time that a TICI 2B/3 was first documented during the mechanical thrombectomy procedure? | text (datetime_mdy, Min: 2015-01-01 00:00, Max: 2099-12-31 23:59), Identifier<br>Custom alignment: RH |
| 171 | calc_door_to_reperfusion<br><br>Show the field ONLY if:<br>([treatment_type(MER)]='1' or [treatment_type(IAtPA)]='1') and [ticidt_precision]<>'0' | Arrival to Reperfusion<br>*Time in Minutes* | calc<br>Calculation: if([doortime_evt]='2', datediff([gs_symptomdatetime], [ticidt], "m", "mdy", true), if([decision_evt]='1', datediff([worse_sympt_dt], [ticidt], "m", "mdy", true), datediff([jc_arrdatetime], [ticidt], "m", "mdy", true))) |
| 172 | calc_puncture_to_reperfusion<br><br>Show the field ONLY if:<br>([treatment_type(MER)]='1' or [treatment_type(IAtPA)]='1') and [ticidt_precision]<>'0' | Puncture to Reperfusion<br>*Time in Minutes* | calc<br>Calculation: datediff([artpunc_dt], [ticidt], "m", "mdy", true) |
| 173 | monitored_bed<br><br>Show the field ONLY if:<br>[gs_notadmit]='2' and [gs_symptomlocation]<>'4' | Section Header: *Monitored Bed*<br><br>✪ At what time was the patient admitted to a Monitored Bed? | text (datetime_mdy, Min: 2010-01-01 00:00, Max: 2099-12-31 23:59), Identifier |
| 174 | calc_door_to_mb<br><br>Show the field ONLY if:<br>[gs_notadmit]='2' and [gs_symptomlocation]<>'4' | Door to Monitored Bed<br>*Time in Minutes - Goal: ≤ 180 Minutes* | calc<br>Calculation: if([gs_symptomlocation]<>4, datediff([jc_arrdatetime], [monitored_bed], "m", "mdy", true),'') |
| 175 | gs_jp_jcahocomm<br><br>Show the field ONLY if:<br>([treatment_recvd]='1' or ([gs_lastknownwell_precision]<>'0' and [calc_lkw_to_arrival]<=1440 and [gs_patientarrival]<>'3') or [att_fields_override]='1' or [gs_jp_jcahocomm]<>'') and [att_fields_override]<>'0' | Section Header: *Follow-up notes*<br><br>✪ Additional comments | notes<br>Field Annotation: @CHARLIMIT=500 |

| | | | |
|---|---|---|---|
| 176 | treatment_delays<br><br>Show the field ONLY if:<br>([treatment_recvd]='1' or [gs_notadmit]<>'1') | Section Header: *Treatment Delays*<br><br>✪ If there were delays in treatment, what were the reasons documented for the delay? | notes<br>Custom alignment: RH |
| 177 | initial_stroke_information_complete | Section Header: *Form Status*<br><br>Complete? | dropdown<br>0 Incomplete<br>1 Unverified<br>2 Complete |
| Instrument: **Admin** (admin) | | | Collapse |
| 178 | desc_transferred_link<br><br>Show the field ONLY if:<br>[transferred_fin]<>'' | Section Header: *Initial Stroke Information:SITE:[site] MRN: [mrn]Patient Name: [jc_lastname], [jc_firstname]Link to Issues Log:Click here to open Clinical Quality Data Team Issues Log record for this patient*<br><br>Click here to open or create record for linked FIN (transferred patient). | descriptive |
| 179 | patient_os_id<br><br>Show the field ONLY if:<br>([site]='SMCA' or [site]='SMCH' or [site]='SMCW' or [site]='UMCB') and ([treatment_recvd]='1' or [gs_notadmit]<>'1') | ✪ Patient ID<br>*GWTG PMT Number Max. 9 chars.* | text<br>Field Annotation: @CHARLIMIT=7 |
| 180 | gs_stroketype | ✪ Final clinical diagnosis related to stroke: | dropdown<br>2 Ischemic stroke<br>3 Transient Ischemic Attack (< 24 hours)<br>4 Subarachnoid Hemorrhage<br>5 Intracerebral Hemorrhage<br>6 Stroke not otherwise specified<br>7 No stroke related diagnosis<br>8 Elective Carotid Intervention only |

| | | | |
|---|---|---|---|
| 181 | gs_mimics<br><br>Show the field ONLY if:<br>([site]='SMCA' or [site]='SMCH' or [site]='SMCW' or [site]='UMCB') and [gs_stroketype] = '7' and ([treatment_recvd]='1' or [gs_notadmit]<>'1') and [hide_nonessential_fields(1)]='0' | ✪ If No Stroke Related Diagnosis | dropdown<br>1 Migraine<br>2 Seizure<br>3 Delirium<br>4 Electrolyte or metabolic imbalance<br>5 Functional disorder<br>6 Other<br>7 Uncertain |
| 182 | cryp_etiology<br><br>Show the field ONLY if:<br>([site]='SMCA' or [site]='SMCH' or [site]='SMCW' or [site]='UMCB') and [gs_stroketype] = '2' and ([treatment_recvd]='1' or [gs_notadmit]<>'1') | Was the stroke etiology documented in the patient medical record: | radio<br>1 Yes<br>2 No<br><br>Custom alignment: RH |
| 183 | cryp_stetio<br><br>Show the field ONLY if:<br>([site]='SMCA' or [site]='SMCH' or [site]='SMCW' or [site]='UMCB') and [cryp_etiology] = '1' | Select documented stroke etiology: | radio<br>1 1: Large-artery atherosclerosis (e.g., carotid or basilar artery stenosis)<br>2 2: Cardioembolism (e.g., atrial fibrillation/flutter, prosthetic heart valve, recent MI)<br>3 3: Small-vessel occlusion (e.g., Subcortical or brain stem lacunar infarction < 1.5 cm)<br>4 4: Stroke of other determined etiology<br>5 5: Cryptogenic Stroke |
| 184 | cryp_othetio<br><br>Show the field ONLY if:<br>([site]='SMCA' or [site]='SMCH' or [site]='SMCW' or [site]='UMCB') and [cryp_stetio] = '4' | Stroke of other determined etiology (if selected, then specify) | radio<br>1 Dissection<br>2 Hypercoagualability<br>3 Other (e.g., vasculopathy or other hematologic disorders) |
| 185 | cryp_crystk<br><br>Show the field ONLY if:<br>([site]='SMCA' or [site]='SMCH' or [site]='SMCW' or [site]='UMCB') and [cryp_stetio] = '5' | Cryptogenic Stroke (if selected, then specify) | radio<br>1 Multiple potential etiologies identified<br>2 Stroke of undetermined etiology<br>3 Unspecified |

| | | | |
|---|---|---|---|
| 186 | dnv_5_statement<br><br>Show the field ONLY if:<br>([site]='SMCA' or [site]='SMCH' or [site]='SMCW' or [site]='UMCB') and [gs_stroketype]='2' and [calc_lkw_to_arrival]>='0' and [calc_lkw_to_arrival]<=1440 and [hide_nonessential_fields(1)]='0' | Was a Thrombectomy Statement Present?<br>(Documentation that an endovascular procedure either was performed or was considered and deemed not to be appropriate or possible for ischemic stroke patients arriving within 24 hours of last known well.) | yesno<br>1 Yes<br>0 No<br><br>Custom alignment: RH<br>Field Annotation: @HIDDEN |
| 187 | gs_comfortonly<br><br>Show the field ONLY if:<br>([treatment_recvd]='1' or [gs_notadmit]<>'1') | When is the earliest documentation of comfort measures only? | dropdown<br>1 Day 0 or 1<br>2 Day 2 or after<br>3 Timing unclear<br>4 Not Documented/UTD |
| 188 | ich_documentation<br><br>Show the field ONLY if:<br>[site]='UMCB' and ([gs_stroketype]='5' or [gs_comfortonly]='1') | ✪ Is there clear documentation for comfort care/palliative care measures established prior to hospital arrival? | radio<br>1 Yes<br>2 Not Documented / UTD<br><br>Custom alignment: RH |
| 189 | desc_arrivaldatetime | Arrival Date/Time:[jc_arrdatetime]From: Initial Stroke Information | descriptive |
| 190 | desc_gs_notadmit | Was the patient admitted?[gs_notadmit]From: Initial Stroke Information | descriptive |
| 191 | jc_admitdate<br><br>Show the field ONLY if:<br>[gs_notadmit] = '2' and [hide_nonessential_fields(1)]='0' | Admit Date | text (date_mdy, Min: 2010-01-01, Max: 2099-12-31), Identifier |
| 192 | desc_gs_notadmitwhy<br><br>Show the field ONLY if:<br>[gs_notadmit] = '1' | Reason Not Admitted[gs_notadmitwhy]From: Initial Stroke Information | descriptive |
| 193 | gs_discdatetime_precision | ✪ Discharge Date Precision | dropdown<br>5 Date & Time (MM/DD/YYYY HH24:MM)<br>3 Date Only (MM/DD/YYYY) |
| 194 | gs_discdatetime | ✪ Discharge Date and Time | text (datetime_mdy, Min: 2010-01-01 00:00, Max: 2099-12-31 23:59), Identifier |

| | | | |
|---|---|---|---|
| 195 | gs_delintra<br><br>Show the field ONLY if:<br>([gs_ptarrtranrsn(1)]='1' or [gs_ptarrtranrsn(2)]='1' or [gs_ptarrtranrsn(3)]='1') and [calc_door_to_transfer]>=90 | ✪ Documented reason for delay in transfer to referral facility? | radio<br>1 Yes<br>2 No/ND<br><br>Custom alignment: RH |
| 196 | gs_rsdeltra<br><br>Show the field ONLY if:<br>[gs_delintra] = '1' | ✪ Specific reason for delay documented in transfer patient | checkbox<br>1 gs_rsdeltra___1 Social/religious<br>2 gs_rsdeltra___2 Initial refusal<br>3 gs_rsdeltra___3 Care team unable to determine eligibility<br>4 gs_rsdeltra___4 Management of concomitant emergent/acute conditions such as cardiopulmonary arrest, respiratory failure (requiring intubation)<br>5 gs_rsdeltra___5 Investigational or experimental protocol for reperfusion<br>6 gs_rsdeltra___6 Delay in stroke diagnosis *<br>7 gs_rsdeltra___7 In-hospital time delay *<br>8 gs_rsdeltra___8 Equipment-related delay *<br>9 gs_rsdeltra___9 Need for additional imaging *<br>10 gs_rsdeltra___10 Catheter lab not available *<br>11 gs_rsdeltra___11 Other * |
| 197 | jc_disposition | For patients discharged on or after 04/01/2011:<br>✪ What was the patient's discharge disposition on the day of discharge? | dropdown<br>1 1 - Home<br>2 2 - Hospice - Home<br>3 3 - Hospice - Health Care Facility<br>4 4 - Acute Care Facility<br>5 5 - Other Health Care Facility<br>6 6 - Expired<br>7 7 - Left Against Medical Advise/AMA<br>8 8 - Not Documented or Unable to Determine (UTD) |

| | | | |
|---|---|---|---|
| 198 | gs_othfacility<br><br>Show the field ONLY if:<br>[jc_disposition] = '5' | ✪ If Other Health Care Facility | dropdown<br>1 \| 1 - Skilled Nursing Facility (SNF)<br>2 \| 2 - Inpatient Rehabilitation Facility (IRF)<br>3 \| 3 - Long Term Care Hospital (LTCH)<br>4 \| 4 - Intermediate Care facility (ICF)<br>5 \| Other |
| 199 | calc_door_to_transfer<br><br>Show the field ONLY if:<br>[gs_notadmitwhy]='1' | Door to Transfer<br>*Time in Minutes - Goal: ≤ 120 Minutes* | calc<br>Calculation: if([gs_notadmitwhy]=1, datediff([jc_arrdatetime], [gs_discdatetime], "m", "mdy", true), '') |
| 200 | gwtg_status<br><br>Show the field ONLY if:<br>(([site] = 'SMCA' or [site] = 'SMCH' or [site] = 'SMCW' or [site] = 'UMCB') and [patient_os_id] <> '' and [initial_stroke_information_complete] = '2' and [admin_complete] = '2' and [clinical_codes_complete] = '2' and [admission_complete] = '2' and [hospitalization_complete] = '2' and [advanced_stroke_care_complete] = '2' and [additional_stroke_information_complete] = '2' and [optional_complete] = '2' and [discharge_complete] = '2') or ([gwtg_status(1)]='1') | Section Header: *Status Fields*<br><br>GWTG Upload Status<br>*Uncheck ONLY if data has changed and needs to be uploaded to GWTG.* | checkbox<br>1 \| gwtg_status___1 \| Record uploaded to GWTG. |
| 201 | premier_status<br><br>Show the field ONLY if:<br>[gs_notadmit] = '2' and [initial_stroke_information_complete] = '2' and [admin_complete] = '2' and [clinical_codes_complete] = '2' and [admission_complete] = '2' and [hospitalization_complete] = '2' and [additional_stroke_information_complete] = '2' and [optional_complete] = '2' and [special_initiatives_complete] = '2' and [discharge_complete] = '2' | Premier Upload Status<br>*All changes must be made in PremierDo NOT clear radio button.* | radio<br>1 \| Record Uploaded to Premier |

| | | | |
|---|---|---|---|
| 202 | admin_complete | Section Header: *Form Status*<br>Complete? | dropdown<br>0 Incomplete<br>1 Unverified<br>2 Complete |
| Instrument: **Clinical Codes** (clinical_codes) | | | Collapse |
| 203 | jc_princicd10diagnosis | Section Header: *Initial Stroke Information:SITE:[site] MRN: [mrn]Patient Name: [jc_lastname], [jc_firstname]Link to Issues Log:Click here to open Clinical Quality Data Team Issues Log record for this patient Diagnosis Codes*<br>✪ ICD-10-CM Principal Diagnosis Code<br>*3-7 char. [NO DECIMALS OR DOTS]Discharge Diagnosis Code from Code Stroke Form: [cs_dsch_dx]* | text |
| 204 | jc_othericd10diag<br>Show the field ONLY if:<br>([site]='SMCA' or [site]='SMCH' or [site]='SMCW' or [site]='UMCB') and [jc_princicd10diagnosis] <> '' and ([treatment_recvd]='1' or [gs_notadmit]<>'1') and [hide_nonessential_fields(1)]='0' | ✪ ICD-10-CM Other Diagnosis Codes<br>*Multiple diagnoses can be entered, separated by a colon (:)* | text |
| 205 | jc_princicd10procedurdt<br>Show the field ONLY if:<br>([site]='SMCA' or [site]='SMCH' or [site]='SMCW' or [site]='UMCB') and ([treatment_recvd]='1' or [gs_notadmit]<>'1') and [hide_nonessential_fields(1)]='0' | Section Header: *Procedure Codes*<br>✪ ICD-10-PCS Principal Procedure Date | text (datetime_mdy, Min: 2010-01-01 00:00, Max: 2099-12-31 23:59), Identifier |
| 206 | jc_princicd10procedure<br>Show the field ONLY if:<br>[jc_princicd10procedurdt] <> '' and [hide_nonessential_fields(1)]='0' | ✪ ICD-10-PCS Principal Procedure Code | text<br>Field Annotation: @CHARLIMIT=7 |
| 207 | jc_othericd10procdate1<br>Show the field ONLY if:<br>[jc_princicd10procedure] <> '' and [hide_nonessential_fields(1)]='0' | ✪ ICD-10-PCS Other Procedure Dates | text (datetime_mdy, Min: 2010-01-01 00:00, Max: 2099-12-31 23:59), Identifier |
| 208 | jc_othericd10proc1<br>Show the field ONLY if:<br>[jc_othericd10procdate1] <> '' and [hide_nonessential_fields(1)]='0' | ✪ ICD-10-PCS Other Procedure Codes | text<br>Field Annotation: @CHARLIMIT=7 |

| | | | |
|---|---|---|---|
| 209 | jc_othericd10procdate2<br>Show the field ONLY if:<br>[jc_othericd10proc1] <> '' and [hide_nonessential_fields(1)]='0' | ✪ ICD-10-PCS Other Procedure Dates | text (datetime_mdy, Min: 2010-01-01 00:00, Max: 2099-12-31 23:59), Identifier |
| 210 | jc_othericd10proc2<br>Show the field ONLY if:<br>[jc_othericd10procdate2] <> '' and [hide_nonessential_fields(1)]='0' | ✪ ICD-10-PCS Other Procedure Codes | text<br>Field Annotation: @CHARLIMIT=7 |
| 211 | jc_othericd10procdate3<br>Show the field ONLY if:<br>[jc_othericd10proc2] <> '' and [hide_nonessential_fields(1)]='0' | ✪ ICD-10-PCS Other Procedure Dates | text (datetime_mdy, Min: 2010-01-01 00:00, Max: 2099-12-31 23:59), Identifier |
| 212 | jc_othericd10proc3<br>Show the field ONLY if:<br>[jc_othericd10procdate3] <> '' and [hide_nonessential_fields(1)]='0' | ✪ ICD-10-PCS Other Procedure Codes | text<br>Field Annotation: @CHARLIMIT=7 |
| 213 | jc_othericd10procdate4<br>Show the field ONLY if:<br>[jc_othericd10proc3] <> '' and [hide_nonessential_fields(1)]='0' | ✪ ICD-10-PCS Other Procedure Dates | text (datetime_mdy, Min: 2010-01-01 00:00, Max: 2099-12-31 23:59), Identifier |
| 214 | jc_othericd10proc4<br>Show the field ONLY if:<br>[jc_othericd10procdate4] <> '' and [hide_nonessential_fields(1)]='0' | ✪ ICD-10-PCS Other Procedure Codes | text<br>Field Annotation: @CHARLIMIT=7 |
| 215 | jc_othericd10procdate5<br>Show the field ONLY if:<br>[jc_othericd10proc4] <> '' and [hide_nonessential_fields(1)]='0' | ✪ ICD-10-PCS Other Procedure Dates | text (datetime_mdy, Min: 2010-01-01 00:00, Max: 2099-12-31 23:59), Identifier |
| 216 | jc_othericd10proc5<br>Show the field ONLY if:<br>[jc_othericd10procdate5] <> '' and [hide_nonessential_fields(1)]='0' | ✪ ICD-10-PCS Other Procedure Codes | text<br>Field Annotation: @CHARLIMIT=7 |
| 217 | jc_othericd10procdate6<br>Show the field ONLY if:<br>[jc_othericd10proc5] <> '' and [hide_nonessential_fields(1)]='0' | ✪ ICD-10-PCS Other Procedure Dates | text (datetime_mdy, Min: 2010-01-01 00:00, Max: 2099-12-31 23:59), Identifier |

| | | | |
|---|---|---|---|
| 218 | jc_othericd10proc6<br><br>Show the field ONLY if:<br>[jc_othericd10procdate6] <> '' and [hide_nonessential_fields(1)]='0' | ✪ ICD-10-PCS Other Procedure Codes | text<br>Field Annotation: @CHARLIMIT=7 |
| 219 | jc_othericd10procdate7<br><br>Show the field ONLY if:<br>[jc_othericd10proc6] <> '' and [hide_nonessential_fields(1)]='0' | ✪ ICD-10-PCS Other Procedure Dates | text (datetime_mdy, Min: 2010-01-01 00:00, Max: 2099-12-31 23:59), Identifier |
| 220 | jc_othericd10proc7<br><br>Show the field ONLY if:<br>[jc_othericd10procdate7] <> '' and [hide_nonessential_fields(1)]='0' | ✪ ICD-10-PCS Other Procedure Codes | text<br>Field Annotation: @CHARLIMIT=7 |
| 221 | jc_othericd10procdate8<br><br>Show the field ONLY if:<br>[jc_othericd10proc7] <> '' and [hide_nonessential_fields(1)]='0' | ✪ ICD-10-PCS Other Procedure Dates | text (datetime_mdy, Min: 2010-01-01 00:00, Max: 2099-12-31 23:59), Identifier |
| 222 | jc_othericd10proc8<br><br>Show the field ONLY if:<br>[jc_othericd10procdate8] <> '' and [hide_nonessential_fields(1)]='0' | ✪ ICD-10-PCS Other Procedure Codes | text<br>Field Annotation: @CHARLIMIT=7 |
| 223 | jc_othericd10procdate9<br><br>Show the field ONLY if:<br>[jc_othericd10proc8] <> '' and [hide_nonessential_fields(1)]='0' | ✪ ICD-10-PCS Other Procedure Dates | text (datetime_mdy, Min: 2010-01-01 00:00, Max: 2099-12-31 23:59), Identifier |
| 224 | jc_othericd10proc9<br><br>Show the field ONLY if:<br>[jc_othericd10procdate9] <> '' and [hide_nonessential_fields(1)]='0' | ✪ ICD-10-PCS Other Procedure Codes | text<br>Field Annotation: @CHARLIMIT=7 |
| 225 | jc_othericd10procdate10<br><br>Show the field ONLY if:<br>[jc_othericd10proc9] <> '' and [hide_nonessential_fields(1)]='0' | ✪ ICD-10-PCS Other Procedure Dates | text (datetime_mdy, Min: 2010-01-01 00:00, Max: 2099-12-31 23:59), Identifier |
| 226 | jc_othericd10proc10<br><br>Show the field ONLY if:<br>[jc_othericd10procdate10] <> '' and [hide_nonessential_fields(1)]='0' | ✪ ICD-10-PCS Other Procedure Codes | text<br>Field Annotation: @CHARLIMIT=7 |

| | | | |
|---|---|---|---|
| 227 | jc_othericd10procdate11<br><br>Show the field ONLY if:<br>[jc_othericd10proc10] <> '' and [hide_nonessential_fields(1)]='0' | ✪ ICD-10-PCS Other Procedure Dates | text (datetime_mdy, Min: 2010-01-01 00:00, Max: 2099-12-31 23:59), Identifier |
| 228 | jc_othericd10proc11<br><br>Show the field ONLY if:<br>[jc_othericd10procdate11] <> '' and [hide_nonessential_fields(1)]='0' | ✪ ICD-10-PCS Other Procedure Codes | text<br>Field Annotation: @CHARLIMIT=7 |
| 229 | jc_othericd10procdate12<br><br>Show the field ONLY if:<br>[jc_othericd10proc11] <> '' and [hide_nonessential_fields(1)]='0' | ✪ ICD-10-PCS Other Procedure Dates | text (datetime_mdy, Min: 2010-01-01 00:00, Max: 2099-12-31 23:59), Identifier |
| 230 | jc_othericd10proc12<br><br>Show the field ONLY if:<br>[jc_othericd10procdate12] <> '' and [hide_nonessential_fields(1)]='0' | ✪ ICD-10-PCS Other Procedure Codes | text<br>Field Annotation: @CHARLIMIT=7 |
| 231 | jc_othericd10procdate13<br><br>Show the field ONLY if:<br>[jc_othericd10proc12] <> '' and [hide_nonessential_fields(1)]='0' | ✪ ICD-10-PCS Other Procedure Dates | text (datetime_mdy, Min: 2010-01-01 00:00, Max: 2099-12-31 23:59), Identifier |
| 232 | jc_othericd10proc13<br><br>Show the field ONLY if:<br>[jc_othericd10procdate13] <> '' and [hide_nonessential_fields(1)]='0' | ✪ ICD-10-PCS Other Procedure Codes | text<br>Field Annotation: @CHARLIMIT=7 |
| 233 | jc_othericd10procdate14<br><br>Show the field ONLY if:<br>[jc_othericd10proc13] <> '' and [hide_nonessential_fields(1)]='0' | ✪ ICD-10-PCS Other Procedure Dates | text (datetime_mdy, Min: 2010-01-01 00:00, Max: 2099-12-31 23:59), Identifier |
| 234 | jc_othericd10proc14<br><br>Show the field ONLY if:<br>[jc_othericd10procdate14] <> '' and [hide_nonessential_fields(1)]='0' | ✪ ICD-10-PCS Other Procedure Codes | text<br>Field Annotation: @CHARLIMIT=7 |
| 235 | jc_othericd10procdate15<br><br>Show the field ONLY if:<br>[jc_othericd10proc14] <> '' and [hide_nonessential_fields(1)]='0' | ✪ ICD-10-PCS Other Procedure Dates | text (datetime_mdy, Min: 2010-01-01 00:00, Max: 2099-12-31 23:59), Identifier |

| | | | |
|---|---|---|---|
| 236 | jc_othericd10proc15<br><br>Show the field ONLY if:<br>[jc_othericd10procdate15] <> '' and [hide_nonessential_fields(1)]='0' | ✪ ICD-10-PCS Other Procedure Codes | text<br>Field Annotation: @CHARLIMIT=7 |
| 237 | jc_othericd10procdate16<br><br>Show the field ONLY if:<br>[jc_othericd10proc15] <> '' and [hide_nonessential_fields(1)]='0' | ✪ ICD-10-PCS Other Procedure Dates | text (datetime_mdy, Min: 2010-01-01 00:00, Max: 2099-12-31 23:59), Identifier |
| 238 | jc_othericd10proc16<br><br>Show the field ONLY if:<br>[jc_othericd10procdate16] <> '' and [hide_nonessential_fields(1)]='0' | ✪ ICD-10-PCS Other Procedure Codes | text<br>Field Annotation: @CHARLIMIT=7 |
| 239 | jc_othericd10procdate17<br><br>Show the field ONLY if:<br>[jc_othericd10proc16] <> '' and [hide_nonessential_fields(1)]='0' | ✪ ICD-10-PCS Other Procedure Dates | text (datetime_mdy, Min: 2010-01-01 00:00, Max: 2099-12-31 23:59), Identifier |
| 240 | jc_othericd10proc17<br><br>Show the field ONLY if:<br>[jc_othericd10procdate17] <> '' and [hide_nonessential_fields(1)]='0' | ✪ ICD-10-PCS Other Procedure Codes | text<br>Field Annotation: @CHARLIMIT=7 |
| 241 | jc_othericd10procdate18<br><br>Show the field ONLY if:<br>[jc_othericd10proc17] <> '' and [hide_nonessential_fields(1)]='0' | ✪ ICD-10-PCS Other Procedure Dates | text (datetime_mdy, Min: 2010-01-01 00:00, Max: 2099-12-31 23:59), Identifier |
| 242 | jc_othericd10proc18<br><br>Show the field ONLY if:<br>[jc_othericd10procdate18] <> '' and [hide_nonessential_fields(1)]='0' | ✪ ICD-10-PCS Other Procedure Codes | text<br>Field Annotation: @CHARLIMIT=7 |
| 243 | jc_othericd10procdate19<br><br>Show the field ONLY if:<br>[jc_othericd10proc18] <> '' and [hide_nonessential_fields(1)]='0' | ✪ ICD-10-PCS Other Procedure Dates | text (datetime_mdy, Min: 2010-01-01 00:00, Max: 2099-12-31 23:59), Identifier |
| 244 | jc_othericd10proc19<br><br>Show the field ONLY if:<br>[jc_othericd10procdate19] <> '' and [hide_nonessential_fields(1)]='0' | ✪ ICD-10-PCS Other Procedure Codes | text<br>Field Annotation: @CHARLIMIT=7 |

| | | | |
|---|---|---|---|
| 245 | jc_othericd10procdate20<br><br>Show the field ONLY if:<br>[jc_othericd10proc19] <> '' and [hide_nonessential_fields(1)]='0' | ✪ ICD-10-PCS Other Procedure Dates | text (datetime_mdy, Min: 2010-01-01 00:00, Max: 2099-12-31 23:59), Identifier |
| 246 | jc_othericd10proc20<br><br>Show the field ONLY if:<br>[jc_othericd10procdate20] <> '' and [hide_nonessential_fields(1)]='0' | ✪ ICD-10-PCS Other Procedure Codes | text<br>Field Annotation: @CHARLIMIT=7 |
| 247 | jc_othericd10procdate21<br><br>Show the field ONLY if:<br>[jc_othericd10proc20] <> '' and [hide_nonessential_fields(1)]='0' | ✪ ICD-10-PCS Other Procedure Dates | text (datetime_mdy, Min: 2010-01-01 00:00, Max: 2099-12-31 23:59) |
| 248 | jc_othericd10proc21<br><br>Show the field ONLY if:<br>[jc_othericd10procdate21] <> '' and [hide_nonessential_fields(1)]='0' | ✪ ICD-10-PCS Other Procedure Codes | text<br>Field Annotation: @CHARLIMIT=7 |
| 249 | jc_othericd10procdate22<br><br>Show the field ONLY if:<br>[jc_othericd10proc21] <> '' and [hide_nonessential_fields(1)]='0' | ✪ ICD-10-PCS Other Procedure Dates | text (datetime_mdy, Min: 2010-01-01 00:00, Max: 2099-12-31 23:59), Identifier |
| 250 | jc_othericd10proc22<br><br>Show the field ONLY if:<br>[jc_othericd10procdate22] <> '' and [hide_nonessential_fields(1)]='0' | ✪ ICD-10-PCS Other Procedure Codes | text<br>Field Annotation: @CHARLIMIT=7 |
| 251 | jc_othericd10procdate23<br><br>Show the field ONLY if:<br>[jc_othericd10proc22] <> '' and [hide_nonessential_fields(1)]='0' | ✪ ICD-10-PCS Other Procedure Dates | text (datetime_mdy, Min: 2010-01-01 00:00, Max: 2099-12-31 23:59), Identifier |
| 252 | jc_othericd10proc23<br><br>Show the field ONLY if:<br>[jc_othericd10procdate23] <> '' and [hide_nonessential_fields(1)]='0' | ✪ ICD-10-PCS Other Procedure Codes | text<br>Field Annotation: @CHARLIMIT=7 |
| 253 | jc_othericd10procdate24<br><br>Show the field ONLY if:<br>[jc_othericd10proc23] <> '' and [hide_nonessential_fields(1)]='0' | ✪ ICD-10-PCS Other Procedure Dates | text (datetime_mdy, Min: 2010-01-01 00:00, Max: 2099-12-31 23:59), Identifier |

| | | | |
|---|---|---|---|
| 254 | jc_othericd10proc24<br><br>Show the field ONLY if:<br>[jc_othericd10procdate24] <> '' and [hide_nonessential_fields(1)]='0' | ✪ ICD-10-PCS Other Procedure Codes | text<br>Field Annotation: @CHARLIMIT=7 |
| 255 | admittingdiagnosisicd10<br><br>Show the field ONLY if:<br>([site] = 'SMCA' or [site] = 'UMCB') and ([treatment_recvd]='1' or [gs_notadmit]<>'1') and [hide_nonessential_fields(1)]='0' | Section Header: *Admitting Diagnosis*<br><br>✪ What was the ICD-10-CM diagnosis code selected as the admitting diagnosis for this patient? | text |
| 256 | icd10stroke<br><br>Show the field ONLY if:<br>[gs_icd10nostroke]<>'1' and [hide_nonessential_fields(1)]='0' | Section Header: *Discharge Diagnosis*<br><br>✪ ICD-10 discharge diagnosis related to stroke<br>*3-7 char. [NO DECIMALS OR DOTS]* | text |
| 257 | gs_icd10nostroke<br><br>Show the field ONLY if:<br>[gs_notadmit]<>'1' or [gs_notadmitwhy]='5' and [hide_nonessential_fields(1)]='0' | ✪ No Stroke or TIA Code | radio<br>1 \| No Stroke or TIA Related ICD-10-CM Code Present |
| 258 | clinical_codes_complete | Section Header: *Form Status*<br><br>Complete? | dropdown<br>0 \| Incomplete<br>1 \| Unverified<br>2 \| Complete |
| Instrument: **Admission** (admission) | | | Collapse |
| 259 | desc_demographics_shazam<br><br>Show the field ONLY if:<br>([treatment_recvd]='1' or [gs_notadmit]<>'1') | Section Header: *Demographics* | descriptive<br>Field Annotation: @HIDDEN |
| 260 | desc_gender_dob | Gender Birth Date Age {jc_gender:icons} {jc_dob:icons} {gs_age:icons} | descriptive |
| 261 | jc_gender<br><br>Show the field ONLY if:<br>([treatment_recvd]='1' or [gs_notadmit]<>'1') | Gender | radio<br>1 \| Male<br>2 \| Female<br>3 \| Unknown<br><br>Custom alignment: RH |

| | | | |
|---|---|---|---|
| 262 | jc_dob<br><br>Show the field ONLY if:<br>([treatment_recvd]='1' or [gs_notadmit]<>'1') | Birth Date | text (date_mdy, Min: 1890-01-01, Max: 2099-12-31), Identifier |
| 263 | gs_age<br><br>Show the field ONLY if:<br>([treatment_recvd]='1' or [gs_notadmit]<>'1') | Age | calc, Identifier<br>Calculation: rounddown(datediff([jc_dob],[jc_arrdatetime],"y","mdy",true),0) |
| 264 | desc_home_zip_pay<br><br>Show the field ONLY if:<br>([treatment_recvd]='1' or [gs_notadmit]<>'1') and [hide_nonessential_fields(1)]='0' | {gs_homeless:icons} Zip Code: {gs_zip1:icons} - {gs_zip2:icons} Payment Source {paysource:icons} | descriptive |
| 265 | gs_homeless<br><br>Show the field ONLY if:<br>([treatment_recvd]='1' or [gs_notadmit]<>'1') and [hide_nonessential_fields(1)]='0' | Homeless | radio<br>1 Homeless |
| 266 | gs_zip1<br><br>Show the field ONLY if:<br>([treatment_recvd]='1' or [gs_notadmit]<>'1') and [gs_homeless] = '' and [hide_nonessential_fields(1)] ='0' | Zip Code | text (zipcode), Identifier |
| 267 | gs_zip2<br><br>Show the field ONLY if:<br>([treatment_recvd]='1' or [gs_notadmit]<>'1') and [gs_homeless] = '' and [hide_nonessential_fields(1)] ='0' | Zip Code Extension | text (integer, Max: 9999), Identifier |

| | | | |
|---|---|---|---|
| 268 | paysource<br><br>Show the field ONLY if:<br>([treatment_recvd]='1' or [gs_notadmit]<>'1') and [hide_nonessential_fields(1)]='0' | Payment Source<br>*Select all that apply.* | checkbox<br>1 \| paysource___1 \| Medicare Title 18<br>2 \| paysource___2 \| Medicaid Title 19<br>3 \| paysource___3 \| Medicare - Private/HMO/PPO/Other<br>4 \| paysource___4 \| Medicaid - Private/HMO/PPO/Other<br>5 \| paysource___5 \| Private/HMO/PPO/Other<br>6 \| paysource___6 \| VA/CHAMPVA/Tricare<br>7 \| paysource___7 \| Self-Pay/No Insurance<br>8 \| paysource___8 \| Other/Not Documented/UTD<br><br>Field Annotation: @NONEOFTHEABOVE='8' |
| 269 | jc_medicare<br><br>Show the field ONLY if:<br>([treatment_recvd]='1' or [gs_notadmit]<>'1') | What is the patient's source of payment for this episode of care? | radio<br>1 \| Medicare<br>2 \| Non-Medicare<br><br>Custom alignment: RH |
| 270 | desc_race_shazam<br><br>Show the field ONLY if:<br>([treatment_recvd]='1' or [gs_notadmit]<>'1') | Section Header: *Race and Ethnicity* | descriptive<br>Field Annotation: @HIDDEN |
| 271 | desc_race | Race {gs_race:icons} {gs_race_asian:icons} {gs_race_native:icons}<br>Hispanic Ethnicity If Yes, Hispanic Ethnicity {sp_ethnic:icons}<br>{sp_ethnic_opt:icons} | descriptive |
| 272 | gs_race<br><br>Show the field ONLY if:<br>([treatment_recvd]='1' or [gs_notadmit]<>'1') | Race | checkbox<br>4 \| gs_race___4 \| American Indian/Alaska Native<br>3 \| gs_race___3 \| Asian<br>2 \| gs_race___2 \| Black or African American<br>5 \| gs_race___5 \| Native Hawaiian/Pacific Islander<br>1 \| gs_race___1 \| White<br>6 \| gs_race___6 \| UTD<br><br>Field Annotation: @NONEOFTHEABOVE="6" |

| | | | |
|---|---|---|---|
| 273 | gs_race_asian<br><br>Show the field ONLY if:<br>[gs_race(3)] = '1' | Race Asian | checkbox<br>1 \| gs_race_asian___1 \| Asian Indian<br>2 \| gs_race_asian___2 \| Chinese<br>3 \| gs_race_asian___3 \| Filipino<br>4 \| gs_race_asian___4 \| Japanese<br>5 \| gs_race_asian___5 \| Korean<br>6 \| gs_race_asian___6 \| Vietnamese<br>7 \| gs_race_asian___7 \| Other Asian |
| 274 | gs_race_native<br><br>Show the field ONLY if:<br>[gs_race(5)] = '1' | Race Native Hawaiian | checkbox<br>1 \| gs_race_native___1 \| Native Hawaiian<br>2 \| gs_race_native___2 \| Guamanian or Chamorro<br>3 \| gs_race_native___3 \| Samoan<br>4 \| gs_race_native___4 \| Other Pacific Islander |
| 275 | sp_ethnic<br><br>Show the field ONLY if:<br>([treatment_recvd]='1' or [gs_nota dmit]<>'1') | Hispanic Ethnicity | radio<br>1 \| Yes<br>2 \| No/UTD<br><br>Custom alignment: RH |
| 276 | sp_ethnic_opt<br><br>Show the field ONLY if:<br>[sp_ethnic] = "1" | If Yes, Hispanic Ethnicity | dropdown<br>1 \| Mexican, Mexican American, Chicano/a<br>2 \| Puerto Rican<br>3 \| Cuban<br>4 \| Another Hispanic, Latino or Spanish Origin |
| 277 | jc_clinical<br><br>Show the field ONLY if:<br>([treatment_recvd]='1' or [gs_nota dmit]<>'1') and [hide_nonessential _fields(1)]='0' | Section Header: *Arrival & Admission Information*<br><br>✪ During this hospital stay, was the patient enrolled in a clinical trial in which patients with the same condition as the measure set were being studied (i.e. STK, VTE)? | radio<br>1 \| Yes<br>2 \| No<br><br>Custom alignment: RH<br>Field Annotation: @DEFAULT='2' |

| | | | |
|---|---|---|---|
| 278 | clinical_trial_name<br><br>Show the field ONLY if:<br>([site]='SMCA' or [site]='SMCH' or [site]='SMCW' or [site]='UMCB') and ([treatment_recvd]='1' or [gs_notadmit]<>'1') and [hide_nonessential_fields(1)]='0' | ✪ If Enrolled in Clinical Trial or Research Study, Name of Trial or Study | text |
| 279 | jc_carotid<br><br>Show the field ONLY if:<br>([treatment_recvd]='1' or [gs_notadmit]<>'1') and [hide_nonessential_fields(1)]='0' | Was this patient admitted for the sole purpose of performance of elective carotid intervention? | radio<br>1 Yes<br>2 No<br><br>Custom alignment: RH |
| 280 | desc_referring_hosp | | descriptive<br>Field Annotation: @HIDDEN |
| 281 | desc_symptomlocation<br><br>Show the field ONLY if:<br>([treatment_recvd]='1' or [gs_notadmit]<>'1') | Patient location when stroke symptoms discovered:From: Initial Stroke Information [gs_symptomlocation] | descriptive |
| 282 | desc_patientarrival<br><br>Show the field ONLY if:<br>([treatment_recvd]='1' or [gs_notadmit]<>'1') | How patient arrived at your hospital:From: Initial Stroke Information [gs_patientarrival] | descriptive |
| 283 | desc_transfer_from_hosp<br><br>Show the field ONLY if:<br>[transfer_from_hosp]<>'' | Patient transferred from: [transfer_from_hosp] | descriptive |
| 284 | desc_gs_ahaid2nd<br><br>Show the field ONLY if:<br>[gs_ahaid2nd]=1 | Patient transferred from: Transfer Hospital Not Documented | descriptive |
| 285 | desc_referringhosp | Referring hospital arrival date/time {gs_refarrdt_precision:icons} {gs_refarrdt:icons} Referring hospital discharge date/time {gs_rechospdt_precision:icons} {gs_rechospdt:icons} If patient transferred to your hospital, select transfer reason(s) {gs_ptrecvdreason:icons} | descriptive |
| 286 | gs_refarrdt_precision<br><br>Show the field ONLY if:<br>[gs_patientarrival] = '3' and [hide_nonessential_fields(1)]='0' | Referring hospital arrival date/time Precision | dropdown<br>5 Date & Time (MM/DD/YYYY HH24:MM)<br>3 Date Only (MM/DD/YYYY)<br>0 Unknown |

<table>
<tr><td>287</td><td>gs_refarrdt<br><br>Show the field ONLY if:<br>[gs_patientarrival] = '3' and [gs_refarrdt_precision] <> '0' and [hide_nonessential_fields(1)]='0'</td><td>Referring hospital arrival date/time</td><td>text (datetime_mdy, Min: 2010-01-01 00:00, Max: 2099-12-31 23:59)</td></tr>
<tr><td>288</td><td>gs_rechospdt_precision<br><br>Show the field ONLY if:<br>[gs_patientarrival] = '3' and [hide_nonessential_fields(1)]='0'</td><td>Referring hospital discharge date/time Precision</td><td>dropdown<table><tr><td>5</td><td>Date & Time (MM/DD/YYYY HH24:MM)</td></tr><tr><td>3</td><td>Date Only (MM/DD/YYYY)</td></tr><tr><td>0</td><td>Unknown</td></tr></table></td></tr>
<tr><td>289</td><td>gs_rechospdt<br><br>Show the field ONLY if:<br>[gs_patientarrival] = '3' and [gs_rechospdt_precision] <> '0' and [hide_nonessential_fields(1)]='0'</td><td>Referring hospital discharge date/time</td><td>text (datetime_mdy, Min: 2010-01-01 00:00, Max: 2099-12-31 23:59)</td></tr>
<tr><td>290</td><td>gs_ptrecvdreason<br><br>Show the field ONLY if:<br>[gs_patientarrival] = '3' and [hide_nonessential_fields(1)]='0'</td><td>If patient transferred to your hospital, select transfer reason(s)</td><td>checkbox<table><tr><td>1</td><td>gs_ptrecvdreason___1</td><td>Evaluation for IV tPA up to 4.5 hours</td></tr><tr><td>2</td><td>gs_ptrecvdreason___2</td><td>Post Management of IV tPA (e.g. Drip and Ship)</td></tr><tr><td>3</td><td>gs_ptrecvdreason___3</td><td>Evaluation for Endovascular thrombectomy</td></tr><tr><td>4</td><td>gs_ptrecvdreason___4</td><td>Advanced stroke care (e.g., Neurocritical care, surgical or other time critical therapy)</td></tr><tr><td>5</td><td>gs_ptrecvdreason___5</td><td>Patient/family request</td></tr><tr><td>6</td><td>gs_ptrecvdreason___6</td><td>Other advanced care (not stroke related)</td></tr><tr><td>7</td><td>gs_ptrecvdreason___7</td><td>Not documented</td></tr></table></td></tr>
<tr><td>291</td><td>jc_edpatient<br><br>Show the field ONLY if:<br>([treatment_recvd]='1' or [gs_notadmit]<>'1')</td><td>Was the patient an ED patient at the facility?</td><td>radio<table><tr><td>1</td><td>Yes</td></tr><tr><td>2</td><td>No</td></tr></table><br>Custom alignment: RH</td></tr>
</table>

| | | | |
|---|---|---|---|
| 292 | c_directadm<br><br>Show the field ONLY if:<br>([site] = 'SMCA' or [site] = 'UMCB') and [jc_edpatient] = '2' and [hide_nonessential_fields(1)]='0' | Was the patient a direct admission to the hospital? | radio<br>1 Yes<br>2 No<br><br>Custom alignment: RH |
| 293 | gs_placercd<br><br>Show the field ONLY if:<br>([site]='SMCA' or [site]='SMCH' or [site]='SMCW' or [site]='UMCB') and ([treatment_recvd]='1' or [gs_notadmit]<>'1') and [hide_nonessential_fields(1)]='0' | Where patient first received care at your hospital: | dropdown<br>1 Emergency Department/Urgent Care<br>2 Direct Admit or direct to floor<br>3 Imaging suite prior to ED<br>4 ND or Cannot be determined |
| 294 | gs_prehosp_ems<br><br>Show the field ONLY if:<br>([site]='SMCA' or [site]='SMCH' or [site]='SMCW' or [site]='UMCB') and ([gs_patientarrival] = '1' or [gs_patientarrival] = '10') | Advanced notification by EMS? | radio<br>1 Yes<br>2 No/ND<br>3 N/A<br><br>Custom alignment: RH<br>Field Annotation: @HIDECHOICE='3' |
| 295 | gs_careunit<br><br>Show the field ONLY if:<br>([site]='SMCA' or [site]='SMCH' or [site]='SMCW' or [site]='UMCB') and ([treatment_recvd]='1' or [gs_notadmit]<>'1') and [hide_nonessential_fields(1)]='0' | ✪ Where was the patient cared for and by whom?<br>*Check all that apply.* | checkbox<br>1 gs_careunit___1 Neuro Admission<br>2 gs_careunit___2 Other Service Admission<br>3 gs_careunit___3 Stroke Consult<br>4 gs_careunit___4 No Stroke Consult<br>5 gs_careunit___5 In Stroke Unit<br>6 gs_careunit___6 Not in Stroke Unit |
| 296 | gs_nomedhx<br><br>Show the field ONLY if:<br>([treatment_recvd]='1' or [gs_notadmit]<>'1') | Section Header: *Medical History*<br><br>No Medical History | radio<br>1 No medical history<br><br>Field Annotation: @HIDDEN |

| | | | |
|---|---|---|---|
| 297 | gs_med_hist<br><br>Show the field ONLY if:<br>[gs_nomedhx] <> '1' and ([treatment_recvd]='1' or [gs_notadmit]<>'1') | Previously known medical hx of:<br>*Select all that apply.* | checkbox<br><br>Field Annotation: @NONEOFTHEABOVE='0' |

| | | |
|---|---|---|
| 0 | gs_med_hist___0 | None |
| 1 | gs_med_hist___1 | Atrial Fib/Flutter |
| 4 | gs_med_hist___4 | CAD/prior MI |
| 5 | gs_med_hist___5 | Carotid Stenosis |
| 14 | gs_med_hist___14 | Current pregnancy (up to 6 weeks post partum) |
| 25 | gs_med_hist___25 | DVT/PE |
| 29 | gs_med_hist___29 | Dementia |
| 24 | gs_med_hist___24 | Depression |
| 6 | gs_med_hist___6 | Diabetes Mellitus |
| 17 | gs_med_hist___17 | Drugs/Alcohol Abuse |
| 10 | gs_med_hist___10 | Dyslipidemia |
| 27 | gs_med_hist___27 | E-Cigarette Use (Vaping) |
| 26 | gs_med_hist___26 | Familial hypercholesterolemia |
| 18 | gs_med_hist___18 | Family History of Stroke |
| 12 | gs_med_hist___12 | HF |
| 19 | gs_med_hist___19 | HRT |
| 28 | gs_med_hist___28 | HX of Emerging Infectious Disease |
| 8 | gs_med_hist___8 | Hypertension |
| 20 | gs_med_hist___20 | Migraine |
| 21 | gs_med_hist___21 | Obesity/Overweight |
| 15 | gs_med_hist___15 | Previous Stroke |
| 16 | gs_med_hist___16 | Previous TIA |
| 2 | gs_med_hist___2 | Prosthetic Heart Valve |
| 7 | gs_med_hist___7 | PVD |
| 22 | gs_med_hist___22 | Renal insufficiency - chronic |
| 13 | gs_med_hist___13 | Sickle Cell |
| 23 | gs_med_hist___23 | Sleep Apnea |
| 9 | gs_med_hist___9 | Smoker |

| | | | |
|---|---|---|---|
| 298 | dmtype<br><br>Show the field ONLY if:<br>[gs_med_hist(6)] = '1' | Diabetes Type | radio<br>1 Type I<br>2 Type II<br>3 ND<br><br>Custom alignment: RH |
| 299 | dmduration<br><br>Show the field ONLY if:<br>[gs_med_hist(6)] = '1' | Duration of Diabetes hx | radio<br>1 < 5 years<br>2 5 - < 10 years<br>3 10 - < 20 years<br>4 >= 20 years<br>5 Unknown |
| 300 | hxinfectopt<br><br>Show the field ONLY if:<br>[gs_med_hist(28)]='1' | Emerging Infectious Disease | checkbox<br>1 hxinfectopt___1 MERS<br>2 hxinfectopt___2 SARS-COV-1<br>3 hxinfectopt___3 SARS-COV-2 (COVID-19)<br>4 hxinfectopt___4 Other infectious respiratory pathogen |
| 301 | gs_med_hist_stroke<br><br>Show the field ONLY if:<br>[gs_med_hist(15)] = '1' | Previous Stroke | checkbox<br>1 gs_med_hist_stroke___1 Ischemic stroke<br>2 gs_med_hist_stroke___2 ICH<br>3 gs_med_hist_stroke___3 SAH<br>4 gs_med_hist_stroke___4 Not Specified<br><br>Field Annotation: @NONEOFTHEABOVE='4' |
| 302 | gs_ambulatprior<br><br>Show the field ONLY if:<br>([site]='SMCA' or [site]='SMCH' or [site]='SMCW' or [site]='UMCB') and ([treatment_recvd]='1' or [gs_notadmit]<>'1') and [hide_nonessential_fields(1)]='0' | Ambulatory status prior to the current event? | dropdown<br>1 Able to ambulate independently (no help from another person) w/ or w/o device<br>2 With assistance (from person)<br>3 Unable to ambulate<br>9 ND |

| | | | |
|---|---|---|---|
| 303 | c_prestrokemrs<br><br>Show the field ONLY if:<br>([site]='SMCA' or [site]='SMCH' or [site]='SMCW' or [site]='UMCB') and [hide_nonessential_fields(1)]='0' | Pre-stroke Modified Rankin Score<br>*GWTG Field - Abstract per coding instructions.* | dropdown<br>1 \| A score value of 0, 1, or 2 was documented in the medical record, OR physician/APN/PA documentation that the patient was able to look after self without daily help prior to this acute stroke episode.<br>2 \| A score value of 3, 4, or 5 was documented in the medical record, OR physician/APN/PA documentation that the patient could NOT look after self without daily help prior to this acute stroke episode.<br>3 \| A score value was not documented, OR unable to determine (UTD) from the medical record documentation. |
| 304 | c_prestrokemrs_score<br><br>Show the field ONLY if:<br>([site]='SMCA' or [site]='SMCH' or [site]='SMCW' or [site]='UMCB') and [hide_nonessential_fields(1)]='0' | Pre-stroke Modified Rankin Score | dropdown<br>0 \| 0 - No symptoms at all<br>1 \| 1 - No significant disability despite symptoms: Able to carry out all usual activities<br>2 \| 2 - Slight disability<br>3 \| 3 - Moderate disability: Requiring some help but able to walk without assistance<br>4 \| 4 - Moderate to severe disability: Unable to walk without assistance and unable to attend to own bodily needs without assistance<br>5 \| 5 - Severe disability: Bedridden, incontinent and requiring constant nursing care and attention<br>9 \| ND |
| 305 | gs_tiaduration<br><br>Show the field ONLY if:<br>([site]='SMCA' or [site]='SMCH' or [site]='SMCW' or [site]='UMCB') and [gs_stroketype] = '3' and ([treatment_recvd]='1' or [gs_notadmit]<>'1') and [hide_nonessential_fields(1)]='0' | Section Header: *Diagnosis & Evaluation*<br><br>Symptom Duration if diagnosis of Transient Ischemic Attack (less than 24 hours) | dropdown<br>1 \| Less then 10 minutes<br>2 \| 10 - 59 minutes<br>3 \| >= 60 minutes<br>4 \| ND |
| 306 | desc_gs_nihssperformed<br><br>Show the field ONLY if:<br>([treatment_recvd]='1' or [gs_notadmit]<>'1') | Initial NIH Stroke Scale:[gs_nihssperformed]From: Initial Stroke Information | descriptive |

| | | | |
|---|---|---|---|
| 307 | desc_gs_nihssproretro<br><br>Show the field ONLY if:<br>[gs_nihssperformed] = '1' and ([treatment_recvd]='1' or [gs_notadmit]<>'1') | if Yes, Initial NIHSS:[gs_nihssproretro]From: Initial Stroke Information | descriptive |
| 308 | desc_gs_nihssadmit<br><br>Show the field ONLY if:<br>[gs_nihssperformed] = '1' and ([treatment_recvd]='1' or [gs_notadmit]<>'1') | Total Score:[gs_nihssadmit]From: Initial Stroke Information | descriptive |
| 309 | show_nihss<br><br>Show the field ONLY if:<br>([site]='SMCA' or [site]='SMCH' or [site]='SMCW' or [site]='UMCB') and ([treatment_recvd]='1' or [gs_notadmit]<>'1') | Show NIHSS exam questions | radio<br>1 Show<br><br>Custom alignment: RH |
| 310 | gs_nihss1a<br><br>Show the field ONLY if:<br>[show_nihss] = '1' | 1.a. Level of consciousness | dropdown<br>2 0-Alert<br>3 1-Not alert, but arousable with minimal stimulation<br>4 2-Not alert, requires repeated stimulation to attend<br>5 3-Coma |
| 311 | gs_nihss1b<br><br>Show the field ONLY if:<br>[show_nihss] = '1' | 1.b. Ask patient the month and their age: | dropdown<br>2 0-Answers both correctly<br>3 1-Answers one correctly<br>4 2-Both incorrect |
| 312 | gs_nihss1c<br><br>Show the field ONLY if:<br>[show_nihss] = '1' | 1.c. Ask patient to open and close eyes; make fist and let go: | dropdown<br>2 0-Obeys both correctly<br>3 1-Obeys one correctly<br>4 2-Both incorrect |
| 313 | gs_nihss2<br><br>Show the field ONLY if:<br>[show_nihss] = '1' | 2. Best gaze (only horizontal eye movement): | dropdown<br>2 0-Normal<br>3 1-Partial gaze palsy<br>4 2-Forced deviation |

| | | | |
|---|---|---|---|
| 314 | gs_nihss3<br><br>Show the field ONLY if:<br>[show_nihss] = '1' | 3. Visual field testing | dropdown<br>2 — 0-No visual field loss<br>3 — 1-Partial hemianopia<br>4 — 2-Complete hemianopia<br>5 — 3-Bilateral hemianopia (blind including cortical blindness) |
| 315 | gs_nihss4<br><br>Show the field ONLY if:<br>[show_nihss] = '1' | 4. Facial paresis (Ask patient to show teeth or raise eyebrows and close eyes tightly): | dropdown<br>2 — 0-Normal symmetrical movement<br>3 — 1-Minor paralysis (flattened nasolabial fold, asymmetry on smiling)<br>4 — 2-Partial paralysis (total or near paralysis of lower face)<br>5 — 3-Complete paralysis of one or both sides |
| 316 | gs_nihss5l<br><br>Show the field ONLY if:<br>[show_nihss] = '1' | 5l. Motor function - left arm | dropdown<br>2 — 0-Normal (extends arms 90 (or 45) degrees for 10 seconds without drift)<br>3 — 1-Drift<br>4 — 2-Some effort against gravity<br>5 — 3-No effort against gravity<br>6 — 4-No movement<br>7 — U-Untestable (Joint fused or limb amputated) |
| 317 | gs_nihss5r<br><br>Show the field ONLY if:<br>[show_nihss] = '1' | 5r. Motor function - right arm: | dropdown<br>2 — 0-Normal (extends arms 90 (or 45) degrees for 10 seconds without drift)<br>3 — 1-Drift<br>4 — 2-Some effort against gravity<br>5 — 3-No effort against gravity<br>6 — 4-No movement<br>7 — U-Untestable (Joint fused or limb amputated) |

| | | | |
|---|---|---|---|
| 318 | gs_nihss6l<br><br>Show the field ONLY if:<br>[show_nihss] = '1' | 6l. Motor function - left leg: | dropdown<br>2 0-Normal (hold leg 30 degrees position for 5 seconds)<br>3 1-Drift<br>4 2-Some effort against gravity<br>5 3-No effort against gravity<br>6 4-No movement<br>7 U-Untestable (Joint fused or limb amputated) |
| 319 | gs_nihss6r<br><br>Show the field ONLY if:<br>[show_nihss] = '1' | 6r. Motor function - right leg: | dropdown<br>2 0-Normal (hold leg 30 degrees position for 5 seconds)<br>3 1-Drift<br>4 2-Some effort against gravity<br>5 3-No effort against gravity<br>6 4-No movement<br>7 U-Untestable (Joint fused or limb amputated) |
| 320 | gs_nihss7<br><br>Show the field ONLY if:<br>[show_nihss] = '1' | 7. Limb ataxia: | dropdown<br>2 0-No ataxia<br>3 1-Present in one limb<br>4 2-Present in two limbs<br>5 U-Untestable (Joint fused or limb amputated) |
| 321 | gs_nihss8<br><br>Show the field ONLY if:<br>[show_nihss] = '1' | 8. Sensory (use pinprick to test arms, legs, trunk and face - compare side to side): | dropdown<br>2 0-Normal<br>3 1-Mild to moderate decrease in sensation<br>4 2-Severe to total sensory loss |
| 322 | gs_nihss9<br><br>Show the field ONLY if:<br>[show_nihss] = '1' | 9. Best language (describe picture, name items, read sentences): | dropdown<br>2 0-No aphasia<br>3 1-Mild to moderate aphasia<br>4 2-Severe aphasia<br>5 3-Mute |

| | | | |
|---|---|---|---|
| 323 | gs_nihss10<br><br>Show the field ONLY if:<br>[show_nihss] = '1' | 10. Dysarthria (read several words): | dropdown<br>2 \| 0-Normal articulation<br>3 \| 1-Mild to moderate slurring of words<br>4 \| 2-Near unintelligible or unable to speak<br>5 \| U-Intubated or other physical barrier |
| 324 | gs_nihss11<br><br>Show the field ONLY if:<br>[show_nihss] = '1' | 11. Extinction and inattention: | dropdown<br>2 \| 0-Normal<br>3 \| 1-Inattention or extinction to bilateral stimulation in one modality<br>4 \| 2-Severe hemi-inattention or hemi-inattention to multiple modalities |
| 325 | c_nihssobtutd<br><br>Show the field ONLY if:<br>([site] = 'SMCA' or [site] = 'UMCB') and [gs_age] >= 18 and [gs_notadmit] = '2' and [jc_admitdate] <> '' and [gs_discdatetime] <> '' and [jc_princicd10diagnosis] <> '' and [jc_princicd10procedure] <> '' and ([treatment_type(IVtPA)]='1' or [treatment_type(IAtPA)]='1' or [treatment_type(MER)]='1') and [hide_nonessential_fields(1)]='0' | Initial NIHSS Score at Hospital Arrival - UTD<br>*First NIHSS score obtained prior to or after hospital arrival.* | radio<br>1 \| UTD |
| 326 | c_nihssobt<br><br>Show the field ONLY if:<br>([site]='SMCA' or [site]='UMCB') and [gs_age]>=18 and [gs_notadmit]='2' and [jc_admitdate]<>'' and [gs_discdatetime]<>'' and (starts_with([jc_princicd10diagnosis],'i63') and [jc_princicd10procedure]<>'' and ([treatment_type(IVtPA)]='1' or [treatment_type(IAtPA)]='1' or [treatment_type(MER)]='1')) and [c_nihssobtutd]<>'1' and [hide_nonessential_fields(1)]='0' | What is the first NIHSS score obtained prior to or after hospital arrival? | text |

| | | | |
|---|---|---|---|
| 327 | c_nihssperf<br><br>Show the field ONLY if:<br>([site] = 'SMCA' or [site] = 'UMCB') and [gs_age] >= 18 and [gs_notadmit] = '2' and [jc_admitdate] <> '' and [gs_discdatetime] <> '' and [jc_princicd10diagnosis] <> '' and [hide_nonessential_fields(1)]='0' | Is there documentation that an initial NIHSS score was done at this hospital? | radio<br>1 Yes<br>2 No<br><br>Custom alignment: RH |
| 328 | c_nihssdt_precision<br><br>Show the field ONLY if:<br>([site] = 'SMCA' or [site] = 'UMCB') and [gs_age] >= 18 and [gs_notadmit] = '2' and [jc_admitdate] <> '' and [gs_discdatetime] <> '' and [jc_princicd10diagnosis] <> '' and [hide_nonessential_fields(1)]='0' | Initial NIHSS Score Date/Time Precision | dropdown<br>5 Date & Time (MM/DD/YYYY HH24:MM)<br>3 Date Only (MM/DD/YYYY)<br>0 Unknown |
| 329 | c_nihssdt<br><br>Show the field ONLY if:<br>([site] = 'SMCA' or [site] = 'UMCB') and [gs_age] >= 18 and [gs_notadmit] = '2' and [jc_admitdate] <> '' and [gs_discdatetime] <> '' and [jc_princicd10diagnosis] <> '' and [c_nihssdt_precision] <> '0' and [hide_nonessential_fields(1)]='0' | What is the date and time that the NIHSS score was first performed at this hospital?<br>*If Date Only, enter 00:00 for time.* | text (datetime_mdy, Min: 2010-01-01 00:00, Max: 2099-12-31 23:59), Identifier |
| 330 | gs_transfernihssnd<br><br>Show the field ONLY if:<br>([site]='SMCA' or [site]='SMCH' or [site]='SMCW' or [site]='UMCB') and [gs_patientarrival] = '3' and ([treatment_recvd]='1' or [gs_notadmit]<>'1') and [hide_nonessential_fields(1)]='0' | Transfer NIHSS Score - ND<br>*NIHSS score obtained from transferring facility.* | radio<br>1 ND |
| 331 | gs_transfernihss<br><br>Show the field ONLY if:<br>([site]='SMCA' or [site]='SMCH' or [site]='SMCW' or [site]='UMCB') and [gs_patientarrival] = '3' and [gs_transfernihssnd] <> '1' and ([treatment_recvd]='1' or [gs_notadmit]<>'1') and [hide_nonessential_fields(1)]='0' | NIHSS score obtained from transferring facility | text |

| | | | |
|---|---|---|---|
| 332 | gs_initexam<br><br>Show the field ONLY if:<br>([site]='SMCA' or [site]='SMCH' or [site]='SMCW' or [site]='UMCB') and ([treatment_recvd]='1' or [gs_notadmit]<>'1') and [hide_nonessential_fields(1)]='0' | Initial exam findings | checkbox<br>1 \| gs_initexam___1 \| Weakness/Paresis<br>2 \| gs_initexam___2 \| Altered Level of Consciousness<br>3 \| gs_initexam___3 \| Aphasia/Language Disturbance<br>4 \| gs_initexam___4 \| Other neurological signs/symptoms<br>5 \| gs_initexam___5 \| No neurological signs/symptoms<br>6 \| gs_initexam___6 \| ND<br><br>Field Annotation: @NONEOFTHEABOVE='5,6' |
| 333 | gs_ambulatadmit<br><br>Show the field ONLY if:<br>([site]='SMCA' or [site]='SMCH' or [site]='SMCW' or [site]='UMCB') and ([treatment_recvd]='1' or [gs_notadmit]<>'1') and [hide_nonessential_fields(1)]='0' | Ambulatory status on admission | dropdown<br>1 \| Able to ambulate independently (no help from another person) w/ or w/o device<br>2 \| With assistance (from person)<br>3 \| Unable to ambulate<br>4 \| ND |
| 334 | gcs_descriptive<br><br>Show the field ONLY if:<br>([site] = 'SMCA' or [site] = 'UMCB') and ([gs_stroketype] = '4' or [gs_stroketype] = '5') and ([treatment_recvd]='1' or [gs_notadmit]<>'1') and [hide_nonessential_fields(1)]='0' | Section Header: *Hemorrhagic Stroke Scales*<br>First Glasgow Coma Scale (GCS) | descriptive |
| 335 | c_emsgcsnd<br><br>Show the field ONLY if:<br>([site] = 'SMCA' or [site] = 'UMCB') and ([gs_stroketype] = '4' or [gs_stroketype] = '5') and ([treatment_recvd]='1' or [gs_notadmit]<>'1') and [hide_nonessential_fields(1)]='0' | GCS Not Documented | radio<br>1 \| ND |
| 336 | c_emsgcseye<br><br>Show the field ONLY if:<br>([site] = 'SMCA' or [site] = 'UMCB') and ([gs_stroketype] = '4' or [gs_stroketype] = '5') and [c_emsgcsnd] <> '1' and ([treatment_recvd]='1' or [gs_notadmit]<>'1') and [hide_nonessential_fields(1)]='0' | GCS Eye | dropdown<br>2 \| 1<br>3 \| 2<br>4 \| 3<br>5 \| 4 |

| | | | |
|---|---|---|---|
| 337 | c_emsgcsintub<br><br>Show the field ONLY if:<br>([site] = 'SMCA' or [site] = 'UMCB') and ([gs_stroketype] = '4' or [gs_stroketype] = '5') and [c_emsgcsnd] < > '1' and ([treatment_recvd]='1' or [gs_notadmit]<>'1') and [hide_nonessential_fields(1)]='0' | Total GCS: Intubated Comprehensive | radio<br>1 Intubated |
| 338 | c_emsgcsvoic<br><br>Show the field ONLY if:<br>([site] = 'SMCA' or [site] = 'UMCB') and ([gs_stroketype] = '4' or [gs_stroketype] = '5') and [c_emsgcsnd] < > '1' and [c_emsgcsintub] <> '1' and ([treatment_recvd]='1' or [gs_notadmit]<>'1') and [hide_nonessential_fields(1)]='0' | GCS Verbal | dropdown<br>2 1<br>3 2<br>4 3<br>5 4<br>6 5 |
| 339 | c_emsgcsmot<br><br>Show the field ONLY if:<br>([site] = 'SMCA' or [site] = 'UMCB') and ([gs_stroketype] = '4' or [gs_stroketype] = '5') and [c_emsgcsnd] < > '1' and ([treatment_recvd]='1' or [gs_notadmit]<>'1') and [hide_nonessential_fields(1)]='0' | GCS Motor | dropdown<br>2 1<br>3 2<br>4 3<br>5 4<br>6 5<br>7 6 |
| 340 | c_emsgcstot<br><br>Show the field ONLY if:<br>([site] = 'SMCA' or [site] = 'UMCB') and ([gs_stroketype] = '4' or [gs_stroketype] = '5') and [c_emsgcsnd] < > '1' and ([treatment_recvd]='1' or [gs_notadmit]<>'1') and [hide_nonessential_fields(1)]='0' | Total GCS<br>*Add GCS scores together* | text (integer, Min: 3, Max: 15) |
| 341 | c_sahna<br><br>Show the field ONLY if:<br>([site]='SMCA' or [site]='SMCH' or [site]='SMCW' or [site]='UMCB') and [gs_stroketype]='4' and [gs_notadmit]<>'1' and [hide_nonessential_fields(1)]='0' | Section Header: *Subarachnoid Hemorrhage (SAH)*<br><br>Is there documentation any time during the hospital stay that the hemorrhage was non-aneurysmal or due to head trauma? | radio<br>1 Yes<br>2 No<br><br>Custom alignment: RH |

| | | | |
|---|---|---|---|
| 342 | c_sahscale<br><br>Show the field ONLY if:<br>([site]='SMCA' or [site]='SMCH' or [site]='SMCW' or [site]='UMCB') and [gs_stroketype]='4' and [gs_notadmit]<>'1' and [hide_nonessential_fields(1)]='0' | Was an initial Hunt and Hess scale done at this hospital? | radio<br>1 Yes<br>2 No<br><br>Custom alignment: RH |
| 343 | c_sahscale_value<br><br>Show the field ONLY if:<br>[c_sahscale]='1' and [hide_nonessential_fields(1)]='0' | If yes, Hunt and Hess score: | text (integer, Min: 1, Max: 5) |
| 344 | c_sahscaledt_precision<br><br>Show the field ONLY if:<br>[c_sahscale]='1' and [hide_nonessential_fields(1)]='0' | Initial Hunt and Hess Scale Date/Time Precision | dropdown<br>5 Date & Time (MM/DD/YYYY HH24:MM)<br>3 Date Only (MM/DD/YYYY)<br>0 Unknown |
| 345 | c_sahscaledt<br><br>Show the field ONLY if:<br>[c_sahscale]='1' and [c_sahscaledt_precision]<>'0' and [hide_nonessential_fields(1)]='0' | What is the date and time that the Hunt and Hess Scale was first performed at this hospital?<br>*If Date Only, enter 00:00 for time.* | text (datetime_mdy, Min: 2010-01-01 00:00, Max: 2099-12-31 23:59), Identifier |
| 346 | c_wfnssah<br><br>Show the field ONLY if:<br>([site]='SMCA' or [site]='SMCH' or [site]='SMCW' or [site]='UMCB') and [gs_stroketype]='4' and [gs_notadmit]<>'1' and [hide_nonessential_fields(1)]='0' | WFNS SAH Grading Scale | text (integer, Min: 1, Max: 5) |
| 347 | dnv_12_avm<br><br>Show the field ONLY if:<br>([site]='SMCA' or [site]='UMCB') and ([gs_stroketype]='4' or [gs_stroketype]='5') and [gs_notadmit]<>'1' and [hide_nonessential_fields(1)]='0' | Section Header: *Arteriovenous Malformation (AVM)*<br><br>Was this an AVM patient? | yesno<br>1 Yes<br>0 No<br><br>Custom alignment: RH<br>Field Annotation: @HIDDEN |

| | | | |
|---|---|---|---|
| 348 | c_ichscore<br><br>Show the field ONLY if:<br>(([site]='SMCA' or [site]='SMCH' or [site]='SMCW') and [gs_stroketype]='5' and [gs_notadmit]<>'1' and [hide_nonessential_fields(1)]='0') or ([site]='UMCB' and [gs_stroketype]='5') | Section Header: *Intracerebral Hemorrhage (ICH)*<br><br>Was an initial ICH score done at this hospital? | radio<br>1 Yes<br>2 No<br><br>Custom alignment: RH |
| 349 | c_ichscore_value<br><br>Show the field ONLY if:<br>[c_ichscore]='1' and ([site]='UMCB' or [hide_nonessential_fields(1)]='0') | If yes, ICH score: | text (integer, Min: 0, Max: 6) |
| 350 | c_ichscoredt_precision<br><br>Show the field ONLY if:<br>[c_ichscore]='1' and [hide_nonessential_fields(1)]='0' | Initial ICH Score Date/Time Precision | dropdown<br>5 Date & Time (MM/DD/YYYY HH24:MM)<br>3 Date Only (MM/DD/YYYY)<br>0 Unknown |
| 351 | c_ichscoredt<br><br>Show the field ONLY if:<br>[c_ichscore]='1' and [c_ichscoredt_precision]<>'0' and [hide_nonessential_fields(1)]='0' | What is the date and time that the ICH score was first performed at this hospital?<br>*If Date Only, enter 00:00 for time.* | text (datetime_mdy, Min: 2010-01-01 00:00, Max: 2099-12-31 23:59), Identifier |
| 352 | ich_volume_nd<br><br>Show the field ONLY if:<br>[site]='UMCB' | ✪ ICH Volume ND | radio<br>1 ND |
| 353 | ich_volume<br><br>Show the field ONLY if:<br>[site]='UMCB' and [ich_volume_nd]<>'1' | ✪ ICH Volume:<br>*cm3 - 2 decimal places required* | text (number_2dp) |
| 354 | ich_ivh<br><br>Show the field ONLY if:<br>[site]='UMCB' | ✪ IVH: | radio<br>1 Present<br>2 Not Present<br><br>Custom alignment: RH |

<table>
<tr><td>355</td><td>ich_etiology<br><br>Show the field ONLY if:<br>[site]='UMCB'</td><td>✪ Was the ICH etiology documented in the patient medical record?</td><td>radio<table><tr><td>1</td><td>Yes</td></tr><tr><td>2</td><td>No</td></tr></table><br>Custom alignment: RH</td></tr>
<tr><td>356</td><td>ich_etiologydoc<br><br>Show the field ONLY if:<br>[ich_etiology]='1'</td><td>✪ Documented ICH etiology<br>Select all that apply.</td><td>checkbox<table>
<tr><td>1</td><td>ich_etiologydoc___1</td><td>Hypertension</td></tr>
<tr><td>2</td><td>ich_etiologydoc___2</td><td>Anticoagulant-associated (warfarin, DOACs)</td></tr>
<tr><td>3</td><td>ich_etiologydoc___3</td><td>Coagulopathy (liver cirrhosis hemophilia, sickle cell anemia, DIC, thrombocytopenia)</td></tr>
<tr><td>4</td><td>ich_etiologydoc___4</td><td>Vascular malformations (cerebral aneurysms, AVM, dural AV fistula, capillary telangiectasia, cavernous malformation)</td></tr>
<tr><td>5</td><td>ich_etiologydoc___5</td><td>Tumor</td></tr>
<tr><td>6</td><td>ich_etiologydoc___6</td><td>Substance use (cocaine, stimulants)</td></tr>
<tr><td>7</td><td>ich_etiologydoc___7</td><td>Amyloid angiopathy</td></tr>
<tr><td>8</td><td>ich_etiologydoc___8</td><td>Spontaneous/idiopathic</td></tr>
<tr><td>9</td><td>ich_etiologydoc___9</td><td>Cortical vein thrombosis and venous sinus thrombosis</td></tr>
<tr><td>10</td><td>ich_etiologydoc___10</td><td>Other determined cause {ich_etiologydoc_other:icons}</td></tr>
</table></td></tr>
<tr><td>357</td><td>ich_etiologydoc_other<br><br>Show the field ONLY if:<br>[ich_etiologydoc(10)]='1'</td><td>✪ Specify Other Determined Cause</td><td>text<br>Field Annotation: @CHARLIMIT=200</td></tr>
<tr><td>358</td><td>c_funcscore<br><br>Show the field ONLY if:<br>([site]='SMCA' or [site]='SMCH' or [site]='SMCW' or [site]='UMCB') and [gs_stroketype]='5' and [gs_notadmit]<>'1' and [hide_nonessential_fields(1)]='0'</td><td>FUNC Score (ICH)</td><td>text</td></tr>
</table>

| | | | |
|---|---|---|---|
| 359 | dnv_12_sm_doc<br><br>Show the field ONLY if:<br>([site]='SMCA' or [site]='SMCH' or [site]='SMCW' or [site]='UMCB') and [gs_stroketype]='5' and [gs_notadmit]<>'1' and [hide_nonessential_fields(1)]='0' | Was a Spetzler-Martin score documented at this hospital? | yesno<br>1 \| Yes<br>0 \| No<br><br>Custom alignment: RH |
| 360 | dnv_12_sm_score<br><br>Show the field ONLY if:<br>[dnv_12_sm_doc]='1' and [hide_nonessential_fields(1)]='0' | If yes, Spetzler-Martin score: | text (integer, Min: 1, Max: 6)<br>Custom alignment: RH |
| 361 | gs_nopriormeds<br><br>Show the field ONLY if:<br>([site]='SMCA' or [site]='SMCH' or [site]='SMCW' or [site]='UMCB') and ([treatment_recvd]='1' or [gs_notadmit]<>'1') | Section Header: *Medication Prior to Admission*<br>No medications prior to admission | radio<br>1 \| No prior medications<br><br>Field Annotation: @HIDDEN |
| 362 | gs_priorantithrom<br><br>Show the field ONLY if:<br>([site]='SMCA' or [site]='SMCH' or [site]='SMCW' or [site]='UMCB') and ([treatment_recvd]='1' or [gs_notadmit]<>'1') | Antiplatelet or Anticoagulant medication(s) | radio<br>1 \| Yes<br>2 \| No/ND<br><br>Custom alignment: RH |
| 363 | gs_priorantithrom1class<br><br>Show the field ONLY if:<br>[gs_priorantithrom]='1' | Prior Antithrombotic 1 - Antithrombotic Class | radio<br>1 \| Antiplatelet<br>2 \| Anticoagulant<br><br>Custom alignment: RH |
| 364 | gs_priorantithrom1a<br><br>Show the field ONLY if:<br>[gs_priorantithrom1class] = '1' | Prior Antithrombotic 1 - Antiplatelet | dropdown<br>3 \| aspirin<br>4 \| ASA/dipyridamole (Aggrenox)<br>6 \| clopidogrel (Plavix)<br>20 \| prasugrel (Effient)<br>21 \| ticagrelor (Brilinta)<br>7 \| ticlopidine (Ticlid)<br>11 \| Other Antiplatelet |

| | | | |
|---|---|---|---|
| 365 | gs_priorantithrom1b<br><br>Show the field ONLY if:<br>[gs_priorantithrom1class] = '2' | Prior Antithrombotic 1 - Anticoagulant | dropdown<br>19 apixaban (Eliquis)<br>15 argatroban<br>16 dabigatran (Pradaxa)<br>17 desirudin (Iprivask)<br>22 edoxaban (Savaysa)<br>12 fondaparinux (Arixtra)<br>9 full dose LMW heparin<br>18 lepirudin (Refludan)<br>14 rivaroxaban (Xarelto)<br>8 Unfractionated heparin IV<br>5 warfarin (Coumadin)<br>13 Other Anticoagulant |
| 366 | gs_priorantithrom2class<br><br>Show the field ONLY if:<br>[gs_priorantithrom1class] <> '' | Prior Antithrombotic 2 - Antithrombotic Class | radio<br>1 Antiplatelet<br>2 Anticoagulant<br><br>Custom alignment: RH |
| 367 | gs_priorantithrom2a<br><br>Show the field ONLY if:<br>[gs_priorantithrom2class] = '1' | Prior Antithrombotic 2 - Antiplatelet | dropdown<br>3 aspirin<br>4 ASA/dipyridamole (Aggrenox)<br>6 clopidogrel (Plavix)<br>20 prasugrel (Effient)<br>21 ticagrelor (Brilinta)<br>7 ticlopidine (Ticlid)<br>11 Other Antiplatelet |

| | | | |
|---|---|---|---|
| 368 | gs_priorantithrom2b<br><br>Show the field ONLY if:<br>[gs_priorantithrom2class] = '2' | Prior Antithrombotic 2 - Anticoagulant | dropdown<br>19 \| apixaban (Eliquis)<br>15 \| argatroban<br>16 \| dabigatran (Pradaxa)<br>17 \| desirudin (Iprivask)<br>22 \| edoxaban (Savaysa)<br>12 \| fondaparinux (Arixtra)<br>9 \| full dose LMW heparin<br>18 \| lepirudin (Refludan)<br>14 \| rivaroxaban (Xarelto)<br>8 \| Unfractionated heparin IV<br>5 \| warfarin (Coumadin)<br>13 \| Other Anticoagulant |
| 369 | gs_priorantithrom3class<br><br>Show the field ONLY if:<br>[gs_priorantithrom2class] <> '' | Prior Antithrombotic 3 - Antithrombotic Class | radio<br>1 \| Antiplatelet<br>2 \| Anticoagulant<br><br>Custom alignment: RH |
| 370 | gs_priorantithrom3a<br><br>Show the field ONLY if:<br>[gs_priorantithrom3class] = '1' | Prior Antithrombotic 3 - Antiplatelet | dropdown<br>3 \| aspirin<br>4 \| ASA/dipyridamole (Aggrenox)<br>6 \| clopidogrel (Plavix)<br>20 \| prasugrel (Effient)<br>21 \| ticagrelor (Brilinta)<br>7 \| ticlopidine (Ticlid)<br>11 \| Other Antiplatelet |

| | | | |
|---|---|---|---|
| 371 | gs_priorantithrom3b<br><br>Show the field ONLY if:<br>[gs_priorantithrom3class] = '2' | Prior Antithrombotic 3 - Anticoagulant | dropdown<br>19 apixaban (Eliquis)<br>15 argatroban<br>16 dabigatran (Pradaxa)<br>17 desirudin (Iprivask)<br>22 edoxaban (Savaysa)<br>12 fondaparinux (Arixtra)<br>9 full dose LMW heparin<br>18 lepirudin (Refludan)<br>14 rivaroxaban (Xarelto)<br>8 Unfractionated heparin IV<br>5 warfarin (Coumadin)<br>13 Other Anticoagulant |
| 372 | gs_antihyperprior<br><br>Show the field ONLY if:<br>([site]='SMCA' or [site]='SMCH' or [site]='SMCW' or [site]='UMCB') and ([treatment_recvd]='1' or [gs_notadmit]<>'1') and [hide_nonessential_fields(1)]='0' | Antihypertensive prior to admission? | radio<br>1 Yes<br>2 No/ND<br><br>Custom alignment: RH |
| 373 | gs_cholredprior<br><br>Show the field ONLY if:<br>([site]='SMCA' or [site]='SMCH' or [site]='SMCW' or [site]='UMCB') and ([treatment_recvd]='1' or [gs_notadmit]<>'1') | Cholesterol reducer prior to admission? | radio<br>1 Yes<br>2 No/ND<br><br>Custom alignment: RH |
| 374 | gs_dmmedprior<br><br>Show the field ONLY if:<br>([site]='SMCA' or [site]='SMCH' or [site]='SMCW' or [site]='UMCB') and ([treatment_recvd]='1' or [gs_notadmit]<>'1') | Anti-hyperglycemic medications prior to admission? | radio<br>1 Yes<br>2 No/ND<br><br>Custom alignment: RH |

| | | | |
|---|---|---|---|
| 375 | dmmedspecadm<br><br>Show the field ONLY if:<br>[gs_dmmedprior] = '1' | If yes, select medications<br>*Select all that apply.* | checkbox<br>1 dmmedspecadm___1 DPP-4 Inhibitors<br>2 dmmedspecadm___2 GLP-1 receptor agonist<br>3 dmmedspecadm___3 Insulin<br>4 dmmedspecadm___4 Metformin<br>5 dmmedspecadm___5 SLGT2 Inhibitor<br>6 dmmedspecadm___6 Sulfonyurea<br>7 dmmedspecadm___7 Thiazolidinedione<br>8 dmmedspecadm___8 Other oral agents<br>9 dmmedspecadm___9 Other injectable/subcutaneous agents |
| 376 | prior_antidepmed<br><br>Show the field ONLY if:<br>([site]='SMCA' or [site]='SMCH' or [site]='SMCW' or [site]='UMCB') and ([treatment_recvd]='1' or [gs_notadmit]<>'1') and [hide_nonessential_fields(1)]='0' | Antidepressant medication | radio<br>1 Yes<br>2 No/ND<br><br>Custom alignment: RH |
| 377 | admission_complete | Section Header: *Form Status*<br><br>Complete? | dropdown<br>0 Incomplete<br>1 Unverified<br>2 Complete |

Instrument: **Hospitalization** (hospitalization)

Collapse

| | | | |
|---|---|---|---|
| 378 | desc_lastknownwell | Section Header: *Symptom Timeline*<br><br>Date/Time patient last known to be well:[gs_lastknownwell]From: Initial Stroke Information | descriptive |
| 379 | desc_gs_symptomdatetime | Date/Time of discovery of stroke or stroke-like symptoms:[gs_symptomdatetime]From: Initial Stroke Information | descriptive |
| 380 | desc_onsetcom<br><br>Show the field ONLY if:<br>([treatment_recvd]='1' or [gs_notadmit]<>'1') | Comments:[gs_pc_onsetcom]From: Initial Stroke Information | descriptive |
| 381 | desc_gs_ctcomp<br><br>Show the field ONLY if:<br>([treatment_recvd]='1' or [gs_notadmit]<>'1') | Section Header: *Brain Imaging*<br><br>Brain imaging completed at your hospital for this episode of care?[gs_ctcomp]From: Initial Stroke Information | descriptive |

| | | | |
|---|---|---|---|
| 382 | desc_gs_ctcompdatetime<br><br>Show the field ONLY if:<br>[gs_ctcomp] = '1' and ([treatment_recvd]='1' or [gs_notadmit]<>'1') | Date/Time Brain Imaging Initiated:[gs_ctcompdatetime]From: Initial Stroke Information | descriptive |
| 383 | gs_firstimage<br><br>Show the field ONLY if:<br>([site]='SMCA' or [site]='SMCH' or [site]='SMCW' or [site]='UMCB') and ([treatment_recvd]='1' or [gs_notadmit]<>'1') and [hide_nonessential_fields(1)]='0' | Interpretation of first brain image after symptom onset, done at any facility: | radio<br>1 \| Acute Hemorrhage<br>2 \| No Acute Hemorrhage<br>3 \| Not available |
| 384 | mer_vasc<br><br>Show the field ONLY if:<br>([site]='SMCA' or [site]='SMCH' or [site]='SMCW' or [site]='UMCB') and [gs_stroketype]='2' | Was Acute Vascular or perfusion imaging (e.g. CTA, MRA, DSA) performed at your hospital? | radio<br>1 \| Yes<br>2 \| No<br><br>Custom alignment: RH |
| 385 | mer_imagecdt_precision<br><br>Show the field ONLY if:<br>[mer_vasc]='1' and [hide_nonessential_fields(1)]='0' | Date/Time 1st vessel or perfusion imaging initiated at your hospital Precision | dropdown<br>5 \| Date & Time (MM/DD/YYYY HH24:MM)<br>3 \| Date Only (MM/DD/YYYY)<br>0 \| Unknown |
| 386 | mer_imagecdt<br><br>Show the field ONLY if:<br>[mer_vasc]='1' and [hide_nonessential_fields(1)]='0' | Date/Time 1st vessel or perfusion imaging initiated at your hospital | text (datetime_mdy, Min: 2010-01-01 00:00, Max: 2099-12-31 23:59) |
| 387 | mer_vascper<br><br>Show the field ONLY if:<br>[mer_vasc]='1' and [hide_nonessential_fields(1)]='0' | If yes, type of imaging<br>*Select all that apply.* | checkbox<br>1 \| mer_vascper___1 \| CTA<br>2 \| mer_vascper___2 \| CT Perfusion<br>3 \| mer_vascper___3 \| MRA<br>4 \| mer_vascper___4 \| MR Perfusion<br>5 \| mer_vascper___5 \| DSA (catheter angiography)<br>0 \| mer_vascper___0 \| Image type not documented<br><br>Field Annotation: @NONEOFTHEABOVE='0' |

| | | | |
|---|---|---|---|
| 388 | mer_targles<br><br>Show the field ONLY if:<br>([site]='SMCA' or [site]='SMCH' or [site]='SMCW' or [site]='UMCB') and [gs_stroketype]='2' | Was a target lesion (large vessel occlusion) visualized? | radio<br>1 \| Yes<br>2 \| No/ND<br><br>Custom alignment: RH |
| 389 | mer_vessels<br><br>Show the field ONLY if:<br>[mer_targles] = '1' | If yes, select site of occlusion: | checkbox<br>1 \| mer_vessels___1 \| ICA<br>2 \| mer_vessels___2 \| MCA<br>3 \| mer_vessels___3 \| Basilar Artery<br>4 \| mer_vessels___4 \| Other Cerebral Artery Branch<br>5 \| mer_vessels___5 \| Vertebral Artery |
| 390 | mer_ica<br><br>Show the field ONLY if:<br>[mer_vessels(1)] = '1' | MER ICA | checkbox<br>1 \| mer_ica___1 \| Intracranial ICA<br>2 \| mer_ica___2 \| Cervical ICA<br>3 \| mer_ica___3 \| Other/UTD |
| 391 | mer_mca<br><br>Show the field ONLY if:<br>[mer_vessels(2)] = '1' | MER MCA | checkbox<br>1 \| mer_mca___1 \| M1<br>2 \| mer_mca___2 \| M2<br>3 \| mer_mca___3 \| Other/UTD |
| 392 | timetracker<br><br>Show the field ONLY if:<br>([treatment_recvd]='1' or [gs_notadmit]<>'1') | Section Header: *Time Tracker Fields*<br><br>Show Time Tracker Fields | checkbox<br>1 \| timetracker___1 \| Show<br><br>Custom alignment: RH |
| 393 | desc_gs_jp_teamna<br><br>Show the field ONLY if:<br>[timetracker(1)] = '1' | Stroke Team:[gs_jp_teamna]From: Initial Stroke Information(Blank if not marked NA) | descriptive |
| 394 | desc_gs_jp_teamdatetime<br><br>Show the field ONLY if:<br>[timetracker(1)] = '1' and [gs_jp_teamna] <> '1' | Date/Time Stroke Team Activated:[gs_jp_teamdatetime]From: Initial Stroke Information | descriptive |
| 395 | desc_gs_jp_arrivdatetime<br><br>Show the field ONLY if:<br>[timetracker(1)] = '1' and [gs_jp_teamna] <> '1' | Date/Time Stroke Team Arrived:[gs_jp_arrivdatetime]From: Initial Stroke Information | descriptive |

| | | | |
|---|---|---|---|
| 396 | desc_gs_jp_edphysna<br><br>Show the field ONLY if:<br>[timetracker(1)] = '1' | ED Physician Assessment:[gs_jp_edphysna]From: Initial Stroke Information(Blank if not marked NA) | descriptive |
| 397 | desc_gs_jp_eddatetime<br><br>Show the field ONLY if:<br>[timetracker(1)] = '1' and [gs_jp_edphysna] <> '1' | Date/Time of ED Physician Assessment:[gs_jp_eddatetime]From: Initial Stroke Information | descriptive |
| 398 | desc_gs_jp_neurosurgna<br><br>Show the field ONLY if:<br>[timetracker(1)] = '1' | Neurosurgical Services Consulted:[gs_jp_neurosurgna]From: Initial Stroke Information(Blank if not marked NA) | descriptive |
| 399 | desc_gs_jp_nsurgdatetime<br><br>Show the field ONLY if:<br>[timetracker(1)] = '1' and [gs_jp_neurosurgna] <> '1' | Date/Time Neurosurgical Services Consulted:[gs_jp_nsurgdatetime]From: Initial Stroke Information | descriptive |
| 400 | desc_gs_jp_ctna<br><br>Show the field ONLY if:<br>[timetracker(1)] = '1' | Brain Imaging:[gs_jp_ctna]From: Initial Stroke Information(Blank if not marked NA) | descriptive |
| 401 | desc_gs_jp_ctorddatetime<br><br>Show the field ONLY if:<br>[timetracker(1)] = '1' and [gs_jp_ctna] <> '1' | Date/Time Brain Imaging Ordered:[gs_jp_ctorddatetime]From: Initial Stroke Information | descriptive |
| 402 | desc_gs_jp_ctrepdatetime<br><br>Show the field ONLY if:<br>[timetracker(1)] = '1' and [gs_jp_ctna] <> '1' | Date/Time Brain Imaging Interpreted:[gs_jp_ctrepdatetime]From: Initial Stroke Information | descriptive |
| 403 | desc_gs_ivtpaordered_na<br><br>Show the field ONLY if:<br>[timetracker(1)] = '1' | IV t-PA Orders:[gs_ivtpaordered_na]From: Initial Stroke Information(Blank if not marked NA) | descriptive |
| 404 | desc_gs_ivtpaordered<br><br>Show the field ONLY if:<br>[timetracker(1)] = '1' and [gs_ivtpaordered_na] <> '1' | Date/Time IV t-PA Ordered:[gs_ivtpaordered]From: Initial Stroke Information | descriptive |
| 405 | desc_gs_jp_labna<br><br>Show the field ONLY if:<br>[timetracker(1)] = '1' | Labs:[gs_jp_labna]From: Initial Stroke Information(Blank if not marked NA) | descriptive |

| | | | |
|---|---|---|---|
| 406 | desc_gs_jp_laborddatetime<br><br>Show the field ONLY if:<br>[timetracker(1)] = '1' and [gs_jp_labna] <> '1' | Date/Time Lab Tests Ordered:[gs_jp_laborddatetime]From: Initial Stroke Information | descriptive |
| 407 | desc_gs_jp_labcpdatetime<br><br>Show the field ONLY if:<br>[timetracker(1)] = '1' and [gs_jp_labna] <> '1' | Date/Time Lab Tests Completed:[gs_jp_labcpdatetime]From: Initial Stroke Information | descriptive |
| 408 | desc_gs_jp_ecgna<br><br>Show the field ONLY if:<br>[timetracker(1)] = '1' | ECG:[gs_jp_ecgna]From: Initial Stroke Information(Blank if not marked NA) | descriptive |
| 409 | desc_gs_jp_ecgorddatetime<br><br>Show the field ONLY if:<br>[timetracker(1)] = '1' and [gs_jp_ecgna] <> '1' | Date/Time ECG Ordered:[gs_jp_ecgorddatetime]From: Initial Stroke Information | descriptive |
| 410 | desc_gs_jp_ecgcpdatetime<br><br>Show the field ONLY if:<br>[timetracker(1)] = '1' and [gs_jp_ecgna] <> '1' | Date/Time ECG Completed:[gs_jp_ecgcpdatetime]From: Initial Stroke Information | descriptive |
| 411 | desc_gs_jp_xrayna<br><br>Show the field ONLY if:<br>[timetracker(1)] = '1' | Chest X-ray:[gs_jp_xrayna]From: Initial Stroke Information(Blank if not marked NA) | descriptive |
| 412 | desc_gs_jp_xorderdatetime<br><br>Show the field ONLY if:<br>[timetracker(1)] = '1' and [gs_jp_xrayna] <> '1' | Date/Time Chest X-ray Ordered:[gs_jp_xorderdatetime]From: Initial Stroke Information | descriptive |
| 413 | desc_gs_jp_xcompdatetime<br><br>Show the field ONLY if:<br>[timetracker(1)] = '1' and [gs_jp_xrayna] <> '1' | Date/Time Chest X-ray Completed:[gs_jp_xcompdatetime]From: Initial Stroke Information | descriptive |
| 414 | desc_gs_jp_jcahocomm<br><br>Show the field ONLY if:<br>[timetracker(1)] = '1' | Additional comments:[gs_jp_jcahocomm]From: Initial Stroke Information | descriptive |

| | | | |
|---|---|---|---|
| 415 | gs_ivthroinit<br><br>Show the field ONLY if:<br>[gs_stroketype] <> '4' and [gs_stroketype] <> '5' and ([treatment_recvd]='1' or [gs_notadmit]<>'1') | Section Header: *IV Thrombolytic TherapyDid this patient receive an Acute Stroke Treatment?[treatment_recvd]*<br><br>✪ IV Thrombolytic initiated at this hospital? | radio<br>1 Yes<br>2 No<br><br>Custom alignment: RH<br>Field Annotation: @DEFAULT='[treatment_type(IVtPA):value]' |
| 416 | desc_ivthrodttm_precision<br><br>Show the field ONLY if:<br>[gs_ivthroinit] = '1' | IV Thrombolytic Initiation Date/Time Precision[ivthrodttm_precision]From: Initial Stroke Information | descriptive |
| 417 | jc_ivthrodttm_precision<br><br>Show the field ONLY if:<br>[gs_ivthroinit] = '1' | ✪ IV Thrombolytic Initiation Date/Time Precision | dropdown<br>5 Date & Time (MM/DD/YYYY HH24:MM)<br>3 Date Only (MM/DD/YYYY)<br>0 Unknown<br><br>Field Annotation: @DEFAULT='[ivthrodttm_precision:value]' |
| 418 | desc_ivthrodttm<br><br>Show the field ONLY if:<br>[gs_ivthroinit] = '1' and [jc_ivthrodttm_precision] <> '0' | IV Thrombolytic Initiation Date/Time[ivthrodttm]From: Initial Stroke Information▲ ▼ copy the date/time into the field below | descriptive, Identifier |
| 419 | jc_ivthrodttm<br><br>Show the field ONLY if:<br>[gs_ivthroinit] = '1' and [jc_ivthrodttm_precision] <> '0' | ✪ IV Thrombolytic Initiation Date/Time | text (datetime_mdy, Min: 2010-01-01 00:00, Max: 2099-12-31 23:59), Identifier<br>Field Annotation: @DEFAULT='[ivthrodttm]' |
| 420 | gs_thrombo<br><br>Show the field ONLY if:<br>[gs_ivthroinit] = '1' | ✪ Thrombolytic Used | radio<br>1 Alteplase (Class 1 evidence)<br>2 Tenecteplase (Class 2b evidence)<br><br>Field Annotation: @DEFAULT='[iv_thrombolytic_type:value]' |
| 421 | gs_altedosend<br><br>Show the field ONLY if:<br>[gs_thrombo]='1' | Alteplase Dose Not Documented | radio<br>1 Alteplase dose ND |
| 422 | gs_altotaldose<br><br>Show the field ONLY if:<br>[gs_thrombo]='1' and [gs_altedosend]<>'1' | Alteplase Total Dose: (mg)<br>*1 Decimal Place Required (GWTG)* | text (number_1dp) |

| | | | |
|---|---|---|---|
| 423 | gs_tenecdosend<br><br>Show the field ONLY if:<br>[gs_thrombo]='2' | ✪ Tenecteplase Dose Not Documented | radio<br>1 \| Tenecteplase dose ND |
| 424 | gs_tenectotaldose<br><br>Show the field ONLY if:<br>[gs_thrombo]='2' and [gs_tenecdo send]<>'1' | ✪ Tenecteplase Total Dose: (mg)<br>*1 Decimal Place Required (GWTG)* | text (number_1dp)<br>Field Annotation: @DEFAULT='[tnk_dose_given]' |
| 425 | gs_tenecreason<br><br>Show the field ONLY if:<br>[gs_thrombo]='2' | Reason for selecting Tenecteplase instead of Alteplase: | radio<br>1 \| Large Vessel Occlusion (LVO) with potential thrombectomy<br>2 \| Mild Stroke<br>3 \| Other |
| 426 | gs_tenecreason_ot<br><br>Show the field ONLY if:<br>[gs_tenecreason] = '3' | Reason for selecting Tenecteplase instead of Alteplase - Other | text<br>Field Annotation: @CHARLIMIT=200 |
| 427 | gs_thrombimg<br><br>Show the field ONLY if:<br>[gs_ivthroinit]='1' and [calc_lkw_to _ivtpa]>=270 | If IV Thrombolytic administered beyond 4.5-hour, was imaging used to identify eligibility? | radio<br>1 \| Yes, Diffusion-FLAIR mismatch<br>2 \| Yes, Core-Perfusion mismatch<br>3 \| None<br>4 \| Other |
| 428 | gs_thrombimg_ot<br><br>Show the field ONLY if:<br>[gs_thrombimg] = '4' | IV thrombolytic beyond 4.5-hour, imaging used to identify eligibility - Other | text<br>Field Annotation: @CHARLIMIT=200 |
| 429 | gs_ivtpanc<br><br>Show the field ONLY if:<br>[gs_stroketype] <> '4' and [gs_stro ketype] <> '5' | Documented exclusions or relative exclusions (contraindications or warnings) for not initiating IV thrombolytic in the 0-3 hr treatment window? | radio<br>1 \| Yes<br>2 \| No<br><br>Custom alignment: RH |

| | | | |
|---|---|---|---|
| 430 | gs_ivtpalatenc<br><br>Show the field ONLY if:<br>([site]='SMCA' or [site]='SMCH' or [site]='SMCW' or [site]='UMCB') and [gs_stroketype] <> '4' and [gs_stroketype] <> '5' and ([gs_ivthroinit] <> '1' or [jc_ivthrodttm] = '' or [gs_lastknownwell] = '' or datediff([gs_lastknownwell], [jc_ivthrodttm], 'm', 'mdy', true) > 180) | Documented exclusions or relative exclusions (contraindications or warnings) for not initiating IV thrombolytic in the 3-4.5 hr treatment window? | radio<br>1 Yes<br>2 No<br><br>Custom alignment: RH |
| 431 | gs_exclusion<br><br>Show the field ONLY if:<br>([site]='SMCA' or [site]='SMCH' or [site]='SMCW' or [site]='UMCB') and [gs_ivtpanc] = '1' | Section Header: *Documented Exclusion CriteriaFor discharge on or after April 1, 2016*<br><br>Exclusion Criteria (contraindications) 0-3 hr treatment window<br>*Select all that apply.* | checkbox<br>1 gs_exclusion___1 C1: Elevated blood pressure (systolic > 185 mm Hg or diastolic > 110 mm Hg) despite treatment<br>2 gs_exclusion___2 C2: Recent intracranial or spinal surgery or significant head trauma, or prior stroke in previous 3 months<br>3 gs_exclusion___3 C3: History of previous intracranial hemorrhage, intracranial neoplasm, arteriovenous malformation, or aneurysm<br>4 gs_exclusion___4 C4: Active internal bleeding<br>5 gs_exclusion___5 C5: Acute bleeding diathesis (low platelet count, increased PTT, INR >= 1.7 or use of NOAC)<br>6 gs_exclusion___6 C6: Symptoms suggest subarachnoid hemorrhage<br>7 gs_exclusion___7 C7: CT demonstrates multilobar infarction (hypodensity >1/3 cerebral hemisphere)<br>8 gs_exclusion___8 C8: Arterial puncture at noncompressible site in previous 7 days<br>9 gs_exclusion___9 C9: Blood glucose concentration < 50 mg/dL (2.7 mmol/L) |

| | | | |
|---|---|---|---|
| 432 | gs_relexclusion<br><br>Show the field ONLY if:<br>([site]='SMCA' or [site]='SMCH' or [site]='SMCW' or [site]='UMCB') and [gs_ivtpanc] = '1' | Relative Exclusion Criteria (Warnings) 0-3 hr treatment window<br>*Select all that apply.* | checkbox<br>1 \| gs_relexclusion___1 \| W1: Care-team unable to determine eligibility<br>2 \| gs_relexclusion___2 \| W2: IV or IA thrombolysis/thrombectomy at an outside hospital prior to arrival<br>3 \| gs_relexclusion___3 \| W3: Life expectancy < 1 year or severe co-morbid illness or CMO on admission<br>4 \| gs_relexclusion___4 \| W4: Pregnancy<br>5 \| gs_relexclusion___5 \| W5: Patient/family refusal<br>6 \| gs_relexclusion___6 \| W6: Rapid improvement<br>7 \| gs_relexclusion___7 \| W7: Stroke severity too mild (non-disabling)<br>8 \| gs_relexclusion___8 \| W8: Recent acute myocardial infarction (within previous 3 months)<br>9 \| gs_relexclusion___9 \| W9: Seizure at onset with postictal residual neurological impairments<br>10 \| gs_relexclusion___10 \| W10: Major surgery or serious trauma within previous 14 days<br>11 \| gs_relexclusion___11 \| W11: Recent gastrointestinal or urinary tract hemorrhage (within previous 21 days)<br><br>Field Annotation: @HIDECHOICE='6' |

| | | | |
|---|---|---|---|
| 433 | gs_exclusion2<br><br>Show the field ONLY if:<br>([site]='SMCA' or [site]='SMCH' or [site]='SMCW' or [site]='UMCB') and [gs_ivtpalatenc] = '1' | Exclusion Criteria (contraindications) 3-4.5 hr treatment window<br>*Select all that apply.* | checkbox<br>1 \| gs_exclusion2___1 \| C1: Elevated blood pressure (systolic > 185 mm Hg or diastolic > 110 mm Hg) despite treatment<br>2 \| gs_exclusion2___2 \| C2: Recent intracranial or spinal surgery or significant head trauma, or prior stroke in previous 3 months<br>3 \| gs_exclusion2___3 \| C3: History of previous intracranial hemorrhage, intracranial neoplasm, arteriovenous malformation, or aneurysm<br>4 \| gs_exclusion2___4 \| C4: Active internal bleeding<br>5 \| gs_exclusion2___5 \| C5: Acute bleeding diathesis (low platelet count, increased PTT, INR >= 1.7 or use of NOAC)<br>6 \| gs_exclusion2___6 \| C6: Symptoms suggest subarachnoid hemorrhage<br>7 \| gs_exclusion2___7 \| C7: CT demonstrates multilobar infarction (hypodensity >1/3 cerebral hemisphere)<br>8 \| gs_exclusion2___8 \| C8: Arterial puncture at noncompressible site in previous 7 days<br>9 \| gs_exclusion2___9 \| C9: Blood glucose concentration < 50 mg/dL (2.7 mmol/L) |

| | | | |
|---|---|---|---|
| 434 | gs_relexclusion2<br><br>Show the field ONLY if:<br>([site]='SMCA' or [site]='SMCH' or [site]='SMCW' or [site]='UMCB') and [gs_ivtpalatenc] = '1' | Relative Exclusion Criteria (Warnings) 3-4.5 hr treatment window<br>*Select all that apply.* | checkbox<br>1 \| gs_relexclusion2___1 \| W1: Care-team unable to determine eligibility<br>2 \| gs_relexclusion2___2 \| W2: IV or IA thrombolysis/thrombectomy at an outside hospital prior to arrival<br>3 \| gs_relexclusion2___3 \| W3: Life expectancy < 1 year or severe co-morbid illness or CMO on admission<br>4 \| gs_relexclusion2___4 \| W4: Pregnancy<br>5 \| gs_relexclusion2___5 \| W5: Patient/family refusal<br>6 \| gs_relexclusion2___6 \| W6: Rapid improvement<br>7 \| gs_relexclusion2___7 \| W7: Stroke severity too mild (non-disabling)<br>8 \| gs_relexclusion2___8 \| W8: Recent acute myocardial infarction (within previous 3 months)<br>9 \| gs_relexclusion2___9 \| W9: Seizure at onset with postictal residual neurological impairments<br>10 \| gs_relexclusion2___10 \| W10: Major surgery or serious trauma within previous 14 days<br>11 \| gs_relexclusion2___11 \| W11: Recent gastrointestinal or urinary tract hemorrhage (within previous 21 days)<br><br>Field Annotation: @HIDECHOICE='6' |
| 435 | gs_addrelexcl<br><br>Show the field ONLY if:<br>([site]='SMCA' or [site]='SMCH' or [site]='SMCW' or [site]='UMCB') and [gs_ivtpalatenc] = '1' | Additional Relative Exclusion Criteria 3-4.5 hr treatment window<br>*Select all that apply.* | checkbox<br>1 \| gs_addrelexcl___1 \| AW1: Age > 80<br>2 \| gs_addrelexcl___2 \| AW2: History of both diabetes and prior ischemic stroke<br>3 \| gs_addrelexcl___3 \| AW3: Taking an oral anticoagulant regardless of INR<br>4 \| gs_addrelexcl___4 \| AW4: Severe stroke (NIHSS > 25) |

| | | | |
|---|---|---|---|
| 436 | gs_noivhosp<br><br>Show the field ONLY if:<br>([site]='SMCA' or [site]='SMCH' or [site]='SMCW' or [site]='UMCB') and [gs_stroketype] <> '4' and [gs_stroketype] <> '5' and ([treatment_recvd]='1' or [gs_notadmit]<>'1') | Section Header: *Hospital-related or Other Factors*<br><br>Other Reasons (Hospital-related or other factors) 0-3 hr treatment window.<br>*Select all that apply.* | checkbox<br>6 \| gs_noivhosp___6 \| Delay in Patient Arrival<br>3 \| gs_noivhosp___3 \| Inhospital Time Delay<br>2 \| gs_noivhosp___2 \| Delay in Stroke diagnosis<br>4 \| gs_noivhosp___4 \| No IV access<br>9 \| gs_noivhosp___9 \| Rapid or Early Improvement<br>7 \| gs_noivhosp___7 \| Advanced Age<br>8 \| gs_noivhosp___8 \| Stroke too severe<br>5 \| gs_noivhosp___5 \| Other |
| 437 | gs_noivthromoth<br><br>Show the field ONLY if:<br>([site]='SMCA' or [site]='SMCH' or [site]='SMCW' or [site]='UMCB') and [gs_noivhosp(5)] = '1' | Specify other reason(s) for no IV thrombolytic in 0-3 hr treatment window. | text<br>Field Annotation: @CHARLIMIT=200 |
| 438 | gs_noivhosp2<br><br>Show the field ONLY if:<br>([site]='SMCA' or [site]='SMCH' or [site]='SMCW' or [site]='UMCB') and [gs_stroketype] <> '4' and [gs_stroketype] <> '5' and ([treatment_recvd]='1' or [gs_notadmit]<>'1') | Other Reasons (Hospital-related or other factors) 3-4.5 hr treatment window.<br>*Select all that apply.* | checkbox<br>6 \| gs_noivhosp2___6 \| Delay in Patient Arrival<br>3 \| gs_noivhosp2___3 \| Inhospital Time Delay<br>2 \| gs_noivhosp2___2 \| Delay in Stroke diagnosis<br>4 \| gs_noivhosp2___4 \| No IV access<br>9 \| gs_noivhosp2___9 \| Rapid or Early Improvement<br>7 \| gs_noivhosp2___7 \| Advanced Age<br>8 \| gs_noivhosp2___8 \| Stroke too severe<br>5 \| gs_noivhosp2___5 \| Other<br><br>Field Annotation: @HIDECHOICE='7,8' |
| 439 | gs_noivthromoth2<br><br>Show the field ONLY if:<br>([site]='SMCA' or [site]='SMCH' or [site]='SMCW' or [site]='UMCB') and [gs_noivhosp2(5)] = '1' | Specify other reason(s) for no IV thrombolytic in 3-4.5 hr treatment window. | text<br>Field Annotation: @CHARLIMIT=200 |

| | | | |
|---|---|---|---|
| 440 | ivtpadelay<br><br>Show the field ONLY if:<br>([site]='SMCA' or [site]='SMCH' or [site]='SMCW' or [site]='UMCB') and [gs_symptomlocation]<>'4' and [gs_ivthroinit]='1' and [calc_door_to_ivtpa]>60 | Section Header: *Cause for IV Thrombolytic delay[treatment_delays]From: Initial Stroke Information*<br><br>If IV Thrombolytic was initiated greater than 60 minutes after hospital arrival, were Eligibility or Medical reason(s) documented as the cause for delay? | radio<br>1 \| Yes<br>2 \| No<br><br>Custom alignment: RH |
| 441 | ivtpadelay45<br><br>Show the field ONLY if:<br>([site]='SMCA' or [site]='SMCH' or [site]='SMCW' or [site]='UMCB') and [gs_symptomlocation]<>'4' and [gs_ivthroinit]='1' and [calc_door_to_ivtpa]>45 | If IV Thrombolytic was initiated greater than 45 minutes after hospital arrival, were Eligibility or Medical reason(s) documented as the cause for delay? | radio<br>1 \| Yes<br>2 \| No<br><br>Custom alignment: RH |
| 442 | ivtpadelay30<br><br>Show the field ONLY if:<br>([site]='SMCA' or [site]='SMCH' or [site]='SMCW' or [site]='UMCB') and [gs_symptomlocation]<>'4' and [gs_ivthroinit]='1' and [calc_door_to_ivtpa]>30 | If IV Thrombolytic was initiated greater than 30 minutes after hospital arrival, were Eligibility or Medical reason(s) documented as the cause for delay? | radio<br>1 \| Yes<br>2 \| No<br><br>Custom alignment: RH |
| 443 | ivtpadelay_er<br><br>Show the field ONLY if:<br>([site]='SMCA' or [site]='SMCH' or [site]='SMCW' or [site]='UMCB') and ([ivtpadelay]='1' or [ivtpadelay45]='1' or [ivtpadelay30]='1') | Eligibility Reason(s)<br>*Select all that apply.* | checkbox<br>1 \| ivtpadelay_er___1 \| Social/Religious<br>2 \| ivtpadelay_er___2 \| Initial refusal<br>3 \| ivtpadelay_er___3 \| Care-team unable to determine eligibility |
| 444 | ivtpadelay_erspec<br><br>Show the field ONLY if:<br>([site]='SMCA' or [site]='SMCH' or [site]='SMCW' or [site]='UMCB') and ([ivtpadelay]='1' or [ivtpadelay45]='1' or [ivtpadelay30]='1') | Specify eligibility reason | text<br>Field Annotation: @CHARLIMIT=60 |

| | | | |
|---|---|---|---|
| 445 | ivtpadelay_mr<br><br>Show the field ONLY if:<br>([site]='SMCA' or [site]='SMCH' or [site]='SMCW' or [site]='UMCB') and ([ivtpadelay]='1' or [ivtpadelay45]='1' or [ivtpadelay30]='1') | Medical Reason(s)<br>*Select all that apply.* | checkbox<br>1 ivtpadelay_mr___1 Hypertension requiring aggressive control with IV medications<br>2 ivtpadelay_mr___2 Further diagnostic evaluation to confirm stroke for patients with hypoglycemia (blood glucose < 50), seizures, or major metabolic disorders<br>3 ivtpadelay_mr___3 Management of concomitant emergent/acute conditions such as cardiopulmonary arrest, respiratory failure (requiring intubation)<br>4 ivtpadelay_mr___4 Investigational or experimental protocol for thrombolysis<br>5 ivtpadelay_mr___5 Need for additional PPE for suspected/confirmed infectious disease |
| 446 | ivtpadelay_mrspec<br><br>Show the field ONLY if:<br>([site]='SMCA' or [site]='SMCH' or [site]='SMCW' or [site]='UMCB') and ([ivtpadelay]='1' or [ivtpadelay45]='1' or [ivtpadelay30]='1') | Specify medical reason | text<br>Field Annotation: @CHARLIMIT=60 |
| 447 | ivtpadelay_hro<br><br>Show the field ONLY if:<br>([site]='SMCA' or [site]='SMCH' or [site]='SMCW' or [site]='UMCB') and ([ivtpadelay]='2' or [ivtpadelay45]='2' or [ivtpadelay30]='2') | Hospital Related or Other Reason(s)<br>*Select all that apply.* | checkbox<br>5 ivtpadelay_hro___5 Need for additional imaging<br>1 ivtpadelay_hro___1 Delay in stroke diagnosis<br>2 ivtpadelay_hro___2 In-hospital time delay<br>3 ivtpadelay_hro___3 Equipment-related delay<br>4 ivtpadelay_hro___4 Other |
| 448 | ivtpadelay_other<br><br>Show the field ONLY if:<br>[ivtpadelay_hro(4)]='1' | Specify Other Reason | text<br>Field Annotation: @CHARLIMIT=60 |
| 449 | gs_ivtpaoutside<br><br>Show the field ONLY if:<br>[gs_patientarrival]='1' or [gs_patientarrival]='10' or [gs_patientarrival]='3' | Section Header: *Other thrombolytic/reperfusion therapyDid this patient receive an Acute Stroke Treatment?[treatment_recvd]*<br><br>IV Thrombolytic at an outside hospital EMS / Mobile Stroke Unit? | radio<br>1 Yes<br>2 No<br><br>Custom alignment: RH |

| | | | |
|---|---|---|---|
| 450 | gs_thrombadmin<br>Show the field ONLY if:<br>[gs_ivtpaoutside]='1' | If yes, select Thrombolytic administered at outside hospital or Mobile Stroke Unit | radio<br>1 Alteplase<br>2 Tenecteplase<br>Custom alignment: RH |
| 451 | emsx_neurochecks<br>Show the field ONLY if:<br>[gs_ivtpaoutside] = '1' | Were neuro checks completed by EMS during transport? | yesno<br>1 Yes<br>0 No<br>Custom alignment: RH |
| 452 | gs_thromexp<br>Show the field ONLY if:<br>([site]='SMCA' or [site]='SMCH' or [site]='SMCW' or [site]='UMCB') and [gs_stroketype] <> '4' and [gs_stroketype] <> '5' and ([treatment_recvd]='1' or [gs_notadmit]<>'1') and [hide_nonessential_fields(1)]='0' | Investigational or experimental protocol for thrombolysis? | radio<br>1 Yes<br>2 No<br>Custom alignment: RH |
| 453 | gs_thromexptype<br>Show the field ONLY if:<br>([site]='SMCA' or [site]='SMCH' or [site]='SMCW' or [site]='UMCB') and [gs_thromexp] = '1' and [hide_nonessential_fields(1)]='0' | If yes, Specify | text<br>Field Annotation: @CHARLIMIT=50 |
| 454 | gs_nothromcomment<br>Show the field ONLY if:<br>([site]='SMCA' or [site]='SMCH' or [site]='SMCW' or [site]='UMCB') and [gs_stroketype] <> '4' and [gs_stroketype] <> '5' and ([treatment_recvd]='1' or [gs_notadmit]<>'1') and [hide_nonessential_fields(1)]='0' | Additional Comments Related to Thrombolytics | text<br>Field Annotation: @CHARLIMIT=60 |
| 455 | gs_iatpa<br>Show the field ONLY if:<br>([site]='SMCA' or [site]='SMCH' or [site]='SMCW' or [site]='UMCB') and [gs_stroketype] <> '4' and [gs_stroketype] <> '5' and ([treatment_recvd]='1' or [gs_notadmit]<>'1') and [hide_nonessential_fields(1)]='0' | Section Header: *Endovascular Therapy*<br>Catheter-based stroke treatment at this hospital? | radio<br>1 Yes<br>2 No<br>Custom alignment: RH |

| | | | |
|---|---|---|---|
| 456 | gs_iatpadatetime_precision<br><br>Show the field ONLY if:<br>([site]='SMCA' or [site]='SMCH' or [site]='SMCW' or [site]='UMCB') and [gs_stroketype] <> '4' and [gs_stroketype] <> '5' and ([treatment_recvd]='1' or [gs_notadmit]<>'1') and [hide_nonessential_fields(1)]='0' | IA Thrombolytic or MER Date/Time Precision | dropdown<br>5 \| Date & Time (MM/DD/YYYY HH24:MM)<br>3 \| Date Only (MM/DD/YYYY)<br>0 \| Unknown |
| 457 | gs_iatpadatetime<br><br>Show the field ONLY if:<br>([site]='SMCA' or [site]='SMCH' or [site]='SMCW' or [site]='UMCB') and [gs_stroketype] <> '4' and [gs_stroketype] <> '5' and [gs_iatpadatetime_precision] <> '0' and ([treatment_recvd]='1' or [gs_notadmit]<>'1') and [hide_nonessential_fields(1)]='0' | IA Thrombolytic or MER Initiation Date/Time<br>IA-tPA Infusion or Device Deployment/Insertion Time (whichever is first) | text (datetime_mdy, Min: 2010-01-01 00:00, Max: 2099-12-31 23:59), Identifier |
| 458 | gs_iatpaoutside<br><br>Show the field ONLY if:<br>([site]='SMCA' or [site]='SMCH' or [site]='SMCW' or [site]='UMCB') and [gs_stroketype] <> '4' and [gs_stroketype] <> '5' and ([treatment_recvd]='1' or [gs_notadmit]<>'1') and [hide_nonessential_fields(1)]='0' | Catheter-based stroke treatment at outside hospital? | radio<br>1 \| Yes<br>2 \| No<br><br>Custom alignment: RH |
| 459 | gs_comp_throm_ther<br><br>Show the field ONLY if:<br>([site]='SMCA' or [site]='SMCH' or [site]='SMCW' or [site]='UMCB') and ([gs_ivthroinit] <> '2' or [gs_ivtpaoutside] <> '2' or [gs_iatpa] <> '2' or [gs_iatpaoutside] <> '2') and [gs_stroketype] <> '4' and [gs_stroketype] <> '5' and ([treatment_recvd]='1' or [gs_notadmit]<>'1') | Section Header: *Complications of Thrombolytic Therapy*<br><br>Complications of Reperfusion Therapy (Thrombolytic or MER)<br>*Select all that apply.* | checkbox<br>1 \| gs_comp_throm_ther___1 \| Symptomatic intracranial hemorrhage < 36 hours<br>2 \| gs_comp_throm_ther___2 \| Life threatening, serious systemic hemorrhage < 36 hours<br>5 \| gs_comp_throm_ther___5 \| Other serious complication<br>3 \| gs_comp_throm_ther___3 \| No serious complications<br>4 \| gs_comp_throm_ther___4 \| UTD<br><br>Field Annotation: @NONEOFTHEABOVE='3,4' |

| | | | |
|---|---|---|---|
| 460 | comp_throm_ther_oth<br><br>Show the field ONLY if:<br>[gs_comp_throm_ther(5)]='1' | Other Serious Complications of Thrombolytic Therapy | checkbox<br>1 comp_throm_ther_oth___1 Angioedema<br>0 comp_throm_ther_oth___0 Other {comp_throm_ther_spec:icons} |
| 461 | comp_throm_ther_spec<br><br>Show the field ONLY if:<br>[comp_throm_ther_oth(0)]='1' | Specify Other Serious Complications of Thrombolytic Therapy | text |
| 462 | gs_comptransf<br><br>Show the field ONLY if:<br>([site]='SMCA' or [site]='SMCH' or [site]='SMCW' or [site]='UMCB') an d ([gs_ivthroinit] <> '2' or [gs_ivtpa outside] <> '2' or [gs_iatpa] <> '2' o r [gs_iatpaoutside] <> '2') and [gs_ stroketype] <> '4' and [gs_strokety pe] <> '5' and ([treatment_recvd] ='1' or [gs_notadmit]<>'1') and [hi de_nonessential_fields(1)]='0' | If bleeding complications occur in patient transferred after IV thrombolytic:<br>(If the patient is not transferred after IV thrombolytic (patient remains at your hospital), then this element is not applicable. Select N/A.) | dropdown<br>2 Symptomatic hemorrhage detected prior to patient transfer<br>3 Symptomatic hemorrhage detected only after patient transfer<br>4 Unable to determine<br>5 N/A<br><br>Custom alignment: LV |
| 463 | ich_dysphagia<br><br>Show the field ONLY if:<br>[site]='UMCB' | Section Header: *Dysphagia Screening*<br><br>✪ Was an initial dysphagia screen performed? | radio<br>1 Yes, within 24 hours of arrival<br>2 Yes, greater than 24 hours after arrival<br>3 No<br>4 NC |
| 464 | gs_npo<br><br>Show the field ONLY if:<br>([treatment_recvd]='1' or [gs_nota dmit]<>'1') | Patient NPO throughout the entire hospital stay? | radio<br>1 Yes<br>2 No/ND<br><br>Custom alignment: RH |
| 465 | gs_dysphagia<br><br>Show the field ONLY if:<br>([treatment_recvd]='1' or [gs_nota dmit]<>'1') | Was patient screened for dysphagia prior to any oral intake including water or medications? | radio<br>1 Yes<br>2 No/ND<br>3 NC<br><br>Custom alignment: RH |

| | | | |
|---|---|---|---|
| 466 | gs_dysphagiaresult<br><br>Show the field ONLY if:<br>[gs_dysphagia] = '1' and [hide_nonessential_fields(1)]='0' | If yes, Dysphagia screening results: | radio<br>1 Pass<br>2 Fail<br>3 ND<br><br>Custom alignment: RH |
| 467 | swallowscreen_failed<br><br>Show the field ONLY if:<br>[gs_dysphagiaresult] = '2' and [hide_nonessential_fields(1)]='0' | Were PO meds or food, including water, documented as given after failing initial swallow screen, and prior to a subsequent swallow screen? | yesno<br>1 Yes<br>0 No<br><br>Custom alignment: RH |
| 468 | ich_intake<br><br>Show the field ONLY if:<br>[site]='UMCB' | ✪ Was first oral intake consistent with any Recommended Dietary Modifications? | radio<br>1 Yes<br>2 No<br><br>Custom alignment: RH |
| 469 | swallowscreen_subsequent<br><br>Show the field ONLY if:<br>([treatment_recvd]='1' or [gs_notadmit]<>'1') and [hide_nonessential_fields(1)]='0' | Were there any subsequent fallouts after the initial swallow screen, during the entire hospital stay? | yesno<br>1 Yes<br>0 No<br><br>Custom alignment: RH |
| 470 | gs_pneumonia<br><br>Show the field ONLY if:<br>([site]='SMCA' or [site]='SMCH' or [site]='SMCW' or [site]='UMCB') and ([treatment_recvd]='1' or [gs_notadmit]<>'1') and [hide_nonessential_fields(1)]='0' | Treatment for Hospital-Acquired Pneumonia | radio<br>1 Yes<br>2 No<br>3 NC<br><br>Custom alignment: RH |
| 471 | jc_vteprophylaxis1<br><br>Show the field ONLY if:<br>[gs_comfortonly] <> '1' and [jc_vteprophylaxis7] <> '1' and [gs_notadmit]<>'1' and [hide_nonessential_fields(1)]='0' | Section Header: *VTE Interventions*<br><br>1- Low dose unfractionated heparin (LDUH) | radio<br>1 LDUH |

| | | | |
|---|---|---|---|
| 472 | jc_vteprophylaxis2<br><br>Show the field ONLY if: [gs_comfortonly] <> '1' and [jc_vteprophylaxis7] <> '1' and [gs_notadmit]<>'1' and [hide_nonessential_fields(1)]='0' | 2- Low molecular weight heparin (LMWH) | radio<br>1 \| LMWH |
| 473 | jc_vteprophylaxis3<br><br>Show the field ONLY if: [gs_comfortonly] <> '1' and [jc_vteprophylaxis7] <> '1' and [gs_notadmit]<>'1' and [hide_nonessential_fields(1)]='0' | 3- Intermittent pneumatic compression devices (IPC) | radio<br>1 \| IPC |
| 474 | jc_vteprophylaxis4<br><br>Show the field ONLY if: ([site]='SMCA' or [site]='SMCH' or [site]='SMCW' or [site]='UMCB') and [gs_comfortonly] <> '1' and [jc_vteprophylaxis7] <> '1' and [gs_notadmit]<>'1' and [hide_nonessential_fields(1)]='0' | 4- Graduated compression stockings (GCS) | radio<br>1 \| GCS |
| 475 | jc_vteprophylaxis5<br><br>Show the field ONLY if: [gs_comfortonly] <> '1' and [jc_vteprophylaxis7] <> '1' and [gs_notadmit]<>'1' and [hide_nonessential_fields(1)]='0' | 5- Factor Xa Inhibitor | radio<br>1 \| Factor Xa Inhibitor |
| 476 | jc_vteprophylaxis6<br><br>Show the field ONLY if: [gs_comfortonly] <> '1' and [jc_vteprophylaxis7] <> '1' and [gs_notadmit]<>'1' and [hide_nonessential_fields(1)]='0' | 6- Warfarin | radio<br>1 \| Warfarin |
| 477 | jc_vteprophylaxis8<br><br>Show the field ONLY if: [gs_comfortonly] <> '1' and [jc_vteprophylaxis7] <> '1' and [gs_notadmit]<>'1' and [hide_nonessential_fields(1)]='0' | 7- Venous foot pumps (VFP) | radio<br>1 \| VFP |

| | | | |
|---|---|---|---|
| 478 | jc_vteprophylaxis9<br><br>Show the field ONLY if:<br>[gs_comfortonly] <> '1' and [jc_vteprophylaxis7] <> '1' and [gs_notadmit]<>'1' and [hide_nonessential_fields(1)]='0' | 8- Oral Factor Xa Inhibitor | radio<br>1 \| Oral Factor Xa Inhibitor |
| 479 | jc_vteprophylaxis10<br><br>Show the field ONLY if:<br>([site]='SMCA' or [site]='SMCH' or [site]='SMCW' or [site]='UMCB') and [gs_comfortonly] <> '1' and [jc_vteprophylaxis7] <> '1' and [gs_notadmit]<>'1' and [hide_nonessential_fields(1)]='0' | 9- Aspirin | radio<br>1 \| Aspirin |
| 480 | jc_vteprophylaxis7<br><br>Show the field ONLY if:<br>[gs_comfortonly] <> '1' and [gs_notadmit]<>'1' and [hide_nonessential_fields(1)]='0' | A- None of the above or not documented or unable to determine from medical record documentation | radio<br>1 \| None, ND, or UTD |
| 481 | jc_vtedate<br><br>Show the field ONLY if:<br>[gs_comfortonly] <> '1' and [gs_notadmit] <> '1' and [jc_vteprophylaxis7] <> '1' and [hide_nonessential_fields(1)]='0' | What date was the VTE prophylaxis administered after hospital admission? | text (date_mdy, Min: 2010-01-01, Max: 2099-12-31), Identifier |
| 482 | ich_ipcdt_precision<br><br>Show the field ONLY if:<br>[site]='UMCB' | ✪ IPC Date/Time Precision | dropdown<br>5 \| Date & Time (MM/DD/YYYY HH24:MM)<br>3 \| Date Only (MM/DD/YYYY)<br>0 \| Unknown |
| 483 | ich_ipcdt<br><br>Show the field ONLY if:<br>[site]='UMCB' and [ich_ipcdt_precision]<>'0' | ✪ What was the date and time the IPC device was placed? | text (datetime_mdy, Min: 2010-01-01 00:00, Max: 2099-12-31 23:59)<br>Custom alignment: RH |
| 484 | ich_ipcdoc<br><br>Show the field ONLY if:<br>[site]='UMCB' | ✪ Is there physician/ APN/ PA documentation why IPC was not used for VTE prophylaxis? | radio<br>1 \| Yes<br>2 \| No<br><br>Custom alignment: RH |

| | | | |
|---|---|---|---|
| 485 | jc_novteadmis<br><br>Show the field ONLY if:<br>[gs_notadmit] <> '1' and [gs_comfortonly] <> '1' and [hide_nonessential_fields(1)]='0' | Is there physician/APN/PA or pharmacist documentation why VTE prophylaxis was not administered at hospital admission? | radio<br>1 Yes<br>2 No<br><br>Custom alignment: RH |
| 486 | jc_reasonoralxa<br><br>Show the field ONLY if:<br>[jc_vteprophylaxis9] = '1' and [hide_nonessential_fields(1)]='0' | Is there physician/APN/PA documentation why Oral Factor Xa Inhibitor was administered for VTE prophylaxis? | radio<br>1 Yes<br>2 No<br><br>Custom alignment: RH |
| 487 | gs_otheranticoag<br><br>Show the field ONLY if:<br>([treatment_recvd]='1' or [gs_notadmit]<>'1') and ([jc_novteadmis]='1' or [jc_reasonoralxa]='1' or [site] = 'SMCH' or [site] = 'SMCW') and [hide_nonessential_fields(1)]='0' | Section Header: *Other In-hospital Treatments and Screening*<br><br>Other Therapeutic Anticoagulation | checkbox<br>1 gs_otheranticoag___1 Unfractionated heparin IV<br>2 gs_otheranticoag___2 Dabigatran (Pradaxa)<br>3 gs_otheranticoag___3 Argatroban<br>4 gs_otheranticoag___4 Desirudin (Iprivask)<br>7 gs_otheranticoag___7 Rivaroxaban (Xarelto)<br>8 gs_otheranticoag___8 apixaban (Eliquis)<br>5 gs_otheranticoag___5 Lepirudin (Refludan)<br>9 gs_otheranticoag___9 Edoxaban (Savaysa)<br>6 gs_otheranticoag___6 Other Anticoagulant |
| 488 | gs_dvtdoc<br><br>Show the field ONLY if:<br>([site]='SMCA' or [site]='SMCH' or [site]='SMCW' or [site]='UMCB') and [gs_comfortonly] <> '1' and ([treatment_recvd]='1' or [gs_notadmit]<>'1') and [hide_nonessential_fields(1)]='0' | Was DVT or PE documented? | radio<br>1 Yes<br>2 No/ND<br><br>Custom alignment: RH |
| 489 | gs_antiplateletadm<br><br>Show the field ONLY if:<br>[gs_comfortonly] <> '1' and ([treatment_recvd]='1' or [gs_notadmit]<>'1') and [hide_nonessential_fields(1)]='0' | Was antithrombotic therapy administered by the end of hospital day 2? | radio<br>1 Yes<br>6 No/ND<br>5 NC<br><br>Custom alignment: RH |

| | | | |
|---|---|---|---|
| 490 | gs_antithrom2type<br><br>Show the field ONLY if:<br>([site]='SMCA' or [site]='SMCH' or [site]='SMCW' or [site]='UMCB') and [gs_antiplateletadm] = '1' and [hide_nonessential_fields(1)]='0' | If yes, (antithrombotic therapy) | checkbox<br>1 gs_antithrom2type___1 Antiplatelet<br>2 gs_antithrom2type___2 Anticoagulant |
| 491 | jc_noantithroday2<br><br>Show the field ONLY if:<br>([treatment_recvd]='1' or [gs_notadmit]<>'1') and [hide_nonessential_fields(1)]='0' | Is there documentation by a physician/advanced practice nurse/physician assistant (physician/APN/PA) or pharmacist in the medical record of a reason for not administering antithrombotic therapy by end of hospital day 2? | radio<br>1 Yes<br>2 No<br><br>Custom alignment: RH |
| 492 | activeinfec | Active bacterial or viral infection at admission or during hospitalization: | checkbox<br>4 activeinfec___4 None<br>2 activeinfec___2 Emerging Infectious Disease<br>3 activeinfec___3 Bacterial infection<br>6 activeinfec___6 Influenza<br>5 activeinfec___5 Seasonal Cold<br>7 activeinfec___7 Other Viral Infection<br><br>Field Annotation: @NONEOFTHEABOVE='4' |
| 493 | activeinfecopt<br><br>Show the field ONLY if:<br>[activeinfec(2)]='1' | Specify Emerging Infectious Disease | checkbox<br>1 activeinfecopt___1 SARS-COV-1<br>2 activeinfecopt___2 SARS-COV-2 (COVID-19)<br>3 activeinfecopt___3 MERS<br>5 activeinfecopt___5 Other Emerging Infectious Disease |
| 494 | activeother | Other Infectious Respiratory Pathogen (Historic) | radio<br>1 Other infectious respiratory pathogen |
| 495 | activecoldflu | Seasonal cold or flu (Historic) | radio<br>1 Seasonal cold or flu |
| 496 | desc_measurements_shazam | Section Header: *Measurements (first measurement upon presentation to your hospital)* | descriptive |
| 497 | c_plateletutd<br><br>Show the field ONLY if:<br>([site] = 'SMCA' or [site] = 'UMCB') and ([treatment_recvd]='1' or [gs_notadmit]<>'1') and [hide_nonessential_fields(1)]='0' | Initial Platelet Count at Hospital Arrival - UTD | radio<br>1 UTD |

| | | | |
|---|---|---|---|
| 498 | c_platelet<br><br>Show the field ONLY if:<br>([site] = 'SMCA' or [site] = 'UMCB') and [c_plateletutd] <> '1' and ([treatment_recvd]='1' or [gs_notadmit]<>'1') and [hide_nonessential_fields(1)]='0' | What is the first platelet count obtained prior to or after hospital arrival? | text (integer, Min: 1, Max: 999999) |
| 499 | gs_inr_nd<br><br>Show the field ONLY if:<br>([site]='SMCA' or [site]='SMCH' or [site]='SMCW' or [site]='UMCB') and ([treatment_recvd]='1' or [gs_notadmit]<>'1') and [hide_nonessential_fields(1)]='0' | INR ND/NC | radio<br>1 ND<br>2 NC<br><br>Custom alignment: RH |
| 500 | gs_inr<br><br>Show the field ONLY if:<br>([site]='SMCA' or [site]='SMCH' or [site]='SMCW' or [site]='UMCB') and [gs_inr_nd] = '' and ([treatment_recvd]='1' or [gs_notadmit]<>'1') and [hide_nonessential_fields(1)]='0' | INR<br>*Round to one decimal place.* | text (number, Min: 0, Max: 25) |
| 501 | c_inrperf<br><br>Show the field ONLY if:<br>([site]='SMCA' or [site]='SMCH' or [site]='SMCW' or [site]='UMCB') and ([treatment_recvd]='1' or [gs_notadmit]<>'1') and [hide_nonessential_fields(1)]='0' | INR Value > 1.4 | radio<br>1 Yes<br>2 No<br><br>Custom alignment: RH |
| 502 | gs_scr_nd<br><br>Show the field ONLY if:<br>([site]='SMCA' or [site]='SMCH' or [site]='SMCW' or [site]='UMCB') and ([treatment_recvd]='1' or [gs_notadmit]<>'1') and [hide_nonessential_fields(1)]='0' | Serum Creatinine ND | radio<br>1 ND |

| | | | |
|---|---|---|---|
| 503 | gs_scr<br><br>Show the field ONLY if:<br>([site]='SMCA' or [site]='SMCH' or [site]='SMCW' or [site]='UMCB') and [gs_scr_nd] <> '1' and ([treatment_recvd]='1' or [gs_notadmit]<>'1') and [hide_nonessential_fields(1)]='0' | Serum Creatinine | text (number_1dp, Max: 999.9) |
| 504 | gs_bloodglucose_nd<br><br>Show the field ONLY if:<br>([site]='SMCA' or [site]='SMCH' or [site]='SMCW' or [site]='UMCB') and ([treatment_recvd]='1' or [gs_notadmit]<>'1') | Blood Glucose ND | dropdown<br>1 ND<br>2 Too Low<br>3 Too High |
| 505 | gs_fastingblood<br><br>Show the field ONLY if:<br>([site]='SMCA' or [site]='SMCH' or [site]='SMCW' or [site]='UMCB') and [gs_bloodglucose_nd] = '' and ([treatment_recvd]='1' or [gs_notadmit]<>'1') | What is the first blood glucose value obtained prior to or after hospital arrival? (required if patient received IV tPA) | text (integer, Min: 10, Max: 1000) |
| 506 | gs_hba1cnd<br><br>Show the field ONLY if:<br>([site]='SMCA' or [site]='SMCH' or [site]='SMCW' or [site]='UMCB') and ([treatment_recvd]='1' or [gs_notadmit]<>'1') and [hide_nonessential_fields(1)]='0' | A1C: ND | radio<br>1 ND |
| 507 | gs_hba1c<br><br>Show the field ONLY if:<br>([site]='SMCA' or [site]='SMCH' or [site]='SMCW' or [site]='UMCB') and [gs_hba1cnd] <> '1' and ([treatment_recvd]='1' or [gs_notadmit]<>'1') and [hide_nonessential_fields(1)]='0' | A1C<br>*NOTICE: GWTG only accepts answers with up to one decimal place.* | text (number, Min: 1, Max: 9999.9) |

| | | | |
|---|---|---|---|
| 508 | gs_lipidsnd<br><br>Show the field ONLY if: ([site]='SMCA' or [site]='SMCH' or [site]='SMCW' or [site]='UMCB') and [gs_lipidsnc] <> '1' and ([treatment_recvd]='1' or [gs_notadmit]<>'1') and [hide_nonessential_fields(1)]='0' | Lipids: ND | radio<br>1 ND |
| 509 | gs_lipidsnc<br><br>Show the field ONLY if: ([site]='SMCA' or [site]='SMCH' or [site]='SMCW' or [site]='UMCB') and [gs_lipidsnd] <> '1' and ([treatment_recvd]='1' or [gs_notadmit]<>'1') and [hide_nonessential_fields(1)]='0' | Lipids: NC | radio<br>1 NC |
| 510 | gs_cholesterol<br><br>Show the field ONLY if: ([site]='SMCA' or [site]='SMCH' or [site]='SMCW' or [site]='UMCB') and [gs_lipidsnd] <> '1' and [gs_lipidsnc] <> '1' and ([treatment_recvd]='1' or [gs_notadmit]<>'1') and [hide_nonessential_fields(1)]='0' | Total Chol | text (integer) |
| 511 | gs_triglycerides<br><br>Show the field ONLY if: ([site]='SMCA' or [site]='SMCH' or [site]='SMCW' or [site]='UMCB') and [gs_lipidsnd] <> '1' and [gs_lipidsnc] <> '1' and ([treatment_recvd]='1' or [gs_notadmit]<>'1') and [hide_nonessential_fields(1)]='0' | Triglycerides | text (integer) |
| 512 | gs_hdl<br><br>Show the field ONLY if: ([site]='SMCA' or [site]='SMCH' or [site]='SMCW' or [site]='UMCB') and [gs_lipidsnd] <> '1' and [gs_lipidsnc] <> '1' and ([treatment_recvd]='1' or [gs_notadmit]<>'1') and [hide_nonessential_fields(1)]='0' | HDL | text (integer, Max: 999) |

| | | | |
|---|---|---|---|
| 513 | gs_ldl<br><br>Show the field ONLY if:<br>([site]='SMCA' or [site]='SMCH' or [site]='SMCW' or [site]='UMCB') and [gs_lipidsnd] <> '1' and [gs_lipidsnc] <> '1' and ([treatment_recvd]='1' or [gs_notadmit]<>'1') and [hide_nonessential_fields(1)]='0' | LDL | text (integer) |
| 514 | gs_vital_nd<br><br>Show the field ONLY if:<br>([site]='SMCA' or [site]='SMCH' or [site]='SMCW' or [site]='UMCB') and ([treatment_recvd]='1' or [gs_notadmit]<>'1') and [hide_nonessential_fields(1)]='0' | Section Header: *Vital Signs*<br><br>Vital Signs: UTD | radio<br>1 UTD |
| 515 | gs_hr<br><br>Show the field ONLY if:<br>([site]='SMCA' or [site]='SMCH' or [site]='SMCW' or [site]='UMCB') and [gs_vital_nd] <> '1' and ([treatment_recvd]='1' or [gs_notadmit]<>'1') and [hide_nonessential_fields(1)]='0' | Heart Rate (beats per minute): | text |
| 516 | gs_systolic<br><br>Show the field ONLY if:<br>([site]='SMCA' or [site]='SMCH' or [site]='SMCW' or [site]='UMCB') and [gs_vital_nd] <> '1' and ([treatment_recvd]='1' or [gs_notadmit]<>'1') | Section Header: *What is the first blood pressure obtained prior to or after hospital arrival?*<br><br>Blood Pressure Systolic<br>*mmHg (Systolic/Diastolic)* | text |
| 517 | gs_diastolic<br><br>Show the field ONLY if:<br>([site]='SMCA' or [site]='SMCH' or [site]='SMCW' or [site]='UMCB') and [gs_vital_nd] <> '1' and ([treatment_recvd]='1' or [gs_notadmit]<>'1') | Blood Pressure Diastolic<br>*mmHg (Systolic/Diastolic)* | text |

| | | | |
|---|---|---|---|
| 518 | gs_heightnd<br>Show the field ONLY if: ([site]='SMCA' or [site]='SMCH' or [site]='SMCW' or [site]='UMCB') and ([treatment_recvd]='1' or [gs_notadmit]<>'1') and [hide_nonessential_fields(1)]='0' | Height: ND | radio<br>1 ND |
| 519 | gs_height<br>Show the field ONLY if: ([site]='SMCA' or [site]='SMCH' or [site]='SMCW' or [site]='UMCB') and [gs_heightnd] <> '1' and ([treatment_recvd]='1' or [gs_notadmit]<>'1') and [hide_nonessential_fields(1)]='0' | Height | text (integer, Max: 999) |
| 520 | gs_heightunit<br>Show the field ONLY if: ([site]='SMCA' or [site]='SMCH' or [site]='SMCW' or [site]='UMCB') and [gs_heightnd] <> '1' and ([treatment_recvd]='1' or [gs_notadmit]<>'1') and [hide_nonessential_fields(1)]='0' | Height unit | radio<br>1 in<br>2 cm<br>Custom alignment: RH |
| 521 | gs_weightnd<br>Show the field ONLY if: ([site]='SMCA' or [site]='SMCH' or [site]='SMCW' or [site]='UMCB') and ([treatment_recvd]='1' or [gs_notadmit]<>'1') and ([hide_nonessential_fields(1)]='0' or (([gs_med_hist(6)]='1' or [gs_diagdm]='1') and [gs_notadmit]='2' and [jc_disposition]<>'6') or [gs_ivthroinit]='1') | Weight: ND | radio<br>1 ND |

| | | | |
|---|---|---|---|
| 522 | gs_weight<br><br>Show the field ONLY if:<br>([site]='SMCA' or [site]='SMCH' or [site]='SMCW' or [site]='UMCB') and [gs_weightnd] <> '1' and ([treatment_recvd]='1' or [gs_notadmit]<>'1') and ([hide_nonessential_fields(1)]='0' or (([gs_med_hist(6)]='1' or [gs_diagdm]='1') and [gs_notadmit]='2' and [jc_disposition]<>'6') or [gs_ivthroinit]='1') | Weight | text (integer, Max: 9999) |
| 523 | gs_weightunit<br><br>Show the field ONLY if:<br>([site]='SMCA' or [site]='SMCH' or [site]='SMCW' or [site]='UMCB') and [gs_weightnd] <> '1' and ([treatment_recvd]='1' or [gs_notadmit]<>'1') and ([hide_nonessential_fields(1)]='0' or (([gs_med_hist(6)]='1' or [gs_diagdm]='1') and [gs_notadmit]='2' and [jc_disposition]<>'6') or [gs_ivthroinit]='1') | Weight unit | radio<br>1 lb<br>2 kg<br><br>Custom alignment: RH |
| 524 | gs_waist_nd<br><br>Show the field ONLY if:<br>([site]='SMCA' or [site]='SMCH' or [site]='SMCW' or [site]='UMCB') and ([treatment_recvd]='1' or [gs_notadmit]<>'1') and [hide_nonessential_fields(1)]='0' | Waist Circumference: ND | radio<br>1 ND |
| 525 | gs_waist<br><br>Show the field ONLY if:<br>([site]='SMCA' or [site]='SMCH' or [site]='SMCW' or [site]='UMCB') and [gs_waist_nd] <> '1' and ([treatment_recvd]='1' or [gs_notadmit]<>'1') and [hide_nonessential_fields(1)]='0' | Waist Circumference | text (number) |

| | | | |
|---|---|---|---|
| 526 | gs_waistunits<br><br>Show the field ONLY if:<br>([site]='SMCA' or [site]='SMCH' or [site]='SMCW' or [site]='UMCB') and [gs_waist_nd] <> '1' and ([treatment_recvd]='1' or [gs_notadmit]<>'1') and [hide_nonessential_fields(1)]='0' | Waist Circumference units | radio<br>1 in<br>2 cm<br><br>Custom alignment: RH |
| 527 | gs_bmind<br><br>Show the field ONLY if:<br>([site]='SMCA' or [site]='SMCH' or [site]='SMCW' or [site]='UMCB') and ([treatment_recvd]='1' or [gs_notadmit]<>'1') and [hide_nonessential_fields(1)]='0' | BMI: ND | radio<br>1 ND |
| 528 | gs_bmi<br><br>Show the field ONLY if:<br>([site]='SMCA' or [site]='SMCH' or [site]='SMCW' or [site]='UMCB') and [gs_bmind] <> '1' and ([treatment_recvd]='1' or [gs_notadmit]<>'1') and [hide_nonessential_fields(1)]='0' | BMI | calc<br>Calculation: round(if([gs_weightunit] = '1', [gs_weight] * 0.453592, [gs_weight]) / ((if([gs_heightunit] = '1', [gs_height] * 0.0254, [gs_height] / 100))^(2)), 2) |
| 529 | hospitalization_complete | Section Header: *Form Status*<br><br>Complete? | dropdown<br>0 Incomplete<br>1 Unverified<br>2 Complete |

Instrument: **Advanced Stroke Care** (advanced_stroke_care)

Collapse

| | | | |
|---|---|---|---|
| 530 | c_iaroutetpa<br><br>Show the field ONLY if:<br>([site]='SMCA' or [site]='UMCB') and [gs_age] >= 18 and [gs_notadmit] = '2' and [jc_admitdate] <> '' and [gs_discdatetime] <> '' and starts_with([jc_princicd10diagnosis], 'i63') and ([jc_princicd10procedure]='3E03017' or [jc_princicd10procedure]='3E03317' or [jc_princicd10procedure]='3E04017' or [jc_princicd10procedure]='3E04317' or [jc_princicd10procedure]='3E05017' or [jc_princicd10procedure]='3E05317' or [jc_princicd10procedure]='3E06017' or [jc_princicd10procedure]='3E06317' or [jc_othericd10proc1]='3E03017' or [jc_othericd10proc1]='3E03317' or [jc_othericd10proc1]='3E04017' or [jc_othericd10proc1]='3E04317' or [jc_othericd10proc1]='3E05017' or [jc_othericd10proc1]='3E05317' or [jc_othericd10proc1]='3E06017' or [jc_othericd10proc1]='3E06317' or [jc_othericd10proc2]='3E03017' or [jc_othericd10proc2]='3E03317' or [jc_othericd10proc2]='3E04017' or [jc_othericd10proc2]='3E04317' or [jc_othericd10proc2]='3E05017' or [jc_othericd10proc2]='3E05317' or [jc_othericd10proc2]='3E06017' or [jc_othericd10proc2]='3E06317') and [jc_carotid] = '2' and [hide_nonessential_fields(1)]='0' | Section Header: *Catheter-based / Endovascular Stroke Treatment*<br><br>Is there documentation that the route of thrombolytic (t-PA) administration was intra-arterial (IA)? | radio<br>1 Yes<br>2 No<br><br>Custom alignment: RH |
| 531 | c_iathrominit<br><br>Show the field ONLY if:<br>[c_iaroutetpa] = '1' or [c_iaroutetpa] = '2' and [hide_nonessential_fields(1)]='0' | Is there documentation that IA thrombolytic therapy was initiated at this hospital? | radio<br>1 Yes<br>2 No<br><br>Custom alignment: RH |

| | | | |
|---|---|---|---|
| 532 | c_iathrominitdt_precision<br><br>Show the field ONLY if:<br>[c_iathrominit] = '1' and [hide_nonessential_fields(1)]='0' | IA Thrombolytic Initiation Date/Time Precision | dropdown<br>5 Date & Time (MM/DD/YYYY HH24:MM)<br>3 Date Only (MM/DD/YYYY)<br>0 Unknown<br><br>Custom alignment: RH |
| 533 | c_iathrominitdt<br><br>Show the field ONLY if:<br>[c_iathrominit] = '1' and [c_iathrominitdt_precision] <> '0' and [hide_nonessential_fields(1)]='0' | What is the date and time that IA thrombolytic therapy was initiated for this patient at this hospital? | text (datetime_mdy, Min: 2010-01-01 00:00, Max: 2099-12-31 23:59), Identifier<br>Custom alignment: RH |
| 534 | c_delayed<br><br>Show the field ONLY if:<br>([site]='SMCA' or [site]='UMCB') and ([c_iaroutetpa]='1' or (([treatment_type(IAtPA)]='1' or [treatment_type(MER)]='1' or [gs_iatpa]='1') and [jc_princicd10procedure]<>'')) and [hide_nonessential_fields(1)]='0' | Is there documentation in the medical record that the first endovascular treatment procedure was initiated greater than 8 hours after arrival at this hospital? | radio<br>1 Yes<br>2 No<br><br>Custom alignment: RH |
| 535 | c_skin<br><br>Show the field ONLY if:<br>([site]='UMCB' or [site]='SMCA') and [hide_nonessential_fields(1)]='0' | Is there documentation of skin puncture at this hospital to access the arterial site selected for endovascular treatment of a cerebral artery occlusion? | radio<br>1 Yes<br>2 No<br><br>Custom alignment: RH<br>Field Annotation: @DEFAULT='[treatment_type(MER):value]' |
| 536 | desc_artpunc_dt_precision<br><br>Show the field ONLY if:<br>[treatment_type(MER)]='1' and [hide_nonessential_fields(1)]='0' | Skin Puncture Date/Time Precision[artpunc_dt_precision]From: Initial Stroke InformationFormerly: Arterial Puncture Date/Time Precision | descriptive |
| 537 | c_artpuncdt_precision<br><br>Show the field ONLY if:<br>if([site]='SMCA' or [site]='UMCB', [c_skin] = '1', [site]='SMCW') or [c_artpuncdt_precision]<>'' and [hide_nonessential_fields(1)]='0' | Skin Puncture Date/Time Precision | dropdown<br>5 Date & Time (MM/DD/YYYY HH24:MM)<br>3 Date Only (MM/DD/YYYY)<br>0 Unknown<br><br>Custom alignment: RH<br>Field Annotation: @DEFAULT='[artpunc_dt_precision:value]' |

| | | | |
|---|---|---|---|
| 538 | desc_artpunc_dt<br><br>Show the field ONLY if:<br>[treatment_type(MER)]='1' and [art punc_dt_precision] <> '0' and [hid e_nonessential_fields(1)]='0' | What is the date and time of skin puncture at this hospital to access the arterial site selected for endovascular treatment of a cerebral artery occlusion?Formerly: Arterial Puncture Date/Time[artpunc_dt]From: Initial Stroke Information▲ ▼ copy the date/time into the field below | descriptive, Identifier |
| 539 | c_artpuncdt<br><br>Show the field ONLY if:<br>if([site]='SMCA' or [site]='UMCB', [c _skin] = '1' and [c_artpuncdt_preci sion] <> '0', [site]='SMCW') or [c_ar tpuncdt]<>'' and [hide_nonessenti al_fields(1)]='0' | What is the date and time of skin puncture at this hospital to access the arterial site selected for endovascular treatment of a cerebral artery occlusion? | text (datetime_mdy, Min: 2010-01-01 00:00, Max: 2099-12-31 23:59), Identifier<br>Custom alignment: RH<br>Field Annotation: @DEFAULT='[artpunc_dt]' |
| 540 | c_ivtpaprior<br><br>Show the field ONLY if:<br>([site]='SMCA' or [site]='UMCB') an d [gs_age]>=18 and [gs_notadmit] ='2' and [jc_admitdate]<>'' and [gs _discdatetime]<>'' and [jc_princicd 10diagnosis]<>'' and [jc_princicd1 0procedure]<>'' and ([treatment_t ype(IVtPA)]='1' or [treatment_type (IAtPA)]='1' or [treatment_type(ME R)]='1') and [hide_nonessential_fiel ds(1)]='0' | Did the patient receive intravenous (IV) thrombolytic (t-PA) therapy at this hospital or a transferring hospital prior to receiving intra-arterial (IA) thrombolytic therapy or mechanical reperfusion therapy at this hospital? | radio<br>1 Yes<br>2 No<br><br>Custom alignment: RH |
| 541 | mer_proc<br><br>Show the field ONLY if:<br>([site]='SMCA' or [site]='SMCW' or [site]='UMCB') and [gs_stroketype] ='2' | Was a mechanical endovascular reperfusion procedure attempted during this episode of care (at this hospital)? | radio<br>1 Yes<br>2 No<br><br>Custom alignment: RH |
| 542 | c_failedthrom<br><br>Show the field ONLY if:<br>([site] = 'SMCA' or [site] = 'SMCW' o r [site] = 'UMCB') and ([gs_strokety pe] = '2' or starts_with([jc_princicd 10diagnosis],'i63')) and [hide_none ssential_fields(1)]='0' | Was a mechanical thrombectomy procedure attempted but unsuccessful or aborted before removal of the LVO? | radio<br>1 Yes<br>2 No<br><br>Custom alignment: RH |
| 543 | mer_notperf<br><br>Show the field ONLY if:<br>[mer_proc] = '2' | Are reasons for not performing mechanical endovascular reperfusion therapy documented? | radio<br>1 Yes<br>2 No<br><br>Custom alignment: RH |

<table>
<tr><td>544</td><td>mer_reasons<br><br>Show the field ONLY if:<br>[mer_notperf] = '1'</td><td>Reasons for not performing mechanical endovascular reperfusion therapy (select all that apply):<br>*These reasons do not exclude from measure population*</td><td>checkbox
<table>
<tr><td>1</td><td>mer_reasons___1</td><td>Significant pre-stroke disability (pre-stroke mRS > 1)</td></tr>
<tr><td>2</td><td>mer_reasons___2</td><td>No evidence of proximal occlusion</td></tr>
<tr><td>3</td><td>mer_reasons___3</td><td>NIHSS < 6</td></tr>
<tr><td>4</td><td>mer_reasons___4</td><td>Brain imaging not favorable/hemorrhage transformation (ASPECTS score < 6)</td></tr>
<tr><td>5</td><td>mer_reasons___5</td><td>Groin puncture could not be initiated within 6 hours of symptom onset</td></tr>
<tr><td>6</td><td>mer_reasons___6</td><td>Anatomical reason - unfavorable vascular anatomy that limits access to the occluded artery</td></tr>
<tr><td>7</td><td>mer_reasons___7</td><td>Patient/family refusal</td></tr>
<tr><td>8</td><td>mer_reasons___8</td><td>MER performed at outside hospital</td></tr>
<tr><td>15</td><td>mer_reasons___15</td><td>Allergy to contrast material</td></tr>
<tr><td>9</td><td>mer_reasons___9</td><td>Equipment-related delay *</td></tr>
<tr><td>10</td><td>mer_reasons___10</td><td>No endovascular specialist available *</td></tr>
<tr><td>11</td><td>mer_reasons___11</td><td>Delay in stroke diagnosis *</td></tr>
<tr><td>12</td><td>mer_reasons___12</td><td>Vascular imaging not performed *</td></tr>
<tr><td>13</td><td>mer_reasons___13</td><td>Advanced Age *</td></tr>
<tr><td>14</td><td>mer_reasons___14</td><td>Other *</td></tr>
</table></td></tr>
<tr><td>545</td><td>mer_reasoth<br><br>Show the field ONLY if:<br>[mer_reasons(14)] = '1'</td><td>Other reason for not performing MER:<br>*50 character limit*</td><td>text<br>Field Annotation: @CHARLIMIT=50</td></tr>
</table>

| | | | |
|---|---|---|---|
| 546 | c_iatype<br><br>Show the field ONLY if:<br>([site] = 'SMCA' or [site] = 'SMCW' or [site] = 'UMCB') and [gs_iatpa] = '1' and [hide_nonessential_fields(1)]='0' | If MER treatment at this hospital, type of treatment: | checkbox<br>2 c_iatype___2 Retrievable stent<br>3 c_iatype___3 Other mechanical clot retrieval device beside stent retrieval<br>4 c_iatype___4 Clot suction device<br>5 c_iatype___5 Intracranial angioplasty, with or without permanent stent<br>6 c_iatype___6 Cervical carotid angioplasty, with or without permanent stent<br>7 c_iatype___7 Other |
| 547 | c_firstpass<br><br>Show the field ONLY if:<br>([site]='SMCA' or [site]='SMCW' or [site]='UMCB') and [gs_age]>=18 and [gs_notadmit]='2' and [jc_admitdate]<>'' and [gs_discdatetime]<>'' and starts_with([jc_princicd10diagnosis],'i63') and [jc_carotid]='2' and (([jc_princicd10procedure]<>'' and [treatment_type(MER)]='1') or [mer_proc]='1') | Is there documentation in the medical record of the first pass of a mechanical reperfusion device to remove a clot occluding a cerebral artery at this hospital? | radio<br>1 Yes<br>2 No<br><br>Custom alignment: RH |
| 548 | desc_firstpassdt_precision<br><br>Show the field ONLY if:<br>[treatment_type(MER)]='1' | First Pass Date/Time Precision[firstpassdt_precision]From: Initial Stroke Information | descriptive |
| 549 | c_firstpassdt_precision<br><br>Show the field ONLY if:<br>((([site] = 'SMCA' or [site] = 'UMCB') and [c_firstpass] = '1') or [site] = 'SMCW') or [c_firstpassdt_precision]<>'' | First Pass Date/Time Precision | dropdown<br>5 Date & Time (MM/DD/YYYY HH24:MM)<br>3 Date Only (MM/DD/YYYY)<br>0 Unknown<br><br>Custom alignment: RH<br>Field Annotation: @DEFAULT='[firstpassdt_precision:value]' |
| 550 | desc_firstpassdt<br><br>Show the field ONLY if:<br>[treatment_type(MER)]='1' and [firstpassdt_precision] <> '0' | What is the date and time of the first pass of a clot retrieval device at this hospital?From: Initial Stroke Information[firstpassdt]▲ ▼ copy the date/time into the field below | descriptive, Identifier |

| | | | |
|---|---|---|---|
| 551 | c_firstpassdt<br><br>Show the field ONLY if:<br>(((([site] = 'SMCA' or [site] = 'UMCB') and [c_firstpass] = '1') or [site] = 'SMCW') and [c_firstpassdt_precision] <> '0') or [c_firstpassdt]<>'' | What is the date and time of the first pass of a clot retrieval device at this hospital? | text (datetime_mdy, Min: 2010-01-01 00:00, Max: 2099-12-31 23:59), Identifier<br>Custom alignment: RH<br>Field Annotation: @DEFAULT='[firstpassdt]' |
| 552 | mer_delay<br><br>Show the field ONLY if:<br>([site] = 'SMCA' or [site] = 'SMCW' or [site] = 'UMCB') and [hide_nonessential_fields(1)]='0' | Is a cause(s) for delay in performing mechanical endovascular reperfusion therapy documented? | radio<br>1 \| Yes<br>2 \| No<br><br>Custom alignment: RH |
| 553 | mer_reasonsdelay<br><br>Show the field ONLY if:<br>[mer_delay] = '1' and [hide_nonessential_fields(1)]='0' | Reasons for delay:<br>** These reasons do not exclude the patient from the MER Time to EVT measures.* | checkbox<br>1 \| mer_reasonsdelay___1 \| Social/religious<br>2 \| mer_reasonsdelay___2 \| Initial refusal<br>3 \| mer_reasonsdelay___3 \| Care-team unable to determine eligibility<br>6 \| mer_reasonsdelay___6 \| Management of concomitant emergent/acute conditions such as cardiopulmonary arrest, respiratory failure (requiring intubation)<br>7 \| mer_reasonsdelay___7 \| Investigational or experimental protocol for thrombolysis<br>14 \| mer_reasonsdelay___14 \| Additional proximal vascular procedure required prior to first-pass (stent)<br>15 \| mer_reasonsdelay___15 \| Need for additional PPE for suspected/confirmed infectious disease<br>8 \| mer_reasonsdelay___8 \| Delay in stroke diagnosis *<br>9 \| mer_reasonsdelay___9 \| In-hospital time delay *<br>10 \| mer_reasonsdelay___10 \| Equipment-related delay *<br>12 \| mer_reasonsdelay___12 \| Need for additional imaging*<br>13 \| mer_reasonsdelay___13 \| Catheter lab not available*<br>11 \| mer_reasonsdelay___11 \| Other * |

| | | | |
|---|---|---|---|
| 554 | mer_othreasons<br><br>Show the field ONLY if:<br>[mer_reasonsdelay(11)] = '1' and [hide_nonessential_fields(1)]='0' | Other reason(s) for delay:<br>*50 character limit* | text<br>Field Annotation: @CHARLIMIT=50 |
| 555 | c_proxdist<br><br>Show the field ONLY if:<br>([site] = 'SMCA' or [site] = 'UMCB') and ([treatment_recvd]='1' or [gs_notadmit]<>'1') and [hide_nonessential_fields(1)]='0' | What is the location of the clot in the cerebral circulation? | dropdown<br>1 The clot location is proximal<br>2 The clot location is distal<br>3 Neither proximal or distal are documented in reference to the location of the clot in the cerebral circulation, OR unable to determine (UTD) from the medical record documentation<br><br>Custom alignment: RH |
| 556 | c_cartocc<br><br>Show the field ONLY if:<br>([site] = 'SMCA' or [site] = 'UMCB') and ([treatment_recvd]='1' or [gs_notadmit]<>'1') and [hide_nonessential_fields(1)]='0' | What cerebral artery is occluded? | dropdown<br>1 Anterior cerebral artery (ACA)<br>2 A1 ACA<br>3 Anterior communicating artery<br>4 Internal carotid artery (ICA)<br>5 ICA terminus (T-lesion; T occlusion)<br>6 Middle cerebral artery (MCA)<br>7 M1 MCA<br>8 M2 MCA<br>9 M3/M4 MCA<br>10 Vertebral artery (VA)<br>11 Basilar artery (BA)<br>12 Posterior cerebral artery (PCA)<br>13 Other cerebral artery branch/segment<br>14 The clinical location of the primary occluded vessel was not documented, OR unable to determine (UTD) from the medical record documentation<br><br>Custom alignment: RH |

| | | | |
|---|---|---|---|
| 557 | c_ticigrade<br><br>Show the field ONLY if:<br>([site] = 'SMCA' or [site] = 'SMCW' or [site] = 'UMCB') and ([treatment_recvd]='1' or [gs_notadmit]<>'1') and [hide_nonessential_fields(1)]='0' | Thrombolysis in Cerebral Infarction (TICI) Post-Treatment Reperfusion Grade | radio<br>1 \| Grade 0<br>2 \| Grade 1<br>3 \| Grade 2a<br>4 \| Grade 2b<br>5 \| Grade 3<br>6 \| ND<br><br>Custom alignment: LH |
| 558 | c_ticigrade2<br><br>Show the field ONLY if:<br>([site] = 'SMCA' or [site] = 'UMCB') and ([treatment_recvd]='1' or [gs_notadmit]<>'1') and [hide_nonessential_fields(1)]='0' | Is there a documented TICI reperfusion gradepost-treatment? | dropdown<br>1 \| 1 - A TICI reperfusion grade greater than or equal to ( >= ) 2B was documented post-treatment<br>2 \| 2 - A TICI reperfusion grade less than ( < ) 2B was documented post-treatment<br>3 \| 3 - A TICI reperfusion grade was not done post-treatment, OR Unable to determine (UTD) from the medical record documentation<br><br>Custom alignment: RH |
| 559 | mer_gradenotach<br><br>Show the field ONLY if:<br>[mer_proc] = '1' and [mer_ticidt] = '' and [hide_nonessential_fields(1)]='0' | TICI Grade 2b or 3 not achieved | radio<br>1 \| not achieved<br><br>Custom alignment: RH<br>Field Annotation: @HIDDEN |
| 560 | desc_ticidt_precision<br><br>Show the field ONLY if:<br>[treatment_type(MER)]='1' and [hide_nonessential_fields(1)]='0' | Date/time TICI grade 2B or 3 Precision[ticidt_precision]From: Initial Stroke Information | descriptive |
| 561 | mer_ticidt_precision<br><br>Show the field ONLY if:<br>([mer_proc]='1' and [mer_gradenotach]='') or [mer_ticidt_precision]<>'' and [hide_nonessential_fields(1)]='0' | Date/time TICI first documented Precision | dropdown<br>5 \| Date & Time (MM/DD/YYYY HH24:MM)<br>3 \| Date Only (MM/DD/YYYY)<br>0 \| Unknown<br><br>Custom alignment: RH<br>Field Annotation: @DEFAULT='[ticidt_precision:value]' |

| | | | |
|---|---|---|---|
| 562 | desc_ticidt<br><br>Show the field ONLY if:<br>[treatment_type(MER)]='1' and [ticidt_precision] <> '0' and [hide_nonessential_fields(1)]='0' | What was the date and time that a TICI 2B/3 was first documented during the mechanical thrombectomy procedure?From: Initial Stroke Information[ticidt]▲ ▼ copy the date/time into the field below | descriptive, Identifier<br>Field Annotation: @DEFAULT='[ticidt]' |
| 563 | mer_ticidt<br><br>Show the field ONLY if:<br>([mer_proc]='1' and [mer_gradenotach]='' and [mer_ticidt_precision]<>'0') or [mer_ticidt]<>'' and [hide_nonessential_fields(1)]='0' | What was the date and time that a TICI was first documented during the mechanical thrombectomy procedure? | text (datetime_mdy, Min: 2015-01-01 00:00, Max: 2099-12-31 23:59), Identifier |
| 564 | c_neuroimaging36h<br><br>Show the field ONLY if:<br>([site] = 'SMCA' or [site]='SMCH' or [site]='SMCW' or [site] = 'UMCB') and ([treatment_recvd]='1' or [gs_notadmit]<>'1') and [hide_nonessential_fields(1)]='0' | Section Header: *Complications*<br><br>Was there a positive finding on brain imaging of parenchymal hematoma, SAH, and/or IVH following IV or IA thrombolytic (t-PA) therapy, or mechanical endovascular reperfusion therapy initiation? | radio<br>1 Yes<br>2 No<br><br>Custom alignment: RH |
| 565 | c_posbrain_precision<br><br>Show the field ONLY if:<br>[c_neuroimaging36h] = '1' and [hide_nonessential_fields(1)]='0' | Positive Brain Image Date/Time Precision | dropdown<br>5 Date & Time (MM/DD/YYYY HH24:MM)<br>3 Date Only (MM/DD/YYYY)<br>0 Unknown<br><br>Custom alignment: RH |
| 566 | c_posbrain<br><br>Show the field ONLY if:<br>[c_neuroimaging36h] = '1' and [c_posbrain_precision] <> '0' and [hide_nonessential_fields(1)]='0' | Positive Brain Image Date/Time | text (datetime_mdy, Min: 2010-01-01 00:00, Max: 2099-12-31 23:59), Identifier<br>Custom alignment: RH |

| | | | |
|---|---|---|---|
| 567 | c_posbrain_result<br><br>Show the field ONLY if:<br>[c_neuroimaging36h] = '1' and [hide_nonessential_fields(1)]='0' | Results of positive brain image<br>*Select all that apply.* | checkbox<br>1 \| c_posbrain_result___1 \| PH2 (Parenchymal Hematoma Type 2)<br>2 \| c_posbrain_result___2 \| IVH (Intraventricular Hemorrhage)<br>3 \| c_posbrain_result___3 \| SAH (Subarachnoid Hemorrhage)<br>4 \| c_posbrain_result___4 \| RIH (Remote site of intraparenchymal hemorrhage outside the area of infarction)<br>6 \| c_posbrain_result___6 \| Other positive finding not listed above<br>5 \| c_posbrain_result___5 \| Not documented<br><br>Field Annotation: @NONEOFTHEABOVE='5' |
| 568 | c_nihss36ivutd<br><br>Show the field ONLY if:<br>([site] = 'SMCA' or [site] = 'UMCB') and [c_posbrain] <> '' and [hide_nonessential_fields(1)]='0' | Unable to determine Highest NIHSS Score Documented Within 36 Hours Following IV Thrombolytic Initiation | radio<br>1 \| UTD<br><br>Custom alignment: RH |
| 569 | c_nihss36iv<br><br>Show the field ONLY if:<br>([site] = 'SMCA' or [site] = 'UMCB') and [c_posbrain] <> '' and [hide_nonessential_fields(1)]='0' | Highest NIHSS Score Documented Within 36 Hours Following IV Thrombolytic Initiation | text (integer, Min: 0, Max: 42)<br>Custom alignment: RH |
| 570 | c_nihssiatpautd<br><br>Show the field ONLY if:<br>([site]='SMCA' or [site]='SMCW' or [site]='UMCB') and [c_nihssiatpamer]='' and [hide_nonessential_fields(1)]='0' | Unable to determine the last NIHSS score documented prior to initiation of IA t-PA or MER at this hospital. | radio<br>1 \| UTD<br><br>Custom alignment: RH |
| 571 | mer_scoreobt<br><br>Show the field ONLY if:<br>([site]='SMCA' or [site]='SMCW' or [site]='UMCB') and [mer_proc]='1' and [c_nihssiatpautd]<>'1' | MER score obtained from:<br>*If Baseline NIHSS, use [gs_nihssadmit] below.* | radio<br>1 \| Baseline NIHSS<br>2 \| Subsequent NIHSS<br><br>Custom alignment: RH |

| | | | |
|---|---|---|---|
| 572 | c_nihssiatpamer<br>Show the field ONLY if:<br>([site]='SMCA' or [site]='SMCW' or [site]='UMCB') and [c_nihssiatpautd]<>'1' and [hide_nonessential_fields(1)]='0' | What is the last NIHSS score documented prior to initiation of IA thrombolytic or MER at this hospital? | text (integer, Min: 0, Max: 42)<br>Custom alignment: RH |
| 573 | c_nihss36iautd<br>Show the field ONLY if:<br>([site] = 'SMCA' or [site] = 'UMCB') and [c_posbrain] <> '' and [hide_nonessential_fields(1)]='0' | Unable to determine Highest NIHSS Score Documented Within 36 Hours Following IA t-PA or MER Initiation | radio<br>1 UTD<br>Custom alignment: RH |
| 574 | c_nihss36ia<br>Show the field ONLY if:<br>([site] = 'SMCA' or [site] = 'UMCB') and [c_posbrain] <> '' and [hide_nonessential_fields(1)]='0' | Highest NIHSS Score Documented Within 36 Hours Following IA t-PA or MER Initiation | text (integer, Min: 0, Max: 42)<br>Custom alignment: RH |
| 575 | c_procoagulant<br>Show the field ONLY if:<br>([site]='SMCA' or [site]='SMCH' or [site]='SMCW' or [site]='UMCB') and [gs_stroketype]='5' and ([treatment_recvd]='1' or [gs_notadmit]<>'1') and [hide_nonessential_fields(1)]='0' | Is there documentation that a procoagulant reversal agent was initiated at this hospital? | radio<br>1 Yes<br>2 No<br>Custom alignment: RH |
| 576 | ich_procoagulant_type<br>Show the field ONLY if:<br>[site]='UMCB' and [c_procoagulant]='1' | ✪ If yes, select reversal agent administered (at this hospital): | radio<br>1 Prothrombin complex concentrates (PCC)<br>2 Fresh Frozen Plasma (FFP)<br>3 Factor Xa Reversal - Andexxa (andexanet alfa)<br>4 Pradaxa Reversal - Praxbind (idarucizumab)<br>5 Other factor complexes (including anti-inhibitor coagulant complex, factor IX complex)<br>6 Other {ich_procoagulant_typeoth:icons} |
| 577 | ich_procoagulant_typeoth<br>Show the field ONLY if:<br>[ich_procoagulant_type]='6' | ✪ Specify Reversal Agent Administered - Other | text |

| | | | |
|---|---|---|---|
| 578 | c_procoagulant_dt_precision<br><br>Show the field ONLY if:<br>[c_procoagulant]='1' and [hide_nonessential_fields(1)]='0' | Date/Time Procoagulant Initiated Precision | dropdown<br>5 Date & Time (MM/DD/YYYY HH24:MM)<br>3 Date Only (MM/DD/YYYY)<br>0 Unknown<br><br>Custom alignment: RH |
| 579 | c_procoagulant_dt<br><br>Show the field ONLY if:<br>[c_procoagulant]='1' and [c_procoagulant_dt_precision]<>'0' and [hide_nonessential_fields(1)]='0' | Date/Time Procoagulant Initiated | text (datetime_mdy, Min: 2010-01-01 00:00, Max: 2099-12-31 23:59), Identifier<br>Custom alignment: RH |
| 580 | ich_procoagulant_prior<br><br>Show the field ONLY if:<br>[site]='UMCB' | ✪ Was a procoagulant reversal agent administered to the patient prior to arrival at your hospital (in MSU or first hospital): | radio<br>1 Yes<br>2 No<br><br>Custom alignment: RH |
| 581 | ich_procoagulant_priortype<br><br>Show the field ONLY if:<br>[ich_procoagulant_prior]='1' | ✪ If yes, select reversal agent administered (prior to arrival): | radio<br>1 Prothrombin complex concentrates (PCC)<br>2 Fresh Frozen Plasma (FFP)<br>3 Factor Xa Reversal - Andexxa (andexanet alfa)<br>4 Pradaxa Reversal - Praxbind (idarucizumab)<br>5 Other factor complexes (including anti-inhibitor coagulant complex, factor IX complex)<br>6 Other {ich_procoagulant_priortypeoth:icons} |
| 582 | ich_procoagulant_priortypeoth<br><br>Show the field ONLY if:<br>[ich_procoagulant_priortype]='6' | ✪ Specify Reversal Agent Administered (prior to arrival) - Other | text |
| 583 | ich_vitk<br><br>Show the field ONLY if:<br>[site]='UMCB' | ✪ Was Vitamin K administered to this patient? | radio<br>1 Yes, prior to arrival (in MSU or first hospital)<br>2 Yes, at your hospital<br>3 No |

| | | | |
|---|---|---|---|
| 584 | c_reasnopro<br><br>Show the field ONLY if:<br>([site]='SMCA' or [site]='SMCH' or [site]='SMCW' or [site]='UMCB') and [gs_stroketype]='5' and ([treatment_recvd]='1' or [gs_notadmit]<>'1') and [hide_nonessential_fields(1)]='0' | Is there documentation by a physician/APN/PA or pharmacist in the medical record of a reason for not administering a procoagulant reversal agent? | radio<br>1 Yes<br>2 No<br><br>Custom alignment: RH |
| 585 | ich_doac<br><br>Show the field ONLY if:<br>[site]='UMCB' and ([c_procoagulant]='2' or [ich_procoagulant_prior]='2') | ✪ If patient is currently taking DOACs, is there documentation in the medical record of a reason for not administering an anticoagulant reversal agent? | radio<br>1 Yes, medical reason<br>2 Yes, patient reason<br>3 Yes, system reason<br>4 No |
| 586 | c_inraftertx<br><br>Show the field ONLY if:<br>[c_procoagulant]='1' and [hide_nonessential_fields(1)]='0' | No documented INR < = 1.4 after tx | radio<br>1 ND<br><br>Custom alignment: RH |
| 587 | c_inr_dt_precision<br><br>Show the field ONLY if:<br>[c_procoagulant]='1' and [c_inraftertx]<>'1' and [hide_nonessential_fields(1)]='0' | INR < = 1.4 after tx Date/Time Precision | dropdown<br>5 Date & Time (MM/DD/YYYY HH24:MM)<br>3 Date Only (MM/DD/YYYY)<br>0 Unknown<br><br>Custom alignment: RH |
| 588 | c_inr_dt<br><br>Show the field ONLY if:<br>[c_procoagulant]='1' and [c_inraftertx]<>'1' and [c_inr_dt_precision]<>'0' and [hide_nonessential_fields(1)]='0' | If initial INR > 1.4 and treated with procoagulant, Date/Time first INR < = 1.4 after treatment: | text (datetime_mdy, Min: 2010-01-01 00:00, Max: 2099-12-31 23:59), Identifier<br>Custom alignment: RH |
| 589 | c_nimodipine<br><br>Show the field ONLY if:<br>([site]='SMCA' or [site]='SMCH' or [site]='SMCW' or [site]='UMCB') and [gs_age]>=18 and [jc_admitdate]<>'' and starts_with([jc_princicd10diagnosis],'i60') and [jc_clinical]='2' and [hide_nonessential_fields(1)]='0' | Section Header: *Hemorrhagic Stroke Treatment*<br><br>Is there documentation that nimodipine was administered at this hospital within 24 hours of diagnosis? | radio<br>1 Yes<br>2 No<br><br>Custom alignment: RH |

| | | | |
|---|---|---|---|
| 590 | c_nimodipine_dt_precision<br><br>Show the field ONLY if:<br>([site]='SMCA' or [site]='SMCH' or [site]='SMCW' or [site]='UMCB') and [c_nimodipine]='1' and [hide_nonessential_fields(1)]='0' | Nimodipine Administration Date/Time Precision | dropdown<br>5 \| Date & Time (MM/DD/YYYY HH24:MM)<br>3 \| Date Only (MM/DD/YYYY)<br>0 \| Unknown<br><br>Custom alignment: RH |
| 591 | c_nimodipine_dt<br><br>Show the field ONLY if:<br>([site]='SMCA' or [site]='SMCH' or [site]='SMCW' or [site]='UMCB') and [c_nimodipine]='1' and [c_nimodipine_dt_precision]<>'0' and [hide_nonessential_fields(1)]='0' | What is the date and time that nimodipine was first administered to this patient at this hospital? | text (datetime_mdy, Min: 2010-01-01 00:00, Max: 2099-12-31 23:59), Identifier<br>Custom alignment: RH |
| 592 | c_reasonnonim<br><br>Show the field ONLY if:<br>([site]='SMCA' or [site]='SMCH' or [site]='SMCW' or [site]='UMCB') and [c_nimodipine]='2' and [hide_nonessential_fields(1)]='0' | Is there documentation by a physician/APN/PA or pharmacist in the medical record of a reason for not administering nimodipine treatment? | radio<br>1 \| Yes<br>2 \| No<br><br>Custom alignment: RH |
| 593 | c_surgicalich<br><br>Show the field ONLY if:<br>([site] = 'SMCA' or [site] = 'UMCB') and [gs_stroketype] = '5' and ([treatment_recvd]='1' or [gs_notadmit]<>'1') and [hide_nonessential_fields(1)]='0' | Surgical treatment for ICH at this hospital? | radio<br>1 \| Yes<br>2 \| No<br><br>Custom alignment: RH |
| 594 | c_surgicalich_type<br><br>Show the field ONLY if:<br>([site] = 'SMCA' or [site] = 'UMCB') and [c_surgicalich] = '1' and ([treatment_recvd]='1' or [gs_notadmit]<>'1') and [hide_nonessential_fields(1)]='0' | Type of surgical treatment for ICH | checkbox<br>1 \| c_surgicalich_type___1 \| External Ventricular Drain (EVD)<br>2 \| c_surgicalich_type___2 \| Endoscopic evacuation<br>3 \| c_surgicalich_type___3 \| Conventional craniotomy and evacuation of clot under direct vision<br>4 \| c_surgicalich_type___4 \| Stereotaxic evacuation<br>5 \| c_surgicalich_type___5 \| Hemicraniectomy without clot evacuation<br>6 \| c_surgicalich_type___6 \| Fibrinolytic infusion via catheter<br>7 \| c_surgicalich_type___7 \| Other |

| | | | |
|---|---|---|---|
| 595 | ich_surgdt_precision<br><br>Show the field ONLY if:<br>[site]='UMCB' and [c_surgicalich]='1' | ✪ Surgical treatment for ICH date/time Precision | dropdown<br>5 Date & Time (MM/DD/YYYY HH24:MM)<br>3 Date Only (MM/DD/YYYY)<br>0 Unknown |
| 596 | ich_surgdt<br><br>Show the field ONLY if:<br>[site]='UMCB' and [c_surgicalich]='1' and [ich_surgdt_precision]<>'0' | ✪ If surgical treatment for ICH at this hospital is yes, what was the procedure date/ time? | text (datetime_mdy, Min: 2010-01-01 00:00, Max: 2099-12-31 23:59) |
| 597 | evac_proc_startdt<br><br>Show the field ONLY if:<br>([site] = 'SMCA' or [site] = 'UMCB') and [c_surgicalich] = '1' and [hide_nonessential_fields(1)]='0' | Evacuation procedure start date/time | text (datetime_mdy) |
| 598 | c_surgicalich_hr<br><br>Show the field ONLY if:<br>([site] = 'SMCA' or [site] = 'UMCB') and [c_surgicalich] = '1' and [hide_nonessential_fields(1)]='0' | If ICH was evacuated, time from ictus to evacuation procedure start was: *(Hours)* | text (number)<br>Custom alignment: RH |
| 599 | ich_corti<br><br>Show the field ONLY if:<br>[site]='UMCB' | ✪ Was the patient treated with an IV or oral corticosteroid? | radio<br>1 Yes, prior to arrival (in MSU or first hospital)<br>2 Yes, at your hospital<br>3 No |
| 600 | ich_reasoncorti<br><br>Show the field ONLY if:<br>[ich_corti]='1' or [ich_corti]='2' | ✪ Documentation of a neurological or other medical reason for prescribing corticosteroids: | radio<br>1 Yes<br>2 No/ND<br><br>Custom alignment: RH |
| 601 | advanced_stroke_care_complete | Section Header: *Form Status*<br><br>Complete? | dropdown<br>0 Incomplete<br>1 Unverified<br>2 Complete |

Instrument: **Additional Stroke Information** (additional_stroke_information)

Collapse

| | | | |
|---|---|---|---|
| 602 | aneurysmal_sah<br><br>Show the field ONLY if:<br>([site]='SMCA' or [site]='SMCH' or [site]='SMCW' or [site]='UMCB') and [gs_stroketype]='4' and [gs_notadmit]<>'1' and [hide_nonessential_fields(1)]='0' | Was this an Aneurysmal SAH? | yesno<br>1 Yes<br>0 No<br><br>Custom alignment: RH |
| 603 | procedures_interventions<br><br>Show the field ONLY if:<br>([site]='SMCA' or [site]='SMCH' or [site]='SMCW' or [site]='UMCB') and [gs_notadmit]<>'1' and [hide_nonessential_fields(1)]='0' | Section Header: *Other Stroke Procedures Performed*<br><br>✪ Surgical Interventions or Other Procedures Performed | checkbox<br>1 procedures_interventions___1 Clipping<br>2 procedures_interventions___2 Coiling<br>3 procedures_interventions___3 Crani<br>4 procedures_interventions___4 Embolization<br>5 procedures_interventions___5 EVD<br>6 procedures_interventions___6 Neuro Angiography<br>0 procedures_interventions___0 Other Stroke Procedure |
| 604 | procedures_other<br><br>Show the field ONLY if:<br>[procedures_interventions(0)]='1' and [hide_nonessential_fields(1)]='0' | ✪ If other stroke procedure, specify: | text<br>Custom alignment: RH |
| 605 | informed_consent_proc<br><br>Show the field ONLY if:<br>([treatment_type(MER)]='1' or [gs_iatpa]='1' or [procedures_interventions(1)]='1' or [procedures_interventions(2)]='1' or [procedures_interventions(3)]='1' or [procedures_interventions(4)]='1' or [procedures_interventions(5)]='1' or [procedures_interventions(6)]='1' or [procedures_interventions(0)]='1') and [hide_nonessential_fields(1)]='0' | Section Header: *Informed Consent Documentation for ANY invasive procedure.*<br><br>Did physician document either that informed consent was discussed with the patient, or that this was an emergent procedure? | yesno<br>1 Yes<br>0 No<br><br>Custom alignment: RH |
| 606 | dnv_18_rupt<br><br>Show the field ONLY if:<br>([site]='SMCA' or [site]='SMCH' or [site]='SMCW' or [site]='UMCB') and ([procedures_interventions(1)]='1' or [procedures_interventions(2)]='1') and [hide_nonessential_fields(1)]='0' | Section Header: *DNV 18: Complications for aneurysm Clipping and/or Coiling*<br><br>Was initial Aneurysm Ruptured or Unruptured? | dropdown<br>1 Ruptured<br>2 Unruptured<br><br>Custom alignment: RH |

| | | | |
|---|---|---|---|
| 607 | dnv_18_complications<br><br>Show the field ONLY if:<br>([site]='SMCA' or [site]='SMCH' or [site]='SMCW' or [site]='UMCB') and ([procedures_interventions(1)] ='1' or [procedures_interventions(2)]='1') and [hide_nonessential_fields(1)]='0' | Did any of the following complications occur?<br>1. Stroke, bleeding or death within 24 hours of the procedure.<br>(Exclude bleeds within 24 hours for ruptured aneurysm patients.)<br>2. Re-bleeding and/or second treatment for residual aneurysm within 30 days of the procedure? | yesno<br>1 Yes<br>0 No<br><br>Custom alignment: RH |
| 608 | dnv_18_bleed<br><br>Show the field ONLY if:<br>[dnv_18_complications] = '1' and [hide_nonessential_fields(1)]='0' | If re-bleeding occurred within 30 days of the procedure, select the appropriate classification of bleed. Otherwise, select NA. | dropdown<br>0 NA<br>1 Pre-procedural<br>2 Procedural<br>3 Post-procedural<br><br>Custom alignment: RH |
| 609 | tpa_tool_nvs_num<br><br>Show the field ONLY if:<br>([site]='SMCA' or [site]='SMCH' or [site]='SMCW' or [site]='UMCB') and ([treatment_type(IAtPA)]='1' or [treatment_type(IVtPA)]='1' or [treatment_type(MER)]='1' or [gs_ivthroinit]='1' or [gs_ivtpaoutside]='1' or [gs_iatpa]='1' or [gs_iatpaoutside] ='1') and [hide_nonessential_fields(1)]='0' | Section Header: *Post IV Thrombolytic Monitoring Compliance*<br><br>✪ Neuro Vital Signs Numerator<br>*Number done correctly.* | text (integer), Required<br>Custom alignment: RH |
| 610 | tpa_tool_nvs_den<br><br>Show the field ONLY if:<br>([site]='SMCA' or [site]='SMCH' or [site]='SMCW' or [site]='UMCB') and ([treatment_type(IAtPA)]='1' or [treatment_type(IVtPA)]='1' or [treatment_type(MER)]='1' or [gs_ivthroinit]='1' or [gs_ivtpaoutside]='1' or [gs_iatpa]='1' or [gs_iatpaoutside] ='1') and [hide_nonessential_fields(1)]='0' | ✪ Neuro Vital Signs Denominator<br>*Number that should have been done.* | text (integer), Required<br>Custom alignment: RH |

| | | | |
|---|---|---|---|
| 611 | tpa_tool_vs_num<br><br>Show the field ONLY if:<br>([site]='SMCA' or [site]='SMCH' or [site]='SMCW' or [site]='UMCB') and ([treatment_type(IAtPA)]='1' or [treatment_type(IVtPA)]='1' or [treatment_type(MER)]='1' or [gs_ivthroinit]='1' or [gs_ivtpaoutside]='1' or [gs_iatpa]='1' or [gs_iatpaoutside]='1') and [hide_nonessential_fields(1)]='0' | ✪ Vital Signs Numerator<br>*Number done correctly.* | text (integer), Required<br>Custom alignment: RH |
| 612 | tpa_tool_vs_den<br><br>Show the field ONLY if:<br>([site]='SMCA' or [site]='SMCH' or [site]='SMCW' or [site]='UMCB') and ([treatment_type(IAtPA)]='1' or [treatment_type(IVtPA)]='1' or [treatment_type(MER)]='1' or [gs_ivthroinit]='1' or [gs_ivtpaoutside]='1' or [gs_iatpa]='1' or [gs_iatpaoutside]='1') and [hide_nonessential_fields(1)]='0' | ✪ Vital Signs Denominator<br>*Number that should have been done.* | text (integer), Required<br>Custom alignment: RH |
| 613 | post_tpa_bp<br><br>Show the field ONLY if:<br>([site]='SMCA' or [site]='SMCH' or [site]='SMCW' or [site]='UMCB') and ([treatment_type(IAtPA)]='1' or [treatment_type(IVtPA)]='1' or [gs_ivthroinit]='1' or [gs_ivtpaoutside]='1') and [hide_nonessential_fields(1)]='0' | ✪ During post-thrombolytic monitoring, were any Blood Pressures documented greater than 180/105? | yesno, Required<br>1 Yes<br>0 No<br><br>Custom alignment: RH |
| 614 | post_tpa_bp_addressed<br><br>Show the field ONLY if:<br>[post_tpa_bp]='1' and [hide_nonessential_fields(1)]='0' | ✪ During post-thrombolytic monitoring, were all Blood Pressures greater than 180/105 addressed appropriately? | yesno, Required<br>1 Yes<br>0 No<br><br>Custom alignment: RH |

| | | | |
|---|---|---|---|
| 615 | desc_tpa_tool_hidden<br><br>Show the field ONLY if:<br>[tpa_tool_md_num]<>'' or [tpa_tool_md_den]<>'' or [tpa_tool_handoff_num]<>'' or [tpa_tool_handoff_den]<>'' and [hide_nonessential_fields(1)]='0' | Hidden Fields (disabled as of 11/01/2019)Clinician Notification: (num/den)[tpa_tool_md_num]/[tpa_tool_md_den]<br>Post Thrombolytic Hand Off: (num/den)<br>[tpa_tool_handoff_num]/[tpa_tool_md_den] | descriptive |
| 616 | calc_post_tpa_compliance<br><br>Show the field ONLY if:<br>([site]='SMCA' or [site]='SMCH' or [site]='SMCW' or [site]='UMCB') and ([treatment_type(IAtPA)]='1' or [treatment_type(IVtPA)]='1' or [treatment_type(MER)]='1' or [gs_ivthroinit]='1' or [gs_ivtpaoutside]='1' or [gs_iatpa]='1' or [gs_iatpaoutside]='1') and [hide_nonessential_fields(1)]='0' | Post IV Thrombolytic Monitoring Compliance<br>*Percent* | calc<br>Calculation: if([tpa_tool_md_num]='' and [tpa_tool_md_den]='' and [tpa_tool_handoff_num]='' and [tpa_tool_handoff_den]='', round((([tpa_tool_nvs_num]/[tpa_tool_nvs_den])+([tpa_tool_vs_num]/[tpa_tool_vs_den]))/.02,1), round((([tpa_tool_nvs_num]/[tpa_tool_nvs_den])+([tpa_tool_vs_num]/[tpa_tool_vs_den])+([tpa_tool_md_num]/[tpa_tool_md_den])+([tpa_tool_handoff_num]/[tpa_tool_handoff_den]))/.04,1))<br>Custom alignment: RH |
| 617 | tpa_tool_notes<br><br>Show the field ONLY if:<br>([site]='SMCA' or [site]='SMCH' or [site]='SMCW' or [site]='UMCB') and ([treatment_type(IAtPA)]='1' or [treatment_type(IVtPA)]='1' or [treatment_type(MER)]='1' or [gs_ivthroinit]='1' or [gs_ivtpaoutside]='1' or [gs_iatpa]='1' or [gs_iatpaoutside]='1') and [hide_nonessential_fields(1)]='0' | ✪ Identify specific areas of Post Thrombolytic, Thrombectomy, and/or Endovascular Therapy Monitoring non-compliance | notes |
| 618 | ev_proc_type_1<br><br>Show the field ONLY if:<br>([site]='SMCA' or [site]='SMCH' or [site]='SMCW' or [site]='UMCB') and ([treatment_type(IAtPA)]='1' or [treatment_type(MER)]='1' or [gs_iatpa]='1' or [gs_iatpaoutside]='1' or [procedures_interventions(2)]='1' or [procedures_interventions(4)]='1' or [procedures_interventions(6)]='1') and [hide_nonessential_fields(1)]='0' | Section Header: *Post Thrombectomy or Endovascular Therapy Monitoring Compliance*<br><br>✪ Procedure Type | dropdown<br>1 Thrombectomy<br>2 Neuro Angiography<br>3 Coiling<br>4 Embolization<br>0 Other<br><br>Custom alignment: RH |

| | | | |
|---|---|---|---|
| 619 | ev_proc_orders_1<br><br>Show the field ONLY if: ([site]='SMCA' or [site]='SMCH' or [site]='SMCW' or [site]='UMCB') and ([treatment_type(IAtPA)]='1' or [treatment_type(MER)]='1' or [gs_iatpa]='1' or [gs_iatpaoutside]='1' or [procedures_interventions(2)]='1' or [procedures_interventions(4)]='1' or [procedures_interventions(6)]='1') and [hide_nonessential_fields(1)]='0' | ✪ Were pulse and site assessment orders present? | yesno<br>1 Yes<br>0 No<br><br>Custom alignment: RH |
| 620 | ev_proc_protocol_1<br><br>Show the field ONLY if: [ev_proc_orders_1]='0' and [hide_nonessential_fields(1)]='0' | ✪ If no orders were present, was unit specific protocol followed? | yesno<br>1 Yes<br>0 No<br><br>Custom alignment: RH |
| 621 | ev_proc_end_1<br><br>Show the field ONLY if: ([site]='SMCA' or [site]='SMCH' or [site]='SMCW' or [site]='UMCB') and ([treatment_type(IAtPA)]='1' or [treatment_type(MER)]='1' or [gs_iatpa]='1' or [gs_iatpaoutside]='1' or [procedures_interventions(2)]='1' or [procedures_interventions(4)]='1' or [procedures_interventions(6)]='1') and [hide_nonessential_fields(1)]='0' | ✪ Hemostasis Achieved Time<br>*Time from when assessments start.* | text (datetime_mdy, Min: 2010-01-01 00:00, Max: 2099-12-31 23:59) |
| 622 | ev_proc_depart_1<br><br>Show the field ONLY if: ([site]='SMCA' or [site]='SMCH' or [site]='SMCW' or [site]='UMCB') and ([treatment_type(IAtPA)]='1' or [treatment_type(MER)]='1' or [gs_iatpa]='1' or [gs_iatpaoutside]='1' or [procedures_interventions(2)]='1' or [procedures_interventions(4)]='1' or [procedures_interventions(6)]='1') and [hide_nonessential_fields(1)]='0' | ✪ Time patient left IR Suite | text (datetime_mdy, Min: 2010-01-01 00:00, Max: 2099-12-31 23:59) |

| | | | |
|---|---|---|---|
| 623 | ev_proc_ir_1<br><br>Show the field ONLY if: ([site]='SMCA' or [site]='SMCH' or [site]='SMCW' or [site]='UMCB') and ([treatment_type(IAtPA)]='1' or [treatment_type(MER)]='1' or [gs_iatpa]='1' or [gs_iatpaoutside]='1' or [procedures_interventions(2)]='1' or [procedures_interventions(4)]='1' or [procedures_interventions(6)]='1') and [hide_nonessential_fields(1)]='0' | ✪ Were all appropriate assessments done in IR Suite. | yesno<br>1 Yes<br>0 No<br><br>Custom alignment: RH |
| 624 | ev_proc_dest_1<br><br>Show the field ONLY if: ([site]='SMCA' or [site]='SMCH' or [site]='SMCW' or [site]='UMCB') and ([treatment_type(IAtPA)]='1' or [treatment_type(MER)]='1' or [gs_iatpa]='1' or [gs_iatpaoutside]='1' or [procedures_interventions(2)]='1' or [procedures_interventions(4)]='1' or [procedures_interventions(6)]='1') and [hide_nonessential_fields(1)]='0' | ✪ Patient destination | dropdown<br>1 DSMCUT - MICU<br>2 DSMCUT - SICU<br>16 DSMCUT - Imaging<br>13 DSMCUT - OR<br>3 DSMCUT - PACU<br>4 DSMCUT - 6th Floor<br>20 DSMCUT - Non-Stroke Unit<br>5 SMCA - ICU<br>17 SMCA - Imaging<br>14 SMCA - OR<br>19 SMCA - PACU<br>22 SMCA - 4 North<br>6 SMCA - 4 South/Southwest<br>7 SMCA - 6 North<br>21 SMCA - 7 North<br>8 SMCA - 6 South<br>9 SMCA - 3rd Floor<br>10 SMCW - ICU<br>18 SMCW - Imaging<br>15 SMCW - OR<br>11 SMCW - PACU<br>12 SMCW - 5th Floor |

| | | | |
|---|---|---|---|
| 625 | ev_proc_arrival_1<br><br>Show the field ONLY if:<br>([site]='SMCA' or [site]='SMCH' or [site]='SMCW' or [site]='UMCB') and ([treatment_type(IAtPA)]='1' or [treatment_type(MER)]='1' or [gs_iatpa]='1' or [gs_iatpaoutside]='1' or [procedures_interventions(2)]='1' or [procedures_interventions(4)]='1' or [procedures_interventions(6)]='1') and [hide_nonessential_fields(1)]='0' | ✪ Destination Arrival Time | text (datetime_mdy, Min: 2010-01-01 00:00, Max: 2099-12-31 23:59) |
| 626 | ev_proc_transit_1<br><br>Show the field ONLY if:<br>([site]='SMCA' or [site]='SMCH' or [site]='SMCW' or [site]='UMCB') and ([treatment_type(IAtPA)]='1' or [treatment_type(MER)]='1' or [gs_iatpa]='1' or [gs_iatpaoutside]='1' or [procedures_interventions(2)]='1' or [procedures_interventions(4)]='1' or [procedures_interventions(6)]='1') and [hide_nonessential_fields(1)]='0' | ✪ Were all appropriate assessments documented between IR Suite and patient destination? | radio<br>1 Yes<br>0 No<br>2 NA<br><br>Custom alignment: RH |

| | | | |
|---|---|---|---|
| 627 | tpa_tool_thrombo_num<br><br>Show the field ONLY if:<br>(([site]='SMCH' or [site]='SMCW') and (([treatment_type(IAtPA)]='1' or [treatment_type(MER)]='1' or [gs_iatpa]='1' or [gs_iatpaoutside]='1' or [procedures_interventions(2)]='1' or [procedures_interventions(4)]='1' or [procedures_interventions(6)]='1' or [diagnostic_tests(NeuroAngio)]='1') or (([treatment_type(IVtPA)]='1' or [gs_ivthroinit]='1' or [gs_ivtpaoutside]='1') and (rounddown(datediff([jc_arrdatetime],'01-01-2019 00:00','d','mdy',true)) > 0) and (rounddown(datediff([gs_discdatetime],'01-01-2019 00:00','d','mdy',true)) > 0)))) or (([site]='SMCA' or [site]='UMCB') and (([treatment_type(IAtPA)]='1' or [treatment_type(MER)]='1' or [gs_iatpa]='1' or [gs_iatpaoutside]='1' or [procedures_interventions(2)]='1' or [procedures_interventions(4)]='1' or [procedures_interventions(6)]='1' or [diagnostic_tests(NeuroAngio)]='1') or (([treatment_type(IVtPA)]='1' or [gs_ivthroinit]='1' or [gs_ivtpaoutside]='1') and (rounddown(datediff([jc_arrdatetime],'11-07-2017 00:00','d','mdy',true)) > 0) and (rounddown(datediff([gs_discdatetime],'11-07-2017 00:00','d','mdy',true)) > 0)))) and [hide_nonessential_fields(1)]='0' | ✪ Pulse & Site Assessment Numerator<br>*Number Done Correctly.* | text (integer)<br>Custom alignment: RH |

| | | | |
|---|---|---|---|
| 628 | tpa_tool_thrombo_den<br><br>Show the field ONLY if:<br>(([site]='SMCH' or [site]='SMCW') and (([treatment_type(IAtPA)]='1' or [treatment_type(MER)]='1' or [gs_iatpa]='1' or [gs_iatpaoutside]='1' or [procedures_interventions(2)]='1' or [procedures_interventions(4)]='1' or [procedures_interventions(6)]='1' or [diagnostic_tests(NeuroAngio)]='1') or (([treatment_type(IVtPA)]='1' or [gs_ivthroinit]='1' or [gs_ivtpaoutside]='1') and (rounddown(datediff([jc_arrdatetime],'01-01-2019 00:00','d','mdy',true)) > 0) and (rounddown(datediff([gs_discdatetime],'01-01-2019 00:00','d','mdy',true)) > 0)))) or (([site]='SMCA' or [site]='UMCB') and (([treatment_type(IAtPA)]='1' or [treatment_type(MER)]='1' or [gs_iatpa]='1' or [gs_iatpaoutside]='1' or [procedures_interventions(2)]='1' or [procedures_interventions(4)]='1' or [procedures_interventions(6)]='1' or [diagnostic_tests(NeuroAngio)]='1') or (([treatment_type(IVtPA)]='1' or [gs_ivthroinit]='1' or [gs_ivtpaoutside]='1') and (rounddown(datediff([jc_arrdatetime],'11-07-2017 00:00','d','mdy',true)) > 0) and (rounddown(datediff([gs_discdatetime],'11-07-2017 00:00','d','mdy',true)) > 0)))) and [hide_nonessential_fields(1)]='0' | ✪ Pulse & Site Assessment Denominator<br>*Number that should have been done.* | text (integer)<br>Custom alignment: RH |

| | | | |
|---|---|---|---|
| 629 | calc_tpa_tool_compliance<br><br>Show the field ONLY if:<br>(([site]='SMCH' or [site]='SMCW') and ([treatment_type(IAtPA)]='1' or [treatment_type(MER)]='1' or [gs_iatpa]='1' or [gs_iatpaoutside]='1' or [treatment_type(IVtPA)]='1' or [gs_ivthroinit]='1' or [gs_ivtpaoutside]='1') and (rounddown(datediff([jc_arrdatetime],'01-01-2019 00:00','d','mdy',true)) > 0) and (rounddown(datediff([gs_discdatetime],'01-01-2019 00:00','d','mdy',true)) > 0)) or (([site]='SMCA' or [site]='UMCB') and ([treatment_type(IAtPA)]='1' or [treatment_type(MER)] ='1' or [gs_iatpa]='1' or [gs_iatpaoutside]='1' or [treatment_type(IVtPA)]='1' or [gs_ivthroinit]='1' or [gs_ivtpaoutside]='1') and (rounddown(datediff([jc_arrdatetime],'11-07-2017 00:00','d','mdy',true)) > 0) and (rounddown(datediff([gs_discdatetime],'11-07-2017 00:00','d','mdy',true)) > 0)) and [hide_nonessential_fields(1)]='0' | Overall Post-tPA/Thrombectomy Monitoring Compliance<br>*Percent* | calc<br>Calculation: round(((([tpa_tool_nvs_num]/[tpa_tool_nvs_den])+([tpa_tool_vs_num]/[tpa_tool_vs_den])+([tpa_tool_md_num]/[tpa_tool_md_den])+([tpa_tool_handoff_num]/[tpa_tool_handoff_den])+([tpa_tool_thrombo_num]/[tpa_tool_thrombo_den]))/.05,1)<br>Custom alignment: RH |
| 630 | calc_ev_proc_assessment_1<br><br>Show the field ONLY if:<br>([site]='SMCA' or [site]='SMCH' or [site]='SMCW' or [site]='UMCB') and ([treatment_type(IAtPA)] = '1' or [treatment_type(MER)] = '1' or [gs_iatpa] = '1' or [gs_iatpaoutside]='1' or [procedures_interventions(2)]='1' or [procedures_interventions(4)]='1' or [procedures_interventions(6)]='1' or [diagnostic_tests(NeuroAngio)] = '1') and [hide_nonessential_fields(1)]='0' | Pulse & Site Assessment Compliance<br>*Percent* | calc<br>Calculation: if([treatment_type(IAtPA)]='1' or [treatment_type(MER)]='1' or [gs_iatpa]='1' or [procedures_interventions(2)]='1' or [procedures_interventions(4)]='1' or [procedures_interventions(6)]='1' or [diagnostic_tests(NeuroAngio)]='1', round([tpa_tool_thrombo_num]/[tpa_tool_thrombo_den]*100,1), '')<br>Custom alignment: RH |

| | | | |
|---|---|---|---|
| 631 | stk_protocol_activated<br><br>Show the field ONLY if:<br>([site]='SMCA' or [site]='SMCH' or [site]='SMCW' or [site]='UMCB') and ([treatment_type(IVtPA)]='1' or [gs_ivthroinit]='1' or [gs_ivtpaoutside]='1') and ([treatment_type(IAtPA)]='1' or [treatment_type(MER)]='1' or [gs_iatpa]='1' or [gs_iatpaoutside]='1') and [hide_nonessential_fields(1)]='0' | ✪ Patient received IV thrombolysis AND thrombectomy. PRIOR to the start of thrombectomy, was Stroke Protocol Activation documented by IR? | yesno<br>1 Yes<br>0 No<br><br>Custom alignment: RH |
| 632 | ev_proc_2<br><br>Show the field ONLY if:<br>[ev_proc_type_1]<>'' and [hide_nonessential_fields(1)]='0' | ✪ 2nd Procedure | checkbox<br>1 ev_proc_2___1 2nd Procedure<br><br>Custom alignment: RH |
| 633 | ev_proc_type_2<br><br>Show the field ONLY if:<br>[ev_proc_2(1)]='1' and [hide_nonessential_fields(1)]='0' | ✪ Procedure Type | dropdown<br>1 Thrombectomy<br>2 Neuro Angiography<br>3 Coiling<br>4 Embolization<br>0 Other<br><br>Custom alignment: RH |
| 634 | ev_proc_orders_2<br><br>Show the field ONLY if:<br>[ev_proc_2(1)]='1' and [hide_nonessential_fields(1)]='0' | ✪ Were pulse and site assessment orders present? | yesno<br>1 Yes<br>0 No<br><br>Custom alignment: RH |
| 635 | ev_proc_protocol_2<br><br>Show the field ONLY if:<br>[ev_proc_orders_2]='0' and [hide_nonessential_fields(1)]='0' | ✪ If no orders were present, was unit specific protocol followed? | yesno<br>1 Yes<br>0 No<br><br>Custom alignment: RH |
| 636 | ev_proc_end_2<br><br>Show the field ONLY if:<br>[ev_proc_2(1)]='1' and [hide_nonessential_fields(1)]='0' | ✪ Hemostasis Achieved Time | text (datetime_mdy, Min: 2010-01-01 00:00, Max: 2099-12-31 23:59) |

| | | | |
|---|---|---|---|
| 637 | ev_proc_depart_2<br><br>Show the field ONLY if:<br>[ev_proc_2(1)]='1' and [hide_nonessential_fields(1)]='0' | ✪ Time patient left IR Suite | text (datetime_mdy, Min: 2010-01-01 00:00, Max: 2099-12-31 23:59) |
| 638 | ev_proc_ir_2<br><br>Show the field ONLY if:<br>[ev_proc_2(1)]='1' and [hide_nonessential_fields(1)]='0' | ✪ Were all appropriate assessments done in IR Suite. | yesno<br>1 Yes<br>0 No<br><br>Custom alignment: RH |
| 639 | ev_proc_dest_2<br><br>Show the field ONLY if:<br>[ev_proc_2(1)]='1' and [hide_nonessential_fields(1)]='0' | ✪ Patient destination | dropdown<br>1 DSMCUT - MICU<br>2 DSMCUT - SICU<br>16 DSMCUT - Imaging<br>13 DSMCUT - OR<br>3 DSMCUT - PACU<br>4 DSMCUT - 6th Floor<br>20 DSMCUT - Non-Stroke Unit<br>5 SMCA - ICU<br>17 SMCA - Imaging<br>14 SMCA - OR<br>19 SMCA - PACU<br>22 SMCA - 4 North<br>6 SMCA - 4 South/Southwest<br>7 SMCA - 6 North<br>21 SMCA - 7 North<br>8 SMCA - 6 South<br>9 SMCA - 3rd Floor<br>10 SMCW - ICU<br>18 SMCW - Imaging<br>15 SMCW - OR<br>11 SMCW - PACU<br>12 SMCW - 5th Floor |

| | | | |
|---|---|---|---|
| 640 | ev_proc_arrival_2<br><br>Show the field ONLY if:<br>[ev_proc_2(1)]='1' and [hide_nonessential_fields(1)]='0' | ✪ Destination Arrival Time | text (datetime_mdy, Min: 2010-01-01 00:00, Max: 2099-12-31 23:59) |
| 641 | ev_proc_transit_2<br><br>Show the field ONLY if:<br>[ev_proc_2(1)]='1' and [hide_nonessential_fields(1)]='0' | ✪ Were all appropriate assessments documented between IR Suite and patient destination? | radio<br>1 \| Yes<br>0 \| No<br>2 \| NA<br><br>Custom alignment: RH |
| 642 | ev_proc_assessment_num_2<br><br>Show the field ONLY if:<br>[ev_proc_2(1)]='1' and [hide_nonessential_fields(1)]='0' | ✪ Pulse & Site Assessment Numerator<br>*Number Done Correctly.* | text (integer)<br>Custom alignment: RH |
| 643 | ev_proc_assessment_den_2<br><br>Show the field ONLY if:<br>[ev_proc_2(1)]='1' and [hide_nonessential_fields(1)]='0' | ✪ Pulse & Site Assessment Denominator<br>*Number that should have been done.* | text (integer)<br>Custom alignment: RH |
| 644 | calc_ev_proc_assessment_2<br><br>Show the field ONLY if:<br>[ev_proc_2(1)]='1' and [hide_nonessential_fields(1)]='0' | Pulse & Site Assessment Compliance<br>*Percent* | calc<br>Calculation:<br>round([ev_proc_assessment_num_2]/[ev_proc_assessment_den_2<br>Custom alignment: RH |
| 645 | ev_proc_3<br><br>Show the field ONLY if:<br>[ev_proc_type_2]<>'' and [hide_nonessential_fields(1)]='0' | ✪ 3rd Procedure | checkbox<br>1 \| ev_proc_3___1 \| 3rd Procedure<br><br>Custom alignment: RH |
| 646 | ev_proc_type_3<br><br>Show the field ONLY if:<br>[ev_proc_3(1)]='1' and [hide_nonessential_fields(1)]='0' | ✪ Procedure Type | dropdown<br>1 \| Thrombectomy<br>2 \| Neuro Angiography<br>3 \| Coiling<br>4 \| Embolization<br>0 \| Other<br><br>Custom alignment: RH |

| | | | |
|---|---|---|---|
| 647 | ev_proc_orders_3<br><br>Show the field ONLY if:<br>[ev_proc_3(1)]='1' and [hide_nonessential_fields(1)]='0' | ✪ Were pulse and site assessment orders present? | yesno<br>1 Yes<br>0 No<br><br>Custom alignment: RH |
| 648 | ev_proc_protocol_3<br><br>Show the field ONLY if:<br>[ev_proc_orders_3]='0' and [hide_nonessential_fields(1)]='0' | ✪ If no orders were present, was unit specific protocol followed? | yesno<br>1 Yes<br>0 No<br><br>Custom alignment: RH |
| 649 | ev_proc_end_3<br><br>Show the field ONLY if:<br>[ev_proc_3(1)]='1' and [hide_nonessential_fields(1)]='0' | ✪ Hemostasis Achieved Time | text (datetime_mdy, Min: 2010-01-01 00:00, Max: 2099-12-31 23:59) |
| 650 | ev_proc_depart_3<br><br>Show the field ONLY if:<br>[ev_proc_3(1)]='1' and [hide_nonessential_fields(1)]='0' | ✪ Time patient left IR Suite | text (datetime_mdy, Min: 2010-01-01 00:00, Max: 2099-12-31 23:59) |
| 651 | ev_proc_ir_3<br><br>Show the field ONLY if:<br>[ev_proc_3(1)]='1' and [hide_nonessential_fields(1)]='0' | ✪ Were all appropriate assessments done in IR Suite. | yesno<br>1 Yes<br>0 No<br><br>Custom alignment: RH |

| | | | |
|---|---|---|---|
| 652 | ev_proc_dest_3<br><br>Show the field ONLY if:<br>[ev_proc_3(1)]='1' and [hide_nonessential_fields(1)]='0' | ✪ Patient destination | dropdown<br>1 DSMCUT - MICU<br>2 DSMCUT - SICU<br>16 DSMCUT - Imaging<br>13 DSMCUT - OR<br>3 DSMCUT - PACU<br>4 DSMCUT - 6th Floor<br>20 DSMCUT - Non-Stroke Unit<br>5 SMCA - ICU<br>17 SMCA - Imaging<br>14 SMCA - OR<br>19 SMCA - PACU<br>22 SMCA - 4 North<br>6 SMCA - 4 South/Southwest<br>7 SMCA - 6 North<br>21 SMCA - 7 North<br>8 SMCA - 6 South<br>9 SMCA - 3rd Floor<br>10 SMCW - ICU<br>18 SMCW - Imaging<br>15 SMCW - OR<br>11 SMCW - PACU<br>12 SMCW - 5th Floor |
| 653 | ev_proc_arrival_3<br><br>Show the field ONLY if:<br>[ev_proc_3(1)]='1' and [hide_nonessential_fields(1)]='0' | ✪ Destination Arrival Time | text (datetime_mdy, Min: 2010-01-01 00:00, Max: 2099-12-31 23:59) |
| 654 | ev_proc_transit_3<br><br>Show the field ONLY if:<br>[ev_proc_3(1)]='1' and [hide_nonessential_fields(1)]='0' | ✪ Were all appropriate assessments documented between IR Suite and patient destination? | radio<br>1 Yes<br>0 No<br>2 NA<br><br>Custom alignment: RH |

| | | | |
|---|---|---|---|
| 655 | ev_proc_assessment_num_3<br><br>Show the field ONLY if:<br>[ev_proc_3(1)]='1' and [hide_nonessential_fields(1)]='0' | ✪ Pulse & Site Assessment Numerator<br>*Number Done Correctly.* | text (integer)<br>Custom alignment: RH |
| 656 | ev_proc_assessment_den_3<br><br>Show the field ONLY if:<br>[ev_proc_3(1)]='1' and [hide_nonessential_fields(1)]='0' | ✪ Pulse & Site Assessment Denominator<br>*Number that should have been done.* | text (integer)<br>Custom alignment: RH |
| 657 | calc_ev_proc_assessment_3<br><br>Show the field ONLY if:<br>[ev_proc_3(1)]='1' and [hide_nonessential_fields(1)]='0' | Pulse & Site Assessment Compliance<br>*Percent* | calc<br>Calculation:<br>round([ev_proc_assessment_num_3]/[ev_proc_assessment_den_3<br>Custom alignment: RH |
| 658 | ev_proc_4<br><br>Show the field ONLY if:<br>[ev_proc_type_3]<>'' and [hide_nonessential_fields(1)]='0' | ✪ 4th Procedure | checkbox<br>1 — ev_proc_4___1 — 4th Procedure<br><br>Custom alignment: RH |
| 659 | ev_proc_type_4<br><br>Show the field ONLY if:<br>[ev_proc_4(1)]='1' and [hide_nonessential_fields(1)]='0' | ✪ Procedure Type | dropdown<br>1 — Thrombectomy<br>2 — Neuro Angiography<br>3 — Coiling<br>4 — Embolization<br>0 — Other<br><br>Custom alignment: RH |
| 660 | ev_proc_orders_4<br><br>Show the field ONLY if:<br>[ev_proc_4(1)]='1' and [hide_nonessential_fields(1)]='0' | ✪ Were pulse and site assessment orders present? | yesno<br>1 — Yes<br>0 — No<br><br>Custom alignment: RH |
| 661 | ev_proc_protocol_4<br><br>Show the field ONLY if:<br>[ev_proc_orders_4]='0' and [hide_nonessential_fields(1)]='0' | ✪ If no orders were present, was unit specific protocol followed? | yesno<br>1 — Yes<br>0 — No<br><br>Custom alignment: RH |

| | | | |
|---|---|---|---|
| 662 | ev_proc_end_4<br><br>Show the field ONLY if:<br>[ev_proc_4(1)]='1' and [hide_nonessential_fields(1)]='0' | ✪ Hemostasis Achieved Time | text (datetime_mdy, Min: 2010-01-01 00:00, Max: 2099-12-31 23:59) |
| 663 | ev_proc_depart_4<br><br>Show the field ONLY if:<br>[ev_proc_4(1)]='1' and [hide_nonessential_fields(1)]='0' | ✪ Time patient left IR Suite | text (datetime_mdy, Min: 2010-01-01 00:00, Max: 2099-12-31 23:59) |
| 664 | ev_proc_ir_4<br><br>Show the field ONLY if:<br>[ev_proc_4(1)]='1' and [hide_nonessential_fields(1)]='0' | ✪ Were all appropriate assessments done in IR Suite. | yesno<br>1 Yes<br>0 No<br><br>Custom alignment: RH |

| | | | |
|---|---|---|---|
| 665 | ev_proc_dest_4<br><br>Show the field ONLY if:<br>[ev_proc_4(1)]='1' and [hide_nonessential_fields(1)]='0' | ✪ Patient destination | dropdown<br>1 DSMCUT - MICU<br>2 DSMCUT - SICU<br>16 DSMCUT - Imaging<br>13 DSMCUT - OR<br>3 DSMCUT - PACU<br>4 DSMCUT - 6th Floor<br>20 DSMCUT - Non-Stroke Unit<br>5 SMCA - ICU<br>17 SMCA - Imaging<br>14 SMCA - OR<br>19 SMCA - PACU<br>22 SMCA - 4 North<br>6 SMCA - 4 South/Southwest<br>7 SMCA - 6 North<br>21 SMCA - 7 North<br>8 SMCA - 6 South<br>9 SMCA - 3rd Floor<br>10 SMCW - ICU<br>18 SMCW - Imaging<br>15 SMCW - OR<br>11 SMCW - PACU<br>12 SMCW - 5th Floor |
| 666 | ev_proc_arrival_4<br><br>Show the field ONLY if:<br>[ev_proc_4(1)]='1' and [hide_nonessential_fields(1)]='0' | ✪ Destination Arrival Time | text (datetime_mdy, Min: 2010-01-01 00:00, Max: 2099-12-31 23:59) |
| 667 | ev_proc_transit_4<br><br>Show the field ONLY if:<br>[ev_proc_4(1)]='1' and [hide_nonessential_fields(1)]='0' | ✪ Were all appropriate assessments documented between IR Suite and patient destination? | radio<br>1 Yes<br>0 No<br>2 NA<br><br>Custom alignment: RH |

| | | | |
|---|---|---|---|
| 668 | ev_proc_assessment_num_4<br><br>Show the field ONLY if:<br>[ev_proc_4(1)]='1' and [hide_nonessential_fields(1)]='0' | ✪ Pulse & Site Assessment Numerator<br>*Number Done Correctly.* | text (integer)<br>Custom alignment: RH |
| 669 | ev_proc_assessment_den_4<br><br>Show the field ONLY if:<br>[ev_proc_4(1)]='1' and [hide_nonessential_fields(1)]='0' | ✪ Pulse & Site Assessment Denominator<br>*Number that should have been done.* | text (integer)<br>Custom alignment: RH |
| 670 | calc_ev_proc_assessment_4<br><br>Show the field ONLY if:<br>[ev_proc_4(1)]='1' and [hide_nonessential_fields(1)]='0' | Pulse & Site Assessment Compliance<br>*Percent* | calc<br>Calculation:<br>round([ev_proc_assessment_num_4]/[ev_proc_assessment_den_4[illegible]<br>Custom alignment: RH |
| 671 | ev_proc_assessment_notes<br><br>Show the field ONLY if:<br>([site]='SMCA' or [site]='SMCH' or [site]='SMCW' or [site]='UMCB') and ([treatment_type(IAtPA)]='1' or [treatment_type(MER)]='1' or [gs_iatpa]='1' or [procedures_interventions(2)]='1' or [procedures_interventions(4)]='1' or [procedures_interventions(6)]='1' or [diagnostic_tests(NeuroAngio)]='1') and [hide_nonessential_fields(1)]='0' | ✪ Identify specific areas of Pulse & Site Assessment non-compliance | notes |
| 672 | dnv_15_21days<br><br>Show the field ONLY if:<br>([site]='SMCA' or [site]='SMCH' or [site]='SMCW' or [site]='UMCB') and [aneurysmal_sah]='1' and [gs_notadmit]<>'1' and [hide_nonessential_fields(1)]='0' | Section Header: *DNV 15: Nimodipine for 21 days or until discharge*<br><br>Was Nimodipine continued for 21 days or until discharge if discharged in less than 21 days? | yesno<br>1 Yes<br>0 No<br><br>Custom alignment: RH |
| 673 | additional_stroke_information_complete | Section Header: *Form Status*<br><br>Complete? | dropdown<br>0 Incomplete<br>1 Unverified<br>2 Complete |

Instrument: **Optional** (optional)

Collapse

| | | | |
|---|---|---|---|
| 674 | gs_comments<br><br>Show the field ONLY if:<br>([site]='SMCA' or [site]='SMCH' or [site]='SMCW' or [site]='UMCB') and ([treatment_recvd]='1' or [gs_notadmit]<>'1') and [hide_nonessential_fields(1)]='0' | Section Header: *Optional Fields*<br>Additional Comments | text<br>Field Annotation: @CHARLIMIT=250 |
| 675 | gs_pmtused<br><br>Show the field ONLY if:<br>([site]='SMCA' or [site]='SMCH' or [site]='SMCW' or [site]='UMCB') and ([treatment_recvd]='1' or [gs_notadmit]<>'1') and [hide_nonessential_fields(1)]='0' | Section Header: *Administrative*<br>✪ PMT used concurrently or retrospectively or combination? | radio<br>1 Concurrently<br>2 Retrospectively<br>3 Combination<br><br>Custom alignment: RH |
| 676 | gs_pc_admorder<br><br>Show the field ONLY if:<br>([site]='SMCA' or [site]='SMCH' or [site]='SMCW' or [site]='UMCB') and ([treatment_recvd]='1' or [gs_notadmit]<>'1') and [hide_nonessential_fields(1)]='0' | ✪ Was a stroke admission order set used in this patient? | radio<br>1 Yes<br>2 No<br><br>Custom alignment: RH |
| 677 | gs_pc_dischchlst<br><br>Show the field ONLY if:<br>([site]='SMCA' or [site]='SMCH' or [site]='SMCW' or [site]='UMCB') and ([treatment_recvd]='1' or [gs_notadmit]<>'1') and [hide_nonessential_fields(1)]='0' | Was a stroke discharge checklist used in this patient? | radio<br>1 Yes<br>2 No<br><br>Custom alignment: RH |
| 678 | gs_ptcontract<br><br>Show the field ONLY if:<br>([site]='SMCA' or [site]='SMCH' or [site]='SMCW' or [site]='UMCB') and ([treatment_recvd]='1' or [gs_notadmit]<>'1') and [hide_nonessential_fields(1)]='0' | Patient adherence contract/compact used? | radio<br>1 Yes<br>2 No<br><br>Custom alignment: RH |
| 679 | optional_complete | Section Header: *Form Status*<br>Complete? | dropdown<br>0 Incomplete<br>1 Unverified<br>2 Complete |

Instrument: **Discharge** (discharge)

Collapse

| | | | |
|---|---|---|---|
| 680 | gs_rankindc<br><br>Show the field ONLY if:<br>([site]='SMCA' or [site]='SMCH' or [site]='SMCW' or [site]='UMCB') and ([gs_notadmit]<>'1' or [jc_disposition]='6') and [hide_nonessential_fields(1)]='0' | Section Header: *Discharge Information*<br><br>Modified Rankin Scale at Discharge | radio<br>1 Yes<br>2 No/ND<br><br>Custom alignment: RH |
| 681 | disch_rankin_na<br><br>Show the field ONLY if:<br>[gs_rankindc]='2' | If (and only if) Modified Rankin Scale at Discharge is not applicable, please specify reason(s). | checkbox<br>1 disch_rankin_na___1 Patient left AMA<br>0 disch_rankin_na___0 Other {disch_rankin_na_oth:icons}<br><br>Field Annotation: PLEASE NOTE: If any options are added to this field, the DNV QM.7 CR.4b CALCULATION MUST BE UPDATED! (calc_dnv_qm7_cr4b) |
| 682 | disch_rankin_na_oth<br><br>Show the field ONLY if:<br>[disch_rankin_na(0)]='1' | Other reason Discharge mRS not applicable | text |
| 683 | gs_rankindcyes<br><br>Show the field ONLY if:<br>[gs_rankindc]='1' and [hide_nonessential_fields(1)]='0' | If Modified Rankin Scale at Discharge: Yes | radio<br>1 Actual<br>2 Estimated from the record<br>3 ND<br><br>Custom alignment: RH |
| 684 | gs_rankindctotal<br><br>Show the field ONLY if:<br>[gs_rankindc]='1' and [hide_nonessential_fields(1)]='0' | Modified Rankin Scale at Discharge Total | calc<br>Calculation: [gs_rankin]-1 |

| | | | |
|---|---|---|---|
| 685 | gs_rankin<br><br>Show the field ONLY if:<br>[gs_rankindc]='1' and [hide_nonessential_fields(1)]='0' | Modified Rankin Scale at Discharge Stroke | radio<br>1 | 0 - No symptoms at all<br>2 | 1 - No significant disability despite symptoms: Able to carry out all usual activities<br>3 | 2 - Slight disability<br>4 | 3 - Moderate disability: Requiring some help but able to walk without assistance<br>5 | 4 - Moderate to severe disability: Unable to walk without assistance and unable to attend to own bodily needs without assistance<br>6 | 5 - Severe disability: Bedridden, incontinent and requiring constant nursing care and attention<br>7 | 6 - Death |
| 686 | gs_funcstatus<br><br>Show the field ONLY if:<br>([site]='SMCA' or [site]='SMCH' or [site]='SMCW' or [site]='UMCB') and [jc_disposition] <> '6' and [gs_notadmit]<>'1' and [hide_nonessential_fields(1)]='0' | Ambulatory status at discharge? | radio<br>2 | Able to ambulate independently (no help from another person) w/ or w/o device<br>3 | With assistance (from person)<br>4 | Unable to ambulate<br>5 | ND |
| 687 | gs_disch_bpnd<br><br>Show the field ONLY if:<br>([site]='SMCA' or [site]='SMCH' or [site]='SMCW' or [site]='UMCB') and [jc_disposition] <> '6' and [gs_notadmit]<>'1' and [hide_nonessential_fields(1)]='0' | Discharge BP: ND | radio<br>1 | ND |
| 688 | gs_disch_systolic<br><br>Show the field ONLY if:<br>([site]='SMCA' or [site]='SMCH' or [site]='SMCW' or [site]='UMCB') and [gs_disch_bpnd] <> '1' and [jc_disposition] <> '6' and [gs_notadmit]<>'1' and [hide_nonessential_fields(1)]='0' | Discharge Blood Pressure (Measurement closest to discharge) Systolic<br>*mmHg (Systolic/Diastolic)* | text (integer, Min: 40, Max: 275) |

| | | | |
|---|---|---|---|
| 689 | gs_disch_diastolic<br><br>Show the field ONLY if:<br>([site]='SMCA' or [site]='SMCH' or [site]='SMCW' or [site]='UMCB') and [gs_disch_bpnd] <> '1' and [jc_disposition] <> '6' and [gs_notadmit] <>'1' and [hide_nonessential_fields(1)]='0' | Discharge Blood Pressure (Measurement closest to discharge) Diastolic<br>*mmHg (Systolic/Diastolic)* | text (integer, Min: 20, Max: 200) |
| 690 | ich_hypmed<br><br>Show the field ONLY if:<br>[site]='UMCB' | ✪ Did the patient receive any antihypertensive medication on the day of or the day prior to discharge? | radio<br>1 Yes<br>2 No/ND<br><br>Custom alignment: RH |
| 691 | gs_antithromboticyesno<br><br>Show the field ONLY if:<br>[gs_notadmit] <> '1' and [hide_nonessential_fields(1)]='0' | Section Header: *Discharge Treatments*<br><br>✪ Antithrombotic therapy approved in stroke - Prescribed? | radio<br>1 Yes<br>2 No/ND<br>3 NC<br><br>Custom alignment: RH |
| 692 | gs_antithrombotic1class<br><br>Show the field ONLY if:<br>([site]='SMCA' or [site]='SMCH' or [site]='SMCW' or [site]='UMCB') and [gs_antithromboticyesno] = '1' and [hide_nonessential_fields(1)] ='0' | Prescribed Antithrombotic 1 - Antithrombotic Class | dropdown<br>1 Antiplatelet<br>2 Anticoagulant |
| 693 | gs_antithrombotic1a<br><br>Show the field ONLY if:<br>[gs_antithrombotic1class]='1' and [hide_nonessential_fields(1)]='0' | Prescribed Antithrombotic 1 - Antiplatelet | dropdown<br>3 aspirin<br>4 ASA/dipyridamole (Aggrenox)<br>6 clopidogrel (Plavix)<br>7 ticlopidine (Ticlid) |

| | | | |
|---|---|---|---|
| 694 | gs_antithrombotic1b<br><br>Show the field ONLY if:<br>[gs_antithrombotic1class]='2' and [hide_nonessential_fields(1)]='0' | Prescribed Antithrombotic 1 - Anticoagulant | dropdown<br>19 apixaban (Eliquis)<br>15 argatroban<br>16 dabigatran (Pradaxa)<br>20 edoxaban (Savaysa)<br>12 fondaparinux (Arixtra)<br>9 full dose LMW heparin<br>18 lepirudin (Refludan)<br>14 rivaroxaban (Xarelto)<br>8 Unfractionated heparin IV<br>5 warfarin (Coumadin) |
| 695 | gs_antithrombotic1dose1<br><br>Show the field ONLY if:<br>[gs_antithrombotic1a]='3' and [hide_nonessential_fields(1)]='0' | Prescribed Antithrombotic 1 - Dosage | dropdown<br>100 75 mg<br>101 81 mg<br>102 125 mg<br>103 162 mg<br>104 300 mg<br>105 325 mg<br>106 Other<br>107 Unknown |
| 696 | gs_antithrombotic1dose2<br><br>Show the field ONLY if:<br>[gs_antithrombotic1a]='4' and [hide_nonessential_fields(1)]='0' | Prescribed Antithrombotic 1 - Dosage | dropdown<br>200 200 mg - 25 mg<br>201 Other<br>202 Unknown |
| 697 | gs_antithrombotic1dose3<br><br>Show the field ONLY if:<br>[gs_antithrombotic1a]='6' and [hide_nonessential_fields(1)]='0' | Prescribed Antithrombotic 1 - Dosage | dropdown<br>400 75 mg<br>401 Other<br>402 Unknown |
| 698 | gs_antithrombotic1dose4<br><br>Show the field ONLY if:<br>[gs_antithrombotic1a]='7' and [hide_nonessential_fields(1)]='0' | Prescribed Antithrombotic 1 - Dosage | dropdown<br>500 250 mg<br>501 Other<br>502 Unknown |

| | | | |
|---|---|---|---|
| 699 | gs_antithrombotic1dose5<br><br>Show the field ONLY if:<br>[gs_antithrombotic1b]='19' and [hide_nonessential_fields(1)]='0' | Prescribed Antithrombotic 1 - Dosage | dropdown<br>700 \| 2.5 mg<br>701 \| 5 mg |
| 700 | gs_antithrombotic1dose6<br><br>Show the field ONLY if:<br>[gs_antithrombotic1b] = '16' and [hide_nonessential_fields(1)]='0' | Prescribed Antithrombotic 1 - Dosage | dropdown<br>900 \| 75 mg<br>901 \| 150 mg<br>902 \| Other<br>903 \| Unknown |
| 701 | gs_antithrombotic1dose7<br><br>Show the field ONLY if:<br>[gs_antithrombotic1b] = '20' and [hide_nonessential_fields(1)]='0' | Prescribed Antithrombotic 1 - Dosage | dropdown<br>904 \| 60 mg<br>905 \| Other<br>906 \| Unknown |
| 702 | gs_antithrombotic1dose8<br><br>Show the field ONLY if:<br>[gs_antithrombotic1b] = '12' and [hide_nonessential_fields(1)]='0' | Prescribed Antithrombotic 1 - Dosage | dropdown<br>700 \| 2.5 mg<br>701 \| 5 mg<br>702 \| 7.5 mg<br>703 \| 10 mg<br>704 \| Other<br>705 \| Unknown |
| 703 | gs_antithrombotic1dose9<br><br>Show the field ONLY if:<br>[gs_antithrombotic1b] = '14' and [hide_nonessential_fields(1)]='0' | Prescribed Antithrombotic 1 - Dosage | dropdown<br>800 \| 10 mg<br>803 \| 20 mg<br>801 \| Other<br>802 \| Unknown |

| | | | |
|---|---|---|---|
| 704 | gs_antithrombotic1dose10<br><br>Show the field ONLY if:<br>[gs_antithrombotic1b] = '5' and [hide_nonessential_fields(1)]='0' | Prescribed Antithrombotic 1 - Dosage | dropdown<br>300 \| 2 mg<br>301 \| 2.5 mg<br>302 \| 3 mg<br>303 \| 4 mg<br>304 \| 5 mg<br>305 \| 6 mg<br>306 \| 7.5 mg<br>307 \| Other<br>308 \| Unknown |
| 705 | gs_antithrombotic1freq<br><br>Show the field ONLY if:<br>[gs_antithrombotic1dose1] <> '' or [gs_antithrombotic1dose2] <> '' or [gs_antithrombotic1dose3] <> '' or [gs_antithrombotic1dose4] <> '' or [gs_antithrombotic1dose5] <> '' or [gs_antithrombotic1dose6] <> '' or [gs_antithrombotic1dose7] <> '' or [gs_antithrombotic1dose8] <> '' or [gs_antithrombotic1dose9] <> '' or [gs_antithrombotic1dose10] <> '' and [hide_nonessential_fields(1)] ='0' | Prescribed Antithrombotic 1 - Frequency | dropdown<br>2 \| Every day<br>3 \| 2 times a day<br>4 \| 3 times a day<br>5 \| 4 times a day<br>6 \| Other<br>7 \| Unknown |
| 706 | gs_antithrombotic2class<br><br>Show the field ONLY if:<br>[gs_antithrombotic1class] <> '' and [hide_nonessential_fields(1)]='0' | Prescribed Antithrombotic 2 - Antithrombotic Class | dropdown<br>1 \| Antiplatelet<br>2 \| Anticoagulant |
| 707 | gs_antithrombotic2a<br><br>Show the field ONLY if:<br>[gs_antithrombotic2class] = '1' and [hide_nonessential_fields(1)]='0' | Prescribed Antithrombotic 2 - Antiplatelet | dropdown<br>3 \| aspirin<br>4 \| ASA/dipyridamole (Aggrenox)<br>6 \| clopidogrel (Plavix)<br>7 \| ticlopidine (Ticlid) |

| | | | |
|---|---|---|---|
| 708 | gs_antithrombotic2b<br><br>Show the field ONLY if:<br>[gs_antithrombotic2class] = '2' and [hide_nonessential_fields(1)]='0' | Prescribed Antithrombotic 2 - Anticoagulant | dropdown<br>19 apixaban (Eliquis)<br>15 argatroban<br>16 dabigatran (Pradaxa)<br>20 edoxaban (Savaysa)<br>12 fondaparinux (Arixtra)<br>9 full dose LMW heparin<br>18 lepirudin (Refludan)<br>14 rivaroxaban (Xarelto)<br>8 Unfractionated heparin IV<br>5 warfarin (Coumadin) |
| 709 | gs_antithrombotic2dose1<br><br>Show the field ONLY if:<br>[gs_antithrombotic2a] = '3' and [hide_nonessential_fields(1)]='0' | Prescribed Antithrombotic 2 - Dosage | dropdown<br>100 75 mg<br>101 81 mg<br>102 125 mg<br>103 162 mg<br>104 300 mg<br>105 325 mg<br>106 Other<br>107 Unknown |
| 710 | gs_antithrombotic2dose2<br><br>Show the field ONLY if:<br>[gs_antithrombotic2a] = '4' and [hide_nonessential_fields(1)]='0' | Prescribed Antithrombotic 2 - Dosage | dropdown<br>200 200 mg - 25 mg<br>201 Other<br>202 Unknown |
| 711 | gs_antithrombotic2dose3<br><br>Show the field ONLY if:<br>[gs_antithrombotic2a] = '6' and [hide_nonessential_fields(1)]='0' | Prescribed Antithrombotic 2 - Dosage | dropdown<br>400 75 mg<br>401 Other<br>402 Unknown |
| 712 | gs_antithrombotic2dose4<br><br>Show the field ONLY if:<br>[gs_antithrombotic2a] = '7' and [hide_nonessential_fields(1)]='0' | Prescribed Antithrombotic 2 - Dosage | dropdown<br>500 250 mg<br>501 Other<br>502 Unknown |

| | | | |
|---|---|---|---|
| 713 | gs_antithrombotic2dose5<br><br>Show the field ONLY if:<br>[gs_antithrombotic2b] = '19' and<br>[hide_nonessential_fields(1)]='0' | Prescribed Antithrombotic 2 - Dosage | dropdown<br>700 \| 2.5 mg<br>701 \| 5 mg |
| 714 | gs_antithrombotic2dose6<br><br>Show the field ONLY if:<br>[gs_antithrombotic2b] = '16' and<br>[hide_nonessential_fields(1)]='0' | Prescribed Antithrombotic 2 - Dosage | dropdown<br>900 \| 75 mg<br>901 \| 150 mg<br>902 \| Other<br>903 \| Unknown |
| 715 | gs_antithrombotic2dose7<br><br>Show the field ONLY if:<br>[gs_antithrombotic2b] = '20' and<br>[hide_nonessential_fields(1)]='0' | Prescribed Antithrombotic 2 - Dosage | dropdown<br>904 \| 60 mg<br>905 \| Other<br>906 \| Unknown |
| 716 | gs_antithrombotic2dose8<br><br>Show the field ONLY if:<br>[gs_antithrombotic2b] = '12' and<br>[hide_nonessential_fields(1)]='0' | Prescribed Antithrombotic 2 - Dosage | dropdown<br>700 \| 2.5 mg<br>701 \| 5 mg<br>702 \| 7.5 mg<br>703 \| 10 mg<br>704 \| Other<br>705 \| Unknown |
| 717 | gs_antithrombotic2dose9<br><br>Show the field ONLY if:<br>[gs_antithrombotic2b] = '14' and<br>[hide_nonessential_fields(1)]='0' | Prescribed Antithrombotic 2 - Dosage | dropdown<br>800 \| 10 mg<br>803 \| 20 mg<br>801 \| Other<br>802 \| Unknown |

| | | | |
|---|---|---|---|
| 718 | gs_antithrombotic2dose10<br><br>Show the field ONLY if:<br>[gs_antithrombotic2b] = '5' and [hide_nonessential_fields(1)]='0' | Prescribed Antithrombotic 2 - Dosage | dropdown<br>300 \| 2 mg<br>301 \| 2.5 mg<br>302 \| 3 mg<br>303 \| 4 mg<br>304 \| 5 mg<br>305 \| 6 mg<br>306 \| 7.5 mg<br>307 \| Other<br>308 \| Unknown |
| 719 | gs_antithrombotic2freq<br><br>Show the field ONLY if:<br>[gs_antithrombotic2dose1] <> '' or [gs_antithrombotic2dose2] <> '' or [gs_antithrombotic2dose3] <> '' or [gs_antithrombotic2dose4] <> '' or [gs_antithrombotic2dose5] <> '' or [gs_antithrombotic2dose6] <> '' or [gs_antithrombotic2dose7] <> '' or [gs_antithrombotic2dose8] <> '' or [gs_antithrombotic2dose9] <> '' or [gs_antithrombotic2dose10] <> '' and [hide_nonessential_fields(1)]='0' | Prescribed Antithrombotic 2 - Frequency | dropdown<br>2 \| Every day<br>3 \| 2 times a day<br>4 \| 3 times a day<br>5 \| 4 times a day<br>6 \| Other<br>7 \| Unknown |
| 720 | gs_antithrombotic3class<br><br>Show the field ONLY if:<br>[gs_antithrombotic2class] <> '' and [hide_nonessential_fields(1)]='0' | Prescribed Antithrombotic 3 - Antithrombotic Class | dropdown<br>1 \| Antiplatelet<br>2 \| Anticoagulant |
| 721 | gs_antithrombotic3a<br><br>Show the field ONLY if:<br>[gs_antithrombotic3class] = '1' and [hide_nonessential_fields(1)]='0' | Prescribed Antithrombotic 3 - Antiplatelet | dropdown<br>3 \| aspirin<br>4 \| ASA/dipyridamole (Aggrenox)<br>6 \| clopidogrel (Plavix)<br>7 \| ticlopidine (Ticlid) |

| | | | |
|---|---|---|---|
| 722 | gs_antithrombotic3b<br><br>Show the field ONLY if:<br>[gs_antithrombotic3class] = '2' and [hide_nonessential_fields(1)]='0' | Prescribed Antithrombotic 3 - Anticoagulant | dropdown<br>19 apixaban (Eliquis)<br>15 argatroban<br>16 dabigatran (Pradaxa)<br>20 edoxaban (Savaysa)<br>12 fondaparinux (Arixtra)<br>9 full dose LMW heparin<br>18 lepirudin (Refludan)<br>14 rivaroxaban (Xarelto)<br>8 Unfractionated heparin IV<br>5 warfarin (Coumadin) |
| 723 | gs_antithrombotic3dose1<br><br>Show the field ONLY if:<br>[gs_antithrombotic3a] = '3' and [hide_nonessential_fields(1)]='0' | Prescribed Antithrombotic 3 - Dosage | dropdown<br>100 75 mg<br>101 81 mg<br>102 125 mg<br>103 162 mg<br>104 300 mg<br>105 325 mg<br>106 Other<br>107 Unknown |
| 724 | gs_antithrombotic3dose2<br><br>Show the field ONLY if:<br>[gs_antithrombotic3a] = '4' and [hide_nonessential_fields(1)]='0' | Prescribed Antithrombotic 3 - Dosage | dropdown<br>200 200 mg - 25 mg<br>201 Other<br>202 Unknown |
| 725 | gs_antithrombotic3dose3<br><br>Show the field ONLY if:<br>[gs_antithrombotic3a] = '6' and [hide_nonessential_fields(1)]='0' | Prescribed Antithrombotic 3 - Dosage | dropdown<br>400 75 mg<br>401 Other<br>402 Unknown |
| 726 | gs_antithrombotic3dose4<br><br>Show the field ONLY if:<br>[gs_antithrombotic3a] = '7' and [hide_nonessential_fields(1)]='0' | Prescribed Antithrombotic 3 - Dosage | dropdown<br>500 250 mg<br>501 Other<br>502 Unknown |

| | | | |
|---|---|---|---|
| 727 | gs_antithrombotic3dose5<br><br>Show the field ONLY if:<br>[gs_antithrombotic3b] = '19' and<br>[hide_nonessential_fields(1)]='0' | Prescribed Antithrombotic 3 - Dosage | dropdown<br>700 2.5 mg<br>701 5 mg |
| 728 | gs_antithrombotic3dose6<br><br>Show the field ONLY if:<br>[gs_antithrombotic3b] = '16' and<br>[hide_nonessential_fields(1)]='0' | Prescribed Antithrombotic 3 - Dosage | dropdown<br>900 75 mg<br>901 150 mg<br>902 Other<br>903 Unknown |
| 729 | gs_antithrombotic3dose7<br><br>Show the field ONLY if:<br>[gs_antithrombotic3b] = '20' and<br>[hide_nonessential_fields(1)]='0' | Prescribed Antithrombotic 3 - Dosage | dropdown<br>904 60 mg<br>905 Other<br>906 Unknown |
| 730 | gs_antithrombotic3dose8<br><br>Show the field ONLY if:<br>[gs_antithrombotic3b] = '12' and<br>[hide_nonessential_fields(1)]='0' | Prescribed Antithrombotic 3 - Dosage | dropdown<br>700 2.5 mg<br>701 5 mg<br>702 7.5 mg<br>703 10 mg<br>704 Other<br>705 Unknown |
| 731 | gs_antithrombotic3dose9<br><br>Show the field ONLY if:<br>[gs_antithrombotic3b] = '14' and<br>[hide_nonessential_fields(1)]='0' | Prescribed Antithrombotic 3 - Dosage | dropdown<br>800 10 mg<br>803 20 mg<br>801 Other<br>802 Unknown |

| | | | |
|---|---|---|---|
| 732 | gs_antithrombotic3dose10<br><br>Show the field ONLY if:<br>[gs_antithrombotic3b] = '5' and [hide_nonessential_fields(1)]='0' | Prescribed Antithrombotic 3 - Dosage | dropdown<br>300 \| 2 mg<br>301 \| 2.5 mg<br>302 \| 3 mg<br>303 \| 4 mg<br>304 \| 5 mg<br>305 \| 6 mg<br>306 \| 7.5 mg<br>307 \| Other<br>308 \| Unknown |
| 733 | gs_antithrombotic3freq<br><br>Show the field ONLY if:<br>[gs_antithrombotic3dose1] <> '' or [gs_antithrombotic3dose2] <> '' or [gs_antithrombotic3dose3] <> '' or [gs_antithrombotic3dose4] <> '' or [gs_antithrombotic3dose5] <> '' or [gs_antithrombotic3dose6] <> '' or [gs_antithrombotic3dose7] <> '' or [gs_antithrombotic3dose8] <> '' or [gs_antithrombotic3dose9] <> '' or [gs_antithrombotic3dose10] <> '' and [hide_nonessential_fields(1)] ='0' | Prescribed Antithrombotic 3 - Frequency | dropdown<br>2 \| Every day<br>3 \| 2 times a day<br>4 \| 3 times a day<br>5 \| 4 times a day<br>6 \| Other<br>7 \| Unknown |
| 734 | gs_antithrombotic4class<br><br>Show the field ONLY if:<br>[gs_antithrombotic3class] <> '' and [hide_nonessential_fields(1)]='0' | Prescribed Antithrombotic 4 - Antithrombotic Class | dropdown<br>1 \| Antiplatelet<br>2 \| Anticoagulant |
| 735 | gs_antithrombotic4a<br><br>Show the field ONLY if:<br>[gs_antithrombotic4class] = '1' and [hide_nonessential_fields(1)]='0' | Prescribed Antithrombotic 4 - Antiplatelet | dropdown<br>3 \| aspirin<br>4 \| ASA/dipyridamole (Aggrenox)<br>6 \| clopidogrel (Plavix)<br>7 \| ticlopidine (Ticlid) |

| | | | |
|---|---|---|---|
| 736 | gs_antithrombotic4b<br><br>Show the field ONLY if:<br>[gs_antithrombotic4class] = '2' and [hide_nonessential_fields(1)]='0' | Prescribed Antithrombotic 4 - Anticoagulant | dropdown<br>19 apixaban (Eliquis)<br>15 argatroban<br>16 dabigatran (Pradaxa)<br>20 edoxaban (Savaysa)<br>12 fondaparinux (Arixtra)<br>9 full dose LMW heparin<br>18 lepirudin (Refludan)<br>14 rivaroxaban (Xarelto)<br>8 Unfractionated heparin IV<br>5 warfarin (Coumadin) |
| 737 | gs_antithrombotic4dose1<br><br>Show the field ONLY if:<br>[gs_antithrombotic4a] = '3' and [hide_nonessential_fields(1)]='0' | Prescribed Antithrombotic 4 - Dosage | dropdown<br>100 75 mg<br>101 81 mg<br>102 125 mg<br>103 162 mg<br>104 300 mg<br>105 325 mg<br>106 Other<br>107 Unknown |
| 738 | gs_antithrombotic4dose2<br><br>Show the field ONLY if:<br>[gs_antithrombotic4a] = '4' and [hide_nonessential_fields(1)]='0' | Prescribed Antithrombotic 4 - Dosage | dropdown<br>200 200 mg - 25 mg<br>201 Other<br>202 Unknown |
| 739 | gs_antithrombotic4dose3<br><br>Show the field ONLY if:<br>[gs_antithrombotic4a] = '6' and [hide_nonessential_fields(1)]='0' | Prescribed Antithrombotic 4 - Dosage | dropdown<br>400 75 mg<br>401 Other<br>402 Unknown |
| 740 | gs_antithrombotic4dose4<br><br>Show the field ONLY if:<br>[gs_antithrombotic4a] = '7' and [hide_nonessential_fields(1)]='0' | Prescribed Antithrombotic 4 - Dosage | dropdown<br>500 250 mg<br>501 Other<br>502 Unknown |

| | | | |
|---|---|---|---|
| 741 | gs_antithrombotic4dose5<br><br>Show the field ONLY if:<br>[gs_antithrombotic4b] = '19' and<br>[hide_nonessential_fields(1)]='0' | Prescribed Antithrombotic 4 - Dosage | dropdown<br>700 — 2.5 mg<br>701 — 5 mg |
| 742 | gs_antithrombotic4dose6<br><br>Show the field ONLY if:<br>[gs_antithrombotic4b] = '16' and<br>[hide_nonessential_fields(1)]='0' | Prescribed Antithrombotic 4 - Dosage | dropdown<br>900 — 75 mg<br>901 — 150 mg<br>902 — Other<br>903 — Unknown |
| 743 | gs_antithrombotic4dose7<br><br>Show the field ONLY if:<br>[gs_antithrombotic4b] = '20' and<br>[hide_nonessential_fields(1)]='0' | Prescribed Antithrombotic 4 - Dosage | dropdown<br>904 — 60 mg<br>905 — Other<br>906 — Unknown |
| 744 | gs_antithrombotic4dose8<br><br>Show the field ONLY if:<br>[gs_antithrombotic4b] = '12' and<br>[hide_nonessential_fields(1)]='0' | Prescribed Antithrombotic 4 - Dosage | dropdown<br>700 — 2.5 mg<br>701 — 5 mg<br>702 — 7.5 mg<br>703 — 10 mg<br>704 — Other<br>705 — Unknown |
| 745 | gs_antithrombotic4dose9<br><br>Show the field ONLY if:<br>[gs_antithrombotic4b] = '14' and<br>[hide_nonessential_fields(1)]='0' | Prescribed Antithrombotic 4 - Dosage | dropdown<br>800 — 10 mg<br>803 — 20 mg<br>801 — Other<br>802 — Unknown |

| | | | |
|---|---|---|---|
| 746 | gs_antithrombotic4dose10<br><br>Show the field ONLY if:<br>[gs_antithrombotic4b] = '5' and [hide_nonessential_fields(1)]='0' | Prescribed Antithrombotic 4 - Dosage | dropdown<br>300 \| 2 mg<br>301 \| 2.5 mg<br>302 \| 3 mg<br>303 \| 4 mg<br>304 \| 5 mg<br>305 \| 6 mg<br>306 \| 7.5 mg<br>307 \| Other<br>308 \| Unknown |
| 747 | gs_antithrombotic4freq<br><br>Show the field ONLY if:<br>[gs_antithrombotic4dose1] <> '' or [gs_antithrombotic4dose2] <> '' or [gs_antithrombotic4dose3] <> '' or [gs_antithrombotic4dose4] <> '' or [gs_antithrombotic4dose5] <> '' or [gs_antithrombotic4dose6] <> '' or [gs_antithrombotic4dose7] <> '' or [gs_antithrombotic4dose8] <> '' or [gs_antithrombotic4dose9] <> '' or [gs_antithrombotic4dose10] <> '' and [hide_nonessential_fields(1)]='0' | Prescribed Antithrombotic 4 - Frequency | dropdown<br>2 \| Every day<br>3 \| 2 times a day<br>4 \| 3 times a day<br>5 \| 4 times a day<br>6 \| Other<br>7 \| Unknown |
| 748 | gs_reasonnoantithrom<br><br>Show the field ONLY if:<br>[gs_antithromboticyesno] = '3' and [jc_disposition] <> '6' and [hide_nonessential_fields(1)]='0' | If NC, documented contraindications<br>*Select all that apply.* | checkbox<br>13 \| gs_reasonnoantithrom___13 \| Allergy to or complication r/t aspirin, ticlopidine, clopidogrel, dipyridamole and warfarin or heparins (hx or current)<br>4 \| gs_reasonnoantithrom___4 \| Patient/Family refused<br>18 \| gs_reasonnoantithrom___18 \| Risk for bleeding or discontinued due to bleeding<br>21 \| gs_reasonnoantithrom___21 \| Serious side effect to medication<br>8 \| gs_reasonnoantithrom___8 \| Terminal illness/Comfort Measures Only<br>22 \| gs_reasonnoantithrom___22 \| Other |

| | | | |
|---|---|---|---|
| 749 | gs_otheranti<br><br>Show the field ONLY if:<br>([site]='SMCA' or [site]='SMCH' or [site]='SMCW' or [site]='UMCB') and [gs_notadmit]<>'1' and [hide_nonessential_fields(1)]='0' | Other Antithrombotic(s) - Prescribed? | radio<br>1 Yes<br>2 No<br><br>Custom alignment: RH |
| 750 | gs_otheranti_med1<br><br>Show the field ONLY if:<br>[gs_otheranti] = '1' and [hide_nonessential_fields(1)]='0' | Other Antithrombotic(s) Med 1 | dropdown<br>1 desirudin (Iprivask)<br>2 ticagrelor (Brilinta)<br>3 prasugrel (Effient) *contraindication in stroke and TIA<br>4 Other |
| 751 | gs_otheranti_dos1a<br><br>Show the field ONLY if:<br>[gs_otheranti_med1] = '1' and [hide_nonessential_fields(1)]='0' | Other Antithrombotic(s) Med 1 - Dose | dropdown<br>3 15 mg<br>5 Other<br>6 Unknown |
| 752 | gs_otheranti_dos1b<br><br>Show the field ONLY if:<br>[gs_otheranti_med1] = '2' and [hide_nonessential_fields(1)]='0' | Other Antithrombotic(s) Med 1 - Dose | dropdown<br>4 90 mg |
| 753 | gs_otheranti_dos1c<br><br>Show the field ONLY if:<br>[gs_otheranti_med1] = '3' and [hide_nonessential_fields(1)]='0' | Other Antithrombotic(s) Med 1 - Dose | dropdown<br>1 5 mg<br>2 10 mg |
| 754 | gs_otheranti_freq1<br><br>Show the field ONLY if:<br>[gs_otheranti_dos1a] <> '' or [gs_otheranti_dos1b] <> '' or [gs_otheranti_dos1c] <> '' and [hide_nonessential_fields(1)]='0' | Other Antithrombotic(s) Med 1 - Frequency | dropdown<br>1 Every Day<br>2 2 times a day<br>3 3 times a day<br>4 4 times a day<br>5 Other<br>6 Unknown |

| | | | |
|---|---|---|---|
| 755 | gs_otheranti_med2<br><br>Show the field ONLY if:<br>[gs_otheranti_med1] <> '' and [hide_nonessential_fields(1)]='0' | Other Antithrombotic(s) Med 2 | dropdown<br>1 desirudin (Iprivask)<br>2 ticagrelor (Brilinta)<br>3 prasugrel (Effient) *contraindication in stroke and TIA<br>4 Other |
| 756 | gs_otheranti_dos2a<br><br>Show the field ONLY if:<br>[gs_otheranti_med2] = '1' and [hide_nonessential_fields(1)]='0' | Other Antithrombotic(s) Med 2 - Dose | dropdown<br>3 15 mg<br>5 Other<br>6 Unknown |
| 757 | gs_otheranti_dos2b<br><br>Show the field ONLY if:<br>[gs_otheranti_med2] = '2' and [hide_nonessential_fields(1)]='0' | Other Antithrombotic(s) Med 2 - Dose | dropdown<br>4 90 mg |
| 758 | gs_otheranti_dos2c<br><br>Show the field ONLY if:<br>[gs_otheranti_med2] = '3' and [hide_nonessential_fields(1)]='0' | Other Antithrombotic(s) Med 2 - Dose | dropdown<br>1 5 mg<br>2 10 mg |
| 759 | gs_otheranti_freq2<br><br>Show the field ONLY if:<br>[gs_otheranti_dos2a] <> '' or [gs_otheranti_dos2b] <> '' or [gs_otheranti_dos2c] <> '' and [hide_nonessential_fields(1)]='0' | Other Antithrombotic(s) Med 2 - Frequency | dropdown<br>1 Every Day<br>2 2 times a day<br>3 3 times a day<br>4 4 times a day<br>5 Other<br>6 Unknown |
| 760 | gs_persistentafib<br><br>Show the field ONLY if:<br>[gs_notadmit]<>'1' and [jc_disposition]<>'6' and [hide_nonessential_fields(1)]='0' | Persistent or paroxysmal Atrial Fibrillation/Flutter | radio<br>1 Yes<br>2 No<br><br>Custom alignment: RH |

| | | | |
|---|---|---|---|
| 761 | gs_afibdischarge<br><br>Show the field ONLY if:<br>[gs_notadmit]<>'1' and ([gs_persistentafib]='1' or [gs_med_hist(1)]='1') and [hide_nonessential_fields(1)]='0' | If atrial fib/flutter or history of PAF documented, was patient discharged on anticoagulation? | radio<br>1 Yes<br>2 No/ND<br>3 NC<br><br>Custom alignment: RH |
| 762 | gs_whynoatrialmeds<br><br>Show the field ONLY if:<br>[gs_afibdischarge] = '3' and [hide_nonessential_fields(1)]='0' | If NC, documented reasons for no anticoagulation<br>*Select all that apply.* | checkbox<br>13 gs_whynoatrialmeds___13 Allergy to or complication r/t warfarin or heparins (hx or current)<br>3 gs_whynoatrialmeds___3 Mental status<br>6 gs_whynoatrialmeds___6 Patient refused<br>1 gs_whynoatrialmeds___1 Risk for bleeding or discontinued due to bleeding<br>2 gs_whynoatrialmeds___2 Risk for falls<br>14 gs_whynoatrialmeds___14 Serious side effect to medication<br>5 gs_whynoatrialmeds___5 Terminal illness/Comfort Measures Only |
| 763 | gs_antihyper<br><br>Show the field ONLY if:<br>([site]='SMCA' or [site]='SMCH' or [site]='SMCW' or [site]='UMCB') and [gs_notadmit] <> '1' and [hide_nonessential_fields(1)]='0' | Antihypertensive Tx<br>*Select all that apply.* | checkbox<br>1 gs_antihyper___1 None prescribed/ND<br>2 gs_antihyper___2 None - contraindicated<br>3 gs_antihyper___3 ACE Inhibitors<br>4 gs_antihyper___4 ARB<br>5 gs_antihyper___5 Beta Blockers<br>6 gs_antihyper___6 Ca++ Channel Blockers<br>7 gs_antihyper___7 Diuretics<br>8 gs_antihyper___8 Other anti-hypertensive med<br><br>Field Annotation: @NONEOFTHEABOVE='1,2' |

| | | | |
|---|---|---|---|
| 764 | gs_cholredtx<br><br>Show the field ONLY if:<br>[gs_notadmit] <> '1' and [hide_nonessential_fields(1)]='0' | ✪ Cholesterol Reducing Tx<br>*Select all that apply.* | checkbox<br>1 \| gs_cholredtx___1 \| None prescribed/ND<br>2 \| gs_cholredtx___2 \| None - contraindicated<br>3 \| gs_cholredtx___3 \| Statin<br>4 \| gs_cholredtx___4 \| Fibrate<br>6 \| gs_cholredtx___6 \| Other med<br>7 \| gs_cholredtx___7 \| Niacin<br>8 \| gs_cholredtx___8 \| Absorption inhibitor<br>9 \| gs_cholredtx___9 \| PCSK 9 inhibitor<br><br>Field Annotation: @NONEOFTHEABOVE='1,2' |
| 765 | gs_statinmed<br><br>Show the field ONLY if:<br>([site]='SMCA' or [site]='SMCH' or [site]='SMCW' or [site]='UMCB') and [gs_cholredtx(3)] = '1' and [hide_nonessential_fields(1)]='0' | ✪ Statin Medication | dropdown<br>3 \| Amlodipine + Atorvastatin (Caduet)<br>6 \| Atorvastatin (Lipitor)<br>11 \| Ezetimibe + Simvastatin (Vytorin)<br>5 \| Fluvastatin (Lescol)<br>14 \| Fluvastatin XL (Lescol XL)<br>2 \| Lovastatin (Altoprev)<br>8 \| Lovastatin (Mevacor)<br>1 \| Lovastatin + Niacin (Advicor)<br>7 \| Pitavastatin (Livalo)<br>9 \| Pravastatin (Pravachol)<br>4 \| Rosuvastatin (Crestor)<br>12 \| Simvastatin (Zocor)<br>10 \| Simvastatin + Niacin (Simcor)<br>13 \| Sitagliptin + Simvastatin (Juvisync)<br><br>Field Annotation: @HIDECHOICE='13' |

| | | | |
|---|---|---|---|
| 766 | gs_statindose1<br><br>Show the field ONLY if:<br>[gs_statinmed] = '1' and [hide_nonessential_fields(1)]='0' | ✪ Statin Total Daily Dose<br>*Lovastatin + Niacin (Advicor)* | dropdown<br>100 20/500 mg<br>101 20/750 mg<br>102 20/1000 mg<br>103 40/1000 mg<br>104 Other<br>105 Unknown<br><br>Field Annotation: @HIDECHOICE='104' |
| 767 | gs_statindose2<br><br>Show the field ONLY if:<br>[gs_statinmed] = '2' and [hide_nonessential_fields(1)]='0' | ✪ Statin Total Daily Dose<br>*Lovastatin (Altoprev)* | dropdown<br>150 20 mg<br>151 40 mg<br>152 60 mg<br>153 Other<br>154 Unknown<br><br>Field Annotation: @HIDECHOICE='153' |
| 768 | gs_statindose3<br><br>Show the field ONLY if:<br>[gs_statinmed] = '3' and [hide_nonessential_fields(1)]='0' | ✪ Statin Total Daily Dose<br>*Amlodipine + Atorvastatin (Caduet)* | dropdown<br>200 2.5/10 mg<br>201 2.5/20 mg<br>202 2.5/40 mg<br>203 5/10 mg<br>204 5/20 mg<br>205 5/40 mg<br>206 5/80 mg<br>207 10/10 mg<br>208 10/20 mg<br>209 10/40 mg<br>210 10/80 mg<br>211 Other<br>212 Unknown<br><br>Field Annotation: @HIDECHOICE='211' |

| | | | |
|---|---|---|---|
| 769 | gs_statindose4<br><br>Show the field ONLY if:<br>[gs_statinmed] = '4' and [hide_nonessential_fields(1)]='0' | ✪ Statin Total Daily Dose<br>*Rosuvastatin (Crestor)* | dropdown<br>300 \| 5 mg<br>301 \| 10 mg<br>302 \| >= 20 mg<br>303 \| Other<br>304 \| Unknown<br><br>Field Annotation: @HIDECHOICE='303' |
| 770 | gs_statindose5<br><br>Show the field ONLY if:<br>[gs_statinmed] = '5' and [hide_nonessential_fields(1)]='0' | ✪ Statin Total Daily Dose<br>*Fluvastatin (Lescol)* | dropdown<br>400 \| 20 mg<br>401 \| 40 mg<br>402 \| 80 mg<br>404 \| Unknown |
| 771 | gs_statindose6<br><br>Show the field ONLY if:<br>[gs_statinmed] = '6' and [hide_nonessential_fields(1)]='0' | ✪ Statin Total Daily Dose<br>*Atorvastatin (Lipitor)* | dropdown<br>500 \| 10 mg<br>501 \| 20 mg<br>502 \| >= 40 mg<br>503 \| Other<br>504 \| Unknown<br><br>Field Annotation: @HIDECHOICE='503' |
| 772 | gs_statindose7<br><br>Show the field ONLY if:<br>[gs_statinmed] = '7' and [hide_nonessential_fields(1)]='0' | ✪ Statin Total Daily Dose<br>*Pitavastatin (Livalo)* | dropdown<br>650 \| 1 mg<br>651 \| 2 mg<br>652 \| 4 mg<br>655 \| Other<br>656 \| Unknown<br><br>Field Annotation: @HIDECHOICE='655' |

| | | | |
|---|---|---|---|
| 773 | gs_statindose8<br><br>Show the field ONLY if:<br>[gs_statinmed] = '8' and [hide_nonessential_fields(1)]='0' | ✪ Statin Total Daily Dose<br>*Lovastatin (Mevacor)* | dropdown<br>704 — 10 mg<br>700 — 20 mg<br>701 — 40 mg<br>702 — Other<br>703 — Unknown<br><br>Field Annotation: @HIDECHOICE='702' |
| 774 | gs_statindose9<br><br>Show the field ONLY if:<br>[gs_statinmed] = '9' and [hide_nonessential_fields(1)]='0' | ✪ Statin Total Daily Dose<br>*Pravastatin (Pravachol)* | dropdown<br>800 — 10 mg<br>801 — 20 mg<br>802 — 40 mg<br>803 — 80 mg<br>804 — Other<br>805 — Unknown<br><br>Field Annotation: @HIDECHOICE='804' |
| 775 | gs_statindose10<br><br>Show the field ONLY if:<br>[gs_statinmed] = '10' and [hide_nonessential_fields(1)]='0' | ✪ Statin Total Daily Dose<br>*Simvastatin + Niacin (Simcor)* | dropdown<br>900 — 20/500 mg<br>901 — 20/750 mg<br>902 — 20/1000 mg<br>903 — 40/500 mg<br>904 — 40/1000 mg<br>905 — 40/2000 mg (2 x 20/1000 mg)<br>906 — Other<br>907 — Unknown<br><br>Field Annotation: @HIDECHOICE='906' |

| | | | |
|---|---|---|---|
| 776 | gs_statindose11<br><br>Show the field ONLY if:<br>[gs_statinmed] = '11' and [hide_nonessential_fields(1)]='0' | ✪ Statin Total Daily Dose<br>*Ezetimibe + Simvastatin (Vytorin)* | dropdown<br>1000 — 10/10 mg<br>1001 — 10/20 mg<br>1002 — 10/40 mg<br>1003 — 10/80 mg<br>1004 — Other<br>1005 — Unknown<br><br>Field Annotation: @HIDECHOICE='1004' |
| 777 | gs_statindose12<br><br>Show the field ONLY if:<br>[gs_statinmed] = '12' and [hide_nonessential_fields(1)]='0' | ✪ Statin Total Daily Dose<br>*Simvastatin (Zocor)* | dropdown<br>1100 — 5 mg<br>1101 — 10 mg<br>1102 — 20 mg<br>1103 — 40 mg<br>1104 — 80 mg<br>1105 — Other<br>1106 — Unknown<br><br>Field Annotation: @HIDECHOICE='1105' |
| 778 | gs_statindose13<br><br>Show the field ONLY if:<br>[gs_statinmed] = '13' and [hide_nonessential_fields(1)]='0' | ✪ Statin Total Daily Dose<br>*Sitagliptin + Simvastatin (Juvisync)* | dropdown<br>1200 — 100mg/10mg<br>1201 — 100mg/20mg<br>1202 — 100mg/40mg<br>1203 — 50mg/10mg<br>1204 — 50mg/20mg<br>1205 — 50mg/40mg<br>1206 — Other<br>1207 — Unknown<br><br>Field Annotation: @HIDECHOICE='1206' |
| 779 | gs_statindose14<br><br>Show the field ONLY if:<br>[gs_statinmed] = '14' and [hide_nonessential_fields(1)]='0' | ✪ Statin Total Daily Dose<br>*Fluvastatin XL (Lescol XL)* | dropdown<br>1300 — 80 mg |
| 780 | gs_noguidedose | ✪ Documented Reason for Not Prescribing Guideline Recommended | checkbox |

| | | | |
|---|---|---|---|
| | Show the field ONLY if:<br>([site]='SMCA' or [site]='SMCH' or [site]='SMCW' or [site]='UMCB') and (((rounddown(datediff([jc_dob], [jc_admitdate], 'y', 'mdy', true), 0)<75) and ((([gs_statinmed]='6' and ([gs_statindose6]='500' or [gs_statindose6]='501')) or ([gs_statinmed]='5' and ([gs_statindose5]='400' or [gs_statindose5]='401' or [gs_statindose5]='402')) or ([gs_statinmed]='9' and ([gs_statindose9]='800' or [gs_statindose9]='801' or [gs_statindose9]='802' or [gs_statindose9]='803')) or ([gs_statinmed]='3' and ([gs_statindose3]='200' or [gs_statindose3]='201' or [gs_statindose3]='203' or [gs_statindose3]='204' or [gs_statindose3]='207' or [gs_statindose3]='208')) or ([gs_statinmed]='11' and ([gs_statindose11]='1000' or [gs_statindose11]='1001' or [gs_statindose11]='1002')) or ([gs_statinmed]='4' and ([gs_statindose4]='300' or [gs_statindose4]='301')) or ([gs_statinmed]='2' and ([gs_statindose2]='150' or [gs_statindose2]='151' or [gs_statindose2]='152')) or ([gs_statinmed]='8' and ([gs_statindose8]='700' or [gs_statindose8]='701' or [gs_statindose8]='704')) or ([gs_statinmed]='1' and ([gs_statindose1]='100' or [gs_statindose1]='101' or [gs_statindose1]='102' or [gs_statindose1]='103')) or ([gs_statinmed]='7' and ([gs_statindose7]='650' or [gs_statindose7]='651' or [gs_statindose7]='652')) or ([gs_statinmed]='12' and ([gs_statindose12]='1100' or [gs_statindose12]='1101' or [gs_statindose12]='1102' or [gs_statindose12]='1103')) or ([gs_statinmed]='14' and [gs_statindose14]='1300') or ([gs_statinmed]='10' and ([gs_statindose10]='900' or [gs_statindose10]='901' or [gs_statindose10]='902' or [gs_statindose10]='903' or [gs_statindose10] | Dose? | 1 \| gs_noguidedose___1 \| Intolerant to moderate (>75yr) or high (≤75yr) intensity statin<br>2 \| gs_noguidedose___2 \| No evidence of atherosclerosis (cerebral, coronary, or peripheral vascular disease)<br>3 \| gs_noguidedose___3 \| Other documented reason<br>0 \| gs_noguidedose___0 \| Unknown/ND<br><br>Field Annotation: @NONEOFTHEABOVE='0' |

| | | | |
|---|---|---|---|
| | ='904' or [gs_statindose10]='905')))) or ((rounddown(datediff([jc_dob], [jc_admitdate], 'y', 'mdy', true), 0)>=75) and (([gs_statinmed]='5' and ([gs_statindose5]='400' or [gs_statindose5]='401')) or ([gs_statinmed]='9' and ([gs_statindose9]='800' or [gs_statindose9]='801')) or ([gs_statinmed]='11' and [gs_statindose11]='1000') or ([gs_statinmed]='2' and [gs_statindose2]='150') or ([gs_statinmed]='8' and ([gs_statindose8]='700' or [gs_statindose8]='704')) or ([gs_statinmed]='1' and ([gs_statindose1]='100' or [gs_statindose1]='101' or [gs_statindose1]='102')) or ([gs_statinmed]='7' and [gs_statindose7]='650') or ([gs_statinmed]='12' and ([gs_statindose12]='1100' or [gs_statindose12]='1101'))))) and [hide_nonessential_fields(1)]='0' | | |
| 781 | jc_nostatindisc<br><br>Show the field ONLY if:<br>[gs_notadmit]<>'1' and [jc_disposition]<>'3' and [jc_disposition]<>'4' and [jc_disposition]<>'6' and [jc_disposition]<>'7' and [hide_nonessential_fields(1)]='0' | ✪ Documented reason for not prescribing a statin medication at discharge? | radio<br>1 Yes<br>2 No<br><br>Custom alignment: RH |
| 782 | gs_intensivestatin<br><br>Show the field ONLY if:<br>([site]='SMCA' or [site]='SMCH' or [site]='SMCW' or [site]='UMCB') and [gs_notadmit]<>'1' and [hide_nonessential_fields(1)]='0' | Intensive Statin Therapy | radio<br>1 Yes<br>2 No/ND<br>3 NC<br><br>Custom alignment: RH<br>Field Annotation: @HIDDEN |
| 783 | gs_diagdm<br><br>Show the field ONLY if:<br>([site]='SMCA' or [site]='SMCH' or [site]='SMCW' or [site]='UMCB') and [gs_notadmit]<>'1' and [hide_nonessential_fields(1)]='0' | New Diagnosis of Diabetes? | radio<br>1 Yes<br>2 No<br>3 ND<br><br>Custom alignment: RH |

| | | | |
|---|---|---|---|
| 784 | gs_diagdmbasis<br><br>Show the field ONLY if:<br>[gs_diagdm]='1' and [hide_nonessential_fields(1)]='0' | Basis for Diagnosis?<br>*Select all that apply.* | checkbox<br>1 gs_diagdmbasis___1 HbA1c<br>2 gs_diagdmbasis___2 Fasting Blood Sugar<br>3 gs_diagdmbasis___3 Oral Glucose Tolerance<br>4 gs_diagdmbasis___4 Test Other |
| 785 | gs_dmmed<br><br>Show the field ONLY if:<br>([site]='SMCA' or [site]='SMCH' or [site]='SMCW' or [site]='UMCB') and [gs_notadmit]<>'1' and [hide_nonessential_fields(1)]='0' | Diabetic Tx | checkbox<br>1 gs_dmmed___1 None prescribed/ND<br>2 gs_dmmed___2 None - contraindicated<br>3 gs_dmmed___3 Insulin<br>4 gs_dmmed___4 Oral agents<br>5 gs_dmmed___5 Other subcutaneous/injectable agents<br><br>Field Annotation: @HIDDEN @NONEOFTHEABOVE='1,2' |
| 786 | presdmtx<br><br>Show the field ONLY if:<br>[hide_nonessential_fields(1)]='0' | Anti-hyperglycemic medications prescribed? | radio<br>1 Yes<br>2 No<br>3 NC<br><br>Custom alignment: RH |
| 787 | classdmtx1<br><br>Show the field ONLY if:<br>[presdmtx] = '1' and [hide_nonessential_fields(1)]='0' | Anti-hyperglycemic Medication Class | dropdown<br>1 Biguanide<br>2 DPP-4 Inhibitor<br>3 GLP-1 receptor agonist<br>4 Insulin<br>5 SGLT2 Inhibitor<br>6 Sulfonylurea<br>7 Thiazolidinedione<br>8 Other subcutaneous/injectable agents<br>9 Other oral agents |
| 788 | meddmtx1_1<br><br>Show the field ONLY if:<br>[classdmtx1]='1' and [hide_nonessential_fields(1)]='0' | Anti-hyperglycemic Medication List | dropdown<br>1 Metformin |

| | | | |
|---|---|---|---|
| 789 | meddmtx1_2<br><br>Show the field ONLY if:<br>[classdmtx1]='2' and [hide_noness ential_fields(1)]='0' | Anti-hyperglycemic Medication List | dropdown<br>2 alogliptin<br>3 linagliptin<br>4 saxagliptin<br>5 sitagliptin |
| 790 | meddmtx1_3<br><br>Show the field ONLY if:<br>[classdmtx1]='3' and [hide_noness ential_fields(1)]='0' | Anti-hyperglycemic Medication List | dropdown<br>6 dulaglutide<br>7 exenatide<br>8 exenatide extended release<br>9 liraglutide<br>10 lixisenatide<br>11 meglitinide<br>12 semaglutide<br><br>Field Annotation: @HIDECHOICE='11' |
| 791 | meddmtx1_4<br><br>Show the field ONLY if:<br>[classdmtx1]='4' and [hide_noness ential_fields(1)]='0' | Anti-hyperglycemic Medication List | dropdown<br>13 Insulin |
| 792 | meddmtx1_5<br><br>Show the field ONLY if:<br>[classdmtx1]='5' and [hide_noness ential_fields(1)]='0' | Anti-hyperglycemic Medication List | dropdown<br>14 canagliflozin<br>15 dapagliflozin<br>16 empagliflozin<br>17 ertugliflozin |
| 793 | meddmtx1_6<br><br>Show the field ONLY if:<br>[classdmtx1]='6' and [hide_noness ential_fields(1)]='0' | Anti-hyperglycemic Medication List | dropdown<br>18 glimepiride<br>19 glipizide<br>20 glyburide |
| 794 | meddmtx1_7<br><br>Show the field ONLY if:<br>[classdmtx1]='7' and [hide_noness ential_fields(1)]='0' | Anti-hyperglycemic Medication List | dropdown<br>21 pioglitazone<br>22 rosiglitazone |

| | | | |
|---|---|---|---|
| 795 | meddmtx1_8<br><br>Show the field ONLY if:<br>[classdmtx1]='8' and [hide_nonessential_fields(1)]='0' | Anti-hyperglycemic Medication List | dropdown<br>23 Other subcutaneous/injectable agents |
| 796 | meddmtx1_9<br><br>Show the field ONLY if:<br>[classdmtx1]='9' and [hide_nonessential_fields(1)]='0' | Anti-hyperglycemic Medication List | dropdown<br>24 Other oral agents |
| 797 | classdmtx2<br><br>Show the field ONLY if:<br>[classdmtx1]<>'' and [hide_nonessential_fields(1)]='0' | Anti-hyperglycemic Medication Class | dropdown<br>1 Biguanide<br>2 DPP-4 Inhibitor<br>3 GLP-1 receptor agonist<br>4 Insulin<br>5 SGLT2 Inhibitor<br>6 Sulfonylurea<br>7 Thiazolidinedione<br>8 Other subcutaneous/injectable agents<br>9 Other oral agents |
| 798 | meddmtx2_1<br><br>Show the field ONLY if:<br>[classdmtx2]='1' and [hide_nonessential_fields(1)]='0' | Anti-hyperglycemic Medication List | dropdown<br>1 Metformin |
| 799 | meddmtx2_2<br><br>Show the field ONLY if:<br>[classdmtx2]='2' and [hide_nonessential_fields(1)]='0' | Anti-hyperglycemic Medication List | dropdown<br>2 alogliptin<br>3 linagliptin<br>4 saxagliptin<br>5 sitagliptin |

| | | | |
|---|---|---|---|
| 800 | meddmtx2_3<br><br>Show the field ONLY if:<br>[classdmtx2]='3' and [hide_nonessential_fields(1)]='0' | Anti-hyperglycemic Medication List | dropdown<br>6 dulaglutide<br>7 exenatide<br>8 exenatide extended release<br>9 liraglutide<br>10 lixisenatide<br>11 meglitinide<br>12 semaglutide<br><br>Field Annotation: @HIDECHOICE='11' |
| 801 | meddmtx2_4<br><br>Show the field ONLY if:<br>[classdmtx2]='4' and [hide_nonessential_fields(1)]='0' | Anti-hyperglycemic Medication List | dropdown<br>13 Insulin |
| 802 | meddmtx2_5<br><br>Show the field ONLY if:<br>[classdmtx2]='5' and [hide_nonessential_fields(1)]='0' | Anti-hyperglycemic Medication List | dropdown<br>14 canagliflozin<br>15 dapagliflozin<br>16 empagliflozin<br>17 ertugliflozin |
| 803 | meddmtx2_6<br><br>Show the field ONLY if:<br>[classdmtx2]='6' and [hide_nonessential_fields(1)]='0' | Anti-hyperglycemic Medication List | dropdown<br>18 glimepiride<br>19 glipizide<br>20 glyburide |
| 804 | meddmtx2_7<br><br>Show the field ONLY if:<br>[classdmtx2]='7' and [hide_nonessential_fields(1)]='0' | Anti-hyperglycemic Medication List | dropdown<br>21 pioglitazone<br>22 rosiglitazone |
| 805 | meddmtx2_8<br><br>Show the field ONLY if:<br>[classdmtx2]='8' and [hide_nonessential_fields(1)]='0' | Anti-hyperglycemic Medication List | dropdown<br>23 Other subcutaneous/injectable agents |
| 806 | meddmtx2_9<br><br>Show the field ONLY if:<br>[classdmtx2]='9' and [hide_nonessential_fields(1)]='0' | Anti-hyperglycemic Medication List | dropdown<br>24 Other oral agents |

| | | | |
|---|---|---|---|
| 807 | classdmtx3<br><br>Show the field ONLY if:<br>[classdmtx2]<>'' and [hide_nonessential_fields(1)]='0' | Anti-hyperglycemic Medication Class | dropdown<br>1 Biguanide<br>2 DPP-4 Inhibitor<br>3 GLP-1 receptor agonist<br>4 Insulin<br>5 SGLT2 Inhibitor<br>6 Sulfonylurea<br>7 Thiazolidinedione<br>8 Other subcutaneous/injectable agents<br>9 Other oral agents |
| 808 | meddmtx3_1<br><br>Show the field ONLY if:<br>[classdmtx3]='1' and [hide_nonessential_fields(1)]='0' | Anti-hyperglycemic Medication List | dropdown<br>1 Metformin |
| 809 | meddmtx3_2<br><br>Show the field ONLY if:<br>[classdmtx3]='2' and [hide_nonessential_fields(1)]='0' | Anti-hyperglycemic Medication List | dropdown<br>2 alogliptin<br>3 linagliptin<br>4 saxagliptin<br>5 sitagliptin |
| 810 | meddmtx3_3<br><br>Show the field ONLY if:<br>[classdmtx3]='3' and [hide_nonessential_fields(1)]='0' | Anti-hyperglycemic Medication List | dropdown<br>6 dulaglutide<br>7 exenatide<br>8 exenatide extended release<br>9 liraglutide<br>10 lixisenatide<br>11 meglitinide<br>12 semaglutide<br><br>Field Annotation: @HIDECHOICE='11' |
| 811 | meddmtx3_4<br><br>Show the field ONLY if:<br>[classdmtx3]='4' and [hide_nonessential_fields(1)]='0' | Anti-hyperglycemic Medication List | dropdown<br>13 Insulin |

| | | | |
|---|---|---|---|
| 812 | meddmtx3_5<br><br>Show the field ONLY if:<br>[classdmtx3]='5' and [hide_noness ential_fields(1)]='0' | Anti-hyperglycemic Medication List | dropdown<br>14 canagliflozin<br>15 dapagliflozin<br>16 empagliflozin<br>17 ertugliflozin |
| 813 | meddmtx3_6<br><br>Show the field ONLY if:<br>[classdmtx3]='6' and [hide_noness ential_fields(1)]='0' | Anti-hyperglycemic Medication List | dropdown<br>18 glimepiride<br>19 glipizide<br>20 glyburide |
| 814 | meddmtx3_7<br><br>Show the field ONLY if:<br>[classdmtx3]='7' and [hide_noness ential_fields(1)]='0' | Anti-hyperglycemic Medication List | dropdown<br>21 pioglitazone<br>22 rosiglitazone |
| 815 | meddmtx3_8<br><br>Show the field ONLY if:<br>[classdmtx3]='8' and [hide_noness ential_fields(1)]='0' | Anti-hyperglycemic Medication List | dropdown<br>23 Other subcutaneous/injectable agents |
| 816 | meddmtx3_9<br><br>Show the field ONLY if:<br>[classdmtx3]='9' and [hide_noness ential_fields(1)]='0' | Anti-hyperglycemic Medication List | dropdown<br>24 Other oral agents |
| 817 | classdmtx4<br><br>Show the field ONLY if:<br>[classdmtx3]<>'' and [hide_noness ential_fields(1)]='0' | Anti-hyperglycemic Medication Class | dropdown<br>1 Biguanide<br>2 DPP-4 Inhibitor<br>3 GLP-1 receptor agonist<br>4 Insulin<br>5 SGLT2 Inhibitor<br>6 Sulfonylurea<br>7 Thiazolidinedione<br>8 Other subcutaneous/injectable agents<br>9 Other oral agents |

| | | | |
|---|---|---|---|
| 818 | meddmtx4_1<br><br>Show the field ONLY if:<br>[classdmtx4]='1' and [hide_noness ential_fields(1)]='0' | Anti-hyperglycemic Medication List | dropdown<br>1 Metformin |
| 819 | meddmtx4_2<br><br>Show the field ONLY if:<br>[classdmtx4]='2' and [hide_noness ential_fields(1)]='0' | Anti-hyperglycemic Medication List | dropdown<br>2 alogliptin<br>3 linagliptin<br>4 saxagliptin<br>5 sitagliptin |
| 820 | meddmtx4_3<br><br>Show the field ONLY if:<br>[classdmtx4]='3' and [hide_noness ential_fields(1)]='0' | Anti-hyperglycemic Medication List | dropdown<br>6 dulaglutide<br>7 exenatide<br>8 exenatide extended release<br>9 liraglutide<br>10 lixisenatide<br>11 meglitinide<br>12 semaglutide<br><br>Field Annotation: @HIDECHOICE='11' |
| 821 | meddmtx4_4<br><br>Show the field ONLY if:<br>[classdmtx4]='4' and [hide_noness ential_fields(1)]='0' | Anti-hyperglycemic Medication List | dropdown<br>13 Insulin |
| 822 | meddmtx4_5<br><br>Show the field ONLY if:<br>[classdmtx4]='5' and [hide_noness ential_fields(1)]='0' | Anti-hyperglycemic Medication List | dropdown<br>14 canagliflozin<br>15 dapagliflozin<br>16 empagliflozin<br>17 ertugliflozin |
| 823 | meddmtx4_6<br><br>Show the field ONLY if:<br>[classdmtx4]='6' and [hide_noness ential_fields(1)]='0' | Anti-hyperglycemic Medication List | dropdown<br>18 glimepiride<br>19 glipizide<br>20 glyburide |

| | | | |
|---|---|---|---|
| 824 | meddmtx4_7<br><br>Show the field ONLY if:<br>[classdmtx4]='7' and [hide_nonessential_fields(1)]='0' | Anti-hyperglycemic Medication List | dropdown<br>21 pioglitazone<br>22 rosiglitazone |
| 825 | meddmtx4_8<br><br>Show the field ONLY if:<br>[classdmtx4]='8' and [hide_nonessential_fields(1)]='0' | Anti-hyperglycemic Medication List | dropdown<br>23 Other subcutaneous/injectable agents |
| 826 | meddmtx4_9<br><br>Show the field ONLY if:<br>[classdmtx4]='9' and [hide_nonessential_fields(1)]='0' | Anti-hyperglycemic Medication List | dropdown<br>24 Other oral agents |
| 827 | dmtxreason<br><br>Show the field ONLY if:<br>([site]='SMCA' or [site]='SMCH' or [site]='SMCW' or [site]='UMCB') and ([presdmtx]='2' or ([presdmtx]='1' and (([classdmtx1]='' or ([classdmtx1]<>'3' and [classdmtx1]<>'5')) and ([classdmtx2]='' or ([classdmtx2]<>'3' and [classdmtx2]<>'5')) and ([classdmtx3]='' or ([classdmtx3]<>'3' and [classdmtx3]<>'5')) and ([classdmtx4]='' or ([classdmtx4]<>'3' and [classdmtx4]<>'5'))))) and [hide_nonessential_fields(1)]='0' | Was there a documented reason for not prescribing a medication with proven CVD benefit? | radio<br>1 Yes<br>2 No/ND<br><br>Custom alignment: RH |
| 828 | dxfupsched<br><br>Show the field ONLY if:<br>[hide_nonessential_fields(1)]='0' | Follow-up appointment scheduled for diabetes management? | radio<br>1 Yes<br>2 No/ND<br>3 NC<br><br>Custom alignment: RH |
| 829 | dxfupscheddt_precision<br><br>Show the field ONLY if:<br>[dxfupsched]='1' and [hide_nonessential_fields(1)]='0' | Date of diabetes management follow-up visit precision | dropdown<br>5 Date & Time (MM/DD/YYYY HH24:MM)<br>3 Date Only (MM/DD/YYYY)<br>0 Unknown<br><br>Custom alignment: RH |

| | | | |
|---|---|---|---|
| 830 | dxfupscheddt<br><br>Show the field ONLY if:<br>[dxfupsched]='1' and [dxfupscheddt_precision]<>'0' and [hide_nonessential_fields(1)]='0' | Date of diabetes management follow-up visit<br>*If Date Only, enter 00:00 for time.* | text (datetime_mdy, Min: 2015-01-01 00:00, Max: 2099-12-31 23:59)<br>Custom alignment: RH |
| 831 | gs_antismokingtx<br><br>Show the field ONLY if:<br>([site]='SMCA' or [site]='SMCH' or [site]='SMCW' or [site]='UMCB') and [gs_med_hist(9)]='1' and [hide_nonessential_fields(1)]='0' | Anti-Smoking Tx | radio<br>1 Yes<br>2 No/ND<br>3 NC<br><br>Custom alignment: RH |
| 832 | gs_smokingmed<br><br>Show the field ONLY if:<br>[gs_antismokingtx]='1' and [gs_smokingmednot]='' and [hide_nonessential_fields(1)]='0' | Smoking Cessation Therapies Prescribed | checkbox<br>1 gs_smokingmed___1 Counseling<br>2 gs_smokingmed___2 Over the Counter Nicotine Replacement Therapy<br>3 gs_smokingmed___3 Prescription Medications<br>4 gs_smokingmed___4 Other |
| 833 | gs_smokingmednot<br><br>Show the field ONLY if:<br>[gs_antismokingtx]='1' and [hide_nonessential_fields(1)]='0' | Smoking Cessation Therapies - Treatment not specified | radio<br>1 Treatment not specified |
| 834 | gs_antidepmed<br><br>Show the field ONLY if:<br>([site]='SMCA' or [site]='SMCH' or [site]='SMCW' or [site]='UMCB') and [gs_notadmit]<>'1' and [hide_nonessential_fields(1)]='0' | Was the patient prescribed any antidepressant class of medication at discharge? | dropdown<br>1 Yes, SSRI<br>2 Yes, any other antidepressant class<br>3 No/ND<br><br>Custom alignment: RH |
| 835 | gs_lifestyle<br><br>Show the field ONLY if:<br>([site]='SMCA' or [site]='SMCH' or [site]='SMCW' or [site]='UMCB') and [gs_notadmit]<>'1' and [hide_nonessential_fields(1)]='0' | Section Header: *Other Lifestyle Interventions*<br>Reducing weight and/or increasing activity recommendations | radio<br>1 Yes<br>2 No/ND<br>3 NC<br><br>Custom alignment: RH |

| | | | |
|---|---|---|---|
| 836 | gs_adadiet<br><br>Show the field ONLY if:<br>([site]='SMCA' or [site]='SMCH' or [site]='SMCW' or [site]='UMCB') and [gs_notadmit]<>'1' and [hide_nonessential_fields(1)]='0' | TLC Diet or equivalent | radio<br>1 Yes<br>2 No/ND<br>3 NC<br><br>Custom alignment: RH |
| 837 | gs_sodiumdiet<br><br>Show the field ONLY if:<br>([site]='SMCA' or [site]='SMCH' or [site]='SMCW' or [site]='UMCB') and [gs_notadmit]<>'1' and [hide_nonessential_fields(1)]='0' | Antihypertensive Diet | radio<br>1 Yes<br>2 No/ND<br>3 NC<br><br>Custom alignment: RH |
| 838 | gs_dmteaching<br><br>Show the field ONLY if:<br>([site]='SMCA' or [site]='SMCH' or [site]='SMCW' or [site]='UMCB') and [gs_notadmit]<>'1' and [hide_nonessential_fields(1)]='0' | Was Diabetes Teaching Provided? | radio<br>1 Yes<br>2 No/ND<br>4 Not Applicable<br><br>Custom alignment: RH |
| 839 | jc_edurisk<br><br>Show the field ONLY if:<br>([treatment_recvd]='1' or [gs_notadmit]<>'1') and ([gs_comfortonly]='' or [gs_comfortonly]='4') and [hide_nonessential_fields(1)]='0' | Section Header: *Stroke Education*<br><br>Risk Factors for Stroke | radio<br>1 Yes<br>2 No<br><br>Custom alignment: RH |
| 840 | jc_eduwarning<br><br>Show the field ONLY if:<br>([treatment_recvd]='1' or [gs_notadmit]<>'1') and ([gs_comfortonly]='' or [gs_comfortonly]='4') and [hide_nonessential_fields(1)]='0' | Stroke Warning Signs and Symptoms | radio<br>1 Yes<br>2 No<br><br>Custom alignment: RH |
| 841 | jc_eduems<br><br>Show the field ONLY if:<br>([treatment_recvd]='1' or [gs_notadmit]<>'1') and ([gs_comfortonly]='' or [gs_comfortonly]='4') and [hide_nonessential_fields(1)]='0' | How to activate EMS for stroke | radio<br>1 Yes<br>2 No<br><br>Custom alignment: RH |

| | | | |
|---|---|---|---|
| 842 | jc_edufollowup<br><br>Show the field ONLY if:<br>([treatment_recvd]='1' or [gs_notadmit]<>'1') and ([gs_comfortonly]='' or [gs_comfortonly]='4') and [hide_nonessential_fields(1)]='0' | Need for followup after discharge | radio<br>1 \| Yes<br>2 \| No<br><br>Custom alignment: RH |
| 843 | jc_edumeds<br><br>Show the field ONLY if:<br>([treatment_recvd]='1' or [gs_notadmit]<>'1') and ([gs_comfortonly]='' or [gs_comfortonly]='4') and [hide_nonessential_fields(1)]='0' | Their prescribed medications | radio<br>1 \| Yes<br>2 \| No<br><br>Custom alignment: RH |
| 844 | edu_thrombolytic<br><br>Show the field ONLY if:<br>([site]='SMCA' or [site]='SMCH' or [site]='SMCW' or [site]='UMCB') and ([treatment_type(IVtPA)]='1' or [gs_ivthroinit]='1' or [gs_ivtpaoutside]='1') and [hide_nonessential_fields(1)]='0' and [gs_comfortonly]<>'1' and [gs_comfortonly]<>'2' and [gs_comfortonly]<>'3' and [jc_disposition]<>'2' and [jc_disposition]<>'3' and [jc_disposition]<>'6' and [jc_disposition]<>'7' and [jc_disposition]<>'8' | ✪ If the patient received IV thrombolysis, was fibrinolytic education documented? | yesno<br>1 \| Yes<br>0 \| No<br><br>Custom alignment: RH |

| | | | |
|---|---|---|---|
| 845 | edu_arteriogram<br><br>Show the field ONLY if:<br>([site]='SMCA' or [site]='SMCH' or [site]='SMCW' or [site]='UMCB') and ([treatment_type(IAtPA)]='1' or [treatment_type(MER)]='1' or [gs_iatpa]='1' or [procedures_interventions(2)]='1' or [procedures_interventions(6)]='1') and ([gs_stroketype]='2' or [gs_stroketype]='3' or [gs_stroketype]='4' or [gs_stroketype]='5' or [gs_stroketype]='6') and [hide_nonessential_fields(1)]='0' and [gs_comfortonly]<>'1' and [gs_comfortonly]<>'2' and [gs_comfortonly]<>'3' and [jc_disposition]<>'2' and [jc_disposition]<>'3' and [jc_disposition]<>'6' and [jc_disposition]<>'7' and [jc_disposition]<>'8' | ✪ If the patient had a thrombectomy, coil, or angiogram, was arteriogram education documented? | yesno<br>1 \| Yes<br>0 \| No<br><br>Custom alignment: RH |
| 846 | jc_assessrehab<br><br>Show the field ONLY if:<br>([jc_disposition]='1' or [jc_disposition]='5' or [jc_disposition]='8') and ([gs_comfortonly]='' or [gs_comfortonly]='4') and [hide_nonessential_fields(1)]='0' | Section Header: *Stroke Rehabilitation*<br><br>Patient assessed for and/or received rehabilitation services during this hospitalization? | radio<br>1 \| Yes<br>2 \| No<br><br>Custom alignment: RH |
| 847 | gs_rehabsvcs<br><br>Show the field ONLY if:<br>([site]='SMCA' or [site]='SMCH' or [site]='SMCW' or [site]='UMCB') and [jc_assessrehab]='1' and [hide_nonessential_fields(1)]='0' | Check all rehab services that patient received or was assessed for: | checkbox<br>1 \| gs_rehabsvcs___1 \| Patient received rehabilitation services during hospitalization<br>2 \| gs_rehabsvcs___2 \| Patient transferred to rehabilitation facility<br>3 \| gs_rehabsvcs___3 \| Patient referred to rehabilitation services following discharge<br>4 \| gs_rehabsvcs___4 \| Patient ineligible to receive rehabilitation services because symptoms resolved<br>5 \| gs_rehabsvcs___5 \| Patient ineligible to receive rehabilitation services due to impairment (i.e. poor prognosis, patient unable to tolerate rehabilitation therapeutic regimen) |

| | | | |
|---|---|---|---|
| 848 | cryp_ultrasound<br><br>Show the field ONLY if:<br>([site]='SMCA' or [site]='SMCH' or [site]='SMCW' or [site]='UMCB') and ([treatment_recvd]='1' or [gs_notadmit]<>'1') | Section Header: *Stroke Diagnostic Tests and Interventions*<br><br>Cardiac Ultrasound | radio<br>1 Performed during this admission or in the 3 months prior<br>2 Planned post discharge<br>3 Not performed or planned |
| 849 | cryp_icmilr<br><br>Show the field ONLY if:<br>([site]='SMCA' or [site]='SMCH' or [site]='SMCW' or [site]='UMCB') and ([treatment_recvd]='1' or [gs_notadmit]<>'1') | Extended Implantable Cardiac Rhythm Monitoring | radio<br>1 Performed during this admission or in the 3 months prior<br>2 Planned post discharge<br>3 Not performed or planned |
| 850 | cryp_imaging<br><br>Show the field ONLY if:<br>([site]='SMCA' or [site]='SMCH' or [site]='SMCW' or [site]='UMCB') and ([treatment_recvd]='1' or [gs_notadmit]<>'1') | Carotid Imaging | radio<br>1 Performed during this admission or in the 3 months prior<br>2 Planned post discharge<br>3 Not performed or planned |
| 851 | cryp_hypercoa<br><br>Show the field ONLY if:<br>([site]='SMCA' or [site]='SMCH' or [site]='SMCW' or [site]='UMCB') and ([treatment_recvd]='1' or [gs_notadmit]<>'1') | Hypercoagulability Testing | radio<br>1 Performed during this admission or in the 3 months prior<br>2 Planned post discharge<br>3 Not performed or planned |
| 852 | cryp_reva<br><br>Show the field ONLY if:<br>([site]='SMCA' or [site]='SMCH' or [site]='SMCW' or [site]='UMCB') and ([treatment_recvd]='1' or [gs_notadmit]<>'1') | Carotid Revascularization | radio<br>1 Performed during this admission or in the 3 months prior<br>2 Planned post discharge<br>3 Not performed or planned |
| 853 | cryp_intracran<br><br>Show the field ONLY if:<br>([site]='SMCA' or [site]='SMCH' or [site]='SMCW' or [site]='UMCB') and ([treatment_recvd]='1' or [gs_notadmit]<>'1') | Intracranial Vascular Imaging | radio<br>1 Performed during this admission or in the 3 months prior<br>2 Planned post discharge<br>3 Not performed or planned |

| | | | |
|---|---|---|---|
| 854 | cryp_rhythm<br><br>Show the field ONLY if:<br>([site]='SMCA' or [site]='SMCH' or [site]='SMCW' or [site]='UMCB') and ([treatment_recvd]='1' or [gs_notadmit]<>'1') | Extended Surface Cardiac Rhythm Monitoring > 7 Days | radio<br>1 Performed during this admission or in the 3 months prior<br>2 Planned post discharge<br>3 Not performed or planned |
| 855 | cryp_cardiacmon<br><br>Show the field ONLY if:<br>([site]='SMCA' or [site]='SMCH' or [site]='SMCW' or [site]='UMCB') and ([treatment_recvd]='1' or [gs_notadmit]<>'1') | Short-term Cardiac Rhythm Monitoring < = 7 Days | radio<br>1 Performed during this admission or in the 3 months prior<br>2 Planned post discharge<br>3 Not performed or planned |
| 856 | navion_status<br><br>Show the field ONLY if:<br>[hide_nonessential_fields(1)]='0' | Section Header: *Navion Status*<br><br>Navion Status | radio<br>1 Ready for Navion<br>2 Assigned to Navion Manager<br>3 Completed by Navion<br>4 Needs Review by Ascension Stroke Coordinator<br>5 Reviewed by Ascension Stroke Coordinator<br>6 Completed by Ascension Stroke Coordinator<br>7 Needs Review by Ascension Stroke Program Manager |
| 857 | navion_date<br><br>Show the field ONLY if:<br>[navion_status]='3' or [navion_status]='4' or [navion_date]<>'' and [hide_nonessential_fields(1)]='0' | Date Completed by Navion | text (date_mdy), Required |
| 858 | navion_manager<br><br>Show the field ONLY if:<br>[navion_status]='2' or [navion_manager]<>'' and [hide_nonessential_fields(1)]='0' | Notes for Navion Manager Review | notes |
| 859 | navion_review<br><br>Show the field ONLY if:<br>[navion_status]='4' or [navion_review]<>'' and [hide_nonessential_fields(1)]='0' | Notes for Ascension Stroke Coordinator Review | notes |

| | | | |
|---|---|---|---|
| 860 | discharge_complete | Section Header: *Form Status*<br>Complete? | dropdown<br>0 Incomplete<br>1 Unverified<br>2 Complete |

Instrument: **Fallouts** (fallouts) Collapse

| | | | |
|---|---|---|---|
| 861 | desc_fallout_stk_1<br><br>Show the field ONLY if:<br>[calc_stk_1]='0' or [calc_gwtg_vte_prophylaxis]='0' | Section Header: *Fallout: VTE ProphylaxisSITE: [site] MRN: [mrn]Patient Name:[jc_lastname], [jc_firstname]Form / PMT ID:[patient_os_id]* | descriptive |
| 862 | fallout_rnmd_stk_1<br><br>Show the field ONLY if:<br>[calc_stk_1]='0' or [calc_gwtg_vte_prophylaxis]='0' | Fallout by RN or MD? | checkbox<br>1 fallout_rnmd_stk_1___1 RN<br>2 fallout_rnmd_stk_1___2 MD<br><br>Custom alignment: RH |
| 863 | fallout_detail_stk_1<br><br>Show the field ONLY if:<br>[calc_stk_1]='0' or [calc_gwtg_vte_prophylaxis]='0' | Fallout Details: VTE Prophylaxis | notes |
| 864 | fallout_loc_stk_1<br><br>Show the field ONLY if:<br>[calc_stk_1]='0' or [calc_gwtg_vte_prophylaxis]='0' | Fallout Location | checkbox<br>1 fallout_loc_stk_1___1 ED<br>2 fallout_loc_stk_1___2 ICU<br>3 fallout_loc_stk_1___3 Floor<br><br>Custom alignment: RH |

| | | | |
|---|---|---|---|
| 865 | fallout_unit_stk_1<br><br>Show the field ONLY if:<br>[calc_stk_1]='0' or [calc_gwtg_vte_prophylaxis]='0' | Fallout Unit | dropdown<br>1 ED<br>2 ICU<br>8 IMC<br>9 SICU<br>10 MICU<br>3 3rd Floor<br>4 4th Floor<br>11 4 East<br>12 4 West<br>5 5th Floor<br>6 6th Floor<br>7 7th Floor<br><br>Custom alignment: RH |
| 866 | fallout_notified_stk_1<br><br>Show the field ONLY if:<br>[calc_stk_1]='0' or [calc_gwtg_vte_prophylaxis]='0' | Fallout Notification Sent To:<br>*Name or Email* | text<br>Custom alignment: RH |
| 867 | fallout_date_stk_1<br><br>Show the field ONLY if:<br>[calc_stk_1]='0' or [calc_gwtg_vte_prophylaxis]='0' | Date Notified | text (date_mdy, Min: 2016-01-01, Max: 2099-12-31)<br>Custom alignment: RH |
| 868 | fallout_alert_stk_1<br><br>Show the field ONLY if:<br>[calc_stk_1]='0' or [calc_gwtg_vte_prophylaxis]='0' | Email Notification: | radio<br>1 Send Email<br>2 Email(s) Sent<br><br>Custom alignment: RH |
| 869 | fallout_verify_stk_1<br><br>Show the field ONLY if:<br>[calc_stk_1]='0' or [calc_gwtg_vte_prophylaxis]='0' | Reviewed by Stroke Coordinator: | checkbox<br>1 fallout_verify_stk_1___1 |
| 870 | fallout_resp_desc_stk_1<br><br>Show the field ONLY if:<br>[calc_stk_1]='0' or [calc_gwtg_vte_prophylaxis]='0' | Fallout Review: [fallout_review_stk_1]<br>Fallout Response: [fallout_response_stk_1] | descriptive |

| | | | |
|---|---|---|---|
| 871 | desc_fallout_stk_2<br><br>Show the field ONLY if:<br>[calc_stk_2] = '0' or [calc_gwtg_antithrombotics] = '0' | Section Header: *Fallout: Discharged on Antithrombotic TherapySITE: [site] MRN: [mrn]Patient Name:[jc_lastname], [jc_firstname]Form / PMT ID:[patient_os_id]* | descriptive |
| 872 | fallout_detail_stk_2<br><br>Show the field ONLY if:<br>[calc_stk_2] = '0' or [calc_gwtg_antithrombotics] = '0' | Fallout Details: Discharged on Antithrombotic Therapy | notes |
| 873 | fallout_loc_stk_2<br><br>Show the field ONLY if:<br>[calc_stk_2] = '0' or [calc_gwtg_antithrombotics] = '0' | Fallout Location | checkbox<br>1 \| fallout_loc_stk_2___1 \| ED<br>2 \| fallout_loc_stk_2___2 \| ICU<br>3 \| fallout_loc_stk_2___3 \| Floor<br><br>Custom alignment: RH |
| 874 | fallout_unit_stk_2<br><br>Show the field ONLY if:<br>[calc_stk_2] = '0' or [calc_gwtg_antithrombotics] = '0' | Fallout Unit | dropdown<br>1 \| ED<br>2 \| ICU<br>8 \| IMC<br>9 \| SICU<br>10 \| MICU<br>3 \| 3rd Floor<br>4 \| 4th Floor<br>11 \| 4 East<br>12 \| 4 West<br>5 \| 5th Floor<br>6 \| 6th Floor<br>7 \| 7th Floor<br><br>Custom alignment: RH |
| 875 | fallout_rnmd_stk_2<br><br>Show the field ONLY if:<br>[calc_stk_2] = '0' or [calc_gwtg_antithrombotics] = '0' | Fallout by RN or MD? | checkbox<br>1 \| fallout_rnmd_stk_2___1 \| RN<br>2 \| fallout_rnmd_stk_2___2 \| MD<br><br>Custom alignment: RH |

| | | | |
|---|---|---|---|
| 876 | fallout_notified_stk_2<br><br>Show the field ONLY if:<br>[calc_stk_2] = '0' or [calc_gwtg_antithrombotics] = '0' | Fallout Notification Sent To:<br>*Name or Email* | text<br>Custom alignment: RH |
| 877 | fallout_date_stk_2<br><br>Show the field ONLY if:<br>[calc_stk_2] = '0' or [calc_gwtg_antithrombotics] = '0' | Date Notified | text (date_mdy, Min: 2016-01-01, Max: 2099-12-31)<br>Custom alignment: RH |
| 878 | fallout_alert_stk_2<br><br>Show the field ONLY if:<br>[calc_stk_2] = '0' or [calc_gwtg_antithrombotics] = '0' | Email Notification: | radio<br>1 \| Send Email<br>2 \| Email(s) Sent<br><br>Custom alignment: RH |
| 879 | fallout_verify_stk_2<br><br>Show the field ONLY if:<br>[calc_stk_2] = '0' or [calc_gwtg_antithrombotics] = '0' | Reviewed by Stroke Coordinator: | checkbox<br>1 \| fallout_verify_stk_2___1 \| |
| 880 | fallout_resp_desc_stk_2<br><br>Show the field ONLY if:<br>[calc_stk_2] = '0' or [calc_gwtg_antithrombotics] = '0' | Fallout Review: [fallout_review_stk_2]<br>Fallout Response: [fallout_response_stk_2] | descriptive |
| 881 | desc_fallout_stk_3<br><br>Show the field ONLY if:<br>[calc_stk_3] = '0' or [calc_gwtg_anticoag_afib] = '0' | Section Header: *Fallout: Anticoagulation Therapy for Atrial Fibrillation/FlutterSITE: [site] MRN: [mrn]Patient Name:[jc_lastname], [jc_firstname]Form / PMT ID:[patient_os_id]* | descriptive |
| 882 | fallout_detail_stk_3<br><br>Show the field ONLY if:<br>[calc_stk_3] = '0' or [calc_gwtg_anticoag_afib] = '0' | Fallout Details: Anticoagulation Therapy for Atrial Fibrillation/Flutter | notes |
| 883 | fallout_loc_stk_3<br><br>Show the field ONLY if:<br>[calc_stk_3] = '0' or [calc_gwtg_anticoag_afib] = '0' | Fallout Location | checkbox<br>1 \| fallout_loc_stk_3___1 \| ED<br>2 \| fallout_loc_stk_3___2 \| ICU<br>3 \| fallout_loc_stk_3___3 \| Floor<br><br>Custom alignment: RH |

| | | | |
|---|---|---|---|
| 884 | fallout_unit_stk_3<br><br>Show the field ONLY if:<br>[calc_stk_3] = '0' or [calc_gwtg_anticoag_afib] = '0' | Fallout Unit | dropdown<br>1 ED<br>2 ICU<br>8 IMC<br>9 SICU<br>10 MICU<br>3 3rd Floor<br>4 4th Floor<br>11 4 East<br>12 4 West<br>5 5th Floor<br>6 6th Floor<br>7 7th Floor<br><br>Custom alignment: RH |
| 885 | fallout_rnmd_stk_3<br><br>Show the field ONLY if:<br>[calc_stk_3] = '0' or [calc_gwtg_anticoag_afib] = '0' | Fallout by RN or MD? | checkbox<br>1 fallout_rnmd_stk_3___1 RN<br>2 fallout_rnmd_stk_3___2 MD<br><br>Custom alignment: RH |
| 886 | fallout_notified_stk_3<br><br>Show the field ONLY if:<br>[calc_stk_3] = '0' or [calc_gwtg_anticoag_afib] = '0' | Fallout Notification Sent To:<br>*Name or Email* | text<br>Custom alignment: RH |
| 887 | fallout_date_stk_3<br><br>Show the field ONLY if:<br>[calc_stk_3] = '0' or [calc_gwtg_anticoag_afib] = '0' | Date Notified | text (date_mdy, Min: 2016-01-01, Max: 2099-12-31)<br>Custom alignment: RH |
| 888 | fallout_alert_stk_3<br><br>Show the field ONLY if:<br>[calc_stk_3] = '0' or [calc_gwtg_anticoag_afib] = '0' | Email Notification: | radio<br>1 Send Email<br>2 Email(s) Sent<br><br>Custom alignment: RH |

| | | | |
|---|---|---|---|
| 889 | fallout_verify_stk_3<br><br>Show the field ONLY if:<br>[calc_stk_3] = '0' or [calc_gwtg_anticoag_afib] = '0' | Reviewed by Stroke Coordinator: | checkbox<br>1 \| fallout_verify_stk_3___1 \| |
| 890 | fallout_resp_desc_stk_3<br><br>Show the field ONLY if:<br>[calc_stk_3] = '0' or [calc_gwtg_anticoag_afib] = '0' | Fallout Review: [fallout_review_stk_3]<br>Fallout Response: [fallout_response_stk_3] | descriptive |
| 891 | desc_fallout_stk_4<br><br>Show the field ONLY if:<br>[calc_stk_4] = '0' or [calc_gwtg_arrive2_treat3] = '0' | Section Header: *Fallout: Thrombolytic TherapySITE: [site] MRN: [mrn]Patient Name: [jc_lastname], [jc_firstname]Form / PMT ID:[patient_os_id]* | descriptive |
| 892 | fallout_detail_stk_4<br><br>Show the field ONLY if:<br>[calc_stk_4] = '0' or [calc_gwtg_arrive2_treat3] = '0' | Fallout Details: Thrombolytic Therapy | notes |
| 893 | fallout_loc_stk_4<br><br>Show the field ONLY if:<br>[calc_stk_4] = '0' or [calc_gwtg_arrive2_treat3] = '0' | Fallout Location | checkbox<br>1 \| fallout_loc_stk_4___1 \| ED<br>2 \| fallout_loc_stk_4___2 \| ICU<br>3 \| fallout_loc_stk_4___3 \| Floor<br><br>Custom alignment: RH |

| | | | |
|---|---|---|---|
| 894 | fallout_unit_stk_4<br><br>Show the field ONLY if:<br>[calc_stk_4] = '0' or [calc_gwtg_arrive2_treat3] = '0' | Fallout Unit | dropdown<br>1 ED<br>2 ICU<br>8 IMC<br>9 SICU<br>10 MICU<br>3 3rd Floor<br>4 4th Floor<br>11 4 East<br>12 4 West<br>5 5th Floor<br>6 6th Floor<br>7 7th Floor<br><br>Custom alignment: RH |
| 895 | fallout_rnmd_stk_4<br><br>Show the field ONLY if:<br>[calc_stk_4] = '0' or [calc_gwtg_arrive2_treat3] = '0' | Fallout by RN or MD? | checkbox<br>1 fallout_rnmd_stk_4___1 RN<br>2 fallout_rnmd_stk_4___2 MD<br><br>Custom alignment: RH |
| 896 | fallout_notified_stk_4<br><br>Show the field ONLY if:<br>[calc_stk_4] = '0' or [calc_gwtg_arrive2_treat3] = '0' | Fallout Notification Sent To:<br>*Name or Email* | text<br>Custom alignment: RH |
| 897 | fallout_date_stk_4<br><br>Show the field ONLY if:<br>[calc_stk_4] = '0' or [calc_gwtg_arrive2_treat3] = '0' | Date Notified | text (date_mdy, Min: 2016-01-01, Max: 2099-12-31)<br>Custom alignment: RH |
| 898 | fallout_alert_stk_4<br><br>Show the field ONLY if:<br>[calc_stk_4] = '0' or [calc_gwtg_arrive2_treat3] = '0' | Email Notification: | radio<br>1 Send Email<br>2 Email(s) Sent<br><br>Custom alignment: RH |

| | | | |
|---|---|---|---|
| 899 | fallout_verify_stk_4<br><br>Show the field ONLY if:<br>[calc_stk_4] = '0' or [calc_gwtg_arrive2_treat3] = '0' | Reviewed by Stroke Coordinator: | checkbox<br>1 \| fallout_verify_stk_4___1 \| |
| 900 | fallout_resp_desc_stk_4<br><br>Show the field ONLY if:<br>[calc_stk_4] = '0' or [calc_gwtg_arrive2_treat3] = '0' | Fallout Review: [fallout_review_stk_4]<br>Fallout Response: [fallout_response_stk_4] | descriptive |
| 901 | desc_fallout_stk_5<br><br>Show the field ONLY if:<br>[calc_stk_5] = '0' or [calc_gwtg_early_antithrom] = '0' | Section Header: *Fallout: Antithrombotic Therapy By End of Hospital Day 2SITE: [site] MRN: [mrn]Patient Name:[jc_lastname], [jc_firstname]Form / PMT ID:[patient_os_id]* | descriptive |
| 902 | fallout_detail_stk_5<br><br>Show the field ONLY if:<br>[calc_stk_5] = '0' or [calc_gwtg_early_antithrom] = '0' | Fallout Details: Antithrombotic Therapy By End of Hospital Day 2 | notes |
| 903 | fallout_loc_stk_5<br><br>Show the field ONLY if:<br>[calc_stk_5] = '0' or [calc_gwtg_early_antithrom] = '0' | Fallout Location | checkbox<br>1 \| fallout_loc_stk_5___1 \| ED<br>2 \| fallout_loc_stk_5___2 \| ICU<br>3 \| fallout_loc_stk_5___3 \| Floor<br><br>Custom alignment: RH |

| | | | |
|---|---|---|---|
| 904 | fallout_unit_stk_5<br><br>Show the field ONLY if:<br>[calc_stk_5] = '0' or [calc_gwtg_early_antithrom] = '0' | Fallout Unit | dropdown<br>1 ED<br>2 ICU<br>8 IMC<br>9 SICU<br>10 MICU<br>3 3rd Floor<br>4 4th Floor<br>11 4 East<br>12 4 West<br>5 5th Floor<br>6 6th Floor<br>7 7th Floor<br><br>Custom alignment: RH |
| 905 | fallout_rnmd_stk_5<br><br>Show the field ONLY if:<br>[calc_stk_5] = '0' or [calc_gwtg_early_antithrom] = '0' | Fallout by RN or MD? | checkbox<br>1 fallout_rnmd_stk_5___1 RN<br>2 fallout_rnmd_stk_5___2 MD<br><br>Custom alignment: RH |
| 906 | fallout_notified_stk_5<br><br>Show the field ONLY if:<br>[calc_stk_5] = '0' or [calc_gwtg_early_antithrom] = '0' | Fallout Notification Sent To:<br>*Name or Email* | text<br>Custom alignment: RH |
| 907 | fallout_date_stk_5<br><br>Show the field ONLY if:<br>[calc_stk_5] = '0' or [calc_gwtg_early_antithrom] = '0' | Date Notified | text (date_mdy, Min: 2016-01-01, Max: 2099-12-31)<br>Custom alignment: RH |
| 908 | fallout_alert_stk_5<br><br>Show the field ONLY if:<br>[calc_stk_5] = '0' or [calc_gwtg_early_antithrom] = '0' | Email Notification: | radio<br>1 Send Email<br>2 Email(s) Sent<br><br>Custom alignment: RH |

| | | | |
|---|---|---|---|
| 909 | fallout_verify_stk_5<br><br>Show the field ONLY if:<br>[calc_stk_5] = '0' or [calc_gwtg_early_antithrom] = '0' | Reviewed by Stroke Coordinator: | checkbox<br>1 \| fallout_verify_stk_5___1 \| |
| 910 | fallout_resp_desc_stk_5<br><br>Show the field ONLY if:<br>[calc_stk_5] = '0' or [calc_gwtg_early_antithrom] = '0' | Fallout Review: [fallout_review_stk_5]<br>Fallout Response: [fallout_response_stk_5] | descriptive |
| 911 | desc_fallout_stk_6<br><br>Show the field ONLY if:<br>[calc_stk_6] = '0' or [calc_gwtg_statin_at_disch] = '0' | Section Header: *Fallout: Discharged on Statin MedicationSITE: [site] MRN: [mrn]Patient Name:[jc_lastname], [jc_firstname]Form / PMT ID:[patient_os_id]* | descriptive |
| 912 | fallout_detail_stk_6<br><br>Show the field ONLY if:<br>[calc_stk_6] = '0' or [calc_gwtg_statin_at_disch] = '0' | Fallout Details: Discharged on Statin Medication | notes |
| 913 | fallout_loc_stk_6<br><br>Show the field ONLY if:<br>[calc_stk_6] = '0' or [calc_gwtg_statin_at_disch] = '0' | Fallout Location | checkbox<br>1 \| fallout_loc_stk_6___1 \| ED<br>2 \| fallout_loc_stk_6___2 \| ICU<br>3 \| fallout_loc_stk_6___3 \| Floor<br><br>Custom alignment: RH |

| | | | |
|---|---|---|---|
| 914 | fallout_unit_stk_6<br><br>Show the field ONLY if:<br>[calc_stk_6] = '0' or [calc_gwtg_statin_at_disch] = '0' | Fallout Unit | dropdown<br>1 ED<br>2 ICU<br>8 IMC<br>9 SICU<br>10 MICU<br>3 3rd Floor<br>4 4th Floor<br>11 4 East<br>12 4 West<br>5 5th Floor<br>6 6th Floor<br>7 7th Floor<br><br>Custom alignment: RH |
| 915 | fallout_rnmd_stk_6<br><br>Show the field ONLY if:<br>[calc_stk_6] = '0' or [calc_gwtg_statin_at_disch] = '0' | Fallout by RN or MD? | checkbox<br>1 fallout_rnmd_stk_6___1 RN<br>2 fallout_rnmd_stk_6___2 MD<br><br>Custom alignment: RH |
| 916 | fallout_notified_stk_6<br><br>Show the field ONLY if:<br>[calc_stk_6] = '0' or [calc_gwtg_statin_at_disch] = '0' | Fallout Notification Sent To:<br>*Name or Email* | text<br>Custom alignment: RH |
| 917 | fallout_date_stk_6<br><br>Show the field ONLY if:<br>[calc_stk_6] = '0' or [calc_gwtg_statin_at_disch] = '0' | Date Notified | text (date_mdy, Min: 2016-01-01, Max: 2099-12-31)<br>Custom alignment: RH |
| 918 | fallout_alert_stk_6<br><br>Show the field ONLY if:<br>[calc_stk_6] = '0' or [calc_gwtg_statin_at_disch] = '0' | Email Notification: | radio<br>1 Send Email<br>2 Email(s) Sent<br><br>Custom alignment: RH |

| | | | |
|---|---|---|---|
| 919 | fallout_verify_stk_6<br><br>Show the field ONLY if:<br>[calc_stk_6] = '0' or [calc_gwtg_statin_at_disch] = '0' | Reviewed by Stroke Coordinator: | checkbox<br>1 \| fallout_verify_stk_6___1 \| |
| 920 | fallout_resp_desc_stk_6<br><br>Show the field ONLY if:<br>[calc_stk_6] = '0' or [calc_gwtg_statin_at_disch] = '0' | Fallout Review: [fallout_review_stk_6]<br>Fallout Response: [fallout_response_stk_6] | descriptive |
| 921 | desc_fallout_stk_8<br><br>Show the field ONLY if:<br>[calc_stk_8] = '0' or [calc_gwtg_stroke_education] = '0' | Section Header: *Fallout: Stroke EducationSITE: [site] MRN: [mrn]Patient Name: [jc_lastname], [jc_firstname]Form / PMT ID:[patient_os_id]* | descriptive |
| 922 | fallout_detail_stk_8<br><br>Show the field ONLY if:<br>[calc_stk_8] = '0' or [calc_gwtg_stroke_education] = '0' | Fallout Details: Stroke Education | notes |
| 923 | fallout_loc_stk_8<br><br>Show the field ONLY if:<br>[calc_stk_8] = '0' or [calc_gwtg_stroke_education] = '0' | Fallout Location | checkbox<br>1 \| fallout_loc_stk_8___1 \| ED<br>2 \| fallout_loc_stk_8___2 \| ICU<br>3 \| fallout_loc_stk_8___3 \| Floor<br><br>Custom alignment: RH |

| | | | |
|---|---|---|---|
| 924 | fallout_unit_stk_8<br><br>Show the field ONLY if:<br>[calc_stk_8] = '0' or [calc_gwtg_stroke_education] = '0' | Fallout Unit | dropdown<br>1 ED<br>2 ICU<br>8 IMC<br>9 SICU<br>10 MICU<br>3 3rd Floor<br>4 4th Floor<br>11 4 East<br>12 4 West<br>5 5th Floor<br>6 6th Floor<br>7 7th Floor<br><br>Custom alignment: RH |
| 925 | fallout_rnmd_stk_8<br><br>Show the field ONLY if:<br>[calc_stk_8] = '0' or [calc_gwtg_stroke_education] = '0' | Fallout by RN or MD? | checkbox<br>1 fallout_rnmd_stk_8___1 RN<br>2 fallout_rnmd_stk_8___2 MD<br><br>Custom alignment: RH |
| 926 | fallout_notified_stk_8<br><br>Show the field ONLY if:<br>[calc_stk_8] = '0' or [calc_gwtg_stroke_education] = '0' | Fallout Notification Sent To:<br>*Name or Email* | text<br>Custom alignment: RH |
| 927 | fallout_date_stk_8<br><br>Show the field ONLY if:<br>[calc_stk_8] = '0' or [calc_gwtg_stroke_education] = '0' | Date Notified | text (date_mdy, Min: 2016-01-01, Max: 2099-12-31)<br>Custom alignment: RH |
| 928 | fallout_alert_stk_8<br><br>Show the field ONLY if:<br>[calc_stk_8] = '0' or [calc_gwtg_stroke_education] = '0' | Email Notification: | radio<br>1 Send Email<br>2 Email(s) Sent<br><br>Custom alignment: RH |

| | | | |
|---|---|---|---|
| 929 | fallout_verify_stk_8<br><br>Show the field ONLY if:<br>[calc_stk_8] = '0' or [calc_gwtg_stroke_education] = '0' | Reviewed by Stroke Coordinator: | checkbox<br>1 \| fallout_verify_stk_8___1 \| |
| 930 | fallout_resp_desc_stk_8<br><br>Show the field ONLY if:<br>[calc_stk_8] = '0' or [calc_gwtg_stroke_education] = '0' | Fallout Review: [fallout_review_stk_8]<br>Fallout Response: [fallout_response_stk_8] | descriptive |
| 931 | desc_fallout_stk_10<br><br>Show the field ONLY if:<br>[calc_stk_10] = '0' or [calc_gwtg_rehab_considered] = '0' | Section Header: *Fallout: Assessed for RehabilitationSITE: [site] MRN: [mrn]Patient Name: [jc_lastname], [jc_firstname]Form / PMT ID:[patient_os_id]* | descriptive |
| 932 | fallout_detail_stk_10<br><br>Show the field ONLY if:<br>[calc_stk_10] = '0' or [calc_gwtg_rehab_considered] = '0' | Fallout Details: Assessed for Rehabilitation | notes |
| 933 | fallout_loc_stk_10<br><br>Show the field ONLY if:<br>[calc_stk_10] = '0' or [calc_gwtg_rehab_considered] = '0' | Fallout Location | checkbox<br>1 \| fallout_loc_stk_10___1 \| ED<br>2 \| fallout_loc_stk_10___2 \| ICU<br>3 \| fallout_loc_stk_10___3 \| Floor<br><br>Custom alignment: RH |

| | | | |
|---|---|---|---|
| 934 | fallout_unit_stk_10<br><br>Show the field ONLY if:<br>[calc_stk_10] = '0' or [calc_gwtg_rehab_considered] = '0' | Fallout Unit | dropdown<br><table><tr><td>1</td><td>ED</td></tr><tr><td>2</td><td>ICU</td></tr><tr><td>8</td><td>IMC</td></tr><tr><td>9</td><td>SICU</td></tr><tr><td>10</td><td>MICU</td></tr><tr><td>3</td><td>3rd Floor</td></tr><tr><td>4</td><td>4th Floor</td></tr><tr><td>11</td><td>4 East</td></tr><tr><td>12</td><td>4 West</td></tr><tr><td>5</td><td>5th Floor</td></tr><tr><td>6</td><td>6th Floor</td></tr><tr><td>7</td><td>7th Floor</td></tr></table><br>Custom alignment: RH |
| 935 | fallout_rnmd_stk_10<br><br>Show the field ONLY if:<br>[calc_stk_10] = '0' or [calc_gwtg_rehab_considered] = '0' | Fallout by RN or MD? | checkbox<br><table><tr><td>1</td><td>fallout_rnmd_stk_10___1</td><td>RN</td></tr><tr><td>2</td><td>fallout_rnmd_stk_10___2</td><td>MD</td></tr></table><br>Custom alignment: RH |
| 936 | fallout_notified_stk_10<br><br>Show the field ONLY if:<br>[calc_stk_10] = '0' or [calc_gwtg_rehab_considered] = '0' | Fallout Notification Sent To:<br>*Name or Email* | text<br>Custom alignment: RH |
| 937 | fallout_date_stk_10<br><br>Show the field ONLY if:<br>[calc_stk_10] = '0' or [calc_gwtg_rehab_considered] = '0' | Date Notified | text (date_mdy, Min: 2016-01-01, Max: 2099-12-31)<br>Custom alignment: RH |
| 938 | fallout_alert_stk_10<br><br>Show the field ONLY if:<br>[calc_stk_10] = '0' or [calc_gwtg_rehab_considered] = '0' | Email Notification: | radio<br><table><tr><td>1</td><td>Send Email</td></tr><tr><td>2</td><td>Email(s) Sent</td></tr></table><br>Custom alignment: RH |

| | | | |
|---|---|---|---|
| 939 | fallout_verify_stk_10<br><br>Show the field ONLY if:<br>[calc_stk_10] = '0' or [calc_gwtg_rehab_considered] = '0' | Reviewed by Stroke Coordinator: | checkbox<br>1 \| fallout_verify_stk_10___1 \| |
| 940 | fallout_resp_desc_stk_10<br><br>Show the field ONLY if:<br>[calc_stk_10] = '0' or [calc_gwtg_rehab_considered] = '0' | Fallout Review: [fallout_review_stk_10]<br>Fallout Response: [fallout_response_stk_10] | descriptive |
| 941 | desc_fallout_swallow<br><br>Show the field ONLY if:<br>[calc_gwtg_dysphagia_screen] = '0' or [calc_swallow_screen] = '0' | Section Header: *Fallout: Swallow ScreenSITE: [site] MRN: [mrn]Patient Name:[jc_lastname], [jc_firstname]Form / PMT ID:[patient_os_id]* | descriptive |
| 942 | fallout_detail_swallow<br><br>Show the field ONLY if:<br>[calc_gwtg_dysphagia_screen] = '0' or [calc_swallow_screen] = '0' | Fallout Details: Swallow Screen | notes |
| 943 | fallout_loc_swallow<br><br>Show the field ONLY if:<br>[calc_gwtg_dysphagia_screen] = '0' or [calc_swallow_screen] = '0' | Fallout Location | checkbox<br>1 \| fallout_loc_swallow___1 \| ED<br>2 \| fallout_loc_swallow___2 \| ICU<br>3 \| fallout_loc_swallow___3 \| Floor<br><br>Custom alignment: RH |

| | | | |
|---|---|---|---|
| 944 | fallout_unit_swallow<br><br>Show the field ONLY if:<br>[calc_gwtg_dysphagia_screen] = '0' or [calc_swallow_screen] = '0' | Fallout Unit | dropdown<br>1 ED<br>2 ICU<br>8 IMC<br>9 SICU<br>10 MICU<br>3 3rd Floor<br>4 4th Floor<br>11 4 East<br>12 4 West<br>5 5th Floor<br>6 6th Floor<br>7 7th Floor<br><br>Custom alignment: RH |
| 945 | fallout_rnmd_swallow<br><br>Show the field ONLY if:<br>[calc_gwtg_dysphagia_screen] = '0' or [calc_swallow_screen] = '0' | Fallout by RN or MD? | checkbox<br>1 fallout_rnmd_swallow___1 RN<br>2 fallout_rnmd_swallow___2 MD<br><br>Custom alignment: RH |
| 946 | fallout_notified_swallow<br><br>Show the field ONLY if:<br>[calc_gwtg_dysphagia_screen] = '0' or [calc_swallow_screen] = '0' | Fallout Notification Sent To:<br>*Name or Email* | text<br>Custom alignment: RH |
| 947 | fallout_date_swallow<br><br>Show the field ONLY if:<br>[calc_gwtg_dysphagia_screen] = '0' or [calc_swallow_screen] = '0' | Date Notified | text (date_mdy, Min: 2016-01-01, Max: 2099-12-31)<br>Custom alignment: RH |
| 948 | fallout_alert_swallow<br><br>Show the field ONLY if:<br>[calc_gwtg_dysphagia_screen] = '0' or [calc_swallow_screen] = '0' | Email Notification: | radio<br>1 Send Email<br>2 Email(s) Sent<br><br>Custom alignment: RH |

| | | | |
|---|---|---|---|
| 949 | fallout_verify_swallow<br><br>Show the field ONLY if:<br>[calc_gwtg_dysphagia_screen] = '0' or [calc_swallow_screen] = '0' | Reviewed by Stroke Coordinator: | checkbox<br>1 \| fallout_verify_swallow___1 \| |
| 950 | fallout_resp_desc_swallow<br><br>Show the field ONLY if:<br>[calc_gwtg_dysphagia_screen] = '0' or [calc_swallow_screen] = '0' | Fallout Review: [fallout_review_swallow]<br>Fallout Response: [fallout_response_swallow] | descriptive |
| 951 | desc_fallout_nihss<br><br>Show the field ONLY if:<br>[calc_gwtg_nihss_reported] = '0' or [calc_dnv_1] = '0' | Section Header: *Fallout: NIHSS ReportedSITE: [site] MRN: [mrn]Patient Name:[jc_lastname], [jc_firstname]Form / PMT ID:[patient_os_id]* | descriptive |
| 952 | fallout_detail_nihss<br><br>Show the field ONLY if:<br>[calc_gwtg_nihss_reported] = '0' or [calc_dnv_1] = '0' | Fallout Details: NIHSS Reported | notes |
| 953 | fallout_loc_nihss<br><br>Show the field ONLY if:<br>[calc_gwtg_nihss_reported] = '0' or [calc_dnv_1] = '0' | Fallout Location | checkbox<br>1 \| fallout_loc_nihss___1 \| ED<br>2 \| fallout_loc_nihss___2 \| ICU<br>3 \| fallout_loc_nihss___3 \| Floor<br><br>Custom alignment: RH |

| | | | |
|---|---|---|---|
| 954 | fallout_unit_nihss<br><br>Show the field ONLY if:<br>[calc_gwtg_nihss_reported] = '0' or [calc_dnv_1] = '0' | Fallout Unit | dropdown<br>1 ED<br>2 ICU<br>8 IMC<br>9 SICU<br>10 MICU<br>3 3rd Floor<br>4 4th Floor<br>11 4 East<br>12 4 West<br>5 5th Floor<br>6 6th Floor<br>7 7th Floor<br><br>Custom alignment: RH |
| 955 | fallout_rnmd_nihss<br><br>Show the field ONLY if:<br>[calc_gwtg_nihss_reported] = '0' or [calc_dnv_1] = '0' | Fallout by RN or MD? | checkbox<br>1 fallout_rnmd_nihss___1 RN<br>2 fallout_rnmd_nihss___2 MD<br><br>Custom alignment: RH |
| 956 | fallout_notified_nihss<br><br>Show the field ONLY if:<br>[calc_gwtg_nihss_reported] = '0' or [calc_dnv_1] = '0' | Fallout Notification Sent To:<br>*Name or Email* | text<br>Custom alignment: RH |
| 957 | fallout_date_nihss<br><br>Show the field ONLY if:<br>[calc_gwtg_nihss_reported] = '0' or [calc_dnv_1] = '0' | Date Notified | text (date_mdy, Min: 2016-01-01, Max: 2099-12-31)<br>Custom alignment: RH |
| 958 | fallout_alert_nihss<br><br>Show the field ONLY if:<br>[calc_gwtg_nihss_reported] = '0' or [calc_dnv_1] = '0' | Email Notification: | radio<br>1 Send Email<br>2 Email(s) Sent<br><br>Custom alignment: RH |

| | | | |
|---|---|---|---|
| 959 | fallout_verify_nihss<br><br>Show the field ONLY if:<br>[calc_gwtg_nihss_reported] = '0' or [calc_dnv_1] = '0' | Reviewed by Stroke Coordinator: | checkbox<br>1 \| fallout_verify_nihss___1 \| |
| 960 | fallout_resp_desc_nihss<br><br>Show the field ONLY if:<br>[calc_gwtg_nihss_reported] = '0' or [calc_dnv_1] = '0' | Fallout Review: [fallout_review_nihss]<br>Fallout Response: [fallout_response_nihss] | descriptive |
| 961 | desc_fallout_mrs<br><br>Show the field ONLY if:<br>[calc_dnv_qm7_cr4a]='0' or [calc_dnv_qm7_cr4b]='0' or [calc_dnv_qm7_cr4c]='0' or [calc_dnv_9]='0' | Section Header: *Fallout: Modified Rankin Scores DocumentedSITE: [site]MRN: [mrn]Patient Name:[jc_lastname], [jc_firstname]GWTG PMT ID:[patient_os_id]QM.7 CR.4a - Pre-Morbid mRS:[calc_dnv_qm7_cr4a]1 = In Numerator 0 = In Measure / Fallout Blank = Not in MeasureQM.7 CR.4b - Discharge mRS:[calc_dnv_qm7_cr4b]QM.7 CR.4c - 90 Day mRS:[calc_dnv_qm7_cr4c]DNV 9:[calc_dnv_9]* | descriptive |
| 962 | fallout_detail_mrs<br><br>Show the field ONLY if:<br>[calc_dnv_qm7_cr4a]='0' or [calc_dnv_qm7_cr4b]='0' or [calc_dnv_qm7_cr4c]='0' or [calc_dnv_9]='0' | Fallout Details: Modified Rankin Scores Documented | notes |
| 963 | fallout_loc_mrs<br><br>Show the field ONLY if:<br>[calc_dnv_qm7_cr4a]='0' or [calc_dnv_qm7_cr4b]='0' or [calc_dnv_qm7_cr4c]='0' or [calc_dnv_9]='0' | Fallout Location | checkbox<br>1 \| fallout_loc_mrs___1 \| ED<br>2 \| fallout_loc_mrs___2 \| ICU<br>3 \| fallout_loc_mrs___3 \| Floor<br><br>Custom alignment: RH |

| | | | |
|---|---|---|---|
| 964 | fallout_unit_mrs<br><br>Show the field ONLY if:<br>[calc_dnv_qm7_cr4a]='0' or [calc_dnv_qm7_cr4b]='0' or [calc_dnv_qm7_cr4c]='0' or [calc_dnv_9]='0' | Fallout Unit | dropdown<br>1 ED<br>2 ICU<br>8 IMC<br>9 SICU<br>10 MICU<br>3 3rd Floor<br>4 4th Floor<br>11 4 East<br>12 4 West<br>5 5th Floor<br>6 6th Floor<br>7 7th Floor<br><br>Custom alignment: RH |
| 965 | fallout_rnmd_mrs<br><br>Show the field ONLY if:<br>[calc_dnv_qm7_cr4a]='0' or [calc_dnv_qm7_cr4b]='0' or [calc_dnv_qm7_cr4c]='0' or [calc_dnv_9]='0' | Fallout by? | checkbox<br>1 fallout_rnmd_mrs___1 RN<br>2 fallout_rnmd_mrs___2 MD<br>3 fallout_rnmd_mrs___3 PT/OT<br><br>Custom alignment: RH |
| 966 | fallout_notified_mrs<br><br>Show the field ONLY if:<br>[calc_dnv_qm7_cr4a]='0' or [calc_dnv_qm7_cr4b]='0' or [calc_dnv_qm7_cr4c]='0' or [calc_dnv_9]='0' | Fallout Notification Sent To:<br>*Name or Email* | text<br>Custom alignment: RH |
| 967 | fallout_date_mrs<br><br>Show the field ONLY if:<br>[calc_dnv_qm7_cr4a]='0' or [calc_dnv_qm7_cr4b]='0' or [calc_dnv_qm7_cr4c]='0' or [calc_dnv_9]='0' | Date Notified | text (date_mdy, Min: 2016-01-01, Max: 2099-12-31)<br>Custom alignment: RH |
| 968 | fallout_alert_mrs<br><br>Show the field ONLY if:<br>[calc_dnv_qm7_cr4a]='0' or [calc_dnv_qm7_cr4b]='0' or [calc_dnv_qm7_cr4c]='0' or [calc_dnv_9]='0' | Email Notification: | radio<br>1 Send Email<br>2 Email(s) Sent<br><br>Custom alignment: RH |

| | | | |
|---|---|---|---|
| 969 | fallout_verify_mrs<br><br>Show the field ONLY if:<br>[calc_dnv_qm7_cr4a]='0' or [calc_dnv_qm7_cr4b]='0' or [calc_dnv_qm7_cr4c]='0' or [calc_dnv_9]='0' | Reviewed by Stroke Coordinator: | checkbox<br>1 \| fallout_verify_mrs___1 \| |
| 970 | fallout_resp_desc_mrs<br><br>Show the field ONLY if:<br>[calc_dnv_qm7_cr4a]='0' or [calc_dnv_qm7_cr4b]='0' or [calc_dnv_qm7_cr4c]='0' or [calc_dnv_9]='0' | Fallout Review: [fallout_review_mrs]<br>Fallout Response: [fallout_response_mrs] | descriptive |
| 971 | desc_fallout_sevscore<br><br>Show the field ONLY if:<br>[calc_dnv_12]='0' | Section Header: *Fallout: Initial Severity Scores DocumentedSITE: [site]MRN: [mrn]Patient Name:[jc_lastname], [jc_firstname]Form / PMT ID:[patient_os_id]* | descriptive |
| 972 | fallout_detail_sevscore<br><br>Show the field ONLY if:<br>[calc_dnv_12]='0' | Fallout Details: Initial Severity Scores Documented | notes |
| 973 | fallout_loc_sevscore<br><br>Show the field ONLY if:<br>[calc_dnv_12]='0' | Fallout Location | checkbox<br>1 \| fallout_loc_sevscore___1 \| ED<br>2 \| fallout_loc_sevscore___2 \| ICU<br>3 \| fallout_loc_sevscore___3 \| Floor<br><br>Custom alignment: RH |

| | | | |
|---|---|---|---|
| 974 | fallout_unit_sevscore<br><br>Show the field ONLY if:<br>[calc_dnv_12]='0' | Fallout Unit | dropdown<br>1 ED<br>2 ICU<br>8 IMC<br>9 SICU<br>10 MICU<br>3 3rd Floor<br>4 4th Floor<br>11 4 East<br>12 4 West<br>5 5th Floor<br>6 6th Floor<br>7 7th Floor<br><br>Custom alignment: RH |
| 975 | fallout_rnmd_sevscore<br><br>Show the field ONLY if:<br>[calc_dnv_12]='0' | Fallout by RN or MD? | checkbox<br>1 fallout_rnmd_sevscore___1 RN<br>2 fallout_rnmd_sevscore___2 MD<br><br>Custom alignment: RH |
| 976 | fallout_notified_sevscore<br><br>Show the field ONLY if:<br>[calc_dnv_12]='0' | Fallout Notification Sent To:<br>*Name or Email* | text<br>Custom alignment: RH |
| 977 | fallout_date_sevscore<br><br>Show the field ONLY if:<br>[calc_dnv_12]='0' | Date Notified | text (date_mdy, Min: 2016-01-01, Max: 2099-12-31)<br>Custom alignment: RH |
| 978 | fallout_alert_sevscore<br><br>Show the field ONLY if:<br>[calc_dnv_12]='0' | Email Notification: | radio<br>1 Send Email<br>2 Email(s) Sent<br><br>Custom alignment: RH |
| 979 | fallout_verify_sevscore<br><br>Show the field ONLY if:<br>[calc_dnv_12]='0' | Reviewed by Stroke Coordinator: | checkbox<br>1 fallout_verify_sevscore___1 |

| | | | |
|---|---|---|---|
| 980 | fallout_resp_desc_sevscore<br><br>Show the field ONLY if:<br>[calc_dnv_12]='0' | Fallout Review: [fallout_review_sevscore]<br>Fallout Response: [fallout_response_sevscore] | descriptive |
| 981 | desc_fallout_fib_edu<br><br>Show the field ONLY if:<br>[calc_fib_edu]='0' | Section Header: *Fallout: Fibrinolytic Therapy EducationSITE: [site] MRN: [mrn]Patient Name:[jc_lastname], [jc_firstname]Form / PMT ID:[patient_os_id]* | descriptive |
| 982 | fallout_detail_fib_edu<br><br>Show the field ONLY if:<br>[calc_fib_edu]='0' | Fallout Details: Fibrinolytic Therapy Education | notes |
| 983 | fallout_loc_fib_edu<br><br>Show the field ONLY if:<br>[calc_fib_edu]='0' | Fallout Location | checkbox<br>1 fallout_loc_fib_edu___1 ED<br>2 fallout_loc_fib_edu___2 ICU<br>3 fallout_loc_fib_edu___3 Floor<br><br>Custom alignment: RH |
| 984 | fallout_unit_fib_edu<br><br>Show the field ONLY if:<br>[calc_fib_edu]='0' | Fallout Unit | dropdown<br>1 ED<br>2 ICU<br>8 IMC<br>9 SICU<br>10 MICU<br>3 3rd Floor<br>4 4th Floor<br>11 4 East<br>12 4 West<br>5 5th Floor<br>6 6th Floor<br>7 7th Floor<br><br>Custom alignment: RH |
| 985 | fallout_rnmd_fib_edu<br><br>Show the field ONLY if:<br>[calc_fib_edu]='0' | Fallout by RN or MD? | checkbox<br>1 fallout_rnmd_fib_edu___1 RN<br>2 fallout_rnmd_fib_edu___2 MD<br><br>Custom alignment: RH |

| | | | |
|---|---|---|---|
| 986 | fallout_notified_fib_edu<br><br>Show the field ONLY if:<br>[calc_fib_edu]='0' | Fallout Notification Sent To:<br>*Name or Email* | text<br>Custom alignment: RH |
| 987 | fallout_date_fib_edu<br><br>Show the field ONLY if:<br>[calc_fib_edu]='0' | Date Notified | text (date_mdy, Min: 2016-01-01, Max: 2099-12-31)<br>Custom alignment: RH |
| 988 | fallout_alert_fib_edu<br><br>Show the field ONLY if:<br>[calc_fib_edu]='0' | Email Notification: | radio<br>1 \| Send Email<br>2 \| Email(s) Sent<br><br>Custom alignment: RH |
| 989 | fallout_verify_fib_edu<br><br>Show the field ONLY if:<br>[calc_fib_edu]='0' | Reviewed by Stroke Coordinator: | checkbox<br>1 \| fallout_verify_fib_edu___1 \| |
| 990 | fallout_resp_desc_fib_edu<br><br>Show the field ONLY if:<br>[calc_fib_edu]='0' | Fallout Review: [fallout_review_fib_edu]<br>Fallout Response: [fallout_response_fib_edu] | descriptive |
| 991 | desc_fallout_thro_edu<br><br>Show the field ONLY if:<br>[calc_thro_edu]='0' | Section Header: *Fallout: Arteriogram EducationSITE: [site] MRN: [mrn]Patient Name: [jc_lastname], [jc_firstname]Form / PMT ID:[patient_os_id]* | descriptive |
| 992 | fallout_detail_thro_edu<br><br>Show the field ONLY if:<br>[calc_thro_edu]='0' | Fallout Details: Arteriogram Education | notes |
| 993 | fallout_loc_thro_edu<br><br>Show the field ONLY if:<br>[calc_thro_edu]='0' | Fallout Location | checkbox<br>1 \| fallout_loc_thro_edu___1 \| ED<br>2 \| fallout_loc_thro_edu___2 \| ICU<br>3 \| fallout_loc_thro_edu___3 \| Floor<br><br>Custom alignment: RH |

| | | | |
|---|---|---|---|
| 994 | fallout_unit_thro_edu<br><br>Show the field ONLY if:<br>[calc_thro_edu]='0' | Fallout Unit | dropdown<br>1 ED<br>2 ICU<br>8 IMC<br>9 SICU<br>10 MICU<br>3 3rd Floor<br>4 4th Floor<br>11 4 East<br>12 4 West<br>5 5th Floor<br>6 6th Floor<br>7 7th Floor<br><br>Custom alignment: RH |
| 995 | fallout_rnmd_thro_edu<br><br>Show the field ONLY if:<br>[calc_thro_edu]='0' | Fallout by RN or MD? | checkbox<br>1 fallout_rnmd_thro_edu___1 RN<br>2 fallout_rnmd_thro_edu___2 MD<br><br>Custom alignment: RH |
| 996 | fallout_notified_thro_edu<br><br>Show the field ONLY if:<br>[calc_thro_edu]='0' | Fallout Notification Sent To:<br>*Name or Email* | text<br>Custom alignment: RH |
| 997 | fallout_date_thro_edu<br><br>Show the field ONLY if:<br>[calc_thro_edu]='0' | Date Notified | text (date_mdy, Min: 2016-01-01, Max: 2099-12-31)<br>Custom alignment: RH |
| 998 | fallout_alert_thro_edu<br><br>Show the field ONLY if:<br>[calc_thro_edu]='0' | Email Notification: | radio<br>1 Send Email<br>2 Email(s) Sent<br><br>Custom alignment: RH |
| 999 | fallout_verify_thro_edu<br><br>Show the field ONLY if:<br>[calc_thro_edu]='0' | Reviewed by Stroke Coordinator: | checkbox<br>1 fallout_verify_thro_edu___1 |

| | | | |
|---|---|---|---|
| 1000 | fallout_resp_desc_thro_edu<br><br>Show the field ONLY if:<br>[calc_thro_edu]='0' | Fallout Review: [fallout_review_thro_edu]<br>Fallout Response: [fallout_response_thro_edu] | descriptive |
| 1001 | desc_fallout_smoking<br><br>Show the field ONLY if:<br>[calc_gwtg_smoking_cess]='0' | Section Header: *Fallout: Smoking CessationSITE: [site] MRN: [mrn]Patient Name: [jc_lastname], [jc_firstname]Form / PMT ID:[patient_os_id]* | descriptive |
| 1002 | fallout_detail_smoking<br><br>Show the field ONLY if:<br>[calc_gwtg_smoking_cess]='0' | Fallout Details: Smoking Cessation | notes |
| 1003 | fallout_loc_smoking<br><br>Show the field ONLY if:<br>[calc_gwtg_smoking_cess]='0' | Fallout Location | checkbox<br>1 \| fallout_loc_smoking___1 \| ED<br>2 \| fallout_loc_smoking___2 \| ICU<br>3 \| fallout_loc_smoking___3 \| Floor<br><br>Custom alignment: RH |
| 1004 | fallout_unit_smoking<br><br>Show the field ONLY if:<br>[calc_gwtg_smoking_cess]='0' | Fallout Unit | dropdown<br>1 \| ED<br>2 \| ICU<br>8 \| IMC<br>9 \| SICU<br>10 \| MICU<br>3 \| 3rd Floor<br>4 \| 4th Floor<br>11 \| 4 East<br>12 \| 4 West<br>5 \| 5th Floor<br>6 \| 6th Floor<br>7 \| 7th Floor<br><br>Custom alignment: RH |
| 1005 | fallout_rnmd_smoking<br><br>Show the field ONLY if:<br>[calc_gwtg_smoking_cess]='0' | Fallout by RN or MD? | checkbox<br>1 \| fallout_rnmd_smoking___1 \| RN<br>2 \| fallout_rnmd_smoking___2 \| MD<br><br>Custom alignment: RH |

| | | | |
|---|---|---|---|
| 1006 | fallout_notified_smoking<br><br>Show the field ONLY if:<br>[calc_gwtg_smoking_cess]='0' | Fallout Notification Sent To:<br>*Name or Email* | text<br>Custom alignment: RH |
| 1007 | fallout_date_smoking<br><br>Show the field ONLY if:<br>[calc_gwtg_smoking_cess]='0' | Date Notified | text (date_mdy, Min: 2016-01-01, Max: 2099-12-31)<br>Custom alignment: RH |
| 1008 | fallout_alert_smoking<br><br>Show the field ONLY if:<br>[calc_gwtg_smoking_cess]='0' | Email Notification: | radio<br>1 \| Send Email<br>2 \| Email(s) Sent<br><br>Custom alignment: RH |
| 1009 | fallout_verify_smoking<br><br>Show the field ONLY if:<br>[calc_gwtg_smoking_cess]='0' | Reviewed by Stroke Coordinator: | checkbox<br>1 \| fallout_verify_smoking___1 \| |
| 1010 | fallout_resp_desc_smoking<br><br>Show the field ONLY if:<br>[calc_gwtg_smoking_cess]='0' | Fallout Review: [fallout_review_smoking]<br>Fallout Response: [fallout_response_smoking] | descriptive |
| 1011 | desc_fallout_ldl<br><br>Show the field ONLY if:<br>[calc_gwtg_ldl_documented]='0' | Section Header: *Fallout: LDL DocumentedSITE: [site] MRN: [mrn]Patient Name: [jc_lastname], [jc_firstname]Form / PMT ID:[patient_os_id]* | descriptive |
| 1012 | fallout_detail_ldl<br><br>Show the field ONLY if:<br>[calc_gwtg_ldl_documented]='0' | Fallout Details: LDL Documented | notes |
| 1013 | fallout_loc_ldl<br><br>Show the field ONLY if:<br>[calc_gwtg_ldl_documented]='0' | Fallout Location | checkbox<br>1 \| fallout_loc_ldl___1 \| ED<br>2 \| fallout_loc_ldl___2 \| ICU<br>3 \| fallout_loc_ldl___3 \| Floor<br><br>Custom alignment: RH |

| | | | |
|---|---|---|---|
| 1014 | fallout_unit_ldl<br><br>Show the field ONLY if:<br>[calc_gwtg_ldl_documented]='0' | Fallout Unit | dropdown<br>1 ED<br>2 ICU<br>8 IMC<br>9 SICU<br>10 MICU<br>3 3rd Floor<br>4 4th Floor<br>11 4 East<br>12 4 West<br>5 5th Floor<br>6 6th Floor<br>7 7th Floor<br><br>Custom alignment: RH |
| 1015 | fallout_rnmd_ldl<br><br>Show the field ONLY if:<br>[calc_gwtg_ldl_documented]='0' | Fallout by RN or MD? | checkbox<br>1 fallout_rnmd_ldl___1 RN<br>2 fallout_rnmd_ldl___2 MD<br><br>Custom alignment: RH |
| 1016 | fallout_notified_ldl<br><br>Show the field ONLY if:<br>[calc_gwtg_ldl_documented]='0' | Fallout Notification Sent To:<br>*Name or Email* | text<br>Custom alignment: RH |
| 1017 | fallout_date_ldl<br><br>Show the field ONLY if:<br>[calc_gwtg_ldl_documented]='0' | Date Notified | text (date_mdy, Min: 2016-01-01, Max: 2099-12-31)<br>Custom alignment: RH |
| 1018 | fallout_alert_ldl<br><br>Show the field ONLY if:<br>[calc_gwtg_ldl_documented]='0' | Email Notification: | radio<br>1 Send Email<br>2 Email(s) Sent<br><br>Custom alignment: RH |
| 1019 | fallout_verify_ldl<br><br>Show the field ONLY if:<br>[calc_gwtg_ldl_documented]='0' | Reviewed by Stroke Coordinator: | checkbox<br>1 fallout_verify_ldl___1 |

| | | | |
|---|---|---|---|
| 1020 | fallout_resp_desc_ldl<br><br>Show the field ONLY if:<br>[calc_gwtg_ldl_documented]='0' | Fallout Review: [fallout_review_ldl]<br>Fallout Response: [fallout_response_ldl] | descriptive |
| 1021 | desc_fallout_ist<br><br>Show the field ONLY if:<br>[calc_gwtg_intensive_statin]='0' | Section Header: *Fallout: Intensive Statin TherapySITE: [site] MRN: [mrn]Patient Name: [jc_lastname], [jc_firstname]Form / PMT ID:[patient_os_id]* | descriptive |
| 1022 | fallout_detail_ist<br><br>Show the field ONLY if:<br>[calc_gwtg_intensive_statin]='0' | Fallout Details: Intensive Statin Therapy | notes |
| 1023 | fallout_loc_ist<br><br>Show the field ONLY if:<br>[calc_gwtg_intensive_statin]='0' | Fallout Location | checkbox<br>1 \| fallout_loc_ist___1 \| ED<br>2 \| fallout_loc_ist___2 \| ICU<br>3 \| fallout_loc_ist___3 \| Floor<br><br>Custom alignment: RH |
| 1024 | fallout_unit_ist<br><br>Show the field ONLY if:<br>[calc_gwtg_intensive_statin]='0' | Fallout Unit | dropdown<br>1 \| ED<br>2 \| ICU<br>8 \| IMC<br>9 \| SICU<br>10 \| MICU<br>3 \| 3rd Floor<br>4 \| 4th Floor<br>11 \| 4 East<br>12 \| 4 West<br>5 \| 5th Floor<br>6 \| 6th Floor<br>7 \| 7th Floor<br><br>Custom alignment: RH |
| 1025 | fallout_rnmd_ist<br><br>Show the field ONLY if:<br>[calc_gwtg_intensive_statin]='0' | Fallout by RN or MD? | checkbox<br>1 \| fallout_rnmd_ist___1 \| RN<br>2 \| fallout_rnmd_ist___2 \| MD<br><br>Custom alignment: RH |

| | | | |
|---|---|---|---|
| 1026 | fallout_notified_ist<br><br>Show the field ONLY if:<br>[calc_gwtg_intensive_statin]='0' | Fallout Notification Sent To:<br>*Name or Email* | text<br>Custom alignment: RH |
| 1027 | fallout_date_ist<br><br>Show the field ONLY if:<br>[calc_gwtg_intensive_statin]='0' | Date Notified | text (date_mdy, Min: 2016-01-01, Max: 2099-12-31)<br>Custom alignment: RH |
| 1028 | fallout_alert_ist<br><br>Show the field ONLY if:<br>[calc_gwtg_intensive_statin]='0' | Email Notification: | radio<br>1 Send Email<br>2 Email(s) Sent<br><br>Custom alignment: RH |
| 1029 | fallout_verify_ist<br><br>Show the field ONLY if:<br>[calc_gwtg_intensive_statin]='0' | Reviewed by Stroke Coordinator: | checkbox<br>1 fallout_verify_ist___1 |
| 1030 | fallout_resp_desc_ist<br><br>Show the field ONLY if:<br>[calc_gwtg_intensive_statin]='0' | Fallout Review: [fallout_review_ist]<br>Fallout Response: [fallout_response_ist] | descriptive |
| 1031 | desc_fallout_cva_poc<br><br>Show the field ONLY if:<br>[calc_cva_poc_fallout] = '0' | Section Header: *Fallout: CVA Plan of CareSITE: [site] MRN: [mrn]Patient Name: [jc_lastname], [jc_firstname]Fallout Details:[cva_poc_notes] Was the CVA Plan of Care Initiated within 24 hours of admission?[cva_poc_init]Was the CVA Plan of Care signed and dated by all RNs?[cva_poc_signed]CVA Plan of Care Compliance (Reviewed and Updated Daily)[calc_cva_poc]Was Stroke Education on Risk Factors initiated on the CVA POC?[cva_poc_risk_fact_init]Were all appropriate patient specific risk factors selected for stroke education on the CVA POC?[cva_poc_risk_fact_ident]* | descriptive |
| 1032 | fallout_notified_cva_poc<br><br>Show the field ONLY if:<br>[calc_cva_poc_fallout] = '0' | Fallout Notification Sent To:<br>*Name or Email* | text<br>Custom alignment: RH |
| 1033 | fallout_date_cva_poc<br><br>Show the field ONLY if:<br>[calc_cva_poc_fallout] = '0' | Date Notified | text (date_mdy, Min: 2016-01-01, Max: 2099-12-31)<br>Custom alignment: RH |
| 1034 | fallout_alert_cva_poc<br><br>Show the field ONLY if:<br>[calc_cva_poc_fallout] = '0' | Email Notification: | radio<br>1 Send Email<br>2 Email(s) Sent<br><br>Custom alignment: RH |
| 1035 | fallout_verify_cva_poc<br><br>Show the field ONLY if:<br>[calc_cva_poc_fallout] = '0' | Reviewed by Stroke Coordinator: | checkbox<br>1 fallout_verify_cva_poc___1 |

| | | | |
|---|---|---|---|
| 1036 | desc_cva_poc_fo_ind_need<br><br>Show the field ONLY if:<br>[calc_cva_poc_fallout] = '0' and [cva_poc_sample]='1' | Question below will only be answered if selected for patient sample: Were individual needs identified for the patient based on their condition and co-morbidities on any POC?[cva_poc_indiv_needs_ident] | descriptive |
| 1037 | fallout_date_cva_poc_2<br><br>Show the field ONLY if:<br>[calc_cva_poc_fallout] = '0' and [cva_poc_indiv_needs_ident]='0' | Date Notified: Individual Needs Identified | text (date_mdy, Min: 2016-01-01, Max: 2099-12-31)<br>Custom alignment: RH |
| 1038 | fallout_email_cva_poc_1<br><br>Show the field ONLY if:<br>[calc_cva_poc_fallout] = '0' | Fallout Notification Email:<br>*Email #1* | text (email)<br>Custom alignment: RH<br>Field Annotation: @HIDDEN |
| 1039 | fallout_email_cva_poc_2<br><br>Show the field ONLY if:<br>[calc_cva_poc_fallout] = '0' and [fallout_email_cva_poc_1] <> '' | Fallout Notification Email:<br>*Email #2* | text (email)<br>Custom alignment: RH<br>Field Annotation: @HIDDEN |
| 1040 | fallout_email_cva_poc_3<br><br>Show the field ONLY if:<br>[calc_cva_poc_fallout] = '0' and [fallout_email_cva_poc_2] <> '' | Fallout Notification Email:<br>*Email #3* | text (email)<br>Custom alignment: RH<br>Field Annotation: @HIDDEN |
| 1041 | fallout_email_cva_poc_4<br><br>Show the field ONLY if:<br>[calc_cva_poc_fallout] = '0' and [fallout_email_cva_poc_3] <> '' | Fallout Notification Email:<br>*Email #4* | text (email)<br>Custom alignment: RH<br>Field Annotation: @HIDDEN |
| 1042 | fallout_email_cva_poc_5<br><br>Show the field ONLY if:<br>[calc_cva_poc_fallout] = '0' and [fallout_email_cva_poc_4] <> '' | Fallout Notification Email:<br>*Email #5* | text (email)<br>Custom alignment: RH<br>Field Annotation: @HIDDEN |
| 1043 | fallout_resp_desc_cva_poc<br><br>Show the field ONLY if:<br>[calc_cva_poc_fallout] = '0' | Fallout Review:[fallout_review_cva_poc]Fallout Response:<br>[fallout_response_cva_poc] | descriptive |
| 1044 | fallout_resp_desc_cva_poc_2<br><br>Show the field ONLY if:<br>[calc_cva_poc_fallout] = '0' and [cva_poc_indiv_needs_ident]='0' | Fallout Review: Individual Needs Identified[fallout_review_cva_poc_2]Fallout Response: Individual Needs Identified[fallout_response_cva_poc_2] | descriptive |

| | | | |
|---|---|---|---|
| 1045 | desc_fallout_post_tpa_bp<br><br>Show the field ONLY if:<br>[site]='SMCH' and [calc_post_tpa_monitoring]='0' | Section Header: *Fallout: Post IV Thrombolytic MonitoringSite: [site] MRN: [mrn]Patient Name:[jc_lastname], [jc_firstname]Fallout Details:[tpa_tool_notes] Neuro Vital Signs Numerator/Denominator:[tpa_tool_nvs_num]/[tpa_tool_nvs_den]Vital Signs Numerator/Denominator:[tpa_tool_vs_num]/[tpa_tool_vs_den]Clinician Notification Numerator/Denominator:[tpa_tool_md_num]/[tpa_tool_md_den]Post tPA Hand Off Numerator/Denominator:[tpa_tool_handoff_num]/[tpa_tool_handoff_den]Post IV tPA Monitoring Compliance[calc_post_tpa_compliance]%*<br><br>ANY BP > 180/105 during monitoring? (SMCH)[post_tpa_bp]ALL BPs > 180/105 addressed appropriately? (SMCH)[post_tpa_bp_addressed] | descriptive |
| 1046 | desc_fallout_post_tpa<br><br>Show the field ONLY if:<br>(([site]='SMCH' or [site]='SMCW') and [calc_post_tpa_monitoring]='0') or ([site]='SMCA' and ([treatment_type(IVtPA)]='1' or [gs_ivthroinit]='1' or [gs_ivtpaoutside]='1') and [tpa_tool_notes]<>'') | | descriptive |
| 1047 | fallout_notified_post_tpa<br><br>Show the field ONLY if:<br>(([site]='SMCH' or [site]='SMCW') and [calc_post_tpa_monitoring]='0') or ([site]='SMCA' and ([treatment_type(IVtPA)]='1' or [gs_ivthroinit]='1' or [gs_ivtpaoutside]='1') and [tpa_tool_notes]<>'') | Fallout Notification Sent To:<br>*Name or Email* | text<br>Custom alignment: RH |
| 1048 | fallout_date_post_tpa<br><br>Show the field ONLY if:<br>(([site]='SMCH' or [site]='SMCW') and [calc_post_tpa_monitoring]='0') or ([site]='SMCA' and ([treatment_type(IVtPA)]='1' or [gs_ivthroinit]='1' or [gs_ivtpaoutside]='1') and [tpa_tool_notes]<>'') | Date Notified | text (date_mdy, Min: 2016-01-01, Max: 2099-12-31)<br>Custom alignment: RH |
| 1049 | fallout_alert_post_tpa<br><br>Show the field ONLY if:<br>(([site]='SMCH' or [site]='SMCW') and [calc_post_tpa_monitoring]='0') or ([site]='SMCA' and ([treatment_type(IVtPA)]='1' or [gs_ivthroinit]='1' or [gs_ivtpaoutside]='1') and [tpa_tool_notes]<>'') | Email Notification: | radio<br>1 Send Email<br>2 Email(s) Sent<br><br>Custom alignment: RH |

| | | | |
|---|---|---|---|
| 1050 | fallout_verify_post_tpa<br><br>Show the field ONLY if:<br>(([site]='SMCH' or [site]='SMCW') and [calc_post_tpa_monitoring]='0') or ([site]='SMCA' and ([treatment_type(IVtPA)]='1' or [gs_ivthroinit]='1' or [gs_ivtpaoutside]='1') and [tpa_tool_notes]<>'') | Reviewed by Stroke Coordinator: | checkbox<br>1 fallout_verify_post_tpa___1 |
| 1051 | fallout_resp_desc_post_tpa<br><br>Show the field ONLY if:<br>(([site]='SMCH' or [site]='SMCW') and [calc_post_tpa_monitoring]='0') or ([site]='SMCA' and ([treatment_type(IVtPA)]='1' or [gs_ivthroinit]='1' or [gs_ivtpaoutside]='1') and [tpa_tool_notes]<>'') | Fallout Review: [fallout_review_nihss]<br>Fallout Response: [fallout_response_nihss] | descriptive |
| 1052 | fallout_info_ev_proc_1<br><br>Show the field ONLY if:<br>[ev_proc_protocol_1]='0' or [ev_proc_ir_1]='0' or [ev_proc_transit_1]='0' or ([calc_ev_proc_assessment_1]<>'' and [calc_ev_proc_assessment_1]<>100) or ([ev_proc_type_1]='1' and [calc_post_tpa_compliance]<>'' and [calc_post_tpa_compliance]<>100) or ([site]='SMCA' and ([ev_proc_assessment_notes]<>'' or [tpa_tool_notes]<>'')) | Section Header: *Fallout: Post IR Assessments Site: [site] MRN: [mrn] Patient Name: [jc_lastname], [jc_firstname] Fallout Details: [ev_proc_assessment_notes]*<br><br>1st Procedure: [ev_proc_type_1] Were Groin & Pedal Pulse Assessment Orders present? [ev_proc_orders_1] If no orders were present, was unit specific protocol followed? [ev_proc_protocol_1] Procedure End Time: [ev_proc_end_1] Time patient left IR Suite: [ev_proc_depart_1] Were all appropriate assessments done in IR Suite? [ev_proc_ir_1] Patient destination: [ev_proc_dest_1] Destination Arrival Time: [ev_proc_arrival_1] Were all appropriate assessments documented between IR Suite and patient destination? [ev_proc_transit_1] Groin & Pedal Pulse Assessment Compliance for this procedure: [calc_ev_proc_assessment_1]% | descriptive |
| 1053 | fallout_info_ev_proc_vs_1<br><br>Show the field ONLY if:<br>(([ev_proc_protocol_1]='0' or [ev_proc_ir_1]='0' or [ev_proc_transit_1]='0' or ([calc_ev_proc_assessment_1]<>'' and [calc_ev_proc_assessment_1]<>100)) and ([ev_proc_type_1]='1' and [calc_post_tpa_compliance]<>'' and [calc_post_tpa_compliance]<>100)) or ([site]='SMCA' and [ev_proc_type_1]='1' and ([ev_proc_assessment_notes]<>'' and [tpa_tool_notes]<>'')) | Neuro Checks Numerator/Denominator: [tpa_tool_nvs_num]/[tpa_tool_nvs_den] Vital Signs Numerator/Denominator: [tpa_tool_vs_num]/[tpa_tool_vs_den] Post IR Neuro Check/Vital Signs Monitoring Compliance: [calc_post_tpa_compliance]% Post Thrombolytic, Thrombectomy, and/or Endovascular Therapy Monitoring notes. [tpa_tool_notes] | descriptive |

| | | | |
|---|---|---|---|
| 1054 | desc_fallout_ev_proc_1<br><br>Show the field ONLY if:<br>[ev_proc_protocol_1]='0' or [ev_proc_ir_1]='0' or [ev_proc_transit_1]='0' or ([calc_ev_proc_assessment_1]<>'' and [calc_ev_proc_assessment_1]<>100) or ([ev_proc_type_1]='1' and [calc_post_tpa_compliance]<>'' and [calc_post_tpa_compliance]<>100) or ([site]='SMCA' and ([ev_proc_assessment_notes]<>'' or [tpa_tool_notes]<>'')) | | descriptive |
| 1055 | fallout_notified_ev_proc_1<br><br>Show the field ONLY if:<br>[ev_proc_protocol_1]='0' or [ev_proc_ir_1]='0' or [ev_proc_transit_1]='0' or ([calc_ev_proc_assessment_1]<>'' and [calc_ev_proc_assessment_1]<>100) or ([ev_proc_type_1]='1' and [calc_post_tpa_compliance]<>'' and [calc_post_tpa_compliance]<>100) or ([site]='SMCA' and ([ev_proc_assessment_notes]<>'' or [tpa_tool_notes]<>'')) | Fallout Notification Sent To:<br>*Name or Email* | text<br>Custom alignment: RH |
| 1056 | fallout_date_ev_proc_1<br><br>Show the field ONLY if:<br>[ev_proc_protocol_1]='0' or [ev_proc_ir_1]='0' or [ev_proc_transit_1]='0' or ([calc_ev_proc_assessment_1]<>'' and [calc_ev_proc_assessment_1]<>100) or ([ev_proc_type_1]='1' and [calc_post_tpa_compliance]<>'' and [calc_post_tpa_compliance]<>100) or ([site]='SMCA' and ([ev_proc_assessment_notes]<>'' or [tpa_tool_notes]<>'')) | Date Notified | text (date_mdy, Min: 2016-01-01, Max: 2099-12-31)<br>Custom alignment: RH |

| | | | |
|---|---|---|---|
| 1057 | fallout_alert_ev_proc_1<br><br>Show the field ONLY if:<br>[ev_proc_protocol_1]='0' or [ev_proc_ir_1]='0' or [ev_proc_transit_1]='0' or ([calc_ev_proc_assessment_1]<>'' and [calc_ev_proc_assessment_1]<>100) or ([ev_proc_type_1]='1' and [calc_post_tpa_compliance]<>'' and [calc_post_tpa_compliance]<>100) or ([site]='SMCA' and ([ev_proc_assessment_notes]<>'' or [tpa_tool_notes]<>'')) and [fallout_verify_ev_proc_1(1)]='0' | Email Notification: | radio<br>1 Send Email<br>2 Email(s) Sent<br><br>Custom alignment: RH |
| 1058 | fallout_verify_ev_proc_1<br><br>Show the field ONLY if:<br>[ev_proc_protocol_1]='0' or [ev_proc_ir_1]='0' or [ev_proc_transit_1]='0' or ([calc_ev_proc_assessment_1]<>'' and [calc_ev_proc_assessment_1]<>100) or ([ev_proc_type_1]='1' and [calc_post_tpa_compliance]<>'' and [calc_post_tpa_compliance]<>100) or ([site]='SMCA' and ([ev_proc_assessment_notes]<>'' or [tpa_tool_notes]<>'')) | Reviewed by Stroke Coordinator: | checkbox<br>1 fallout_verify_ev_proc_1___1 |
| 1059 | fallout_resp_dsc_ev_proc_1<br><br>Show the field ONLY if:<br>[ev_proc_protocol_1]='0' or [ev_proc_ir_1]='0' or [ev_proc_transit_1]='0' or ([calc_ev_proc_assessment_1]<>'' and [calc_ev_proc_assessment_1]<>100) or ([ev_proc_type_1]='1' and [calc_post_tpa_compliance]<>'' and [calc_post_tpa_compliance]<>100) or ([site]='SMCA' and ([ev_proc_assessment_notes]<>'' or [tpa_tool_notes]<>'')) | Fallout Review:[fallout_review_ev_proc_1]Fallout Response:[fallout_response_ev_proc_1] | descriptive |

| | | | |
|---|---|---|---|
| 1060 | fallout_info_ev_proc_2<br><br>Show the field ONLY if: [ev_proc_protocol_2]='0' or [ev_proc_ir_2]='0' or [ev_proc_transit_2]='0' or ([calc_ev_proc_assessment_2]<>'' and [calc_ev_proc_assessment_2]<>100) or ([ev_proc_type_2]='1' and [calc_post_tpa_compliance]<>'' and [calc_post_tpa_compliance]<>100) or ([site]='SMCA' and [ev_proc_type_2]<>'' and ([ev_proc_assessment_notes]<>'' or [tpa_tool_notes]<>'')) | 2nd Procedure: [ev_proc_type_2] Were Groin & Pedal Pulse Assessment Orders present? [ev_proc_orders_2] If no orders were present, was unit specific protocol followed? [ev_proc_protocol_2] Procedure End Time: [ev_proc_end_2] Time patient left IR Suite: [ev_proc_depart_2] Were all appropriate assessments done in IR Suite? [ev_proc_ir_2] Patient destination: [ev_proc_dest_2] Destination Arrival Time: [ev_proc_arrival_2] Were all appropriate assessments documented between IR Suite and patient destination? [ev_proc_transit_2] Groin & Pedal Pulse Assessment Compliance for this procedure: [calc_ev_proc_assessment_2]% | descriptive |
| 1061 | fallout_info_ev_proc_vs_2<br><br>Show the field ONLY if: (([ev_proc_protocol_2]='0' or [ev_proc_ir_2]='0' or [ev_proc_transit_2]='0' or ([calc_ev_proc_assessment_2]<>'' and [calc_ev_proc_assessment_2]<>100)) and ([ev_proc_type_2]='1' and [calc_post_tpa_compliance]<>'' and [calc_post_tpa_compliance]<>100)) or ([site]='SMCA' and [ev_proc_type_2]='1' and ([ev_proc_assessment_notes]<>'' and [tpa_tool_notes]<>'')) | Neuro Checks Numerator/Denominator: [tpa_tool_nvs_num]/[tpa_tool_nvs_den] Vital Signs Numerator/Denominator: [tpa_tool_vs_num]/[tpa_tool_vs_den] Post IR Neuro Check/Vital Signs Monitoring Compliance: [calc_post_tpa_compliance]% Post Thrombolytic, Thrombectomy, and/or Endovascular Therapy Monitoring notes. [tpa_tool_notes] | descriptive |
| 1062 | desc_fallout_ev_proc_2<br><br>Show the field ONLY if: [ev_proc_protocol_2]='0' or [ev_proc_ir_2]='0' or [ev_proc_transit_2]='0' or ([calc_ev_proc_assessment_2]<>'' and [calc_ev_proc_assessment_2]<>100) or ([ev_proc_type_2]='1' and [calc_post_tpa_compliance]<>'' and [calc_post_tpa_compliance]<>100) or ([site]='SMCA' and [ev_proc_type_2]<>'' and ([ev_proc_assessment_notes]<>'' or [tpa_tool_notes]<>'')) | | descriptive |

| | | | |
|---|---|---|---|
| 1063 | fallout_notified_ev_proc_2<br><br>Show the field ONLY if:<br>[ev_proc_protocol_2]='0' or [ev_proc_ir_2]='0' or [ev_proc_transit_2]='0' or ([calc_ev_proc_assessment_2]<>'' and [calc_ev_proc_assessment_2]<>100) or ([ev_proc_type_2]='1' and [calc_post_tpa_compliance]<>'' and [calc_post_tpa_compliance]<>100) or ([site]='SMCA' and [ev_proc_type_2]<>'' and ([ev_proc_assessment_notes]<>'' or [tpa_tool_notes]<>'')) | Fallout Notification Sent To:<br>*Name or Email* | text<br>Custom alignment: RH |
| 1064 | fallout_date_ev_proc_2<br><br>Show the field ONLY if:<br>[ev_proc_protocol_2]='0' or [ev_proc_ir_2]='0' or [ev_proc_transit_2]='0' or ([calc_ev_proc_assessment_2]<>'' and [calc_ev_proc_assessment_2]<>100) or ([ev_proc_type_2]='1' and [calc_post_tpa_compliance]<>'' and [calc_post_tpa_compliance]<>100) or ([site]='SMCA' and [ev_proc_type_2]<>'' and ([ev_proc_assessment_notes]<>'' or [tpa_tool_notes]<>'')) | Date Notified | text (date_mdy, Min: 2016-01-01, Max: 2099-12-31)<br>Custom alignment: RH |
| 1065 | fallout_alert_ev_proc_2<br><br>Show the field ONLY if:<br>[ev_proc_protocol_2]='0' or [ev_proc_ir_2]='0' or [ev_proc_transit_2]='0' or ([calc_ev_proc_assessment_2]<>'' and [calc_ev_proc_assessment_2]<>100) or ([ev_proc_type_2]='1' and [calc_post_tpa_compliance]<>'' and [calc_post_tpa_compliance]<>100) or ([site]='SMCA' and ([ev_proc_assessment_notes]<>'' or [tpa_tool_notes]<>'')) and [fallout_verify_ev_proc_2(1)]='0' | Email Notification: | radio<br>1 \| Send Email<br>2 \| Email(s) Sent<br><br>Custom alignment: RH |

| | | | |
|---|---|---|---|
| 1066 | fallout_verify_ev_proc_2<br><br>Show the field ONLY if: [ev_proc_protocol_2]='0' or [ev_proc_ir_2]='0' or [ev_proc_transit_2]='0' or ([calc_ev_proc_assessment_2]<>'' and [calc_ev_proc_assessment_2]<>100) or ([ev_proc_type_2]='1' and [calc_post_tpa_compliance]<>'' and [calc_post_tpa_compliance]<>100) or ([site]='SMCA' and [ev_proc_type_2]<>'' and ([ev_proc_assessment_notes]<>'' or [tpa_tool_notes]<>'')) | Reviewed by Stroke Coordinator: | checkbox<br>1 \| fallout_verify_ev_proc_2___1 |
| 1067 | fallout_resp_dsc_ev_proc_2<br><br>Show the field ONLY if: [ev_proc_protocol_2]='0' or [ev_proc_ir_2]='0' or [ev_proc_transit_2]='0' or ([calc_ev_proc_assessment_2]<>'' and [calc_ev_proc_assessment_2]<>100) or ([ev_proc_type_2]='1' and [calc_post_tpa_compliance]<>'' and [calc_post_tpa_compliance]<>100) or ([site]='SMCA' and [ev_proc_type_2]<>'' and ([ev_proc_assessment_notes]<>'' or [tpa_tool_notes]<>'')) | Fallout Review:[fallout_review_ev_proc_2]Fallout Response: [fallout_response_ev_proc_2] | descriptive |
| 1068 | fallout_info_ev_proc_3<br><br>Show the field ONLY if: [ev_proc_protocol_3]='0' or [ev_proc_ir_3]='0' or [ev_proc_transit_3]='0' or ([calc_ev_proc_assessment_3]<>'' and [calc_ev_proc_assessment_3]<>100) or ([ev_proc_type_3]='1' and [calc_post_tpa_compliance]<>'' and [calc_post_tpa_compliance]<>100) or ([site]='SMCA' and [ev_proc_type_3]<>'' and ([ev_proc_assessment_notes]<>'' or [tpa_tool_notes]<>'')) | 3rd Procedure: [ev_proc_type_3] Were Groin & Pedal Pulse Assessment Orders present? [ev_proc_orders_3] If no orders were present, was unit specific protocol followed? [ev_proc_protocol_3] Procedure End Time: [ev_proc_end_3] Time patient left IR Suite: [ev_proc_depart_3] Were all appropriate assessments done in IR Suite? [ev_proc_ir_3] Patient destination: [ev_proc_dest_3] Destination Arrival Time: [ev_proc_arrival_3] Were all appropriate assessments documented between IR Suite and patient destination? [ev_proc_transit_3] Groin & Pedal Pulse Assessment Compliance for this procedure: [calc_ev_proc_assessment_3]% | descriptive |

| | | | |
|---|---|---|---|
| 1069 | fallout_info_ev_proc_vs_3<br><br>Show the field ONLY if:<br>(([ev_proc_protocol_3]='0' or [ev_proc_ir_3]='0' or [ev_proc_transit_3]='0' or ([calc_ev_proc_assessment_3]<>'' and [calc_ev_proc_assessment_3]<>100)) and ([ev_proc_type_3]='1' and [calc_post_tpa_compliance]<>'' and [calc_post_tpa_compliance]<>100)) or ([site]='SMCA' and [ev_proc_type_3]='1' and ([ev_proc_assessment_notes]<>'' and [tpa_tool_notes]<>'')) | Neuro Checks Numerator/Denominator: [tpa_tool_nvs_num]/[tpa_tool_nvs_den] Vital Signs Numerator/Denominator: [tpa_tool_vs_num]/[tpa_tool_vs_den] Post IR Neuro Check/Vital Signs Monitoring Compliance: [calc_post_tpa_compliance]% Post Thrombolytic, Thrombectomy, and/or Endovascular Therapy Monitoring notes. [tpa_tool_notes] | descriptive |
| 1070 | desc_fallout_ev_proc_3<br><br>Show the field ONLY if:<br>[ev_proc_protocol_3]='0' or [ev_proc_ir_3]='0' or [ev_proc_transit_3]='0' or ([calc_ev_proc_assessment_3]<>'' and [calc_ev_proc_assessment_3]<>100) or ([ev_proc_type_3]='1' and [calc_post_tpa_compliance]<>'' and [calc_post_tpa_compliance]<>100) or ([site]='SMCA' and [ev_proc_type_3]<>'' and ([ev_proc_assessment_notes]<>'' or [tpa_tool_notes]<>'')) | | descriptive |
| 1071 | fallout_notified_ev_proc_3<br><br>Show the field ONLY if:<br>[ev_proc_protocol_3]='0' or [ev_proc_ir_3]='0' or [ev_proc_transit_3]='0' or ([calc_ev_proc_assessment_3]<>'' and [calc_ev_proc_assessment_3]<>100) or ([ev_proc_type_3]='1' and [calc_post_tpa_compliance]<>'' and [calc_post_tpa_compliance]<>100) or ([site]='SMCA' and [ev_proc_type_3]<>'' and ([ev_proc_assessment_notes]<>'' or [tpa_tool_notes]<>'')) | Fallout Notification Sent To:<br>*Name or Email* | text<br>Custom alignment: RH |

| | | | |
|---|---|---|---|
| 1072 | fallout_date_ev_proc_3<br><br>Show the field ONLY if:<br>[ev_proc_protocol_3]='0' or [ev_proc_ir_3]='0' or [ev_proc_transit_3]='0' or ([calc_ev_proc_assessment_3]<>'' and [calc_ev_proc_assessment_3]<>100) or ([ev_proc_type_3]='1' and [calc_post_tpa_compliance]<>'' and [calc_post_tpa_compliance]<>100) or ([site]='SMCA' and [ev_proc_type_3]<>'' and ([ev_proc_assessment_notes]<>'' or [tpa_tool_notes]<>'')) | Date Notified | text (date_mdy, Min: 2016-01-01, Max: 2099-12-31)<br>Custom alignment: RH |
| 1073 | fallout_alert_ev_proc_3<br><br>Show the field ONLY if:<br>[ev_proc_protocol_3]='0' or [ev_proc_ir_3]='0' or [ev_proc_transit_3]='0' or ([calc_ev_proc_assessment_3]<>'' and [calc_ev_proc_assessment_3]<>100) or ([ev_proc_type_3]='1' and [calc_post_tpa_compliance]<>'' and [calc_post_tpa_compliance]<>100) or ([site]='SMCA' and ([ev_proc_assessment_notes]<>'' or [tpa_tool_notes]<>'')) and [fallout_verify_ev_proc_3(1)]='0' | Email Notification: | radio<br>1 Send Email<br>2 Email(s) Sent<br><br>Custom alignment: RH |
| 1074 | fallout_verify_ev_proc_3<br><br>Show the field ONLY if:<br>[ev_proc_protocol_3]='0' or [ev_proc_ir_3]='0' or [ev_proc_transit_3]='0' or ([calc_ev_proc_assessment_3]<>'' and [calc_ev_proc_assessment_3]<>100) or ([ev_proc_type_3]='1' and [calc_post_tpa_compliance]<>'' and [calc_post_tpa_compliance]<>100) or ([site]='SMCA' and [ev_proc_type_3]<>'' and ([ev_proc_assessment_notes]<>'' or [tpa_tool_notes]<>'')) | Reviewed by Stroke Coordinator: | checkbox<br>1 fallout_verify_ev_proc_3___1 |

| | | | |
|---|---|---|---|
| 1075 | fallout_resp_dsc_ev_proc_3<br>Show the field ONLY if: [ev_proc_protocol_3]='0' or [ev_proc_ir_3]='0' or [ev_proc_transit_3]='0' or ([calc_ev_proc_assessment_3]<>'' and [calc_ev_proc_assessment_3]<>100) or ([ev_proc_type_3]='1' and [calc_post_tpa_compliance]<>'' and [calc_post_tpa_compliance]<>100) or ([site]='SMCA' and [ev_proc_type_3]<>'' and ([ev_proc_assessment_notes]<>'' or [tpa_tool_notes]<>'')) | Fallout Review:[fallout_review_ev_proc_3]Fallout Response: [fallout_response_ev_proc_3] | descriptive |
| 1076 | fallout_info_ev_proc_4<br>Show the field ONLY if: [ev_proc_protocol_4]='0' or [ev_proc_ir_4]='0' or [ev_proc_transit_4]='0' or ([calc_ev_proc_assessment_4]<>'' and [calc_ev_proc_assessment_4]<>100) or ([ev_proc_type_4]='1' and [calc_post_tpa_compliance]<>'' and [calc_post_tpa_compliance]<>100) or ([site]='SMCA' and [ev_proc_type_4]<>'' and ([ev_proc_assessment_notes]<>'' or [tpa_tool_notes]<>'')) | 4th Procedure: [ev_proc_type_4] Were Groin & Pedal Pulse Assessment Orders present? [ev_proc_orders_4] If no orders were present, was unit specific protocol followed? [ev_proc_protocol_4] Procedure End Time: [ev_proc_end_4] Time patient left IR Suite: [ev_proc_depart_4] Were all appropriate assessments done in IR Suite? [ev_proc_ir_4] Patient destination: [ev_proc_dest_4] Destination Arrival Time: [ev_proc_arrival_4] Were all appropriate assessments documented between IR Suite and patient destination? [ev_proc_transit_4] Groin & Pedal Pulse Assessment Compliance for this procedure: [calc_ev_proc_assessment_4]% | descriptive |
| 1077 | fallout_info_ev_proc_vs_4<br>Show the field ONLY if: (([ev_proc_protocol_4]='0' or [ev_proc_ir_4]='0' or [ev_proc_transit_4]='0' or ([calc_ev_proc_assessment_4]<>'' and [calc_ev_proc_assessment_4]<>100)) and ([ev_proc_type_4]='1' and [calc_post_tpa_compliance]<>'' and [calc_post_tpa_compliance]<>100)) or ([site]='SMCA' and [ev_proc_type_4]='1' and ([ev_proc_assessment_notes]<>'' and [tpa_tool_notes]<>'')) | Neuro Checks Numerator/Denominator: [tpa_tool_nvs_num]/[tpa_tool_nvs_den] Vital Signs Numerator/Denominator: [tpa_tool_vs_num]/[tpa_tool_vs_den] Post IR Neuro Check/Vital Signs Monitoring Compliance: [calc_post_tpa_compliance]% Post Thrombolytic, Thrombectomy, and/or Endovascular Therapy Monitoring notes. [tpa_tool_notes] | descriptive |

| | | | |
|---|---|---|---|
| 1078 | desc_fallout_ev_proc_4<br><br>Show the field ONLY if:<br>[ev_proc_protocol_4]='0' or [ev_proc_ir_4]='0' or [ev_proc_transit_4]='0' or ([calc_ev_proc_assessment_4]<>'' and [calc_ev_proc_assessment_4]<>100) or ([ev_proc_type_4]='1' and [calc_post_tpa_compliance]<>'' and [calc_post_tpa_compliance]<>100) or ([site]='SMCA' and [ev_proc_type_4]<>'' and ([ev_proc_assessment_notes]<>'' or [tpa_tool_notes]<>'')) | | descriptive |
| 1079 | fallout_notified_ev_proc_4<br><br>Show the field ONLY if:<br>[ev_proc_protocol_4]='0' or [ev_proc_ir_4]='0' or [ev_proc_transit_4]='0' or ([calc_ev_proc_assessment_4]<>'' and [calc_ev_proc_assessment_4]<>100) or ([ev_proc_type_4]='1' and [calc_post_tpa_compliance]<>'' and [calc_post_tpa_compliance]<>100) or ([site]='SMCA' and [ev_proc_type_4]<>'' and ([ev_proc_assessment_notes]<>'' or [tpa_tool_notes]<>'')) | Fallout Notification Sent To:<br>*Name or Email* | text<br>Custom alignment: RH |
| 1080 | fallout_date_ev_proc_4<br><br>Show the field ONLY if:<br>[ev_proc_protocol_4]='0' or [ev_proc_ir_4]='0' or [ev_proc_transit_4]='0' or ([calc_ev_proc_assessment_4]<>'' and [calc_ev_proc_assessment_4]<>100) or ([ev_proc_type_4]='1' and [calc_post_tpa_compliance]<>'' and [calc_post_tpa_compliance]<>100) or ([site]='SMCA' and [ev_proc_type_4]<>'' and ([ev_proc_assessment_notes]<>'' or [tpa_tool_notes]<>'')) | Date Notified | text (date_mdy, Min: 2016-01-01, Max: 2099-12-31)<br>Custom alignment: RH |

| | | | |
|---|---|---|---|
| 1081 | fallout_alert_ev_proc_4<br><br>Show the field ONLY if:<br>[ev_proc_protocol_4]='0' or [ev_proc_ir_4]='0' or [ev_proc_transit_4]='0' or ([calc_ev_proc_assessment_4]<>'' and [calc_ev_proc_assessment_4]<>100) or ([ev_proc_type_4]='1' and [calc_post_tpa_compliance]<>'' and [calc_post_tpa_compliance]<>100) or ([site]='SMCA' and ([ev_proc_assessment_notes]<>'' or [tpa_tool_notes]<>'')) and [fallout_verify_ev_proc_4(1)]='0' | Email Notification: | radio<br>1 Send Email<br>2 Email(s) Sent<br><br>Custom alignment: RH |
| 1082 | fallout_verify_ev_proc_4<br><br>Show the field ONLY if:<br>[ev_proc_protocol_4]='0' or [ev_proc_ir_4]='0' or [ev_proc_transit_4]='0' or ([calc_ev_proc_assessment_4]<>'' and [calc_ev_proc_assessment_4]<>100) or ([ev_proc_type_4]='1' and [calc_post_tpa_compliance]<>'' and [calc_post_tpa_compliance]<>100) or ([site]='SMCA' and [ev_proc_type_4]<>'' and ([ev_proc_assessment_notes]<>'' or [tpa_tool_notes]<>'')) | Reviewed by Stroke Coordinator: | checkbox<br>1 fallout_verify_ev_proc_4___1 |
| 1083 | fallout_resp_dsc_ev_proc_4<br><br>Show the field ONLY if:<br>[ev_proc_protocol_4]='0' or [ev_proc_ir_4]='0' or [ev_proc_transit_4]='0' or ([calc_ev_proc_assessment_4]<>'' and [calc_ev_proc_assessment_4]<>100) or ([ev_proc_type_4]='1' and [calc_post_tpa_compliance]<>'' and [calc_post_tpa_compliance]<>100) or ([site]='SMCA' and [ev_proc_type_4]<>'' and ([ev_proc_assessment_notes]<>'' or [tpa_tool_notes]<>'')) | Fallout Review:[fallout_review_ev_proc_4]Fallout Response:[fallout_response_ev_proc_4] | descriptive |
| 1084 | fallouts_complete | Section Header: *Form Status*<br><br>Complete? | dropdown<br>0 Incomplete<br>1 Unverified<br>2 Complete |

Instrument: **Fallout Review** (fallout_review)

Collapse

| | | | |
|---|---|---|---|
| 1085 | fallout_review_stk_1<br><br>Show the field ONLY if:<br>[calc_stk_1]='0' or [calc_gwtg_vte_prophylaxis]='0' | Section Header: *Fallout: VTE ProphylaxisSITE: [site] MRN: [mrn]Patient Name:[jc_lastname], [jc_firstname]Fallout Details:[fallout_detail_stk_1]Fallout Unit:[fallout_unit_stk_1]Date Notified:[fallout_date_stk_1]Notification Sent To:[fallout_notified_stk_1]*<br><br>Fallout Validation<br>*If flagged for further review, include reason in response below.Selecting Further Review Needed will trigger notification email.* | radio<br>1 Reviewed and Acknowledged<br>2 Further Review Needed by Stroke Coordinator |
| 1086 | desc_fallout_rev_ack_stk_1<br><br>Show the field ONLY if:<br>[fallout_review_stk_1]='1' | Fallout Reviewed and Acknowledged:<br><br>Please provide Discovery Details and Action Plan(s) below. | descriptive |
| 1087 | desc_fallout_fur_rev_stk_1<br><br>Show the field ONLY if:<br>[fallout_review_stk_1]='2' | Further Review Needed by Stroke Coordinator:<br><br>Please provide Discovery Details and Reason(s) for Further Review. | descriptive |
| 1088 | fallout_response_stk_1<br><br>Show the field ONLY if:<br>[fallout_review_stk_1]='1' or [fallout_review_stk_1]='2' | VTE Prophylaxis Fallout Response / Discovery Details: | notes |
| 1089 | fallout_ap_indiv_stk_1<br><br>Show the field ONLY if:<br>[fallout_review_stk_1]='1' | Fallout Action Plan - Individual: | notes |
| 1090 | fallout_ap_unit_stk_1<br><br>Show the field ONLY if:<br>[fallout_review_stk_1]='1' | Fallout Action Plan - Unit: | notes |
| 1091 | fallout_review_stk_2<br><br>Show the field ONLY if:<br>[calc_stk_2] = '0' or [calc_gwtg_antithrombotics] = '0' | Section Header: *Fallout: Discharged on Antithrombotic TherapySITE: [site]MRN: [mrn]Patient Name:[jc_lastname], [jc_firstname]Fallout Details:[fallout_detail_stk_2]Fallout Unit:[fallout_unit_stk_2]Date Notified:[fallout_date_stk_2]Notification Sent To: [fallout_notified_stk_2]*<br><br>Fallout Validation<br>*If flagged for further review, include reason in response below.Selecting Further Review Needed will trigger notification email.* | radio<br>1 Reviewed / Acknowledged<br>2 Further Review Needed |
| 1092 | desc_fallout_rev_ack_stk_2<br><br>Show the field ONLY if:<br>[fallout_review_stk_2]='1' | Fallout Reviewed and Acknowledged:<br><br>Please provide Discovery Details and Action Plan(s) below. | descriptive |
| 1093 | desc_fallout_fur_rev_stk_2<br><br>Show the field ONLY if:<br>[fallout_review_stk_2]='2' | Further Review Needed by Stroke Coordinator:<br><br>Please provide Discovery Details and Reason(s) for Further Review. | descriptive |

| | | | |
|---|---|---|---|
| 1094 | fallout_response_stk_2<br><br>Show the field ONLY if:<br>[fallout_review_stk_2]='1' or [fallout_review_stk_2]='2' | Discharged on Antithrombotic Therapy Fallout Response / Discovery Details: | notes |
| 1095 | fallout_ap_indiv_stk_2<br><br>Show the field ONLY if:<br>[fallout_review_stk_2]='1' | Fallout Action Plan - Individual: | notes |
| 1096 | fallout_ap_unit_stk_2<br><br>Show the field ONLY if:<br>[fallout_review_stk_2]='1' | Fallout Action Plan - Unit: | notes |
| 1097 | fallout_review_stk_3<br><br>Show the field ONLY if:<br>[calc_stk_3] = '0' or [calc_gwtg_anticoag_afib] = '0' | Section Header: *Fallout: Anticoagulation Therapy for Atrial Fibrillation/FlutterSITE: [site]MRN: [mrn]Patient Name:[jc_lastname], [jc_firstname]Fallout Details: [fallout_detail_stk_3]Fallout Unit:[fallout_unit_stk_3]Date Notified: [fallout_date_stk_3]Notification Sent To:[fallout_notified_stk_3]*<br><br>Fallout Validation<br>*If flagged for further review, include reason in response below.Selecting Further Review Needed will trigger notification email.* | radio<br>1 Reviewed / Acknowledged<br>2 Further Review Needed |
| 1098 | desc_fallout_rev_ack_stk_3<br><br>Show the field ONLY if:<br>[fallout_review_stk_3]='1' | Fallout Reviewed and Acknowledged:<br><br>Please provide Discovery Details and Action Plan(s) below. | descriptive |
| 1099 | desc_fallout_fur_rev_stk_3<br><br>Show the field ONLY if:<br>[fallout_review_stk_3]='2' | Further Review Needed by Stroke Coordinator:<br><br>Please provide Discovery Details and Reason(s) for Further Review. | descriptive |
| 1100 | fallout_response_stk_3<br><br>Show the field ONLY if:<br>[fallout_review_stk_3]='1' or [fallout_review_stk_3]='2' | Anticoagulation Therapy for Atrial Fibrillation/Flutter Fallout Response / Discovery Details: | notes |
| 1101 | fallout_ap_indiv_stk_3<br><br>Show the field ONLY if:<br>[fallout_review_stk_3]='1' | Fallout Action Plan - Individual: | notes |
| 1102 | fallout_ap_unit_stk_3<br><br>Show the field ONLY if:<br>[fallout_review_stk_3]='1' | Fallout Action Plan - Unit: | notes |
| 1103 | fallout_review_stk_4<br><br>Show the field ONLY if:<br>[calc_stk_4] = '0' or [calc_gwtg_arrive2_treat3] = '0' | Section Header: *Fallout: Thrombolytic TherapySITE: [site] MRN: [mrn]Patient Name: [jc_lastname], [jc_firstname]Fallout Details:[fallout_detail_stk_4]Fallout Unit: [fallout_unit_stk_4]Date Notified:[fallout_date_stk_4]Notification Sent To: [fallout_notified_stk_4]*<br><br>Fallout Validation<br>*If flagged for further review, include reason in response below.Selecting Further Review Needed will trigger notification email.* | radio<br>1 Reviewed / Acknowledged<br>2 Further Review Needed |

| | | | |
|---|---|---|---|
| 1104 | desc_fallout_rev_ack_stk_4<br><br>Show the field ONLY if:<br>[fallout_review_stk_4]='1' | Fallout Reviewed and Acknowledged:<br><br>Please provide Discovery Details and Action Plan(s) below. | descriptive |
| 1105 | desc_fallout_fur_rev_stk_4<br><br>Show the field ONLY if:<br>[fallout_review_stk_4]='2' | Further Review Needed by Stroke Coordinator:<br><br>Please provide Discovery Details and Reason(s) for Further Review. | descriptive |
| 1106 | fallout_response_stk_4<br><br>Show the field ONLY if:<br>[fallout_review_stk_4]='1' or [fallout_review_stk_4]='2' | Thrombolytic Therapy Fallout Response / Discovery Details: | notes |
| 1107 | fallout_ap_indiv_stk_4<br><br>Show the field ONLY if:<br>[fallout_review_stk_4]='1' | Fallout Action Plan - Individual: | notes |
| 1108 | fallout_ap_unit_stk_4<br><br>Show the field ONLY if:<br>[fallout_review_stk_4]='1' | Fallout Action Plan - Unit: | notes |
| 1109 | fallout_review_stk_5<br><br>Show the field ONLY if:<br>[calc_stk_5] = '0' or [calc_gwtg_early_antithrom] = '0' | Section Header: *Fallout: Antithrombotic Therapy By End of Hospital Day 2SITE: [site]MRN: [mrn]Patient Name:[jc_lastname], [jc_firstname]Fallout Details:[fallout_detail_stk_5]Fallout Unit:[fallout_unit_stk_5]Date Notified:[fallout_date_stk_5]Notification Sent To: [fallout_notified_stk_5]*<br><br>Fallout Validation<br>*If flagged for further review, include reason in response below.Selecting Further Review Needed will trigger notification email.* | radio<br>1 Reviewed / Acknowledged<br>2 Further Review Needed |
| 1110 | desc_fallout_rev_ack_stk_5<br><br>Show the field ONLY if:<br>[fallout_review_stk_5]='1' | Fallout Reviewed and Acknowledged:<br><br>Please provide Discovery Details and Action Plan(s) below. | descriptive |
| 1111 | desc_fallout_fur_rev_stk_5<br><br>Show the field ONLY if:<br>[fallout_review_stk_5]='2' | Further Review Needed by Stroke Coordinator:<br><br>Please provide Discovery Details and Reason(s) for Further Review. | descriptive |
| 1112 | fallout_response_stk_5<br><br>Show the field ONLY if:<br>[fallout_review_stk_5]='1' or [fallout_review_stk_5]='2' | Antithrombotic Therapy By End of Hospital Day 2 Fallout Response / Discovery Details: | notes |
| 1113 | fallout_ap_indiv_stk_5<br><br>Show the field ONLY if:<br>[fallout_review_stk_5]='1' | Fallout Action Plan - Individual: | notes |

| | | | |
|---|---|---|---|
| 1114 | fallout_ap_unit_stk_5<br><br>Show the field ONLY if:<br>[fallout_review_stk_5]='1' | Fallout Action Plan - Unit: | notes |
| 1115 | fallout_review_stk_6<br><br>Show the field ONLY if:<br>[calc_stk_6] = '0' or [calc_gwtg_statin_at_disch] = '0' | Section Header: *Fallout: Discharged on Statin MedicationSITE: [site] MRN: [mrn]Patient Name:[jc_lastname], [jc_firstname]Fallout Details:[fallout_detail_stk_6]Fallout Unit: [fallout_unit_stk_6]Date Notified:[fallout_date_stk_6]Notification Sent To: [fallout_notified_stk_6]*<br><br>Fallout Validation<br>*If flagged for further review, include reason in response below.Selecting Further Review Needed will trigger notification email.* | radio<br>1 Reviewed / Acknowledged<br>2 Further Review Needed |
| 1116 | desc_fallout_rev_ack_stk_6<br><br>Show the field ONLY if:<br>[fallout_review_stk_6]='1' | Fallout Reviewed and Acknowledged:<br><br>Please provide Discovery Details and Action Plan(s) below. | descriptive |
| 1117 | desc_fallout_fur_rev_stk_6<br><br>Show the field ONLY if:<br>[fallout_review_stk_6]='2' | Further Review Needed by Stroke Coordinator:<br><br>Please provide Discovery Details and Reason(s) for Further Review. | descriptive |
| 1118 | fallout_response_stk_6<br><br>Show the field ONLY if:<br>[fallout_review_stk_6]='1' or [fallout_review_stk_6]='2' | Discharged on Statin Medication Fallout Response / Discovery Details: | notes |
| 1119 | fallout_ap_indiv_stk_6<br><br>Show the field ONLY if:<br>[fallout_review_stk_6]='1' | Fallout Action Plan - Individual: | notes |
| 1120 | fallout_ap_unit_stk_6<br><br>Show the field ONLY if:<br>[fallout_review_stk_6]='1' | Fallout Action Plan - Unit: | notes |
| 1121 | fallout_review_stk_8<br><br>Show the field ONLY if:<br>[calc_stk_8] = '0' or [calc_gwtg_stroke_education] = '0' | Section Header: *Fallout: Stroke EducationSITE: [site] MRN: [mrn]Patient Name: [jc_lastname], [jc_firstname]Fallout Details:[fallout_detail_stk_8]Fallout Unit: [fallout_unit_stk_8]Date Notified:[fallout_date_stk_8]Notification Sent To: [fallout_notified_stk_8]*<br><br>Fallout Validation<br>*If flagged for further review, include reason in response below.Selecting Further Review Needed will trigger notification email.* | radio<br>1 Reviewed / Acknowledged<br>2 Further Review Needed |
| 1122 | desc_fallout_rev_ack_stk_8<br><br>Show the field ONLY if:<br>[fallout_review_stk_8]='1' | Fallout Reviewed and Acknowledged:<br><br>Please provide Discovery Details and Action Plan(s) below. | descriptive |
| 1123 | desc_fallout_fur_rev_stk_8<br><br>Show the field ONLY if:<br>[fallout_review_stk_8]='2' | Further Review Needed by Stroke Coordinator:<br><br>Please provide Discovery Details and Reason(s) for Further Review. | descriptive |

| | | | |
|---|---|---|---|
| 1124 | fallout_response_stk_8<br><br>Show the field ONLY if:<br>[fallout_review_stk_8]='1' or [fallout_review_stk_8]='2' | Stroke Education Fallout Response / Discovery Details: | notes |
| 1125 | fallout_ap_indiv_stk_8<br><br>Show the field ONLY if:<br>[fallout_review_stk_8]='1' | Fallout Action Plan - Individual: | notes |
| 1126 | fallout_ap_unit_stk_8<br><br>Show the field ONLY if:<br>[fallout_review_stk_8]='1' | Fallout Action Plan - Unit: | notes |
| 1127 | fallout_review_stk_10<br><br>Show the field ONLY if:<br>[calc_stk_10] = '0' or [calc_gwtg_rehab_considered] = '0' | Section Header: *Fallout: Assessed for RehabilitationSITE: [site] MRN: [mrn]Patient Name:[jc_lastname], [jc_firstname]Fallout Details:[fallout_detail_stk_10]Fallout Unit:[fallout_unit_stk_10]Date Notified:[fallout_date_stk_10]Notification Sent To:[fallout_notified_stk_10]*<br><br>Fallout Validation<br>*If flagged for further review, include reason in response below.Selecting Further Review Needed will trigger notification email.* | radio<br>1 Reviewed / Acknowledged<br>2 Further Review Needed |
| 1128 | desc_fallout_rev_ack_stk_10<br><br>Show the field ONLY if:<br>[fallout_review_stk_10]='1' | Fallout Reviewed and Acknowledged:<br><br>Please provide Discovery Details and Action Plan(s) below. | descriptive |
| 1129 | desc_fallout_fur_rev_stk_10<br><br>Show the field ONLY if:<br>[fallout_review_stk_10]='2' | Further Review Needed by Stroke Coordinator:<br><br>Please provide Discovery Details and Reason(s) for Further Review. | descriptive |
| 1130 | fallout_response_stk_10<br><br>Show the field ONLY if:<br>[fallout_review_stk_10]='1' or [fallout_review_stk_10]='2' | Assessed for Rehabilitation Fallout Response / Discovery Details: | notes |
| 1131 | fallout_ap_indiv_stk_10<br><br>Show the field ONLY if:<br>[fallout_review_stk_10]='1' | Fallout Action Plan - Individual: | notes |
| 1132 | fallout_ap_unit_stk_10<br><br>Show the field ONLY if:<br>[fallout_review_stk_10]='1' | Fallout Action Plan - Unit: | notes |
| 1133 | fallout_review_swallow<br><br>Show the field ONLY if:<br>[calc_gwtg_dysphagia_screen] = '0' or [calc_swallow_screen] = '0' | Section Header: *Fallout: Swallow ScreenSITE: [site] MRN: [mrn]Patient Name:[jc_lastname], [jc_firstname]Fallout Details:[fallout_detail_swallow]Fallout Unit:[fallout_unit_swallow]Date Notified:[fallout_date_swallow]Notification Sent To:[fallout_notified_swallow]*<br><br>Fallout Validation<br>*If flagged for further review, include reason in response below.Selecting Further Review Needed will trigger notification email.* | radio<br>1 Reviewed / Acknowledged<br>2 Further Review Needed |

| | | | |
|---|---|---|---|
| 1134 | desc_fallout_rev_ack_swallow<br><br>Show the field ONLY if:<br>[fallout_review_swallow]='1' | Fallout Reviewed and Acknowledged:<br><br>Please provide Discovery Details and Action Plan(s) below. | descriptive |
| 1135 | desc_fallout_fur_rev_swallow<br><br>Show the field ONLY if:<br>[fallout_review_swallow]='2' | Further Review Needed by Stroke Coordinator:<br><br>Please provide Discovery Details and Reason(s) for Further Review. | descriptive |
| 1136 | fallout_response_swallow<br><br>Show the field ONLY if:<br>[fallout_review_swallow]='1' or [fall out_review_swallow]='2' | Swallow Screen Fallout Response / Discovery Details: | notes |
| 1137 | fallout_ap_indiv_swallow<br><br>Show the field ONLY if:<br>[fallout_review_swallow]='1' | Fallout Action Plan - Individual: | notes |
| 1138 | fallout_ap_unit_swallow<br><br>Show the field ONLY if:<br>[fallout_review_swallow]='1' | Fallout Action Plan - Unit: | notes |
| 1139 | fallout_review_nihss<br><br>Show the field ONLY if:<br>[calc_gwtg_nihss_reported] = '0' or [calc_dnv_1] = '0' | Section Header: *Fallout: NIHSS ReportedSITE: [site] MRN: [mrn]Patient Name:[jc_lastname], [jc_firstname]Fallout Details:[fallout_detail_nihss]Fallout Unit:[fallout_unit_nihss]Date Notified:[fallout_date_nihss]Notification Sent To:[fallout_notified_nihss]*<br><br>Fallout Validation<br>*If flagged for further review, include reason in response below.Selecting Further Review Needed will trigger notification email.* | radio<br>1 Reviewed / Acknowledged<br>2 Further Review Needed |
| 1140 | desc_fallout_rev_ack_nihss<br><br>Show the field ONLY if:<br>[fallout_review_nihss]='1' | Fallout Reviewed and Acknowledged:<br><br>Please provide Discovery Details and Action Plan(s) below. | descriptive |
| 1141 | desc_fallout_fur_rev_nihss<br><br>Show the field ONLY if:<br>[fallout_review_nihss]='2' | Further Review Needed by Stroke Coordinator:<br><br>Please provide Discovery Details and Reason(s) for Further Review. | descriptive |
| 1142 | fallout_response_nihss<br><br>Show the field ONLY if:<br>[fallout_review_nihss]='1' or [fallou t_review_nihss]='2' | NIHSS Reported Fallout Response / Discovery Details: | notes |
| 1143 | fallout_ap_indiv_nihss<br><br>Show the field ONLY if:<br>[fallout_review_nihss]='1' | Fallout Action Plan - Individual: | notes |
| 1144 | fallout_ap_unit_nihss<br><br>Show the field ONLY if:<br>[fallout_review_nihss]='1' | Fallout Action Plan - Unit: | notes |

| | | | |
|---|---|---|---|
| 1145 | fallout_review_mrs<br><br>Show the field ONLY if: [calc_dnv_qm7_cr4a]='0' or [calc_dnv_qm7_cr4b]='0' or [calc_dnv_qm7_cr4c]='0' or [calc_dnv_9]='0' | Section Header: *Fallout: Modified Rankin Scores DocumentedSITE: [site] MRN: [mrn]Patient Name:[jc_lastname], [jc_firstname]Fallout Details:[fallout_detail_mrs]Fallout Unit: [fallout_unit_mrs]Date Notified:[fallout_date_mrs]Notification Sent To:[fallout_notified_mrs]*<br><br>Fallout Validation<br>*If flagged for further review, include reason in response below.Selecting Further Review Needed will trigger notification email.* | radio<br>1 Reviewed / Acknowledged<br>2 Further Review Needed |
| 1146 | desc_fallout_rev_ack_mrs<br><br>Show the field ONLY if: [fallout_review_mrs]='1' | Fallout Reviewed and Acknowledged:<br><br>Please provide Discovery Details and Action Plan(s) below. | descriptive |
| 1147 | desc_fallout_fur_rev_mrs<br><br>Show the field ONLY if: [fallout_review_mrs]='2' | Further Review Needed by Stroke Coordinator:<br><br>Please provide Discovery Details and Reason(s) for Further Review. | descriptive |
| 1148 | fallout_response_mrs<br><br>Show the field ONLY if: [fallout_review_mrs]='1' or [fallout_review_mrs]='2' | Modified Rankin Scores Documented Fallout Response / Discovery Details: | notes |
| 1149 | fallout_ap_indiv_mrs<br><br>Show the field ONLY if: [fallout_review_mrs]='1' | Fallout Action Plan - Individual: | notes |
| 1150 | fallout_ap_unit_mrs<br><br>Show the field ONLY if: [fallout_review_mrs]='1' | Fallout Action Plan - Unit: | notes |
| 1151 | fallout_review_sevscore<br><br>Show the field ONLY if: [calc_dnv_12]='0' | Section Header: *Fallout: Initial Severity Scores DocumentedSITE: [site] MRN: [mrn]Patient Name:[jc_lastname], [jc_firstname]Fallout Details:[fallout_detail_sevscore]Fallout Unit: [fallout_unit_sevscore]Date Notified:[fallout_date_sevscore]Notification Sent To: [fallout_notified_sevscore]*<br><br>Fallout Validation<br>*If flagged for further review, include reason in response below.Selecting Further Review Needed will trigger notification email.* | radio<br>1 Reviewed / Acknowledged<br>2 Further Review Needed |
| 1152 | desc_fallout_rev_ack_sevscore<br><br>Show the field ONLY if: [fallout_review_sevscore]='1' | Fallout Reviewed and Acknowledged:<br><br>Please provide Discovery Details and Action Plan(s) below. | descriptive |
| 1153 | desc_fallout_fur_rev_sevscore<br><br>Show the field ONLY if: [fallout_review_sevscore]='2' | Further Review Needed by Stroke Coordinator:<br><br>Please provide Discovery Details and Reason(s) for Further Review. | descriptive |
| 1154 | fallout_response_sevscore<br><br>Show the field ONLY if: [fallout_review_sevscore]='1' or [fallout_review_sevscore]='2' | Initial Severity Scores Documented Fallout Response / Discovery Details: | notes |

| | | | |
|---|---|---|---|
| 1155 | fallout_ap_indiv_sevscore<br><br>Show the field ONLY if:<br>[fallout_review_sevscore]='1' | Fallout Action Plan - Individual: | notes |
| 1156 | fallout_ap_unit_sevscore<br><br>Show the field ONLY if:<br>[fallout_review_sevscore]='1' | Fallout Action Plan - Unit: | notes |
| 1157 | fallout_review_fib_edu<br><br>Show the field ONLY if:<br>[calc_fib_edu]='0' | Section Header: *Fallout: Fibrinolytic Therapy EducationSITE: [site] MRN: [mrn]Patient Name:[jc_lastname], [jc_firstname]Fallout Details:[fallout_detail_fib_edu]Fallout Unit: [fallout_unit_fib_edu]Date Notified:[fallout_date_fib_edu]Notification Sent To: [fallout_notified_fib_edu]*<br><br>Fallout Validation<br>*If flagged for further review, include reason in response below.Selecting Further Review Needed will trigger notification email.* | radio<br>1 Reviewed / Acknowledged<br>2 Further Review Needed |
| 1158 | desc_fallout_rev_ack_fib_edu<br><br>Show the field ONLY if:<br>[fallout_review_fib_edu]='1' | Fallout Reviewed and Acknowledged:<br><br>Please provide Discovery Details and Action Plan(s) below. | descriptive |
| 1159 | desc_fallout_fur_rev_fib_edu<br><br>Show the field ONLY if:<br>[fallout_review_fib_edu]='2' | Further Review Needed by Stroke Coordinator:<br><br>Please provide Discovery Details and Reason(s) for Further Review. | descriptive |
| 1160 | fallout_response_fib_edu<br><br>Show the field ONLY if:<br>[fallout_review_fib_edu]='1' or [fallout_review_fib_edu]='2' | Fibrinolytic Therapy Education Fallout Response / Discovery Details: | notes |
| 1161 | fallout_ap_indiv_fib_edu<br><br>Show the field ONLY if:<br>[fallout_review_fib_edu]='1' | Fallout Action Plan - Individual: | notes |
| 1162 | fallout_ap_unit_fib_edu<br><br>Show the field ONLY if:<br>[fallout_review_fib_edu]='1' | Fallout Action Plan - Unit: | notes |
| 1163 | fallout_review_thro_edu<br><br>Show the field ONLY if:<br>[calc_thro_edu]='0' | Section Header: *Fallout: Arteriogram EducationSITE: [site] MRN: [mrn]Patient Name: [jc_lastname], [jc_firstname]Fallout Details:[fallout_detail_thro_edu]Fallout Unit: [fallout_unit_thro_edu]Date Notified:[fallout_date_thro_edu]Notification Sent To: [fallout_notified_thro_edu]*<br><br>Fallout Validation<br>*If flagged for further review, include reason in response below.Selecting Further Review Needed will trigger notification email.* | radio<br>1 Reviewed / Acknowledged<br>2 Further Review Needed |
| 1164 | desc_fallout_rev_ack_thro_edu<br><br>Show the field ONLY if:<br>[fallout_review_thro_edu]='1' | Fallout Reviewed and Acknowledged:<br><br>Please provide Discovery Details and Action Plan(s) below. | descriptive |

| | | | |
|---|---|---|---|
| 1165 | desc_fallout_fur_rev_thro_edu<br><br>Show the field ONLY if:<br>[fallout_review_thro_edu]='2' | Further Review Needed by Stroke Coordinator:<br><br>Please provide Discovery Details and Reason(s) for Further Review. | descriptive |
| 1166 | fallout_response_thro_edu<br><br>Show the field ONLY if:<br>[fallout_review_thro_edu]='1' or [fallout_review_thro_edu]='2' | Arteriogram Education Fallout Response / Discovery Details: | notes |
| 1167 | fallout_ap_indiv_thro_edu<br><br>Show the field ONLY if:<br>[fallout_review_thro_edu]='1' | Fallout Action Plan - Individual: | notes |
| 1168 | fallout_ap_unit_thro_edu<br><br>Show the field ONLY if:<br>[fallout_review_thro_edu]='1' | Fallout Action Plan - Unit: | notes |
| 1169 | fallout_review_smoking<br><br>Show the field ONLY if:<br>[calc_gwtg_smoking_cess]='0' | Section Header: *Fallout: Smoking CessationSITE: [site] MRN: [mrn]Patient Name: [jc_lastname], [jc_firstname]Fallout Details:[fallout_detail_smoking]Fallout Unit: [fallout_unit_smoking]Date Notified:[fallout_date_smoking]Notification Sent To: [fallout_notified_smoking]*<br><br>Fallout Validation<br>*If flagged for further review, include reason in response below.Selecting Further Review Needed will trigger notification email.* | radio<br>1 Reviewed / Acknowledged<br>2 Further Review Needed |
| 1170 | desc_fallout_rev_ack_smoking<br><br>Show the field ONLY if:<br>[fallout_review_smoking]='1' | Fallout Reviewed and Acknowledged:<br><br>Please provide Discovery Details and Action Plan(s) below. | descriptive |
| 1171 | desc_fallout_fur_rev_smoking<br><br>Show the field ONLY if:<br>[fallout_review_smoking]='2' | Further Review Needed by Stroke Coordinator:<br><br>Please provide Discovery Details and Reason(s) for Further Review. | descriptive |
| 1172 | fallout_response_smoking<br><br>Show the field ONLY if:<br>[fallout_review_smoking]='1' or [fallout_review_smoking]='2' | Smoking Cessation Fallout Response / Discovery Details: | notes |
| 1173 | fallout_ap_indiv_smoking<br><br>Show the field ONLY if:<br>[fallout_review_smoking]='1' | Fallout Action Plan - Individual: | notes |
| 1174 | fallout_ap_unit_smoking<br><br>Show the field ONLY if:<br>[fallout_review_smoking]='1' | Fallout Action Plan - Unit: | notes |

| | | | |
|---|---|---|---|
| 1175 | fallout_review_ldl<br><br>Show the field ONLY if:<br>[calc_gwtg_ldl_documented]='0' | Section Header: *Fallout: LDL DocumentedSITE: [site] MRN: [mrn]Patient Name: [jc_lastname], [jc_firstname]Fallout Details:[fallout_detail_ldl]Fallout Unit: [fallout_unit_ldl]Date Notified:[fallout_date_ldl]Notification Sent To:[fallout_notified_ldl]*<br><br>Fallout Validation<br>*If flagged for further review, include reason in response below.Selecting Further Review Needed will trigger notification email.* | radio<br>1 Reviewed / Acknowledged<br>2 Further Review Needed |
| 1176 | desc_fallout_rev_ack_ldl<br><br>Show the field ONLY if:<br>[fallout_review_ldl]='1' | Fallout Reviewed and Acknowledged:<br><br>Please provide Discovery Details and Action Plan(s) below. | descriptive |
| 1177 | desc_fallout_fur_rev_ldl<br><br>Show the field ONLY if:<br>[fallout_review_ldl]='2' | Further Review Needed by Stroke Coordinator:<br><br>Please provide Discovery Details and Reason(s) for Further Review. | descriptive |
| 1178 | fallout_response_ldl<br><br>Show the field ONLY if:<br>[fallout_review_ldl]='1' or [fallout_review_ldl]='2' | LDL Documented Fallout Response / Discovery Details: | notes |
| 1179 | fallout_ap_indiv_ldl<br><br>Show the field ONLY if:<br>[fallout_review_ldl]='1' | Fallout Action Plan - Individual: | notes |
| 1180 | fallout_ap_unit_ldl<br><br>Show the field ONLY if:<br>[fallout_review_ldl]='1' | Fallout Action Plan - Unit: | notes |
| 1181 | fallout_review_ist<br><br>Show the field ONLY if:<br>[calc_gwtg_intensive_statin]='0' | Section Header: *Fallout: Intensive Statin TherapySITE: [site] MRN: [mrn]Patient Name: [jc_lastname], [jc_firstname]Fallout Details:[fallout_detail_ist]Fallout Unit: [fallout_unit_ist]Date Notified:[fallout_date_ist]Notification Sent To:[fallout_notified_ist]*<br><br>Fallout Validation<br>*If flagged for further review, include reason in response below.Selecting Further Review Needed will trigger notification email.* | radio<br>1 Reviewed / Acknowledged<br>2 Further Review Needed |
| 1182 | desc_fallout_fur_rev_ist<br><br>Show the field ONLY if:<br>[fallout_review_ist]='2' | Further Review Needed by Stroke Coordinator:<br><br>Please provide Discovery Details and Reason(s) for Further Review. | descriptive |
| 1183 | fallout_response_ist<br><br>Show the field ONLY if:<br>[fallout_review_ist]='1' or [fallout_review_ist]='2' | Intensive Statin Therapy Fallout Response / Discovery Details: | notes |

| | | | |
|---|---|---|---|
| 1184 | fallout_review_cva_poc<br><br>Show the field ONLY if:<br>[calc_cva_poc_fallout] = '0' | Section Header: *Fallout: CVA Plan of CareSITE: [site] MRN: [mrn]Patient Name: [jc_lastname], [jc_firstname]Fallout Details:[cva_poc_notes]Date Notified: [fallout_date_cva_poc]Notification Sent To:[fallout_email_cva_poc_1] [fallout_email_cva_poc_2] [fallout_email_cva_poc_3] [fallout_email_cva_poc_4] [fallout_email_cva_poc_5] Was the CVA Plan of Care Initiated within 24 hours of admission?[cva_poc_init]Was the CVA Plan of Care signed and dated by all RNs?[cva_poc_signed]CVA Plan of Care Compliance (Reviewed and Updated Daily)[calc_cva_poc]Was Stroke Education on Risk Factors initiated on the CVA POC?[cva_poc_risk_fact_init]Were all appropriate patient specific risk factors selected for stroke education on the CVA POC?[cva_poc_risk_fact_ident]*<br><br>Fallout Validation<br>*If flagged for further review, include reason in response below.Selecting Further Review Needed will trigger notification email.* | radio<br>1 \| Reviewed / Acknowledged<br>2 \| Further Review Needed |
| 1185 | desc_fallout_rev_ack_cva_poc<br><br>Show the field ONLY if:<br>[fallout_review_cva_poc]='1' | Fallout Reviewed and Acknowledged:<br><br>Please provide Discovery Details and Action Plan(s) below. | descriptive |
| 1186 | desc_fallout_fur_rev_cva_poc<br><br>Show the field ONLY if:<br>[fallout_review_cva_poc]='2' | Further Review Needed by Stroke Coordinator:<br><br>Please provide Discovery Details and Reason(s) for Further Review. | descriptive |
| 1187 | fallout_response_cva_poc<br><br>Show the field ONLY if:<br>[fallout_review_cva_poc]='1' or [fallout_review_cva_poc]='2' | CVA Plan of Care Fallout Response / Discovery Details: | notes |
| 1188 | fallout_ap_indiv_cva_poc<br><br>Show the field ONLY if:<br>[fallout_review_cva_poc]='1' | Fallout Action Plan - Individual: | notes |
| 1189 | fallout_ap_unit_cva_poc<br><br>Show the field ONLY if:<br>[fallout_review_cva_poc]='1' | Fallout Action Plan - Unit: | notes |
| 1190 | desc_cva_poc_fr_ind_need<br><br>Show the field ONLY if:<br>[calc_cva_poc_fallout] = '0' and [cva_poc_sample]='1' | Question below will only be answered if selected for patient sample: Were individual needs identified for the patient based on their condition and co-morbidities on any POC?[cva_poc_indiv_needs_ident] | descriptive |
| 1191 | desc_cva_poc_fr_dn<br><br>Show the field ONLY if:<br>[calc_cva_poc_fallout] = '0' and [cva_poc_indiv_needs_ident]='0' | Date Notified: Individual Needs Identified[fallout_date_cva_poc_2] | descriptive |
| 1192 | fallout_review_cva_poc_2<br><br>Show the field ONLY if:<br>[calc_cva_poc_fallout] = '0' and [cva_poc_indiv_needs_ident]='0' | Fallout Validation: Individual Needs Identified<br>*If flagged for further review, include reason in response below.Selecting Further Review Needed will trigger notification email.* | radio<br>1 \| Reviewed / Acknowledged<br>2 \| Further Review Needed |

| | | | |
|---|---|---|---|
| 1193 | desc_fallout_rev_ack_cva_poc_2<br><br>Show the field ONLY if:<br>[fallout_review_cva_poc_2]='1' | Fallout Reviewed and Acknowledged:<br><br>Please provide Discovery Details and Action Plan(s) below. | descriptive |
| 1194 | desc_fallout_fur_rev_cva_poc_2<br><br>Show the field ONLY if:<br>[fallout_review_cva_poc_2]='2' | Further Review Needed by Stroke Coordinator:<br><br>Please provide Discovery Details and Reason(s) for Further Review. | descriptive |
| 1195 | fallout_response_cva_poc_2<br><br>Show the field ONLY if:<br>[fallout_review_cva_poc_2]='1' or [fallout_review_cva_poc_2]='2' | Individual Needs Identified Fallout Response / Discovery Details: | notes |
| 1196 | fallout_ap_indiv_cva_poc_2<br><br>Show the field ONLY if:<br>[fallout_review_cva_poc_2]='1' | Fallout Action Plan - Individual: | notes |
| 1197 | fallout_ap_unit_cva_poc_2<br><br>Show the field ONLY if:<br>[fallout_review_cva_poc_2]='1' | Fallout Action Plan - Unit: | notes |
| 1198 | desc_fo_rev_post_tpa_bp<br><br>Show the field ONLY if:<br>[site]='SMCH' and [calc_post_tpa_monitoring]='0' | Section Header: *Fallout/Feedback: Post IV Thrombolytic MonitoringSite: [site] MRN: [mrn]Patient Name:[jc_lastname], [jc_firstname]Fallout Details:[tpa_tool_notes]Date Notified:[fallout_date_post_tpa]Notification Sent To:[fallout_notified_post_tpa] Neuro Vital Signs Numerator/Denominator:[tpa_tool_nvs_num]/[tpa_tool_nvs_den]Vital Signs Numerator/Denominator:[tpa_tool_vs_num]/[tpa_tool_vs_den]Clinician Notification Numerator/Denominator:[tpa_tool_md_num]/[tpa_tool_md_den]Post tPA Hand Off Numerator/Denominator:[tpa_tool_handoff_num]/[tpa_tool_handoff_den]Post IV tPA Monitoring Compliance[calc_post_tpa_compliance]%*<br><br>ANY BP > 180/105 during monitoring? (SMCH)[post_tpa_bp]ALL BPs > 180/105 addressed appropriately? (SMCH)[post_tpa_bp_addressed] | descriptive |
| 1199 | fallout_review_post_tpa<br><br>Show the field ONLY if:<br>(([site]='SMCH' or [site]='SMCW') and [calc_post_tpa_monitoring]='0') or ([site]='SMCA' and ([treatment_type(IVtPA)]='1' or [gs_ivthroinit]='1' or [gs_ivtpaoutside]='1') and [tpa_tool_notes]<>'') | Fallout Validation<br>*If flagged for further review, include reason in response below.Selecting Further Review Needed will trigger notification email.* | radio<br>1 Reviewed / Acknowledged<br>2 Further Review Needed |
| 1200 | desc_fallout_rev_ack_post_tpa<br><br>Show the field ONLY if:<br>[fallout_review_post_tpa]='1' | Fallout Reviewed and Acknowledged:<br><br>Please provide Discovery Details and Action Plan(s) below. | descriptive |
| 1201 | desc_fallout_fur_rev_post_tpa<br><br>Show the field ONLY if:<br>[fallout_review_post_tpa]='2' | Further Review Needed by Stroke Coordinator:<br><br>Please provide Discovery Details and Reason(s) for Further Review. | descriptive |

| | | | |
|---|---|---|---|
| 1202 | fallout_response_post_tpa<br><br>Show the field ONLY if: [fallout_review_post_tpa]='1' or [fallout_review_post_tpa]='2' | Post IV Thrombolytic Monitoring Fallout Response / Discovery Details: | notes |
| 1203 | fallout_ap_indiv_post_tpa<br><br>Show the field ONLY if: [fallout_review_post_tpa]='1' | Fallout Action Plan - Individual: | notes |
| 1204 | fallout_ap_unit_post_tpa<br><br>Show the field ONLY if: [fallout_review_post_tpa]='1' | Fallout Action Plan - Unit: | notes |
| 1205 | fallout_rev_info_ev_proc_1<br><br>Show the field ONLY if: [ev_proc_protocol_1]='0' or [ev_proc_ir_1]='0' or [ev_proc_transit_1]='0' or ([calc_ev_proc_assessment_1]<>'' and [calc_ev_proc_assessment_1]<>100) or ([ev_proc_type_1]='1' and [calc_post_tpa_compliance]<>'' and [calc_post_tpa_compliance]<>100) or ([site]='SMCA' and ([ev_proc_assessment_notes]<>'' or [tpa_tool_notes]<>'')) | Section Header: *Fallout: Post IR AssessmentsSite: [site] MRN: [mrn]Patient Name: [jc_lastname], [jc_firstname]Fallout Details:[ev_proc_assessment_notes]*<br><br>1st Procedure: [ev_proc_type_1] Were Groin & Pedal Pulse Assessment Orders present? [ev_proc_orders_1] If no orders were present, was unit specific protocol followed? [ev_proc_protocol_1] Procedure End Time: [ev_proc_end_1] Time patient left IR Suite: [ev_proc_depart_1] Were all appropriate assessments done in IR Suite? [ev_proc_ir_1] Patient destination: [ev_proc_dest_1] Destination Arrival Time: [ev_proc_arrival_1] Were all appropriate assessments documented between IR Suite and patient destination? [ev_proc_transit_1] Groin & Pedal Pulse Assessment Compliance for this procedure: [calc_ev_proc_assessment_1]% Date Notified: [fallout_date_ev_proc_1] Notification Sent To: [fallout_notified_ev_proc_1] | descriptive |
| 1206 | fallout_rev_info_ev_proc_vs_1<br><br>Show the field ONLY if: (([ev_proc_protocol_1]='0' or [ev_proc_ir_1]='0' or [ev_proc_transit_1]='0' or ([calc_ev_proc_assessment_1]<>'' and [calc_ev_proc_assessment_1]<>100)) and ([ev_proc_type_1]='1' and [calc_post_tpa_compliance]<>'' and [calc_post_tpa_compliance]<>100)) or ([site]='SMCA' and [ev_proc_type_1]='1' and ([ev_proc_assessment_notes]<>'' and [tpa_tool_notes]<>'')) | Neuro Checks Numerator/Denominator: [tpa_tool_nvs_num]/[tpa_tool_nvs_den] Vital Signs Numerator/Denominator: [tpa_tool_vs_num]/[tpa_tool_vs_den] Post IR Neuro Check/Vital Signs Monitoring Compliance: [calc_post_tpa_compliance]% Post Thrombolytic, Thrombectomy, and/or Endovascular Therapy Monitoring notes. [tpa_tool_notes] | descriptive |

| | | | |
|---|---|---|---|
| 1207 | fallout_review_ev_proc_1<br><br>Show the field ONLY if: [ev_proc_protocol_1]='0' or [ev_proc_ir_1]='0' or [ev_proc_transit_1]='0' or ([calc_ev_proc_assessment_1]<>'' and [calc_ev_proc_assessment_1]<>100) or ([ev_proc_type_1]='1' and [calc_post_tpa_compliance]<>'' and [calc_post_tpa_compliance]<>100) or ([site]='SMCA' and ([ev_proc_assessment_notes]<>'' or [tpa_tool_notes]<>'')) | Fallout Validation<br>*If flagged for further review, include reason in response below.Selecting Further Review Needed will trigger notification email.* | radio<br>1 \| Reviewed / Acknowledged<br>2 \| Further Review Needed |
| 1208 | desc_fallout_rev_ack_ev_proc_1<br><br>Show the field ONLY if: [fallout_review_ev_proc_1]='1' | Fallout Reviewed and Acknowledged:<br><br>Please provide Discovery Details and Action Plan(s) below. | descriptive |
| 1209 | desc_fallout_fur_rev_ev_proc_1<br><br>Show the field ONLY if: [fallout_review_ev_proc_1]='2' | Further Review Needed by Stroke Coordinator:<br><br>Please provide Discovery Details and Reason(s) for Further Review. | descriptive |
| 1210 | fallout_response_ev_proc_1<br><br>Show the field ONLY if: [fallout_review_ev_proc_1]='1' or [fallout_review_ev_proc_1]='2' | Post IR Assessments Fallout Response / Discovery Details: | notes |
| 1211 | fallout_ap_indiv_ev_proc_1<br><br>Show the field ONLY if: [fallout_review_ev_proc_1]='1' | Fallout Action Plan - Individual: | notes |
| 1212 | fallout_ap_unit_ev_proc_1<br><br>Show the field ONLY if: [fallout_review_ev_proc_1]='1' | Fallout Action Plan - Unit: | notes |
| 1213 | fallout_rev_info_ev_proc_2<br><br>Show the field ONLY if: [ev_proc_protocol_2]='0' or [ev_proc_ir_2]='0' or [ev_proc_transit_2]='0' or ([calc_ev_proc_assessment_2]<>'' and [calc_ev_proc_assessment_2]<>100) or ([ev_proc_type_2]='1' and [calc_post_tpa_compliance]<>'' and [calc_post_tpa_compliance]<>100) or ([site]='SMCA' and [ev_proc_type_2]<>'' and ([ev_proc_assessment_notes]<>'' or [tpa_tool_notes]<>'')) | 2nd Procedure: [ev_proc_type_2] Were Groin & Pedal Pulse Assessment Orders present? [ev_proc_orders_2] If no orders were present, was unit specific protocol followed? [ev_proc_protocol_2] Procedure End Time: [ev_proc_end_2] Time patient left IR Suite: [ev_proc_depart_2] Were all appropriate assessments done in IR Suite? [ev_proc_ir_2] Patient destination: [ev_proc_dest_2] Destination Arrival Time: [ev_proc_arrival_2] Were all appropriate assessments documented between IR Suite and patient destination? [ev_proc_transit_2] Groin & Pedal Pulse Assessment Compliance for this procedure: [calc_ev_proc_assessment_2]% Date Notified: [fallout_date_ev_proc_2] Notification Sent To: [fallout_notified_ev_proc_2] | descriptive |

| | | | |
|---|---|---|---|
| 1214 | fallout_rev_info_ev_proc_vs_2<br><br>Show the field ONLY if: (([ev_proc_protocol_2]='0' or [ev_proc_ir_2]='0' or [ev_proc_transit_2]='0' or ([calc_ev_proc_assessment_2]<>'' and [calc_ev_proc_assessment_2]<>100)) and ([ev_proc_type_2]='1' and [calc_post_tpa_compliance]<>'' and [calc_post_tpa_compliance]<>100)) or ([site]='SMCA' and [ev_proc_type_2]='1' and ([ev_proc_assessment_notes]<>'' and [tpa_tool_notes]<>'')) | Neuro Checks Numerator/Denominator: [tpa_tool_nvs_num]/[tpa_tool_nvs_den] Vital Signs Numerator/Denominator: [tpa_tool_vs_num]/[tpa_tool_vs_den] Post IR Neuro Check/Vital Signs Monitoring Compliance: [calc_post_tpa_compliance]% Post Thrombolytic, Thrombectomy, and/or Endovascular Therapy Monitoring notes. [tpa_tool_notes] | descriptive |
| 1215 | fallout_review_ev_proc_2<br><br>Show the field ONLY if: [ev_proc_protocol_2]='0' or [ev_proc_ir_2]='0' or [ev_proc_transit_2]='0' or ([calc_ev_proc_assessment_2]<>'' and [calc_ev_proc_assessment_2]<>100) or ([ev_proc_type_2]='1' and [calc_post_tpa_compliance]<>'' and [calc_post_tpa_compliance]<>100) or ([site]='SMCA' and [ev_proc_type_2]<>'' and ([ev_proc_assessment_notes]<>'' or [tpa_tool_notes]<>'')) | Fallout Validation<br>*If flagged for further review, include reason in response below.Selecting Further Review Needed will trigger notification email.* | radio<br>1 Reviewed / Acknowledged<br>2 Further Review Needed |
| 1216 | desc_fallout_rev_ack_ev_proc_2<br><br>Show the field ONLY if: [fallout_review_ev_proc_2]='1' | Fallout Reviewed and Acknowledged:<br><br>Please provide Discovery Details and Action Plan(s) below. | descriptive |
| 1217 | desc_fallout_fur_rev_ev_proc_2<br><br>Show the field ONLY if: [fallout_review_ev_proc_2]='2' | Further Review Needed by Stroke Coordinator:<br><br>Please provide Discovery Details and Reason(s) for Further Review. | descriptive |
| 1218 | fallout_response_ev_proc_2<br><br>Show the field ONLY if: [fallout_review_ev_proc_2]='1' or [fallout_review_ev_proc_2]='2' | Post IR Assessments Fallout Response / Discovery Details: | notes |
| 1219 | fallout_ap_indiv_ev_proc_2<br><br>Show the field ONLY if: [fallout_review_ev_proc_2]='1' | Fallout Action Plan - Individual: | notes |

| | | | |
|---|---|---|---|
| 1220 | fallout_ap_unit_ev_proc_2<br><br>Show the field ONLY if:<br>[fallout_review_ev_proc_2]='1' | Fallout Action Plan - Unit: | notes |
| 1221 | fallout_rev_info_ev_proc_3<br><br>Show the field ONLY if:<br>[ev_proc_protocol_3]='0' or [ev_proc_ir_3]='0' or [ev_proc_transit_3]='0' or ([calc_ev_proc_assessment_3]<>'' and [calc_ev_proc_assessment_3]<>100) or ([ev_proc_type_3]='1' and [calc_post_tpa_compliance]<>'' and [calc_post_tpa_compliance]<>100) or ([site]='SMCA' and [ev_proc_type_3]<>'' and ([ev_proc_assessment_notes]<>'' or [tpa_tool_notes]<>'')) | 3rd Procedure: [ev_proc_type_3] Were Groin & Pedal Pulse Assessment Orders present? [ev_proc_orders_3] If no orders were present, was unit specific protocol followed? [ev_proc_protocol_3] Procedure End Time: [ev_proc_end_3] Time patient left IR Suite: [ev_proc_depart_3] Were all appropriate assessments done in IR Suite? [ev_proc_ir_3] Patient destination: [ev_proc_dest_3] Destination Arrival Time: [ev_proc_arrival_3] Were all appropriate assessments documented between IR Suite and patient destination? [ev_proc_transit_3] Groin & Pedal Pulse Assessment Compliance for this procedure: [calc_ev_proc_assessment_3]% Date Notified: [fallout_date_ev_proc_3] Notification Sent To: [fallout_notified_ev_proc_3] | descriptive |
| 1222 | fallout_rev_info_ev_proc_vs_3<br><br>Show the field ONLY if:<br>(([ev_proc_protocol_3]='0' or [ev_proc_ir_3]='0' or [ev_proc_transit_3]='0' or ([calc_ev_proc_assessment_3]<>'' and [calc_ev_proc_assessment_3]<>100)) and ([ev_proc_type_3]='1' and [calc_post_tpa_compliance]<>'' and [calc_post_tpa_compliance]<>100)) or ([site]='SMCA' and [ev_proc_type_3]='1' and ([ev_proc_assessment_notes]<>'' and [tpa_tool_notes]<>'')) | Neuro Checks Numerator/Denominator: [tpa_tool_nvs_num]/[tpa_tool_nvs_den] Vital Signs Numerator/Denominator: [tpa_tool_vs_num]/[tpa_tool_vs_den] Post IR Neuro Check/Vital Signs Monitoring Compliance: [calc_post_tpa_compliance]% Post Thrombolytic, Thrombectomy, and/or Endovascular Therapy Monitoring notes. [tpa_tool_notes] | descriptive |
| 1223 | fallout_review_ev_proc_3<br><br>Show the field ONLY if:<br>[ev_proc_protocol_3]='0' or [ev_proc_ir_3]='0' or [ev_proc_transit_3]='0' or ([calc_ev_proc_assessment_3]<>'' and [calc_ev_proc_assessment_3]<>100) or ([ev_proc_type_3]='1' and [calc_post_tpa_compliance]<>'' and [calc_post_tpa_compliance]<>100) or ([site]='SMCA' and [ev_proc_type_3]<>'' and ([ev_proc_assessment_notes]<>'' or [tpa_tool_notes]<>'')) | Fallout Validation<br>*If flagged for further review, include reason in response below.Selecting Further Review Needed will trigger notification email.* | radio<br>1 Reviewed / Acknowledged<br>2 Further Review Needed |

| | | | |
|---|---|---|---|
| 1224 | desc_fallout_rev_ack_ev_proc_3<br><br>Show the field ONLY if:<br>[fallout_review_ev_proc_3]='1' | Fallout Reviewed and Acknowledged:<br><br>Please provide Discovery Details and Action Plan(s) below. | descriptive |
| 1225 | desc_fallout_fur_rev_ev_proc_3<br><br>Show the field ONLY if:<br>[fallout_review_ev_proc_3]='2' | Further Review Needed by Stroke Coordinator:<br><br>Please provide Discovery Details and Reason(s) for Further Review. | descriptive |
| 1226 | fallout_response_ev_proc_3<br><br>Show the field ONLY if:<br>[fallout_review_ev_proc_3]='1' or [fallout_review_ev_proc_3]='2' | Post IR Assessments Fallout Response / Discovery Details: | notes |
| 1227 | fallout_ap_indiv_ev_proc_3<br><br>Show the field ONLY if:<br>[fallout_review_ev_proc_3]='1' | Fallout Action Plan - Individual: | notes |
| 1228 | fallout_ap_unit_ev_proc_3<br><br>Show the field ONLY if:<br>[fallout_review_ev_proc_3]='1' | Fallout Action Plan - Unit: | notes |
| 1229 | fallout_rev_info_ev_proc_4<br><br>Show the field ONLY if:<br>[ev_proc_protocol_4]='0' or [ev_proc_ir_4]='0' or [ev_proc_transit_4]='0' or ([calc_ev_proc_assessment_4]<>'' and [calc_ev_proc_assessment_4]<>100) or ([ev_proc_type_4]='1' and [calc_post_tpa_compliance]<>'' and [calc_post_tpa_compliance]<>100) or ([site]='SMCA' and [ev_proc_type_4]<>'' and ([ev_proc_assessment_notes]<>'' or [tpa_tool_notes]<>'')) | 4th Procedure: [ev_proc_type_4] Were Groin & Pedal Pulse Assessment Orders present? [ev_proc_orders_4] If no orders were present, was unit specific protocol followed? [ev_proc_protocol_4] Procedure End Time: [ev_proc_end_4] Time patient left IR Suite: [ev_proc_depart_4] Were all appropriate assessments done in IR Suite? [ev_proc_ir_4] Patient destination: [ev_proc_dest_4] Destination Arrival Time: [ev_proc_arrival_4] Were all appropriate assessments documented between IR Suite and patient destination? [ev_proc_transit_4] Groin & Pedal Pulse Assessment Compliance for this procedure: [calc_ev_proc_assessment_4]% Date Notified: [fallout_date_ev_proc_4] Notification Sent To: [fallout_notified_ev_proc_4] | descriptive |

| | | | |
|---|---|---|---|
| 1230 | fallout_rev_info_ev_proc_vs_4<br><br>Show the field ONLY if: (([ev_proc_protocol_4]='0' or [ev_proc_ir_4]='0' or [ev_proc_transit_4]='0' or ([calc_ev_proc_assessment_4]<>'' and [calc_ev_proc_assessment_4]<>100)) and ([ev_proc_type_4]='1' and [calc_post_tpa_compliance]<>'' and [calc_post_tpa_compliance]<>100)) or ([site]='SMCA' and [ev_proc_type_4]='1' and ([ev_proc_assessment_notes]<>'' and [tpa_tool_notes]<>'')) | Neuro Checks Numerator/Denominator: [tpa_tool_nvs_num]/[tpa_tool_nvs_den] Vital Signs Numerator/Denominator: [tpa_tool_vs_num]/[tpa_tool_vs_den] Post IR Neuro Check/Vital Signs Monitoring Compliance: [calc_post_tpa_compliance]% Post Thrombolytic, Thrombectomy, and/or Endovascular Therapy Monitoring notes. [tpa_tool_notes] | descriptive |
| 1231 | fallout_review_ev_proc_4<br><br>Show the field ONLY if: [ev_proc_protocol_4]='0' or [ev_proc_ir_4]='0' or [ev_proc_transit_4]='0' or ([calc_ev_proc_assessment_4]<>'' and [calc_ev_proc_assessment_4]<>100) or ([ev_proc_type_4]='1' and [calc_post_tpa_compliance]<>'' and [calc_post_tpa_compliance]<>100) or ([site]='SMCA' and [ev_proc_type_4]<>'' and ([ev_proc_assessment_notes]<>'' or [tpa_tool_notes]<>'')) | Fallout Validation<br>*If flagged for further review, include reason in response below.Selecting Further Review Needed will trigger notification email.* | radio<br>1 Reviewed / Acknowledged<br>2 Further Review Needed |
| 1232 | desc_fallout_rev_ack_ev_proc_4<br><br>Show the field ONLY if: [fallout_review_ev_proc_4]='1' | Fallout Reviewed and Acknowledged:<br><br>Please provide Discovery Details and Action Plan(s) below. | descriptive |
| 1233 | desc_fallout_fur_rev_ev_proc_4<br><br>Show the field ONLY if: [fallout_review_ev_proc_4]='2' | Further Review Needed by Stroke Coordinator:<br><br>Please provide Discovery Details and Reason(s) for Further Review. | descriptive |
| 1234 | fallout_response_ev_proc_4<br><br>Show the field ONLY if: [fallout_review_ev_proc_4]='1' or [fallout_review_ev_proc_4]='2' | Post IR Assessments Fallout Response / Discovery Details: | notes |
| 1235 | fallout_ap_indiv_ev_proc_4<br><br>Show the field ONLY if: [fallout_review_ev_proc_4]='1' | Fallout Action Plan - Individual: | notes |

| | | | |
|---|---|---|---|
| 1236 | fallout_ap_unit_ev_proc_4<br><br>Show the field ONLY if:<br>[fallout_review_ev_proc_4]='1' | Fallout Action Plan - Unit: | notes |
| 1237 | fallout_review_complete | Section Header: *Form Status*<br><br>Complete? | dropdown<br>0 Incomplete<br>1 Unverified<br>2 Complete |

Instrument: **Communication Log** (communication_log)

Collapse

| | | | |
|---|---|---|---|
| 1238 | desc_comm_log | Communication Log:SITE: [site]<br>MRN: [mrn]Patient Name:[jc_lastname], [jc_firstname]Link to Issues Log:Open Issues Log | descriptive |
| 1239 | comm_shazam | Communication Log Shazam | descriptive |
| 1240 | comm_method1 | Method of Communication: | dropdown<br>1 Email<br>2 Phone<br>3 Tiger Text |
| 1241 | comm_name1 | Communication Sent To (Name): | text |
| 1242 | comm_date1 | Communication Date: | text (date_mdy) |
| 1243 | comm_role1 | Role: | radio<br>1 RN<br>2 MD<br><br>Custom alignment: RH |
| 1244 | comm_type1 | Communication Type: | dropdown<br>4 Coding Query<br>1 Documentation Request<br>2 Fallout Clarification<br>3 Stroke Education |
| 1245 | comm_detail1 | Communication Detail: | notes |
| 1246 | comm_status1 | Communication Status<br>*Enter Earliest Due Date below.* | dropdown<br>1 Follow-up Needed<br>2 Resolved<br>3 Unresolved |

| | | | |
|---|---|---|---|
| 1247 | comm_method2<br><br>Show the field ONLY if:<br>[comm_method1]<>'' | Method of Communication: | dropdown<br>1 Email<br>2 Phone<br>3 Tiger Text |
| 1248 | comm_name2<br><br>Show the field ONLY if:<br>[comm_method1]<>'' | Communication Sent To (Name): | text |
| 1249 | comm_date2<br><br>Show the field ONLY if:<br>[comm_method1]<>'' | Communication Date: | text (date_mdy) |
| 1250 | comm_role2<br><br>Show the field ONLY if:<br>[comm_method1]<>'' | Role: | radio<br>1 RN<br>2 MD<br><br>Custom alignment: RH |
| 1251 | comm_type2<br><br>Show the field ONLY if:<br>[comm_method1]<>'' | Communication Type: | dropdown<br>4 Coding Query<br>1 Documentation Request<br>2 Fallout Clarification<br>3 Stroke Education |
| 1252 | comm_detail2<br><br>Show the field ONLY if:<br>[comm_method1]<>'' | Communication Detail: | notes |
| 1253 | comm_status2<br><br>Show the field ONLY if:<br>[comm_method1]<>'' | Communication Status<br>*Enter Earliest Due Date below.* | dropdown<br>1 Follow-up Needed<br>2 Resolved<br>3 Unresolved |
| 1254 | comm_method3<br><br>Show the field ONLY if:<br>[comm_method2]<>'' | Method of Communication: | dropdown<br>1 Email<br>2 Phone<br>3 Tiger Text |
| 1255 | comm_name3<br><br>Show the field ONLY if:<br>[comm_method2]<>'' | Communication Sent To (Name): | text |

| | | | |
|---|---|---|---|
| 1256 | comm_date3<br><br>Show the field ONLY if:<br>[comm_method2]<>'' | Communication Date: | text (date_mdy) |
| 1257 | comm_role3<br><br>Show the field ONLY if:<br>[comm_method2]<>'' | Role: | radio<br>1 RN<br>2 MD<br><br>Custom alignment: RH |
| 1258 | comm_type3<br><br>Show the field ONLY if:<br>[comm_method2]<>'' | Communication Type: | dropdown<br>4 Coding Query<br>1 Documentation Request<br>2 Fallout Clarification<br>3 Stroke Education |
| 1259 | comm_detail3<br><br>Show the field ONLY if:<br>[comm_method2]<>'' | Communication Detail: | notes |
| 1260 | comm_status3<br><br>Show the field ONLY if:<br>[comm_method2]<>'' | Communication Status<br>*Enter Earliest Due Date below.* | dropdown<br>1 Follow-up Needed<br>2 Resolved<br>3 Unresolved |
| 1261 | comm_due<br><br>Show the field ONLY if:<br>[comm_status1] = '1' or [comm_status2] = '1' or [comm_status3] = '1' | Follow-up Due By: | text (date_mdy) |
| 1262 | communication_log_complete | Section Header: *Form Status*<br><br>Complete? | dropdown<br>0 Incomplete<br>1 Unverified<br>2 Complete |

Instrument: **IRR** (irr)

Collapse

| | | | |
|---|---|---|---|
| 1263 | irr_isi | Initial Stroke Information Re-abstraction agreement? | yesno<br>1 Yes<br>0 No<br><br>Custom alignment: RH |
| 1264 | irr_isi_exp<br><br>Show the field ONLY if:<br>[irr_isi] = '0' | Explain differences between abstractions in Initial Stroke Data instrument? | notes |
| 1265 | irr_adm | Admin Re-abstraction agreement? | yesno<br>1 Yes<br>0 No<br><br>Custom alignment: RH |
| 1266 | irr_adm_exp<br><br>Show the field ONLY if:<br>[irr_adm] = '0' | Explain differences between abstractions in Admin instrument? | notes |
| 1267 | irr_cc | Clinical Codes Re-abstraction agreement? | yesno<br>1 Yes<br>0 No<br><br>Custom alignment: RH |
| 1268 | irr_cc_exp<br><br>Show the field ONLY if:<br>[irr_cc] = '0' | Explain differences between abstractions in Clinical Codes instrument? | notes |
| 1269 | irr_admission<br><br>Show the field ONLY if:<br>[admission_complete]='2' | Admission Re-abstraction agreement? | yesno<br>1 Yes<br>0 No<br><br>Custom alignment: RH |
| 1270 | irr_admission_exp<br><br>Show the field ONLY if:<br>[irr_admission] = '0' | Explain differences between abstractions in Admission instrument? | notes |

| | | | |
|---|---|---|---|
| 1271 | irr_hosp<br><br>Show the field ONLY if:<br>[hospitalization_complete]='2' | Hospitalization Re-abstraction agreement? | yesno<br>1 Yes<br>0 No<br><br>Custom alignment: RH |
| 1272 | irr_hosp_exp<br><br>Show the field ONLY if:<br>[irr_hosp] = '0' | Explain differences between abstractions in Hospitalization instrument? | notes |
| 1273 | irr_asc<br><br>Show the field ONLY if:<br>[advanced_stroke_care_complete]<br>='2' | Advance Strone Care Re-abstraction agreement? | yesno<br>1 Yes<br>0 No<br><br>Custom alignment: RH |
| 1274 | irr_asc_exp<br><br>Show the field ONLY if:<br>[irr_asc] = '0' | Explain differences between abstractions in Advance Stroke Care instrument? | notes |
| 1275 | irr_asi<br><br>Show the field ONLY if:<br>[additional_stroke_information_co<br>mplete]='2' | Additional Stroke Information Re-abstraction agreement? | yesno<br>1 Yes<br>0 No<br><br>Custom alignment: RH |
| 1276 | irr_asi_exp<br><br>Show the field ONLY if:<br>[irr_asi] = '0' | Explain differences between abstractions in Additional Stroke Information instrument? | notes |
| 1277 | irr_opt<br><br>Show the field ONLY if:<br>[optional_complete]='2' | Optional Re-abstraction agreement? | yesno<br>1 Yes<br>0 No<br><br>Custom alignment: RH |
| 1278 | irr_opt_exp<br><br>Show the field ONLY if:<br>[irr_opt] = '0' | Explain differences between abstractions in Optional instrument? | notes |

| | | | |
|---|---|---|---|
| 1279 | irr_spec<br><br>Show the field ONLY if:<br>[special_initiatives_complete]='2' | Special Initiatives Re-abstraction agreement? | yesno<br>1 Yes<br>0 No<br><br>Custom alignment: RH |
| 1280 | irr_spec_exp<br><br>Show the field ONLY if:<br>[irr_spec] = '0' | Explain differences between abstractions in Special Initiatives instrument? | notes |
| 1281 | irr_dis<br><br>Show the field ONLY if:<br>[discharge_complete]='2' | Discharge Re-abstraction agreement? | yesno<br>1 Yes<br>0 No<br><br>Custom alignment: RH |
| 1282 | irr_dis_exp<br><br>Show the field ONLY if:<br>[irr_dis] = '0' | Explain differences between abstractions in Discharge instrument? | notes |
| 1283 | irr_complete | Section Header: *Form Status*<br><br>Complete? | dropdown<br>0 Incomplete<br>1 Unverified<br>2 Complete |
| Instrument: **Code Stroke** (code_stroke) | | | Collapse |
| 1284 | cs_activation_dt | Code Stroke Initial Activation Date/Time: | text (datetime_mdy, Min: 2010-01-01 00:00, Max: 2099-12-31 23:59), Identifier<br>Custom alignment: RH |
| 1285 | cs_notification_dt | Date/Time Code Stroke Notification sent to Stroke Team: | text (datetime_mdy, Min: 2010-01-01 00:00, Max: 2099-12-31 23:59), Identifier<br>Custom alignment: RH |
| 1286 | calc_cs_notification_time | Minutes from Code Stroke Activation to Notification | calc<br>Calculation: datediff([cs_activation_dt], [cs_notification_dt], "m", "mdy", true) |
| 1287 | cs_notification_delay<br><br>Show the field ONLY if:<br>[calc_cs_notification_time] >= 5 | Reason for Code Stroke Notification Delay<br>(Time from Code Stroke Activation to Notification ≥ 5 minutes.) | text<br>Custom alignment: RH |

| | | | |
|---|---|---|---|
| 1288 | stroke_alert_activation | Who activated 324-TIME? | dropdown<br>1 CRT<br>2 ED<br>3 EMS<br>4 Other<br><br>Custom alignment: RH |
| 1289 | cs_trans_neurologist | Transfer to Neurologist? | yesno<br>1 Yes<br>0 No<br><br>Custom alignment: RH |
| 1290 | desc_cs_neurologist_name<br>Show the field ONLY if:<br>[cs_neurologist_name]<>'' or [cs_neuro_notified_dt]<>'' | Name of Neurologist Notified:[cs_neurologist_name]Time Neurologist Notified:[cs_neuro_notified_dt]From: Code Stroke | descriptive |
| 1291 | cs_neurologist_name | Name of Neurologist Notified: | text<br>Custom alignment: RH |
| 1292 | cs_ptinfo_provided | Age, LKW, Deficits provided? | yesno<br>1 Yes<br>0 No<br><br>Custom alignment: RH |
| 1293 | cs_notes | Code Stroke Notes: | notes<br>Custom alignment: RH |
| 1294 | cs_dsch_dx | Discharge Diagnosis Code<br>*[jc_princicd10diagnosis] from Clinical Codes* | text<br>Custom alignment: RH |
| 1295 | cs_dsch_dx_desc | Discharge Diagnosis Description | text<br>Custom alignment: RH |
| 1296 | cs_secondary_dx | Secondary Stroke Diagnosis Code<br>*[jc_othericd10diag] from Clinical Codes* | text<br>Custom alignment: RH |
| 1297 | cs_secondary_dx_desc | Secondary Stroke Diagnosis Description | text<br>Custom alignment: RH |

| | | | |
|---|---|---|---|
| 1298 | code_stroke_complete | Section Header: *Form Status*<br>Complete? | dropdown<br>0 Incomplete<br>1 Unverified<br>2 Complete |
| Instrument: **Lab Data** (lab_data) | | | Collapse |
| 1299 | lab_collection_dt<br><br>Show the field ONLY if:<br>[gs_jp_labna] <> '1' | Section Header: *Initial Stroke Information:SITE: [site] MRN: [mrn]Patient Name: [jc_lastname], [jc_firstname]Date/Time Lab Tests Ordered:[gs_jp_laborddatetime]Date/Time Lab Tests Completed:[gs_jp_labcpdatetime]Labs Ordered to Labs Completed: [calc_order_to_labcp] minutes*<br>Blood Collection Date/Time | text (datetime_mdy, Min: 2010-01-01 00:00, Max: 2099-12-31 23:59), Identifier |
| 1300 | calc_door_to_lab_collect<br><br>Show the field ONLY if:<br>[gs_jp_labna] <> '1' | Door to Labs Collected | calc<br>Calculation: if([gs_symptomlocation]='4',datediff([gs_symptomdatetime], [lab_collection_dt], "m", "mdy", true), datediff([jc_arrdatetime], [lab_collection_dt], "m", "mdy", true))<br>Field Annotation: @HIDDEN |
| 1301 | calc_order_to_lab_collect<br><br>Show the field ONLY if:<br>[gs_jp_labna] <> '1' | Labs Ordered to Labs Collected | calc<br>Calculation: datediff([gs_jp_laborddatetime], [lab_collection_dt], "m", "mdy", true) |
| 1302 | lab_received_dt<br><br>Show the field ONLY if:<br>[gs_jp_labna] <> '1' | Lab Specimen Received Date/Time | text (datetime_mdy, Min: 2010-01-01 00:00, Max: 2099-12-31 23:59), Identifier |
| 1303 | calc_labs_collect_to_received<br><br>Show the field ONLY if:<br>[gs_jp_labna] <> '1' | Labs Collected to Labs Received Time | calc<br>Calculation: datediff([lab_collection_dt], [lab_received_dt], "m", "mdy", true) |
| 1304 | calc_lab_results<br><br>Show the field ONLY if:<br>[gs_jp_labna] <> '1' | Labs Received to Completed Time | calc<br>Calculation: datediff([lab_received_dt], [gs_jp_labcpdatetime], "m", "mdy", true) |
| 1305 | lab_delay_reason<br><br>Show the field ONLY if:<br>[gs_jp_labna] <> '1' and [calc_order_to_labcp] > 45 | Reason for delay.<br>(Labs Ordered to Labs Completed over 45 minutes) | notes |
| 1306 | lab_pi_comments<br><br>Show the field ONLY if:<br>[gs_jp_labna] <> '1' | Process Improvement Comments | notes |

| | | | |
|---|---|---|---|
| 1307 | lab_data_complete | Section Header: *Form Status*<br>Complete? | dropdown<br>0 Incomplete<br>1 Unverified<br>2 Complete |
| Instrument: **Imaging Data** (imaging_data) Collapse | | | |
| 1308 | img_complete_dt<br>Show the field ONLY if:<br>[gs_jp_ctna] <> '1' | Section Header: *Initial Stroke Information:SITE: [site] MRN: [mrn]Patient Name: [jc_lastname], [jc_firstname]Initial brain imaging type completed?[imaging_type]Date/Time Brain Imaging Ordered:[gs_jp_ctorddatetime]Date/Time Brain Imaging Initiated: [gs_ctcompdatetime]Door to Imaging Initiated:[calc_door_to_ctcomp] minutesDate/Time Brain Imaging Interpreted:[gs_jp_ctrepdatetime]Door to Imaging Interpreted: [calc_door_to_ctrep] minutes*<br>Study Complete<br>*Date/Time Imaging Completed* | text (datetime_mdy, Min: 2010-01-01 00:00, Max: 2099-12-31 23:59), Identifier |
| 1309 | calc_img_complete_to_rep<br>Show the field ONLY if:<br>[gs_jp_ctna] <> '1' | Study Interpretation Time<br>(Study Complete to Results Called)<br>*Time in Minutes* | calc<br>Calculation: datediff([img_complete_dt], [gs_jp_ctrepdatetime], "m", "mdy", true) |
| 1310 | radiologist_notified<br>Show the field ONLY if:<br>[gs_jp_ctna] <> '1' | Radiologist Notified<br>*Date/Time Radiologist Notified* | text (datetime_mdy, Min: 2010-01-01 00:00, Max: 2099-12-31 23:59), Identifier |
| 1311 | calc_radiologist_to_rep<br>Show the field ONLY if:<br>[gs_jp_ctna] <> '1' | Radiologist Read Time<br>(Radiologist Notified to Results Called)<br>*Time in Minutes* | calc<br>Calculation: datediff([radiologist_notified], [gs_jp_ctrepdatetime], "m", "mdy", true) |
| 1312 | img_delay_reason<br>Show the field ONLY if:<br>[gs_jp_ctna] <> '1' and [calc_door_to_ctrep] > 45 | Reason for delay.<br>(Door to Imaging Interpreted over 45 minutes) | notes |
| 1313 | img_pi_comments<br>Show the field ONLY if:<br>[gs_jp_ctna] <> '1' | Process Improvement Comments | notes |
| 1314 | imaging_data_complete | Section Header: *Form Status*<br>Complete? | dropdown<br>0 Incomplete<br>1 Unverified<br>2 Complete |
| Instrument: **IR/Cath Lab Data** (ircath_lab_data) Collapse | | | |

| | | | |
|---|---|---|---|
| 1315 | ir_after_hours<br><br>Show the field ONLY if:<br>[treatment_type(MER)]='1' | Section Header: *Initial Stroke Information: SITE: [site] MRN: [mrn] Patient Name: [jc_lastname], [jc_firstname] Arrival Date/Time: [jc_arrdatetime] Skin Puncture Date/Time: [artpunc_dt] Door to Groin: [calc_door_to_artpunc] minutes Endovascular Decision to Treat Date/Time: [mer_decision]*<br><br>After hours / Weekend | radio<br>1 \| After Hours / Weekend |
| 1316 | ir_team_activated_dt<br><br>Show the field ONLY if:<br>[treatment_type(MER)]='1' | IR/Cath Lab Team Activation Date/Time | text (datetime_mdy, Min: 2010-01-01 00:00, Max: 2099-12-31 23:59)<br>5 \| Date & Time (MM/DD/YYYY HH24:MM)<br>3 \| Date Only (MM/DD/YYYY)<br>0 \| Unknown |
| 1317 | calc_ir_decision_to_activ<br><br>Show the field ONLY if:<br>[treatment_type(MER)]='1' | Decision to IR/Cath Lab Team Activation<br>*Time in Minutes* | calc<br>Calculation: datediff([mer_decision], [ir_team_activated_dt], "m", "mdy", true) |
| 1318 | ir_team_arrival_dt<br><br>Show the field ONLY if:<br>[treatment_type(MER)]='1' | IR/Cath Lab Team Arrival Date/Time | text (datetime_mdy, Min: 2010-01-01 00:00, Max: 2099-12-31 23:59)<br>5 \| Date & Time (MM/DD/YYYY HH24:MM)<br>3 \| Date Only (MM/DD/YYYY)<br>0 \| Unknown |
| 1319 | calc_ir_acti_to_arrival<br><br>Show the field ONLY if:<br>[treatment_type(MER)]='1' | IR/Cath Lab Team Activation to Arrival<br>*Time in Minutes* | calc<br>Calculation: datediff([ir_team_activated_dt], [ir_team_arrival_dt], "m", "mdy", true) |
| 1320 | ir_pt_arrival_dt<br><br>Show the field ONLY if:<br>[treatment_type(MER)]='1' | Patient Arrival to IR/Cath Lab Date/Time | text (datetime_mdy, Min: 2010-01-01 00:00, Max: 2099-12-31 23:59)<br>5 \| Date & Time (MM/DD/YYYY HH24:MM)<br>3 \| Date Only (MM/DD/YYYY)<br>0 \| Unknown |
| 1321 | calc_ir_acti_to_pt_arrival<br><br>Show the field ONLY if:<br>[treatment_type(MER)]='1' | IR/Cath Lab Team Activation to Patient Arrival<br>*Time in Minutes* | calc<br>Calculation: datediff([ir_team_activated_dt], [ir_pt_arrival_dt], "m", "mdy", true) |
| 1322 | calc_ir_arriv_to_pt_arriv<br><br>Show the field ONLY if:<br>[treatment_type(MER)]='1' | IR/Cath Lab Team Arrival to Patient Arrival<br>*Time in Minutes* | calc<br>Calculation: datediff([ir_team_arrival_dt], [ir_pt_arrival_dt], "m", "mdy", true) |
| 1323 | calc_ir_arrival_artpunc<br><br>Show the field ONLY if:<br>[treatment_type(MER)]='1' | Patient IR/Cath Lab Arrival to Skin Puncture (Groin Time)<br>*Time in Minutes - SMCA Goal: ≤ 15 Minutes* | calc<br>Calculation: datediff([ir_pt_arrival_dt], [artpunc_dt], "m", "mdy", true) |

| | | | |
|---|---|---|---|
| 1324 | ir_mer_delay_rsn<br><br>Show the field ONLY if:<br>[treatment_type(MER)]='1' and [calc_door_to_artpunc]>90 | Reasons for Delay in Door to Groin<br>(Time from Door to Groin > 90 minutes.) | checkbox<br>1 \| ir_mer_delay_rsn___1 \| Anesthesia<br>2 \| ir_mer_delay_rsn___2 \| Consenting<br>3 \| ir_mer_delay_rsn___3 \| Provider Not Available<br>4 \| ir_mer_delay_rsn___4 \| Pt transport to the procedure area<br>0 \| ir_mer_delay_rsn___0 \| Other {ir_mer_delay_rsn_oth} |
| 1325 | ir_mer_delay_rsn_oth<br><br>Show the field ONLY if:<br>[ir_mer_delay_rsn(0)]='1' | Specify Other Reasons for Delay in Door to Groin | text<br>Field Annotation: @CHARLIMIT=200 |
| 1326 | ir_pi_comments<br><br>Show the field ONLY if:<br>[treatment_type(MER)]='1' | Process Improvement Comments | notes |
| 1327 | ircath_lab_data_complete | Section Header: *Form Status*<br><br>Complete? | dropdown<br>0 \| Incomplete<br>1 \| Unverified<br>2 \| Complete |
| Instrument: **Post Discharge Followup** (post_discharge_followup) | | | Collapse |
| 1328 | lost_to_fup_30<br><br>Show the field ONLY if:<br>[postdis_mrs_scale_30]='' | Section Header: *Initial Stroke Information:SITE:[site]MRN:[mrn]Patient Name:[jc_lastname],[jc_firstname]Discharge Date:[gs_discdatetime]Days Since Stroke Onset:[calc_days_post_onset](until 90 day mRS or 365 days)Disposition:[jc_disposition]If Other Facility:[gs_othfacility]*<br><br>Lost To Follow-Up (30 Day post-discharge mRS) | radio<br>0 \| LTF |
| 1329 | lost_to_fup_90<br><br>Show the field ONLY if:<br>[fu_death]<>'1' and [postdis_mrs_scale]='' | Lost To Follow-Up (90 Day post-discharge mRS) | radio<br>0 \| LTF |
| 1330 | fu_lost | Lost To Follow-Up | radio<br>0 \| LTF<br><br>Field Annotation: @HIDDEN |
| 1331 | fu_death<br><br>Show the field ONLY if:<br>[lost_to_fup_90] = '' | Is patient deceased? | radio<br>1 \| Yes<br>2 \| No<br><br>Custom alignment: RH |

| | | | |
|---|---|---|---|
| 1332 | fu_datedeath_precision<br><br>Show the field ONLY if:<br>[fu_death] = '1' | If date of death not known, select "Unknown" | dropdown<br><table><tr><td>3</td><td>MM/DD/YYYY</td></tr><tr><td>0</td><td>Unknown</td></tr></table> |
| 1333 | fu_datedeath<br><br>Show the field ONLY if:<br>[fu_death] = '1' and [fu_datedeath_precision] = '3' | Date of Death | text (date_mdy, Min: 2010-01-01, Max: 2099-12-31), Identifier |
| 1334 | postdis_mrs_scale_30<br><br>Show the field ONLY if:<br>[lost_to_fup_30] = '' | Section Header: *30 Day mRS*<br><br>Post Discharge Modified Rankin Scale (30 day) | radio<br><table><tr><td>1</td><td>Yes</td></tr><tr><td>2</td><td>No</td></tr></table><br>Custom alignment: RH |
| 1335 | postdis_mrs_date_30<br><br>Show the field ONLY if:<br>[postdis_mrs_scale_30] = '1' | Date Post Discharge Modified Rankin Scale Performed (30 day) | text (date_mdy, Min: 2010-01-01, Max: 2099-12-31), Identifier<br>Custom alignment: RH |
| 1336 | postdis_mrs_score_30<br><br>Show the field ONLY if:<br>[postdis_mrs_scale_30] = '1' | If yes, Modified Rankin Total Score (30 day) | radio<br><table><tr><td>1</td><td>0 - No symptoms at all</td></tr><tr><td>2</td><td>1 - No significant disability despite symptoms; able to carry out all usual duties and activities</td></tr><tr><td>3</td><td>2 - Slight disability; unable to carry out all previous activities, but able to look after own affairs without assistance</td></tr><tr><td>4</td><td>3 - Moderate disability; requiring some help, but able to walk without assistance</td></tr><tr><td>5</td><td>4 - Moderately severe disability; unable to walk without assistance and unable to attend to own bodily needs without assistance</td></tr><tr><td>6</td><td>5 - Severe disability; bedridden, incontinent and requiring constant nursing care and attention</td></tr><tr><td>7</td><td>6 - Dead</td></tr></table><br>Custom alignment: LV |
| 1337 | postdis_mrs_scale<br><br>Show the field ONLY if:<br>[lost_to_fup_90] = '' | Section Header: *90 Day mRS*<br><br>Post Discharge Modified Rankin Scale (90 Day) | radio<br><table><tr><td>1</td><td>Yes</td></tr><tr><td>2</td><td>No</td></tr></table><br>Custom alignment: RH |

| | | | |
|---|---|---|---|
| 1338 | postdis_mrs_date<br><br>Show the field ONLY if:<br>[postdis_mrs_scale] = '1' | Date Post Discharge Modified Rankin Scale Performed (90 Day) | text (date_mdy, Min: 2010-01-01, Max: 2099-12-31), Identifier<br>Custom alignment: RH |
| 1339 | postdis_mrs_score<br><br>Show the field ONLY if:<br>[postdis_mrs_scale] = '1' | If yes, Modified Rankin Total Score (90 Day) | radio<br>1 \| 0 - No symptoms at all<br>2 \| 1 - No significant disability despite symptoms; able to carry out all usual duties and activities<br>3 \| 2 - Slight disability; unable to carry out all previous activities, but able to look after own affairs without assistance<br>4 \| 3 - Moderate disability; requiring some help, but able to walk without assistance<br>5 \| 4 - Moderately severe disability; unable to walk without assistance and unable to attend to own bodily needs without assistance<br>6 \| 5 - Severe disability; bedridden, incontinent and requiring constant nursing care and attention<br>7 \| 6 - Dead<br><br>Custom alignment: LV |
| 1340 | dnv_9_90daymrs | 90 Day mRS | calc<br>Calculation: [postdis_mrs_score]-1 |
| 1341 | fu_aslcds<br><br>Show the field ONLY if:<br>[postdis_mrs_score] = '2' or [postdis_mrs_score] = '3' or [postdis_mrs_score] = '4' or [postdis_mrs_score] = '5' or [postdis_mrs_score] = '6' | Austin Speech Labs Communication Disability Scale<br>**Needs caregiver assistance* | dropdown<br>0 \| 0 - No speech or language deficits.<br>1 \| 1 - Minor communication deficits, but without communication limitations.<br>2 \| 2 - Functional limitations in communication, but not ADLs.<br>3 \| 3 - Limitations in communicating needs or communication deficits limiting independence in ADLs.*<br>4 \| 4 - Requires caregiver assistance and inference for most communications of basic needs.*<br>5 \| 5 - No effective communication.* |
| 1342 | post_discharge_followup_complete | Section Header: *Form Status*<br>Complete? | dropdown<br>0 \| Incomplete<br>1 \| Unverified<br>2 \| Complete |

Instrument: **sICH Review** (sich_review)

Collapse

| | | | |
|---|---|---|---|
| 1343 | sich_results | Section Header: *sICH Information:SITE:[site]MRN:[mrn]Patient Name:[jc_lastname], [jc_firstname]IV t-PA initiated at this hospital?[gs_ivthroinit]Date/Time IV tPA initiated: [jc_ivthrodttm]IA catheter-based treatment at this hospital?[gs_iatpa]IA t-PA or MER Initiation Date/Time:[gs_iatpadatetime]Was there a positive finding on brain imaging of parenchymal hematoma, SAH, and/or IVH following IV or IA thrombolytic (t-PA) therapy, or mechanical endovascular reperfusion therapy initiation?[c_neuroimaging36h]Positive Brain Image Date/Time:[c_posbrain]Results of positive brain image: [c_posbrain_result:checked]Complications of Thrombolytic Therapy: [gs_comp_throm_ther:checked]*<br>Results of positive brain image | notes |
| 1344 | sich_deterioration | Evidence in the chart of worsening neuro status? | radio<br>1 Yes<br>0 No<br>2 UTD<br>Custom alignment: RH |
| 1345 | sich_nihss | Is there a documented NIHSS score associated with hemorrhage/worsening in neuro status? | yesno<br>1 Yes<br>0 No<br>Custom alignment: RH |
| 1346 | sich_nihss_score | If yes, NIHSS score: | text (integer, Min: 0, Max: 42)<br>Custom alignment: RH |
| 1347 | sich_phys_symptomatic | Section Header: *For physician documentation only:*<br>Is this a symptomatic ICH? | yesno<br>1 Yes<br>0 No<br>Custom alignment: RH |
| 1348 | sich_phys_category | Category | dropdown<br>HI1 HI1<br>HI2 HI2<br>PH1 PH1<br>PH2 PH2<br>Custom alignment: RH |
| 1349 | sich_phys_comments | Physician Comments | notes<br>Custom alignment: RH |

| | | | |
|---|---|---|---|
| 1350 | sich_phys_adjudicated | Adjudicated | yesno<br>1 Yes<br>0 No<br><br>Custom alignment: RH |
| 1351 | sich_review_complete | Section Header: *Form Status*<br>Complete? | dropdown<br>0 Incomplete<br>1 Unverified<br>2 Complete |
| Instrument: **M&M Review** (mm_review) | | | Collapse |
| 1352 | mm_complication_findings | Section Header: *M&M ReviewSITE:[site]MRN:[mrn]Patient Name:[jc_lastname], [jc_firstname]How patient arrived at your hospital[gs_patientarrival]Patient location when stroke symptoms discovered[gs_symptomlocation]Arrival Date/Time[jc_arrdatetime] Discharge Date and Time[gs_discdatetime]What was the patient's discharge disposition? [jc_disposition]Type of Acute Stroke Treatment received by patient: [treatment_type:checked]Other Surgical Interventions and Procedures Performed: [procedures_interventions:checked] [procedures_other]*<br>Complication Findings | notes |
| 1353 | mm_action_items | Action Items | notes |
| 1354 | mm_review_date | Review Date | text (date_mdy) |
| 1355 | mm_review_complete | Section Header: *Form Status*<br>Complete? | dropdown<br>0 Incomplete<br>1 Unverified<br>2 Complete |
| Instrument: **Measure Calculations** (measure_calculations) | | | Collapse |
| 1356 | measure_report | Measure Report | truefalse<br>1 True<br>0 False<br><br>Custom alignment: RH |

| | | | |
|---|---|---|---|
| 1357 | calc_stk_1 | Section Header: *Stroke Core MeasuresStroke (STK) National Hospital Inpatient Quality MeasuresVenous Thromboembolism (VTE) ProphylaxisSTK-1:Ischemic and hemorrhage stroke patients who received VTE prophylaxis or have documentation why no VTE prophylaxis was given the day of or the day after hospital admission.*<br>STK-1<br>*1 = Meets Measure0 = Does Not Meet MeasureBlank = Not in Measure* | calc<br>Calculation: if(([gs_stroketype]=2 or [gs_stroketype]=4 or [gs_stroketype]=5) and [gs_age]>=18 and datediff([jc_admitdate], [gs_discdatetime], "m", "mdy", true)/1440<=120 and [jc_clinical]=2 and [jc_carotid]=2 and [gs_comfortonly]<>1 and datediff([jc_admitdate], [gs_discdatetime], "m", "mdy", true)/1440>=2 and ([jc_vteprophylaxis1]=1 or [jc_vteprophylaxis2]=1 or [jc_vteprophylaxis3]=1 or [jc_vteprophylaxis4]=1 or [jc_vteprophylaxis5]=1 or [jc_vteprophylaxis6]=1 or [jc_vteprophylaxis8]=1 or [jc_vteprophylaxis9]=1 or [jc_vteprophylaxis10]=1 or [jc_vteprophylaxis7]=1) and (([jc_vteprophylaxis7]=1 and [jc_vteprophylaxis1]<>1 and [jc_vteprophylaxis2]<>1 and [jc_vteprophylaxis3]<>1 and [jc_vteprophylaxis4]<>1 and [jc_vteprophylaxis5]<>1 and [jc_vteprophylaxis6]<>1 and [jc_vteprophylaxis8]<>1 and [jc_vteprophylaxis9]<>1 and [jc_vteprophylaxis10]<>1 and ([jc_novteadmis]=1 or [jc_novteadmis]=2)) or ([jc_vteprophylaxis1]<>1 and [jc_vteprophylaxis2]<>1 and [jc_vteprophylaxis3]<>1 and [jc_vteprophylaxis5]<>1 and [jc_vteprophylaxis6]<>1 and [jc_vteprophylaxis8]<>1 and (([jc_vteprophylaxis9]=1 and ([jc_reasonoralxa]=1 or [jc_reasonoralxa]=2)) or ([jc_vteprophylaxis9]<>1 and ([jc_novteadmis]=1 or [jc_novteadmis]=2)))) or (([jc_vteprophylaxis1]=1 or [jc_vteprophylaxis2]=1 or [jc_vteprophylaxis3]=1 or [jc_vteprophylaxis5]=1 or [jc_vteprophylaxis6]=1 or [jc_vteprophylaxis8]=1) and [jc_admitdate]<=[jc_vtedate])), if((([jc_vteprophylaxis1]<>1 and [jc_vteprophylaxis2]<>1 and [jc_vteprophylaxis3]<>1 and [jc_vteprophylaxis5]<>1 and [jc_vteprophylaxis6]<>1 and [jc_vteprophylaxis8]<>1) and (([jc_vteprophylaxis7]=1 and [jc_vteprophylaxis4]<>1 and [jc_vteprophylaxis9]<>1 and [jc_vteprophylaxis10]<>1 and [jc_novteadmis]=1) or ([jc_vteprophylaxis9]<>1 and [jc_novteadmis]=1) or ([jc_vteprophylaxis9]=1 and [jc_reasonoralxa]=1))) or (([jc_vteprophylaxis1]=1 or [jc_vteprophylaxis2]=1 or [jc_vteprophylaxis3]=1 or [jc_vteprophylaxis5]=1 or [jc_vteprophylaxis6]=1 or [jc_vteprophylaxis8]=1) and datediff([jc_admitdate],[jc_vtedate], 'd', 'mdy', true)< 2), 1, 0), '') |

| | | | |
|---|---|---|---|
| 1358 | calc_stk_2 | Section Header: *Discharged on Antithrombotic TherapySTK-2:Ischemic stroke patients prescribed antithrombotic therapy at hospital discharge.*<br>STK-2<br>*1 = Meets Measure0 = Does Not Meet MeasureBlank = Not in Measure* | calc<br>Calculation: if([gs_stroketype]=2 and [gs_age]>=18 and datediff([jc_admitdate], [gs_discdatetime], "m", "mdy", true)/1440<=120 and [gs_comfortonly]=4 and [jc_clinical]=2 and [jc_carotid]=2 and ([jc_disposition]=1 or [jc_disposition]=5 or [jc_disposition]=8) and ([gs_antithromboticyesno]=1 or (([gs_antithromboticyesno]=2 or [gs_antithromboticyesno]=3) and [gs_reasonnoantithrom(13)]=0 and [gs_reasonnoantithrom(4)]=0 and [gs_reasonnoantithrom(18)]=0 and [gs_reasonnoantithrom(21)]=0 and [gs_reasonnoantithrom(8)]=0 and [gs_reasonnoantithrom(22)]=0)), if([gs_antithromboticyesno]=1, 1, 0), '') |
| 1359 | calc_stk_3 | Section Header: *Anticoagulation Therapy for Atrial Fibrillation/FlutterSTK-3:Ischemic stroke patients with atrial fibrillation/flutter who are prescribed anticoagulation therapy at hospital discharge.*<br>STK-3<br>*1 = Meets Measure0 = Does Not Meet MeasureBlank = Not in Measure* | calc<br>Calculation: if([gs_stroketype]=2 and [gs_age]>=18 and datediff([jc_admitdate], [gs_discdatetime], "m", "mdy", true)/1440<=120 and [gs_comfortonly]=4 and [jc_clinical]=2 and [jc_carotid]=2 and ([jc_disposition]=1 or [jc_disposition]=5 or [jc_disposition]=8) and ([gs_med_hist(1)]=1 or [gs_persistentafib]=1) and [gs_afibdischarge]<>3, if([gs_afibdischarge]=1, 1, 0), '') |
| 1360 | calc_stk_4 | Section Header: *Thrombolytic TherapySTK-4:Acute ischemic stroke patients who arrive at this hospital within 2 hours of time last known well and for whom IV t-PA was initiated at this hospital within 3 hours of time last known well.*<br>STK-4<br>*1 = Meets Measure0 = Does Not Meet MeasureBlank = Not in Measure* | calc<br>Calculation: if([gs_stroketype]=2 and [gs_age]>=18 and datediff([jc_admitdate], [gs_discdatetime], "m", "mdy", true)/1440<=120 and [jc_clinical]=2 and [jc_carotid]=2 and [jc_edpatient]=1 and [calc_lkw_to_arrival]>=0 and [calc_lkw_to_arrival]<=120 and (([gs_ivthroinit]=1 and (datediff([gs_lastknownwell], [jc_ivthrodttm], "m", "mdy", true)>270 or (datediff([gs_lastknownwell], [jc_ivthrodttm], "m", "mdy", true)>180 and datediff([gs_lastknownwell], [jc_ivthrodttm], "m", "mdy", true)<=270 and [gs_ivtpanc]=2) or (datediff([gs_lastknownwell], [jc_ivthrodttm], "m", "mdy", true)>=0 and datediff([gs_lastknownwell], [jc_ivthrodttm], "m", "mdy", true)<=180))) or ([gs_ivthroinit]=2 and [gs_ivtpanc]=2 and [gs_ivtpalatenc]=2)), if([gs_ivthroinit]=1 and datediff([gs_lastknownwell], [jc_ivthrodttm], "m", "mdy", true)>=0 and datediff([gs_lastknownwell], [jc_ivthrodttm], "m", "mdy", true)<=180, 1, 0), '') |

| | | | |
|---|---|---|---|
| 1361 | calc_stk_5 | Section Header: *Antithrombotic Therapy By End of Hospital Day 2STK-5:Ischemic stroke patients administered antithrombotic therapy by the end of hospital day 2.*<br>STK-5<br>*1 = Meets Measure0 = Does Not Meet MeasureBlank = Not in Measure* | calc<br>Calculation: if([gs_stroketype]=2 and [gs_age]>=18 and datediff([jc_admitdate], [gs_discdatetime], "m", "mdy", true)/1440<=120 and [jc_clinical]=2 and [jc_carotid]=2 and [gs_comfortonly]<>1 and roundup(datediff([jc_arrdatetime], [gs_discdatetime], "d", "mdy", true), 0)>=2 and [gs_ivthroinit]<>1 and [gs_iatpa]<>1 and [gs_ivtpaoutside]<>1 and [gs_iatpaoutside]<>1 and ([gs_antiplateletadm]=1 or [gs_antiplateletadm]=6), if([gs_antiplateletadm]=1, 1, 0), '') |
| 1362 | calc_stk_6 | Section Header: *Discharged on Statin MedicationSTK-6:Ischemic stroke patients who are prescribed statin medication at hospital discharge.*<br>STK-6<br>*1 = Meets Measure0 = Does Not Meet MeasureBlank = Not in Measure* | calc<br>Calculation: if([gs_stroketype]=2 and [gs_age]>=18 and datediff([jc_admitdate], [gs_discdatetime], "m", "mdy", true)/1440<=120 and [gs_comfortonly]=4 and [jc_clinical]=2 and [jc_carotid]=2 and ([jc_disposition]=1 or [jc_disposition]=5 or [jc_disposition]=8) and ([gs_cholredtx(3)]=1 or [jc_nostatindisc]=2), if([gs_cholredtx(3)]=1, 1, 0), '') |
| 1363 | calc_stk_8 | Section Header: *Stroke EducationSTK-8:Ischemic or hemorrhagic stroke patients or their caregivers who were given educational materials during the hospital stay addressing all of the following: activation of emergency medical system, need for follow-up after discharge, medications prescribed at discharge, risk factors for stroke, and warning signs and symptoms of stroke.*<br>STK-8<br>*1 = Meets Measure0 = Does Not Meet MeasureBlank = Not in Measure* | calc<br>Calculation: if(([gs_stroketype]=2 or [gs_stroketype]=4 or [gs_stroketype]=5) and [gs_age]>=18 and datediff([jc_admitdate], [gs_discdatetime], 'd', 'mdy', true)<=120 and [gs_comfortonly]=4 and [jc_clinical]=2 and [jc_carotid]=2 and ([jc_disposition]=1 or [jc_disposition]=8) and ([jc_edurisk]=1 or [jc_edurisk]=2) and ([jc_eduwarning]=1 or [jc_eduwarning]=2) and ([jc_eduems]=1 or [jc_eduems]=2) and ([jc_edufollowup]=1 or [jc_edufollowup]=2) and ([jc_edumeds]=1 or [jc_edumeds]=2), if([jc_edurisk]=1 and [jc_eduwarning]=1 and [jc_eduems]=1 and [jc_edufollowup]=1 and [jc_edumeds]=1, 1, 0), '') |
| 1364 | calc_stk_10 | Section Header: *Assessed for RehabilitationSTK-10:Ischemic or hemorrhagic stroke patients who were assessed for rehabilitation services.*<br>STK-10<br>*1 = Meets Measure0 = Does Not Meet MeasureBlank = Not in Measure* | calc<br>Calculation: if(([gs_stroketype]=2 or [gs_stroketype]=4 or [gs_stroketype]=5) and [gs_age]>=18 and datediff([jc_admitdate], [gs_discdatetime], "m", "mdy", true)/1440<=120 and [jc_clinical]=2 and [jc_carotid]=2 and ([jc_disposition]=1 or [jc_disposition]=5 or [jc_disposition]=8) and ([jc_assessrehab]=1 or [jc_assessrehab]=2), if([jc_assessrehab]=1, 1, 0), '') |
| 1365 | calc_stk_acs | Section Header: *Appropriate Care ScoreSTK-ACS:Percentage of patients that received all stroke care elements as appropriate.*<br>STK-ACS<br>*1 = Meets Measure0 = Does Not Meet MeasureBlank = Not in Measure* | calc<br>Calculation: if([calc_stk_1]=1 or [calc_stk_2]=1 or [calc_stk_3]=1 or [calc_stk_4]=1 or [calc_stk_5]=1 or [calc_stk_6]=1 or [calc_stk_8]=1 or [calc_stk_10]=1 or [calc_stk_1]=0 or [calc_stk_2]=0 or [calc_stk_3]=0 or [calc_stk_4]=0 or [calc_stk_5]=0 or [calc_stk_6]=0 or [calc_stk_8]=0 or [calc_stk_10]=0, if([calc_stk_1]<>0 and [calc_stk_2]<>0 and [calc_stk_3]<>0 and [calc_stk_4]<>0 and [calc_stk_5]<>0 and [calc_stk_6]<>0 and [calc_stk_8]<>0 and [calc_stk_10]<>0, 1, 0), '') |

| | | | |
|---|---|---|---|
| 1366 | calc_dnv_1 | Section Header: *DNV GL Comprehensive Stroke Center Metrics for Measuring Processes and QualityRequired metrics are 1, 2, 3, 4, 6, 7, 9, 12, 15, 18, 19, 27, 28, 29.DNV Metric 1: (required)Percentage of all ischemic stroke/TIA patients who have a deficit at the time of the initial RN note, ED Physician or Neurology consultation note for whom an NIHSS score is documented.*<br>DNV 1<br>*1 = In Numerator0 = In MeasureBlank = Not in Measure* | calc<br>Calculation: if(([gs_stroketype]=2 or ([gs_stroketype]=3 and [gs_symptresolve]<>1)) and [gs_notadmit]=2, if([gs_nihssperformed]=1, 1, 0), '') |
| 1367 | calc_dnv_2 | Section Header: *DNV Metric 2: (required)Percentage of ischemic stroke patients eligible for intravenous thrombolysis who receive it within the appropriate time window.*<br>DNV 2<br>*1 = In Numerator0 = In MeasureBlank = Not in Measure* | calc<br>Calculation: if([gs_stroketype]=2 and [calc_lkw_to_arrival]<=180 and ([gs_ivtpanc]<>1 or [gs_ivtpalatenc]<>1) and ([gs_ivtpaoutside]<>'1' or (rounddown(datediff([gs_discdatetime],'10-01-2018 00:00','d','mdy',true)) > 0)), if(datediff([gs_lastknownwell], [jc_ivthrodttm], 'm', 'mdy', true)>=0 and datediff([gs_lastknownwell], [jc_ivthrodttm], 'm', 'mdy', true)<=270, 1, 0), '') |
| 1368 | calc_dnv_3_45min | Section Header: *DNV Metric 3: (required)Percentage of patients who are treated for acute ischemic stroke with intravenous thrombolysis whose treatment is started within 60 minutes after arrival.*<br>DNV 3 ≤ 45 Minutes<br>*1 = In Numerator0 = In MeasureBlank = Not in Measure* | calc<br>Calculation: if([gs_stroketype]=2 and [jc_clinical]<>1 and [calc_door_to_ivtpa]>=0, if([calc_door_to_ivtpa]>=0 and [calc_door_to_ivtpa]<=45, 1, 0), '') |
| 1369 | calc_dnv_3_60min | DNV 3 ≤ 60 Minutes<br>*1 = In Numerator0 = In MeasureBlank = Not in Measure* | calc<br>Calculation: if([gs_stroketype]=2 and [jc_clinical]<>1 and [calc_door_to_ivtpa]>=0, if([calc_door_to_ivtpa]>=0 and [calc_door_to_ivtpa]<=60, 1, 0), '') |
| 1370 | calc_dnv_4 | Section Header: *DNV Metric 4: (required)Median time from arrival to the start of initial imaging workup for all patients who arrive within 8 hours of last known well.*<br>DNV 4<br>*Door to CT Initiated* | calc<br>Calculation: if([gs_stroketype]=2 and [calc_lkw_to_arrival]>=0 and [calc_lkw_to_arrival]<=480 and ([gs_notadmit]=2 or [gs_notadmitwhy]=1) and [calc_door_to_ctcomp]>=0, [calc_door_to_ctcomp], '') |
| 1371 | calc_dnv_4_8_to_24hrs | DNV 4 (8 to 24 hrs)<br>*Door to CT Initiated* | calc<br>Calculation: if([gs_stroketype]=2 and [calc_lkw_to_arrival]>480 and [calc_lkw_to_arrival]<=1440 and ([gs_notadmit]=2 or [gs_notadmitwhy]=1) and [calc_door_to_ctcomp]>=0, [calc_door_to_ctcomp], '') |
| 1372 | calc_dnv_5 | Section Header: *DNV Metric 5:Percentage of ischemic stroke patients seen within 24 hours of the time they were last known well who have documentation that an endovascular recanalization procedure either was performed, or was considered and deemed not to be appropriate or possible. A reason should be documented if an endovascular procedure was not performed.Retired as a Metric as of October 1, 2018. Now embedded in the standards MR.4 CR.6b*<br>DNV 5<br>*1 = In Numerator0 = In MeasureBlank = Not in Measure* | calc<br>Calculation: if([gs_stroketype]=2 and [calc_lkw_to_arrival]>=0 and [calc_lkw_to_arrival]<=1440 and ([gs_notadmit]=2 or [gs_notadmitwhy]=1), if([gs_iatpa]=1 or [dnv_5_statement]=1, 1, 0), '') |

| | | | |
|---|---|---|---|
| 1373 | calc_dnv_6 | Section Header: *DNV Metric 6: (required)Median time from arrival to start of treatment for acute ischemic stroke patients undergoing an endovascular intervention.*<br>DNV 6<br>*Door to Endovascular Treatment* | calc<br>Calculation: if([gs_stroketype]=2, if([firstpassdt]>0, [calc_door_to_device], if([c_firstpassdt]>0, if([doortime_evt]='2', datediff([gs_symptomdatetime], [c_firstpassdt], "m", "mdy", true), if([decision_evt]='1', datediff([worse_sympt_dt], [c_firstpassdt], "m", "mdy", true), datediff([jc_arrdatetime], [c_firstpassdt], "m", "mdy", true))), if([gs_iatpadatetime]>0, if([doortime_evt]='2', datediff([gs_symptomdatetime], [gs_iatpadatetime], "m", "mdy", true), if([decision_evt]='1', datediff([worse_sympt_dt], [gs_iatpadatetime], "m", "mdy", true), datediff([jc_arrdatetime], [gs_iatpadatetime], "m", "mdy", true))), ''))), '') |
| 1374 | calc_dnv_7 | Section Header: *DNV Metric 7: (required)Ischemic stroke patients who develop a symptomatic intracranial hemorrhage (i.e., clinical deterioration ≥ 4 point increase on NIHSS and brain image finding of parenchymal hematoma, or subarachnoid hemorrhage, or intraventricular hemorrhage) within (≤) 36 hours after the onset of treatment with intra-venous (IV) or intra-arterial (IA) thrombolytic (t-PA) therapy, or mechanical endovascular reperfusion procedure (i.e., mechanical endovascular thrombectomy with a clot retrieval device).*<br>DNV 7<br>*1 = In Numerator (sICH)0 = In Measure (No sICH)Blank = Not in Measure* | calc<br>Calculation: if([gs_stroketype]='2' and ([gs_ivthroinit]='1' or [gs_ivtpaoutside]='1' or [gs_iatpa]='1' or [gs_iatpaoutside]='1'), if([sich_phys_symptomatic]=1, 1, 0), '') |
| 1375 | calc_dnv_7_2015 | DNV 7 (From 2015 CSC 2.0 Requirements)<br>*1 = In Numerator (sICH)0 = In Measure (No sICH)Blank = Not in Measure* | calc<br>Calculation: if(([gs_ivthroinit]=1 or [gs_ivtpaoutside]=1) and [gs_notadmit]=2, if([sich_phys_symptomatic]=1, 1, 0), '') |
| 1376 | calc_dnv_7a | Section Header: *DNV Metric 7a: (required)Ischemic stroke patients who develop a symptomatic intracranial hemorrhage (i.e., clinical deterioration ≥ 4-point increase on NIHSS and brain image finding of parenchymal hematoma, or subarachnoid hemorrhage, or intraventricular hemorrhage) within (≤) 36 hours after the onset of treatment with intra-venous (IV) thrombolytic (t-PA) therapy only.*<br>DNV 7a<br>*1 = In Numerator (sICH)0 = In Measure (No sICH)Blank = Not in Measure* | calc<br>Calculation: if([gs_stroketype]='2' and ([gs_ivthroinit]='1' or [gs_ivtpaoutside]='1') and [gs_iatpa]<>'1' and [gs_iatpaoutside]<>'1', if([sich_phys_symptomatic]=1, 1, 0), '') |
| 1377 | calc_dnv_7b | Section Header: *DNV Metric 7b: (required)Ischemic stroke patients who develop a symptomatic intracranial hemorrhage (i.e., clinical deterioration ≥ 4-point increase on NIHSS and brain image finding of parenchymal hematoma, or subarachnoid hemorrhage, or intraventricular hemorrhage) within (≤) 36 hours after the onset of treatment with IA thrombolytic (t-PA) therapy or mechanical endovascular reperfusion therapy (i.e., mechanical endovascular thrombectomy with a clot retrieval device).*<br>DNV 7b<br>*1 = In Numerator (sICH)0 = In Measure (No sICH)Blank = Not in Measure* | calc<br>Calculation: if([gs_stroketype]='2' and [gs_ivthroinit]<>'1' and [gs_ivtpaoutside]<>'1' and ([gs_iatpa]='1' or [gs_iatpaoutside]='1'), if([sich_phys_symptomatic]=1, 1, 0), '') |
| 1378 | calc_dnv_8 | Section Header: *DNV Metric 8:Percentage of acute ischemic stroke patients treated with endovascular interventions that develop significant intracranial hemorrhage within 36 hours of treatment.Retired as a Metric as of October 1, 2018. Now part of Metric 7.*<br>DNV 8 tPA/MER<br>*1 = In Numerator (sICH)0 = In Measure (No sICH)Blank = Not in Measure* | calc<br>Calculation: if([gs_stroketype]=2 and ([gs_iatpa]=1 or [gs_iatpaoutside]=1) and ([gs_ivthroinit]=1 or [gs_ivtpaoutside]=1) and [gs_notadmit]=2, if([sich_phys_symptomatic]=1, 1, 0), '') |

| | | | |
|---|---|---|---|
| 1379 | calc_dnv_8b | DNV 8 MER only<br>*1 = In Numerator (sICH)0 = In Measure (No sICH)Blank = Not in Measure* | calc<br>Calculation: if([gs_stroketype]=2 and ([gs_iatpa]=1 or [gs_iatpaoutside]=1) and ([gs_ivthroinit]<>1 and [gs_ivtpaoutside]<>1) and [gs_notadmit]=2, if([sich_phys_symptomatic]=1, 1, 0), '') |
| 1380 | calc_dnv_qm7_cr4a | Section Header: *DNV Metric 9: (required)Percentage of acute ischemic stroke patients who are treated with intravenous thrombolysis or who undergo endovascular recanalization procedure for whom there is documentation of a pre-morbid, discharge and 90-day mRS score.*<br>DNV QM.7 CR.4a - Pre-Morbid Modified Rankin Score by or at discharge<br>*1 = In Numerator0 = In MeasureBlank = Not in Measure* | calc<br>Calculation: if(([gs_stroketype]=2 or [gs_stroketype]=3 or [gs_stroketype]=4 or [gs_stroketype]=5 or [gs_stroketype]=6) and [gs_notadmit]=2 and rounddown(datediff('10-01-2018 00:00',[gs_discdatetime],'d','mdy',true))>=0, if(([c_prestrokemrs]=1 or [c_prestrokemrs]=2), 1, 0), '') |
| 1381 | calc_dnv_qm7_cr4b | DNV QM.7 CR.4b - Modified Rankin Score at (or by) discharge<br>*1 = In Numerator0 = In MeasureBlank = Not in Measure* | calc<br>Calculation: if(([gs_stroketype]=2 or [gs_stroketype]=3 or [gs_stroketype]=4 or [gs_stroketype]=5 or [gs_stroketype]=6) and [gs_notadmit]=2 and rounddown(datediff('10-01-2018 00:00',[gs_discdatetime],'d','mdy',true))>=0, if(([gs_rankin]=1 or [gs_rankin]=2 or [gs_rankin]=3 or [gs_rankin]=4 or [gs_rankin]=5 or [gs_rankin]=6 or [gs_rankin]=7), 1, 0), '') |
| 1382 | calc_dnv_qm7_cr4c | DNV QM.7 CR.4c - Modified Rankin Score 90 days after onset<br>*1 = In Numerator0 = In MeasureBlank = Not in Measure* | calc<br>Calculation: if([gs_stroketype]=2 and ([gs_ivthroinit]=1 or [gs_ivtpaoutside]=1 or [gs_iatpa]=1 or [gs_iatpaoutside]=1) and [gs_notadmit]=2 and rounddown(datediff('10-01-2018 00:00',[gs_discdatetime],'d','mdy',true))>=0 and if([gs_symptomdatetime]<>'', rounddown(datediff([gs_symptomdatetime], 'today', 'd', 'mdy', true)), if([gs_lastknownwell]<>'', rounddown(datediff([gs_lastknownwell], 'today', 'd', 'mdy', true)), rounddown(datediff([jc_arrdatetime], 'today', 'd', 'mdy', true))))>75, if([lost_to_fup_90]=0 or [postdis_mrs_score]=1 or [postdis_mrs_score]=2 or [postdis_mrs_score]=3 or [postdis_mrs_score]=4 or [postdis_mrs_score]=5 or [postdis_mrs_score]=6 or [postdis_mrs_score]=7, 1, 0), '') |

| | | | |
|---|---|---|---|
| 1383 | calc_dnv_9 | DNV 9<br>*1 = In Numerator0 = In MeasureBlank = Not in Measure* | calc<br>Calculation: if([gs_stroketype]=2 and ([gs_ivthroinit]=1 or [gs_ivtpaoutside]=1 or [gs_iatpa]=1 or [gs_iatpaoutside]=1) and [gs_notadmit]=2, if(rounddown(datediff('10-01-2018 00:00',[gs_discdatetime],'d','mdy',true))>=0, if([calc_dnv_qm7_cr4a]=1 and [calc_dnv_qm7_cr4b]=1 and ([calc_dnv_qm7_cr4c]=1 or [calc_dnv_qm7_cr4c]=''), 1, 0), if(rounddown(datediff('10-01-2018 00:00', [gs_discdatetime],'d','mdy',true))< 0 and if([gs_symptomdatetime]<>'', rounddown(datediff([gs_symptomdatetime],'today','d','mdy',true) if([gs_lastknownwell]<>'', rounddown(datediff([gs_lastknownwell],'today','d','mdy',true)), rounddown(datediff([jc_arrdatetime],'today','d','mdy',true))))>75, if([lost_to_fup_90]=0 or [postdis_mrs_score]=1 or [postdis_mrs_score]=2 or [postdis_mrs_score]=3 or [postdis_mrs_score]=4 or [postdis_mrs_score]=5 or [postdis_mrs_score]=6 or [postdis_mrs_score]=7, 1, 0), '')), '') |
| 1384 | calc_dnv_10 | Section Header: *DNV Metric 10: (Optional)Percentage of patients undergoing CEA, or carotid angioplasty or stenting, with stroke or death within 30 days of the procedure.*<br>DNV 10<br>Data not currently available.<br>*1 = In Numerator0 = In MeasureBlank = Not in Measure* | calc<br>Calculation: '' |
| 1385 | calc_dnv_11 | Section Header: *DNV Metric 11: (Optional)Percentage of patients undergoing intracranial angioplasty and/or stenting for atherosclerotic disease with stroke or death within 30 days of the procedure.*<br>DNV 11<br>Data not currently available.<br>*1 = In Numerator0 = In MeasureBlank = Not in Measure* | calc<br>Calculation: '' |
| 1386 | calc_dnv_12 | Section Header: *DNV Metric 12: (required)Percentage of SAH and ICH stroke patients for whom a severity measurement is performed prior to surgical intervention or within 6 hours of hospital arrival for patients who do not undergo surgical intervention, and documented in the medical record.*<br>DNV 12<br>*1 = In Numerator0 = In MeasureBlank = Not in Measure* | calc<br>Calculation: if(([gs_stroketype]='4' or [gs_stroketype]='5') and [gs_notadmit]='2', if([gs_stroketype]='4' and ([c_sahscale]='1' or [c_wfnssah]>0), 1, if([gs_stroketype]='5' and [c_ichscore]='1', 1, 0)), '') |
| 1387 | calc_dnv_12a | Section Header: *DNV Metric 12a: (required)Percentage of SAH stroke patients for whom a severity measurement is performed prior to surgical intervention or within 6 hours of hospital arrival for patients who do not undergo surgical intervention, and documented in the medical record.*<br>DNV 12a<br>*1 = In Numerator0 = In MeasureBlank = Not in Measure* | calc<br>Calculation: if([gs_stroketype]='4' and [gs_notadmit]='2', if([c_sahscale]='1' or [c_wfnssah]>0, 1, 0), '') |
| 1388 | calc_dnv_12b | Section Header: *DNV Metric 12b: (required)Percentage of ICH stroke patients for whom a severity measurement is performed prior to surgical intervention or within 6 hours of hospital arrival for patients who do not undergo surgical intervention, and documented in the medical record.*<br>DNV 12b<br>*1 = In Numerator0 = In MeasureBlank = Not in Measure* | calc<br>Calculation: if([gs_stroketype]='5' and [gs_notadmit]='2', if([c_ichscore]='1', 1, 0), '') |

| | | | |
|---|---|---|---|
| 1389 | calc_dnv_13 | Section Header: *DNV Metric 13: (retired 08-01-2018)Median time from admission to start of procedure intended to obliterate a ruptured aneurysm by surgical clipping or endovascular coiling for patients who arrive within 48 hours of the hemorrhage that led directly to admission.*<br>DNV 13<br>*Time in Hours* | calc<br>Calculation: if([calc_lkw_to_arrival]< 2880 and [gs_stroketype]=4 and [aneurysmal_sah]=1, round(datediff([monitored_bed], [dnv_13_proc_start], "h", "mdy", true), 0), '') |
| 1390 | calc_dnv_14 | Section Header: *DNV Metric 14: (Optional)Percentage of patients with aneurysmal SAH arriving within 48 hours of hemorrhage for whom a coiling or clipping procedure was not started within 36 hours of arrival who have a documented reason for not having undergone coiling or clipping within 36 hours of arrival.*<br>DNV 14<br>*1 = In Numerator0 = In MeasureBlank = Not in Measure* | calc<br>Calculation: if([calc_lkw_to_arrival]< 2880 and [gs_stroketype]=4 and [aneurysmal_sah]=1 and (([procedures_interventions(1)]='0' and [procedures_interventions(2)]='0') or datediff([jc_arrdatetime], [dnv_13_proc_start], "h", "mdy", true)>36), if([dnv_13_excl_reason]=1, 1, 0), '') |
| 1391 | calc_dnv_15 | Section Header: *DNV Metric 15: (required)Percentage of patients with documented aneurysmal SAH for whom Nimodipine treatment (60 mg every 4 hours or 30 mg every 2 hours) is started within 24 hours of diagnosis and for whom such treatment is continued until 21 days after the hemorrhage or until discharge if they are discharged less than 21 days after the SAH.*<br>DNV 15<br>*1 = In Numerator0 = In MeasureBlank = Not in Measure* | calc<br>Calculation: if([gs_stroketype]=4 and [aneurysmal_sah]=1, if(([c_nimodipine]=1 and [dnv_15_21days]=1) or [c_reasonnonim]=1, 1, 0), '') |
| 1392 | calc_dnv_16 | Section Header: *DNV Metric 16: (Optional)Percentage of SAH patients with diminished level of consciousness and ventriculomegaly who are treated with EVD.*<br>DNV 16<br>Data not currently available.<br>*1 = In Numerator0 = In MeasureBlank = Not in Measure* | calc<br>Calculation: '' |
| 1393 | calc_dnv_17 | Section Header: *DNV Metric 17: (Optional)Median frequency of noninvasive monitoring performed for surveillance for vasospasm in patients with aneurysmal SAH during the period between 3 and 14 days after SAH.*<br>DNV 17<br>Data not currently available. | calc<br>Calculation: '' |
| 1394 | calc_dnv_18 | Section Header: *DNV Metric 18: (required)Complication rates for aneurysm coiling and clipping: Patients undergoing coiling (18a, 18b) or clipping (18c, 18d) of a ruptured (18b, 18d) or unruptured (18a, 18c) aneurysm who have complications of death, stroke or bleeds withing 24 hours of the procedure, or any re-bleeding and/or second treatment for residual aneurysm within 30 days of the procedure.*<br>DNV 18<br>*1 = In Numerator0 = In MeasureBlank = Not in Measure* | calc<br>Calculation: if(([dnv_18_complications]=0 or [dnv_18_complications]=1) and ([procedures_interventions(2)]='1' or [procedures_interventions(1)]='1'), if([dnv_18_complications]=1, 1, 0), '') |
| 1395 | calc_dnv_18a | DNV 18a<br>*1 = In Numerator0 = In MeasureBlank = Not in Measure* | calc<br>Calculation: if(([dnv_18_complications]=0 or [dnv_18_complications]=1) and [procedures_interventions(2)]='1' and [dnv_18_rupt]=2, if([dnv_18_complications]=1, 1, 0), '') |

| | | | |
|---|---|---|---|
| 1396 | calc_dnv_18b | DNV 18b<br>*1 = In Numerator0 = In MeasureBlank = Not in Measure* | calc<br>Calculation: if(([dnv_18_complications]=0 or [dnv_18_complications]=1) and [procedures_interventions(2)]='1' and [dnv_18_rupt]=1, if([dnv_18_complications]=1, 1, 0), '') |
| 1397 | calc_dnv_18c | DNV 18c<br>*1 = In Numerator0 = In MeasureBlank = Not in Measure* | calc<br>Calculation: if(([dnv_18_complications]=0 or [dnv_18_complications]=1) and [procedures_interventions(1)]='1' and [dnv_18_rupt]=2, if([dnv_18_complications]=1, 1, 0), '') |
| 1398 | calc_dnv_18d | DNV 18d<br>*1 = In Numerator0 = In MeasureBlank = Not in Measure* | calc<br>Calculation: if(([dnv_18_complications]=0 or [dnv_18_complications]=1) and [procedures_interventions(1)]='1' and [dnv_18_rupt]=1, if([dnv_18_complications]=1, 1, 0), '') |
| 1399 | calc_dnv_18_bleeds | DNV 18 Bleeding Complications<br>*1 = Pre-procedural2 = Procedural3 = Post-proceduralBlank = Not in Measure* | calc<br>Calculation: if([dnv_18_bleed]=1 or [dnv_18_bleed]=2 or [dnv_18_bleed]=3, if([dnv_18_bleed]=1, 1, if([dnv_18_bleed]=2, 2, if([dnv_18_bleed]=3, 3, 0))), '') |
| 1400 | calc_dnv_19 | Section Header: *DNV Metric 19: (required)Percentage of intracerebral hemorrhage (ICH) stroke patients with an INR value > 1.4 at hospital arrival who are treated with a procoagulant reversal agent (i.e., fresh frozen plasma, recombinant factor VIIa, prothrombin complex concentrates).*<br>DNV 19<br>*1 = In Numerator0 = In MeasureBlank = Not in Measure* | calc<br>Calculation: if([gs_stroketype]=5 and [gs_notadmit]='2' and ([gs_inr]>1.4 or [c_inrperf]='1') and [c_reasnopro]<>'1' and [gs_comfortonly]='4', if([c_procoagulant]='1', 1, 0), '') |
| 1401 | calc_dnv_19_2015 | DNV 19 (From 2015 CSC 2.0 Requirements)<br>*Time in Minutes* | calc<br>Calculation: if([gs_stroketype]=5 and [gs_inr]>1.4 and [c_procoagulant_dt]>0 and datediff([jc_arrdatetime], [c_procoagulant_dt], "m", "mdy", true)>=0, datediff([jc_arrdatetime], [c_procoagulant_dt], "m", "mdy", true), '') |
| 1402 | calc_dnv_20 | Section Header: *DNV Metric 20: (Optional)Percentage of patients undergoing surgical or endovascular treatment of an AVM with stroke or death within 30 days of the procedure.*<br>DNV 20<br>Data not currently available.<br>*1 = In Numerator0 = In MeasureBlank = Not in Measure* | calc<br>Calculation: '' |
| 1403 | calc_dnv_21 | Section Header: *DNV Metric 21: (Optional)Percentage of patients with ischemic or hemorrhagic stroke or TIA transferred from another hospital to the CSC with documentation of the time from the first call from the transferring hospital to the CSC (to a member of a stroke program or to a centralized transfer center) to arrival time at the CSC.*<br>DNV 21<br>*1 = In Numerator0 = In MeasureBlank = Not in Measure* | calc<br>Calculation: if(([gs_stroketype]=2 or [gs_stroketype]=3 or [gs_stroketype]=4 or [gs_stroketype]=5) and [gs_patientarrival]=3, if([transfer_firstcall]>0 and [jc_arrdatetime]>0, 1, 0), '') |

| | | | |
|---|---|---|---|
| 1404 | calc_dnv_22 | Section Header: *DNV Metric 22: (Optional)Percentage of patients admitted to each type of unit to which patients with ischemic or hemorrhagic stroke or TIA are initially admitted (e.g. neurological/neurosurgical ICU, medical ICU, surgical ICU, general ICU, coronary care unit, burn ICU, stroke unit, other intermediate-level-of-care unit, neurology floor, or other floor). A separate percentage should be calculated for each type of unit.*<br>DNV 22<br>Data not currently available.<br>*1 = In Numerator0 = In MeasureBlank = Not in Measure* | calc<br>Calculation: '' |
| 1405 | calc_dnv_23 | Section Header: *DNV Metric 23: (retired 08-01-2018)Percentage of patients with stroke or death within 24 hours of diagnostic neuroangiography.*<br>DNV 23<br>*1 = In Numerator0 = In MeasureBlank = Not in Measure* | calc<br>Calculation: if([diagnostic_tests(NeuroAngio)]=1, [dnv_23], '') |
| 1406 | calc_dnv_24 | Section Header: *DNV Metric 24: (Optional)Percentage of patients who have a diagnosis of ischemic or hemorrhagic stroke who undergo EVD and then develop ventriculitis.*<br>DNV 24<br>Data not currently available.<br>*1 = In Numerator0 = In MeasureBlank = Not in Measure* | calc<br>Calculation: '' |
| 1407 | calc_dnv_25 | Section Header: *DNV Metric 25: (Optional)Median number of days from admission to completion of evaluations for physical therapy, occupational therapy, speech-language pathology, and rehabilitation medicine, unless there is documentation on admission that some or all of these evaluations are not needed or that the patient cannot tolerate them because of medical instability.*<br>DNV 25<br>Data not currently available.<br>*Number of Days* | calc<br>Calculation: '' |
| 1408 | calc_dnv_26 | Section Header: *DNV Metric 26: (Optional)Percentage of patients admitted with diagnoses of ischemic stroke, SAH, AVM, intracranial hemorrhage, extracranial cervical stenosis, intracranial stenosis, or TIA who are enrolled in a clinical research study.*<br>DNV 26<br>*1 = In Numerator0 = In MeasureBlank = Not in Measure* | calc<br>Calculation: if(([gs_stroketype]=2 or [gs_stroketype]=3 or [gs_stroketype]=4 or [gs_stroketype]=5) and [gs_notadmit]=2, if([jc_clinical]=1, 1, 0), '') |
| 1409 | calc_dnv_27 | Section Header: *DNV Metric 27: (required)Percentage of ischemic stroke patients with a post-treatment reperfusion grade of TICI 2B or higher in the vascular territory beyond the target arterial occlusion at the end of treatment with intra-arterial (IA) thrombolytic (t-PA) therapy and/or mechanical endovascular reperfusion therapy.*<br>DNV 27<br>*1 = In Numerator0 = In MeasureBlank = Not in Measure* | calc<br>Calculation: if([gs_stroketype]='2' and [gs_iatpa]='1' and [gs_notadmit]='2', if([c_ticigrade]='4' or [c_ticigrade]='5' or [mer_ticidt]<>'' or [ticidt]<>'', 1, 0), '') |
| 1410 | calc_dnv_28 | Section Header: *DNV Metric 28: (required)Percentage of ischemic stroke patients with a large vessel cerebral occlusion (i.e., internal carotid artery (ICA) or ICA terminus (T-lesion; T-occlusion), middle cerebral artery (MCA) M1 or M2, basilar artery) who receive mechanical endovascular reperfusion (MER) therapy (time of first pass or deployment of device) within 120 minutes (>/= 0 min. and*<br>DNV 28<br>*1 = In Numerator0 = In MeasureBlank = Not in Measure* | calc<br>Calculation: if([gs_stroketype]='2' and [gs_notadmit]='2' and [gs_symptomlocation]<>'4' and [mer_targles]='1' and ([c_firstpass]='1' or [firstpassdt]<>'' or [c_firstpassdt]<>''), if(([ticidt]<>'' and [calc_door_to_reperfusion]<=150) or ([mer_ticidt]<>'' and datediff([jc_arrdatetime], [mer_ticidt], "m", "mdy", true)<=150), 1, 0), '') |

| | | | |
|---|---|---|---|
| 1411 | calc_dnv_29 | Section Header: *DNV Metric 29: (required)Percentage of ischemic stroke patients with a large vessel cerebral occlusion (i.e., internal carotid artery (ICA) or ICA terminus (T-lesion; T-occlusion), middle cerebral artery (MCA) M1 or M2, basilar artery) who receive mechanical endovascular reperfusion (MER) therapy (time of first pass or deployment of device) and achieve TICI 2B or higher less than (*<br>DNV 29<br>*1 = In Numerator0 = In MeasureBlank = Not in Measure* | calc<br>Calculation: if([gs_stroketype]='2' and [gs_notadmit]=2 and [mer_targles]='1' and ([c_firstpass]='1' or [firstpassdt]<>'' or [c_firstpassdt]<>''), if(([ticidt]<>'' and [artpunc_dt]<>'' and [calc_puncture_to_reperfusion]<=60) or ([mer_ticidt]<>'' and [c_artpuncdt]<>'' and datediff([c_artpuncdt], [mer_ticidt], "m", "mdy", true)<=60), 1, 0), '') |
| 1412 | calc_gwtg_arrive2_treat3 | Section Header: *GWTG-Stroke Achievement MeasuresTarget: ≥ 85% on all (Bronze: 90 days; Silver: 12 consecutive months; Gold: 24 consecutive months)IV rt-PA Arrive by 2 Hours, Treat by 3 Hours:Percent of acute ischemic stroke patients who arrive at the hospital within 120 minutes (2 hours) of time last known well and for whom IV t-PA was initiated at this hospital within 180 minutes (3 hours) of time last known well.*<br>IV rt-PA Arrive by 2 Hours, Treat by 3 Hours<br>*1 = Meets Measure0 = Does Not Meet MeasureBlank = Not in measure* | calc<br>Calculation: if([gs_stroketype]=2 and [gs_age]>=18 and [gs_symptomlocation]<>4 and [gs_ivtpaoutside]<>1 and [calc_lkw_to_arrival]>=0 and [calc_lkw_to_arrival]<=120 and ([gs_ivtpanc]<>1 or ([gs_exclusion(1)]<>1 and [gs_exclusion(2)]<>1 and [gs_exclusion(3)]<>1 and [gs_exclusion(4)]<>1 and [gs_exclusion(5)]<>1 and [gs_exclusion(6)]<>1 and [gs_exclusion(7)]<>1 and [gs_exclusion(8)]<>1 and [gs_exclusion(9)]<>1 and [gs_relexclusion(1)]<>1 and [gs_relexclusion(2)]<>1 and [gs_relexclusion(3)]<>1 and [gs_relexclusion(4)]<>1 and [gs_relexclusion(5)]<>1 and [gs_relexclusion(6)]<>1 and [gs_relexclusion(7)]<>1 and [gs_relexclusion(8)]<>1 and [gs_relexclusion(9)]<>1 and [gs_relexclusion(10)]<>1 and [gs_relexclusion(11)]<>1)) and [gs_lastknownwell_precision]<>3 and [jc_arrdatetime_precision]<>3 and ([gs_ivthroinit]<>1 or ([jc_ivthrodttm_precision]=5 and [jc_ivthrodttm]<>'')) and [jc_clinical]<>1 and [jc_carotid]<>1, if([gs_ivthroinit]=1 and datediff([gs_lastknownwell], [jc_ivthrodttm], "m", "mdy", true) <=180, 1, 0), '') |
| 1413 | calc_gwtg_early_antithrom | Section Header: *Early Antithrombotics:Percent of patients with ischemic stroke or TIA who receive antithrombotic therapy by the end of hospital day two.*<br>Early Antithrombotics<br>*1 = Meets Measure0 = Does Not Meet MeasureBlank = Not in measure* | calc<br>Calculation: if(([gs_stroketype]=2 or [gs_stroketype]=3) and [gs_age]>=18 and [gs_symptomlocation]<>4 and [gs_comfortonly]<>1 and roundup(datediff([jc_arrdatetime], [gs_discdatetime], "d", "mdy", true), 0)>=2 and [gs_antiplateletadm]<>5 and [gs_ivthroinit]<>1 and [gs_ivtpaoutside]<>1 and [gs_iatpa]<>1 and [gs_iatpaoutside] <>1 and [gs_notadmit]<>1 and [jc_clinical]<>1 and [jc_carotid] <>1, if([gs_antiplateletadm]=1, 1, 0), '') |

| | | | |
|---|---|---|---|
| 1414 | calc_gwtg_vte_prophylaxis | Section Header: *VTE Prophylaxis:Percent of patients with an ischemic stroke, or a hemorrhagic stroke, or stroke not otherwise specified who receive VTE prophylaxis the day of or the day after hospital admission.*<br>VTE Prophylaxis<br>*1 = Meets Measure0 = Does Not Meet MeasureBlank = Not in measure* | calc<br>Calculation: if(([gs_stroketype]=2 or [gs_stroketype]=4 or [gs_stroketype]=5 or [gs_stroketype]=6) and [gs_age]>=18 and [gs_symptomlocation]<>4 and [gs_comfortonly]<>1 and datediff([jc_admitdate], [gs_discdatetime], "m", "mdy", true)/1440>=2 and [jc_admitdate]<=[jc_vtedate] and [gs_notadmit]<>1 and [jc_clinical]<>1 and [jc_carotid]<>1, if(((([jc_vteprophylaxis1]=1 or [jc_vteprophylaxis2]=1 or [jc_vteprophylaxis3]=1 or [jc_vteprophylaxis5]=1 or [jc_vteprophylaxis6]=1 or [jc_vteprophylaxis8]=1) or ([jc_vteprophylaxis9]=1 and [jc_reasonoralxa]=1)) and datediff([jc_admitdate],[jc_vtedate], 'd', 'mdy', true)< 2) or [jc_novteadmis]=1, 1, 0), '') |
| 1415 | calc_gwtg_antithrombotics | Section Header: *Antithrombotics:Percent of patients with an ischemic stroke or TIA prescribed antithrombotic therapy at discharge.*<br>Antithrombotics<br>*1 = Meets Measure0 = Does Not Meet MeasureBlank = Not in measure* | calc<br>Calculation: if(([gs_stroketype]=2 or [gs_stroketype]=3) and [gs_age]>=18 and [gs_symptomlocation]<>4 and [gs_comfortonly]=4 and [jc_disposition]<>2 and [jc_disposition]<>3 and [jc_disposition]<>4 and [jc_disposition]<>6 and [jc_disposition]<>7 and [gs_antithromboticyesno]<>3 and ([gs_antithromboticyesno]<>2 or ([gs_otheranti]<>1 or ([gs_otheranti_med1]<>1 and [gs_otheranti_med1]<>2 and [gs_otheranti_med2]<>1 and [gs_otheranti_med2]<>2))) and [gs_notadmit]<>1 and [jc_clinical]<>1 and [jc_carotid]<>1, if([gs_antithromboticyesno]=1, 1, 0), '') |
| 1416 | calc_gwtg_anticoag_afib | Section Header: *Anticoag for AFib/AFlutter:Percent of patients with an ischemic stroke or TIA with atrial fibrillation/flutter discharged on anticoagulation therapy.*<br>Anticoag for AFib/AFlutter<br>*1 = Meets Measure0 = Does Not Meet MeasureBlank = Not in measure* | calc<br>Calculation: if(([gs_stroketype]=2 or [gs_stroketype]=3) and ([gs_med_hist(1)]=1 or [gs_persistentafib]=1) and [gs_age]>=18 and [gs_symptomlocation]<>4 and [gs_comfortonly]=4 and [jc_disposition]<>2 and [jc_disposition]<>3 and [jc_disposition]<>4 and [jc_disposition]<>6 and [jc_disposition]<>7 and [gs_afibdischarge]<>3 and [gs_notadmit]<>1 and [jc_clinical]<>1 and [jc_carotid]<>1, if([gs_afibdischarge]=1, 1, 0), '') |
| 1417 | calc_gwtg_smoking_cess | Section Header: *Smoking Cessation:Percent of patients with ischemic or hemorrhagic stroke, or TIA with a history of smoking cigarettes, who are, or whose caregivers are, given smoking cessation advice or counseling during hospital stay.*<br>Smoking Cessation<br>*1 = Meets Measure0 = Does Not Meet MeasureBlank = Not in measure* | calc<br>Calculation: if(([gs_stroketype]=2 or [gs_stroketype]=3 or [gs_stroketype]=4 or [gs_stroketype]=5 or [gs_stroketype]=6) and [gs_age]>=18 and [gs_symptomlocation]<>4 and [gs_comfortonly]=4 and [jc_disposition]<>2 and [jc_disposition]<>3 and [jc_disposition]<>4 and [jc_disposition]<>6 and [jc_disposition]<>7 and [gs_med_hist(9)]=1 and [gs_antismokingtx]<>3 and [gs_notadmit]<>1 and [jc_clinical]<>1 and [jc_carotid]<>1, if([gs_antismokingtx]=1, 1, 0), '') |

| | | | |
|---|---|---|---|
| 1418 | calc_gwtg_statin_at_disch | Section Header: *Statin Prescribed at Discharge:Percent of ischemic stroke or TIA patients who are discharged on Statin Medication.*<br>Statin Prescribed at Discharge<br>*1 = Meets Measure0 = Does Not Meet MeasureBlank = Not in measure* | calc<br>Calculation: if(([gs_stroketype]=2 or [gs_stroketype]=3) and [gs_age]>=18 and [gs_symptomlocation]<>4 and [gs_comfortonly]=4 and [jc_disposition]<>2 and [jc_disposition]<>3 and [jc_disposition]<>4 and [jc_disposition]<>6 and [jc_disposition]<>7 and [jc_nostatindisc]<>1 and [gs_notadmit]<>1 and [jc_clinical]<>1 and [jc_carotid]<>1, if([gs_cholredtx(3)]=1, 1, 0), '') |
| 1419 | calc_gwtg_dysphagia_screen | Section Header: *GWTG-Stroke Quality MeasuresTarget: ≥ 75% on 5 of 8 GWTG - Stroke Quality Measures (Silver Plus or Gold Plus)Dysphagia Screen:Percent of Stroke patients who undergo screening for dysphagia with an evidence-based bedside testing protocol approved by the hospital before being given any food, fluids, or medication by mouth.*<br>Dysphagia Screen<br>*1 = Meets Measure0 = Does Not Meet MeasureBlank = Not in measure* | calc<br>Calculation: if(([gs_stroketype]=2 or [gs_stroketype]=4 or [gs_stroketype]=5 or [gs_stroketype]=6) and [gs_age]>=18 and [gs_symptomlocation]<>4 and [gs_npo]<>1 and [gs_dysphagia]<>3 and [jc_clinical]<>1 and [jc_carotid]<>1 and [gs_notadmit]=2 and ([gs_dysphagia]=1 or [gs_dysphagia]=2 or [gs_dysphagia]=3 or [hospitalization_complete]=2), if([gs_dysphagia]=1, 1, 0), '') |
| 1420 | calc_gwtg_stroke_education | Section Header: *Stroke Education:Percent of patients with Stroke or TIA or their caregivers who were given education and/or educational materials during the hospital stay addressing ALL of the following: personal risk factors for stroke, warning signs for stroke, activation of emergency medical system, need for follow-up after discharge, and medications prescribed.*<br>Stroke Education<br>*1 = Meets Measure0 = Does Not Meet MeasureBlank = Not in measure* | calc<br>Calculation: if(([gs_stroketype]=2 or [gs_stroketype]=3 or [gs_stroketype]=4 or [gs_stroketype]=5 or [gs_stroketype]=6) and [gs_age]>=18 and [gs_symptomlocation]<>4 and [gs_comfortonly]=4 and [jc_disposition]<>2 and [jc_disposition]<>3 and [jc_disposition]<>4 and [jc_disposition]<>6 and [jc_disposition]<>7 and [gs_notadmit]<>1 and [jc_clinical]<>1 and [jc_carotid]<>1, if([jc_eduems]=1 and [jc_edufollowup]=1 and [jc_edumeds]=1 and [jc_edurisk]=1 and [jc_eduwarning]=1, 1, 0), '') |
| 1421 | calc_gwtg_rehab_considered | Section Header: *Rehabilitation Considered:Percent of patients with Stroke who were assessed for rehabilitation services.*<br>Rehabilitation Considered<br>*1 = Meets Measure0 = Does Not Meet MeasureBlank = Not in measure* | calc<br>Calculation: if(([gs_stroketype]=2 or [gs_stroketype]=4 or [gs_stroketype]=5 or [gs_stroketype]=6) and [gs_age]>=18 and [gs_symptomlocation]<>4 and [gs_comfortonly]=4 and [jc_disposition]<>2 and [jc_disposition]<>3 and [jc_disposition]<>4 and [jc_disposition]<>6 and [jc_disposition]<>7 and [gs_notadmit]<>1 and [jc_clinical]<>1 and [jc_carotid]<>1, if([jc_assessrehab]=1, 1, 0), '') |

| | | | |
|---|---|---|---|
| 1422 | calc_gwtg_time_to_ivtpa_60 | Section Header: *Target: Stroke Honor Roll: Door-to-needle times within 60 minutes for at least 75 percent of applicable patients.Target: Stroke Honor Roll-Elite: Door-to-needle times within 60 minutes for at least 85 percent of applicable patients.Time to IV Thrombolytic Therapy - 60 min:Percent of acute ischemic stroke patients receiving intravenous tissue plasminogen activator (alteplase) therapy during the hospital stay who have a time from hospital arrival to initiation of thrombolytic therapy administration (door-to-needle time) of 60 minutes or less.*<br>Time to Intravenous Thrombolytic Therapy - 60 min<br>*1 = Meets Measure0 = Does Not Meet MeasureBlank = Not in measure* | calc<br>Calculation: if([gs_stroketype]=2 and [gs_ivthroinit]=1 and [gs_age]>=18 and [gs_symptomlocation]<>4 and [jc_arrdatetime_precision]=5 and [jc_ivthrodttm_precision]=5 and [jc_ivthrodttm]>=[jc_arrdatetime] and [gs_lastknownwell_precision]=5 and datediff([gs_lastknownwell], [jc_ivthrodttm], "m", "mdy", true) <=270 and [gs_patientarrival]<>3 and ([ivtpadelay]<>1 or ([ivtpadelay_er(1)]<>1 and [ivtpadelay_er(2)]<>1 and [ivtpadelay_er(3)]<>1 and [ivtpadelay_mr(1)]<>1 and [ivtpadelay_mr(2)]<>1 and [ivtpadelay_mr(3)]<>1 and [ivtpadelay_mr(4)]<>1)) and [jc_clinical]<>1, if(datediff([jc_arrdatetime], [jc_ivthrodttm], "m", "mdy", true) <=60, 1, 0), '') |
| 1423 | calc_gwtg_ldl_documented | Section Header: *LDL Documented:Percent of ischemic stroke or TIA patients with a documented Lipid profile.*<br>LDL Documented<br>*1 = Meets Measure0 = Does Not Meet MeasureBlank = Not in measure* | calc<br>Calculation: if(([gs_stroketype]=2 or [gs_stroketype]=3) and [gs_age]>=18 and [gs_symptomlocation]<>4 and [gs_comfortonly]=4 and [jc_disposition]<>2 and [jc_disposition]<>3 and [jc_disposition]<>4 and [jc_disposition]<>6 and [jc_disposition]<>7 and [gs_lipidsnc] <>1 and [gs_notadmit]<>1 and [jc_clinical]<>1 and [jc_carotid] <>1, if([gs_lipidsnd]<>1 and ([gs_ldl]>=0 or ([gs_cholesterol]>=0 and [gs_triglycerides]>=0 and [gs_hdl]>=0)), 1, 0), '') |
| 1424 | calc_gwtg_intensive_statin | Section Header: *Intensive Statin Therapy:Percentage of Ischemic Stroke and TIA patients who are prescribed high-intensity statin therapy at discharge OR, if > 75 years of age, are prescribed at least moderate- intensity statin therapy at discharge.*<br>Intensive Statin Therapy<br>*1 = Meets Measure0 = Does Not Meet MeasureBlank = Not in measure* | calc<br>Calculation: if(([gs_stroketype]=2 or [gs_stroketype]=3) and [gs_age]>=18 and [gs_symptomlocation]<>4 and [jc_disposition]<>2 and [jc_disposition]<>3 and [jc_disposition]<>4 and [jc_disposition]<>6 and [jc_disposition]<>7 and [gs_notadmit]<>1 and [jc_carotid]<>1 and [jc_clinical]<>1 and [gs_comfortonly]<>1 and [gs_comfortonly]<>2 and [gs_comfortonly]<>3 and ([gs_cholredtx(2)]<>1 or [jc_nostatindisc]<>1) and (([gs_cholredtx(4)]<>1 and [gs_cholredtx(6)]<>1 and [gs_cholredtx(7)]<>1 and [gs_cholredtx(8)]<>1 and [gs_cholredtx(9)]<>1) or [jc_nostatindisc]<>1 or [gs_cholredtx(3)]=1) and ([gs_age]>75 or (([gs_statinmed]<>2 or [gs_statindose2]<>151) and ([gs_statinmed]<>4 or ([gs_statindose4]<>300 and [gs_statindose4]<>301)) and ([gs_statinmed]<>5 or [gs_statindose5]<>402) and ([gs_statinmed]<>6 or ([gs_statindose6]<>500 and [gs_statindose6]<>501)) and ([gs_statinmed]<>7 or ([gs_statindose7]<>651 and [gs_statindose7]<>652)) and ([gs_statinmed]<>8 or [gs_statindose8]<>701) and ([gs_statinmed]<>9 or ([gs_statindose9]<>802 and [gs_statindose9]<>803)) and ([gs_statinmed]<>12 or ([gs_statindose12]<>1102 and [gs_statindose12]<>1103)) and |

| | | | |
|---|---|---|---|
| | | | ([gs_statinmed]<>14 or [gs_statindose14]<>1300)) or ([gs_noguidedose(1)]<>1 and [gs_noguidedose(2)]<>1 and [gs_noguidedose(3)]<>1)) and ((([gs_statinmed]<>5 or ([gs_statindose5]<>400 and [gs_statindose5]<>401)) and ([gs_statinmed]<>7 or [gs_statindose7]<>650) and ([gs_statinmed]<>8 or ([gs_statindose8]<>704 and [gs_statindose8]<>700)) and ([gs_statinmed]<>9 or [gs_statindose9]<>800 or [gs_statindose9]<>801) and ([gs_statinmed]<>12 or [gs_statindose12]<>1101)) or ([gs_noguidedose(1)]<>1 and [gs_noguidedose(2)]<>1 and [gs_noguidedose(3)]<>1)), if((([gs_statinmed]=3 and ([gs_statindose3]=202 or [gs_statindose3]=205 or [gs_statindose3]=209)) or ([gs_statinmed]=4 and [gs_statindose4]=302) or ([gs_statinmed]=6 and [gs_statindose6]=502) or ([gs_statinmed]=11 and [gs_statindose11]=1003) or ([gs_statinmed]=12 and [gs_statindose12]=1104)) or ([gs_age]>75 and (([gs_statinmed]=2 and [gs_statindose2]=151) or ([gs_statinmed]=4 and ([gs_statindose4]=300 or [gs_statindose4]=301)) or ([gs_statinmed]=5 and [gs_statindose5]=402) or ([gs_statinmed]=6 and ([gs_statindose6]=500 or [gs_statindose6]=501)) or ([gs_statinmed]=7 and ([gs_statindose7]=651 or [gs_statindose7]=652)) or ([gs_statinmed]=8 and [gs_statindose8]=701) or ([gs_statinmed]=9 and ([gs_statindose9]=802 or [gs_statindose9]=803)) or ([gs_statinmed]=12 and ([gs_statindose12]=1102 or [gs_statindose12]=1103)) or ([gs_statinmed]=14 and [gs_statindose14]=1300))), 1, 0), '') |
| 1425 | calc_gwtg_arrive3_treat4 | Section Header: *IV rt-PA Arrive by 3.5 Hours, Treat by 4.5 Hours:Percent of acute ischemic stroke patients who arrive at the hospital within 210 minutes (3.5 hours) of time last known well and for whom IV t-PA was initiated at this hospital within 270 minutes (4.5 hours) of time last known well.*<br>IV rt-PA Arrive by 3.5 Hours, Treat by 4.5 Hours<br>*1 = Meets Measure0 = Does Not Meet MeasureBlank = Not in measure* | calc<br>Calculation: if([gs_stroketype]=2 and [gs_age]>=18 and [gs_symptomlocation]<>4 and [gs_ivtpaoutside]<>1 and [calc_lkw_to_arrival]>=0 and [calc_lkw_to_arrival]<=210 and ([calc_lkw_to_arrival]>120 or [gs_ivtpanc]<>1 or ([gs_exclusion(1)]<>1 and [gs_exclusion(2)]<>1 and [gs_exclusion(3)]<>1 and [gs_exclusion(4)]<>1 and [gs_exclusion(5)]<>1 and [gs_exclusion(6)]<>1 and [gs_exclusion(7)]<>1 and [gs_exclusion(8)]<>1 and [gs_exclusion(9)]<>1 and [gs_relexclusion(1)]<>1 and [gs_relexclusion(2)]<>1 and [gs_relexclusion(3)]<>1 and [gs_relexclusion(4)]<>1 and [gs_relexclusion(5)]<>1 and [gs_relexclusion(6)]<>1 and [gs_relexclusion(7)]<>1 and [gs_relexclusion(8)]<>1 and [gs_relexclusion(9)]<>1 and [gs_relexclusion(10)]<>1 and [gs_relexclusion(11)]<>1) or [gs_ivtpalatenc]<>1 or ([gs_exclusion2(1)]<>1 and [gs_exclusion2(2)]<>1 and [gs_exclusion2(3)]<>1 and [gs_exclusion2(4)]<>1 and [gs_exclusion2(5)]<>1 and [gs_exclusion2(6)]<>1 and [gs_exclusion2(7)]<>1 and |

| | | | |
|---|---|---|---|
| | | | [gs_exclusion2(8)]<>1 and [gs_exclusion2(9)]<>1 and [gs_relexclusion2(1)]<>1 and [gs_relexclusion2(2)]<>1 and [gs_relexclusion2(3)]<>1 and [gs_relexclusion2(4)]<>1 and [gs_relexclusion2(5)]<>1 and [gs_relexclusion2(6)]<>1 and [gs_relexclusion2(7)]<>1 and [gs_relexclusion2(8)]<>1 and [gs_relexclusion2(9)]<>1 and [gs_relexclusion2(10)]<>1 and [gs_relexclusion2(11)]<>1 and [gs_addrelexcl(1)]<>1 and [gs_addrelexcl(2)]<>1 and [gs_addrelexcl(3)]<>1 and [gs_addrelexcl(4)]<>1)) and ([calc_lkw_to_arrival]<=120 or [gs_ivtpalatenc]<>1 or ([gs_exclusion2(1)]<>1 and [gs_exclusion2(2)]<>1 and [gs_exclusion2(3)]<>1 and [gs_exclusion2(4)]<>1 and [gs_exclusion2(5)]<>1 and [gs_exclusion2(6)]<>1 and [gs_exclusion2(7)]<>1 and [gs_exclusion2(8)]<>1 and [gs_exclusion2(9)]<>1 and [gs_relexclusion2(1)]<>1 and [gs_relexclusion2(2)]<>1 and [gs_relexclusion2(3)]<>1 and [gs_relexclusion2(4)]<>1 and [gs_relexclusion2(5)]<>1 and [gs_relexclusion2(6)]<>1 and [gs_relexclusion2(7)]<>1 and [gs_relexclusion2(8)]<>1 and [gs_relexclusion2(9)]<>1 and [gs_relexclusion2(10)]<>1 and [gs_relexclusion2(11)]<>1 and [gs_addrelexcl(1)]<>1 and [gs_addrelexcl(2)]<>1 and [gs_addrelexcl(3)]<>1 and [gs_addrelexcl(4)]<>1)) and ([calc_lkw_to_arrival]>180 or [gs_ivtpanc]<>1 or ([gs_exclusion(2)]<>1 and [gs_exclusion(3)]<>1 and [gs_exclusion(4)]<>1 and [gs_exclusion(5)]<>1 and [gs_exclusion(6)]<>1 and [gs_exclusion(7)]<>1 and [gs_relexclusion(2)]<>1 and [gs_relexclusion(3)]<>1 and [gs_relexclusion(4)]<>1 and [gs_relexclusion(10)]<>1)) and [gs_lastknownwell_precision]<>3 and [jc_arrdatetime_precision]<>3 and ([gs_ivthroinit]<>1 or [jc_ivthrodttm_precision]=5) and [jc_clinical]<>1 and [jc_carotid]<>1, if([gs_ivthroinit]=1 and datediff([gs_lastknownwell], [jc_ivthrodttm], "m", "mdy", true) <=270, 1, 0), '') |
| 1426 | calc_gwtg_nihss_reported | Section Header: *NIHSS Reported:Percent of ischemic stroke and stroke not otherwise specified patients with a score reported for NIH Stroke Scale (Initial).*<br>NIHSS Reported<br>*1 = Meets Measure0 = Does Not Meet MeasureBlank = Not in measure* | calc<br>Calculation: if(([gs_stroketype]=2 or [gs_stroketype]=6) and [gs_age]>=18 and [gs_symptomlocation]<>4 and [gs_symptresolve]<>1 and [jc_clinical]<>1 and [jc_carotid]<>1 and [gs_notadmit]=2, if([gs_nihssperformed]=1 and [gs_nihssadmit]>=0, 1, 0), '') |

| | | | |
|---|---|---|---|
| 1427 | calc_gwtg_time_to_ivtpa_45 | Section Header: *GWTG-Stroke Reporting MeasuresTarget: Stroke Honor Roll-Elite Plus: Door-to-needle times within 45 minutes for at least 75 percent of applicable patients AND Door-to-needle times within 30 minutes for at least 50 percent of applicable patients.Time to IV Thrombolytic Therapy - 45 min:Percent of acute ischemic stroke patients receiving intravenous tissue plasminogen activator (alteplase) therapy during the hospital stay who have a time from hospital arrival to initiation of thrombolytic therapy administration (door-to-needle time) of 45 minutes or less.*<br>Time to Intravenous Thrombolytic Therapy - 45 min<br>*1 = Meets Measure0 = Does Not Meet MeasureBlank = Not in measure* | calc<br>Calculation: if([gs_stroketype]=2 and [gs_ivthroinit]=1 and [gs_age]>=18 and [gs_symptomlocation]<>4 and [jc_arrdatetime_precision]=5 and [jc_ivthrodttm_precision]=5 and [jc_ivthrodttm]>=[jc_arrdatetime] and [gs_lastknownwell_precision]=5 and datediff([gs_lastknownwell], [jc_ivthrodttm], "m", "mdy", true) <=270 and [gs_patientarrival]<>3 and ([ivtpadelay]<>1 or ([ivtpadelay_er(1)]<>1 and [ivtpadelay_er(2)]<>1 and [ivtpadelay_er(3)]<>1 and [ivtpadelay_mr(1)]<>1 and [ivtpadelay_mr(2)]<>1 and [ivtpadelay_mr(3)]<>1 and [ivtpadelay_mr(4)]<>1)) and [jc_clinical]<>1, if(datediff([jc_arrdatetime], [jc_ivthrodttm], "m", "mdy", true) <=45, 1, 0), '') |
| 1428 | calc_gwtg_time_to_ivtpa_30 | Section Header: *Time to IV Thrombolytic Therapy - 30 min:Percent of acute ischemic stroke patients receiving intravenous tissue plasminogen activator (alteplase) therapy during the hospital stay who have a time from hospital arrival to initiation of thrombolytic therapy administration (door-to-needle time) of 30 minutes or less.*<br>Time to Intravenous Thrombolytic Therapy - 30 min<br>*1 = Meets Measure0 = Does Not Meet MeasureBlank = Not in measure* | calc<br>Calculation: if([gs_stroketype]=2 and [gs_ivthroinit]=1 and [gs_age]>=18 and [gs_symptomlocation]<>4 and [jc_arrdatetime_precision]=5 and [jc_ivthrodttm_precision]=5 and [jc_ivthrodttm]>=[jc_arrdatetime] and [gs_lastknownwell_precision]=5 and datediff([gs_lastknownwell], [jc_ivthrodttm], "m", "mdy", true) <=270 and [gs_patientarrival]<>3 and ([ivtpadelay]<>1 or ([ivtpadelay_er(1)]<>1 and [ivtpadelay_er(2)]<>1 and [ivtpadelay_er(3)]<>1 and [ivtpadelay_mr(1)]<>1 and [ivtpadelay_mr(2)]<>1 and [ivtpadelay_mr(3)]<>1 and [ivtpadelay_mr(4)]<>1)) and [jc_clinical]<>1, if(datediff([jc_arrdatetime], [jc_ivthrodttm], "m", "mdy", true) <=30, 1, 0), '') |
| 1429 | calc_gwtg_time_to_ivtpa | Section Header: *Time to Intravenous Thrombolytic Therapy:Time from hospital arrival to initiation of thrombolytic therapy administration for ischemic stroke patients treated at this hospital.*<br>Time to Intravenous Thrombolytic Therapy<br>*Time in Minutes* | calc<br>Calculation: if([gs_stroketype]=2 and [gs_ivthroinit]=1 and [gs_age]>=18 and [gs_symptomlocation]<>4 and [jc_arrdatetime_precision]=5 and [jc_ivthrodttm_precision]=5 and [jc_ivthrodttm]>=[jc_arrdatetime] and [gs_lastknownwell_precision]=5 and datediff([gs_lastknownwell], [jc_ivthrodttm], 'm', 'mdy', true) <=270 and [gs_patientarrival]<>3 and ([ivtpadelay]<>1 or ([ivtpadelay_er(1)]=0 and [ivtpadelay_er(2)]=0 and [ivtpadelay_er(3)]=0 and [ivtpadelay_mr(1)]=0 and [ivtpadelay_mr(2)]=0 and [ivtpadelay_mr(3)]=0 and [ivtpadelay_mr(4)]=0)) and [jc_clinical]<>1, datediff([jc_arrdatetime], [ivthrodttm], 'm', 'mdy', true), '') |

| | | | |
|---|---|---|---|
| 1430 | calc_cva_poc_fallout | Section Header: *Special InitiativesCVA POC:Description*<br>CVA POC<br>*1 = Meets Measure0 = Does Not Meet MeasureBlank = Not in measure* | calc<br>Calculation: if([gs_notadmit]='2' and [cva_poc_na(1)]=0 and [additional_stroke_information_complete]=2 and ([cva_poc_init]=0 or [cva_poc_init]=1), if([cva_poc_init]='1' and [cva_poc_signed]='1' and [calc_cva_poc]='100' and [cva_poc_risk_fact_init]='1' and [cva_poc_risk_fact_ident]='1' and [cva_poc_indiv_needs_ident]<>'0', 1, 0), '') |
| 1431 | calc_swallow_screen | Section Header: *Swallow Screen:Percent of Stroke patients who pass a screening for dysphagia with an evidence-based bedside testing protocol approved by the hospital before being given, or having documented as given, any food, fluids, or medication by mouth.*<br>Swallow Screen<br>*1 = Meets Measure0 = Does Not Meet MeasureBlank = Not in measure* | calc<br>Calculation: if(([gs_stroketype]=2 or [gs_stroketype]=3 or [gs_stroketype]=4 or [gs_stroketype]=5 or [gs_stroketype]=6) and [gs_age]>=18 and [gs_dysphagia]<>3 and [jc_clinical]<>1 and [jc_carotid]<>1 and [gs_notadmit]=2 and ([gs_dysphagia]=1 or [gs_dysphagia]=2 or [gs_dysphagia]=3 or [hospitalization_complete]=2), if(([gs_dysphagia]=1 and [gs_dysphagiaresult]=1 and [swallowscreen_subsequent]=0) or ([gs_dysphagia]=1 and [gs_dysphagiaresult]=2 and [swallowscreen_failed]=0 and [swallowscreen_subsequent]=0), 1, 0), '') |

| | | | |
|---|---|---|---|
| 1432 | calc_post_ir_assessment | Section Header: *Post IR Assessments:Stroke patients undergoing any endovascular procedure for whom post IR assessments have been completed per protocol and/or hospital policy.*<br><br>Post IR Assessments<br>*1 = Meets Measure0 = Does Not Meet MeasureBlank = Not in measure* | calc<br>Calculation: if ( ( [treatment_type(IAtPA)]=1 or [treatment_type(MER)]=1 or [gs_iatpa]=1 or [procedures_interventions(2)]=1 or [procedures_interventions(4)]=1 or [procedures_interventions(6)]=1 ), if ( if ( [ev_proc_type_1]<>'', if ( [ev_proc_protocol_1]<>0 and [ev_proc_ir_1]<>0 and [ev_proc_transit_1]<>0 and [calc_ev_proc_assessment_1]=100, if ( [ev_proc_type_1]=1, if ( [calc_post_tpa_compliance]=100, 1, 0 ), 1 ), 0 ), '' )=0 or if ( [ev_proc_type_2]<>'', if ( [ev_proc_protocol_2]<>0 and [ev_proc_ir_2]<>0 and [ev_proc_transit_2]<>0 and [calc_ev_proc_assessment_2]=100, if ( [ev_proc_type_2]=1, if ( [calc_post_tpa_compliance]=100, 1, 0 ), 1 ), 0 ), '' )=0 or if ( [ev_proc_type_3]<>'', if ( [ev_proc_protocol_3]<>0 and [ev_proc_ir_3]<>0 and [ev_proc_transit_3]<>0 and [calc_ev_proc_assessment_3]=100, if ( [ev_proc_type_3]=1, if ( [calc_post_tpa_compliance]=100, 1, 0 ), 1 ), 0 ), '' )=0 or if ( [ev_proc_type_4]<>'', if ( [ev_proc_protocol_4]<>0 and [ev_proc_ir_4]<>0 and [ev_proc_transit_4]<>0 and [calc_ev_proc_assessment_4]=100, if ( [ev_proc_type_4]=1, if ( [calc_post_tpa_compliance]=100, 1, 0 ), 1 ), 0 ), '' )=0, 0, if ( if ( [ev_proc_type_1]<>'', if ( [ev_proc_protocol_1]<>0 and [ev_proc_ir_1]<>0 and [ev_proc_transit_1]<>0 and [calc_ev_proc_assessment_1]=100, if ( [ev_proc_type_1]=1, if ( [calc_post_tpa_compliance]=100, 1, 0 ), 1 ), 0 ), '' )='' and if ( [ev_proc_type_2]<>'', if ( [ev_proc_protocol_2]<>0 and [ev_proc_ir_2]<>0 and [ev_proc_transit_2]<>0 and [calc_ev_proc_assessment_2]=100, if ( [ev_proc_type_2]=1, if ( [calc_post_tpa_compliance]=100, 1, 0 ), 1 ), 0 ), '' )='' and if ( [ev_proc_type_3]<>'', if ( [ev_proc_protocol_3]<>0 and [ev_proc_ir_3]<>0 and [ev_proc_transit_3]<>0 and [calc_ev_proc_assessment_3]=100, if ( [ev_proc_type_3]=1, if ( [calc_post_tpa_compliance]=100, 1, 0 ), 1 ), 0 ), '' )='' and if ( [ev_proc_type_4]<>'', if ( [ev_proc_protocol_4]<>0 and [ev_proc_ir_4]<>0 and [ev_proc_transit_4]<>0 and [calc_ev_proc_assessment_4]=100, if ( [ev_proc_type_4]=1, if ( [calc_post_tpa_compliance]=100, 1, 0 ), 1 ), 0 ), '' )='', '', 1 ) ), '' ) |
| 1433 | calc_post_tpa_monitoring | Section Header: *Post IV Thrombolytic Monitoring:Stroke patients receiving IV Thrombolytic, Thrombectomy, and/or Endovascular Therapy for whom Post IV Thrombolytic Monitoring has been completed per protocol and/or hospital policy.*<br><br>Post IV Thrombolytic Monitoring<br>*1 = Meets Measure0 = Does Not Meet MeasureBlank = Not in measure* | calc<br>Calculation: if(([treatment_type(IAtPA)]='1' or [treatment_type(IVtPA)]='1' or [treatment_type(MER)]='1' or [gs_ivthroinit]='1' or [gs_ivtpaoutside]='1'), if([calc_post_tpa_compliance]=100 and [post_tpa_bp_addressed]<>0, 1, 0), '') |

| | | | |
|---|---|---|---|
| 1434 | calc_fib_edu | Section Header: *Fibrinolytic Therapy Education:Stroke patients receiving IV Thrombolytic for whom Fibrinolytic Therapy Education has been completed and documented.*<br>Fibrinolytic Therapy Education<br>*1 = Meets Measure0 = Does Not Meet MeasureBlank = Not in measure* | calc<br>Calculation: if(([treatment_type(IVtPA)]='1' or [gs_ivthroinit]='1' or [gs_ivtpaoutside]='1') and [gs_comfortonly]<>'1' and [gs_comfortonly]<>'2' and [gs_comfortonly]<>'3' and [jc_disposition]<>'2' and [jc_disposition]<>'3' and [jc_disposition]<>'6' and [jc_disposition]<>'7' and [jc_disposition]<>'8', if([edu_thrombolytic]=1, 1, 0), '') |
| 1435 | calc_thro_edu | Section Header: *Arteriogram Education:Stroke patients receiving Thrombectomy, Coil, and/or Angiogram for whom Arteriogram Education has been completed and documented.*<br>Arteriogram Education<br>*1 = Meets Measure0 = Does Not Meet MeasureBlank = Not in measure* | calc<br>Calculation: if(([treatment_type(IAtPA)]='1' or [treatment_type(MER)]='1' or [gs_iatpa]='1' or [procedures_interventions(2)]='1' or [procedures_interventions(6)]='1') and ([gs_stroketype]='2' or [gs_stroketype]='3' or [gs_stroketype]='4' or [gs_stroketype]='5' or [gs_stroketype]='6') and [gs_comfortonly]<>'1' and [gs_comfortonly]<>'2' and [gs_comfortonly]<>'3' and [jc_disposition]<>'2' and [jc_disposition]<>'3' and [jc_disposition]<>'6' and [jc_disposition]<>'7' and [jc_disposition]<>'8', if([edu_arteriogram]=1, 1, 0), '') |
| 1436 | calc_flush_tnk_combined | Section Header: *Normal Saline Flush was documented Before and After Tenecteplase given*<br>Flush Documented Before and After Tenecteplase | calc<br>Calculation: if([iv_thrombolytic_type]='2',([flush_before_tnk] + [flush_after_tnk]) / .02,'') |
| 1437 | measure_calculations_complete | Section Header: *Form Status*<br>Complete? | dropdown<br>0 Incomplete<br>1 Unverified<br>2 Complete |

Instrument: **Report Calculations Only** (report_calculations_only)

Collapse

| | | | |
|---|---|---|---|
| 1438 | calc_days_post_onset | Days Since Stroke Onset<br>*(until 90 day mRS or 365 days)* | calc<br>Calculation: if([initial_stroke_information_complete]=2 and [admin_complete]=2 and [clinical_codes_complete]=2 and [admission_complete]=2 and [hospitalization_complete]=2 and [advanced_stroke_care_complete]=2 and [additional_stroke_information_complete]=2 and [optional_complete]=2 and [discharge_complete]=2 and [gs_notadmit]=2, if([postdis_mrs_date]='', if([gs_symptomdatetime]<>'', if(rounddown(datediff([gs_symptomdatetime], 'today', 'd', 'mdy'), 0)< 365, rounddown(datediff([gs_symptomdatetime], 'today', 'd', 'mdy'), 0), 365), if([gs_lastknownwell]<>'', if(rounddown(datediff([gs_lastknownwell], 'today', 'd', 'mdy'), 0)< 365, rounddown(datediff([gs_lastknownwell], 'today', 'd', 'mdy'), 0), 365), if(rounddown(datediff([jc_arrdatetime], 'today', 'd', 'mdy'), 0)< 365, rounddown(datediff([jc_arrdatetime], 'today', 'd', 'mdy'), 0), 365))), if([gs_symptomdatetime]<>'', rounddown(datediff([gs_symptomdatetime], [postdis_mrs_date], 'd', 'mdy'), 0), if([gs_lastknownwell]<>'', rounddown(datediff([gs_lastknownwell], [postdis_mrs_date], 'd', 'mdy'), 0), rounddown(datediff([jc_arrdatetime], [postdis_mrs_date], 'd', 'mdy'), 0)))), '') |
| 1439 | calc_needle_to_artpunc | Needle to Groin<br>*Time in Minutes* | calc<br>Calculation: if([ivthrodttm]<>'' and [artpunc_dt]<>'', datediff([ivthrodttm], [artpunc_dt], 'm', 'mdy', true), if([jc_ivthrodttm]<>'' and [c_artpuncdt]<>'', datediff([jc_ivthrodttm], [c_artpuncdt], 'm', 'mdy', true), '')) |
| 1440 | calc_needle_to_device | Needle to Device<br>*Time in Minutes* | calc<br>Calculation: if([ivthrodttm]<>'' and [firstpassdt]<>'', datediff([ivthrodttm], [firstpassdt], 'm', 'mdy', true), if([jc_ivthrodttm]<>'' and [c_firstpassdt]<>'', datediff([jc_ivthrodttm], [c_firstpassdt], 'm', 'mdy', true), '')) |
| 1441 | calc_needle_to_reperfusion | Needle to Reperfusion<br>*Time in Minutes* | calc<br>Calculation: if([ivthrodttm]<>'' and [ticidt]<>'', datediff([ivthrodttm], [ticidt], 'm', 'mdy', true), if([jc_ivthrodttm]<>'' and [mer_ticidt]<>'', datediff([jc_ivthrodttm], [mer_ticidt], 'm', 'mdy', true), '')) |
| 1442 | calc_needle_to_transfer | Needle to Transfer<br>*Time in Minutes* | calc<br>Calculation: if([gs_notadmitwhy]=1, if([ivthrodttm]<>'', datediff([ivthrodttm], [gs_discdatetime], 'm', 'mdy', true), if([jc_ivthrodttm]<>'', datediff([jc_ivthrodttm], [gs_discdatetime], 'm', 'mdy', true), '')), '') |
| 1443 | calc_transfer_out | Transfer Out | calc<br>Calculation: if([gs_notadmitwhy]=1 or [jc_disposition]=4,1,'') |

| | | | |
|---|---|---|---|
| 1444 | calc_ischemic_stroke | Ischemic Stroke | calc<br>Calculation: if([gs_stroketype]=2 and [gs_age]>=18 and [jc_clinical]<>1 and [jc_carotid]<>1, 1, '') |
| 1445 | calc_ischemic_tx_tpa | Ischemic Stroke Treated with tPA | calc<br>Calculation: if([gs_stroketype]=2 and [gs_age]>=18 and [jc_clinical]<>1 and [jc_carotid]<>1, if([gs_ivthroinit]=1, 1, 0), '') |
| 1446 | calc_ischemic_tx_mer | Ischemic Stroke Treated with MER | calc<br>Calculation: if([gs_stroketype]=2 and [gs_age]>=18 and [jc_clinical]<>1 and [jc_carotid]<>1, if([gs_iatpa]=1, 1, 0), '') |
| 1447 | desc_c_ipp | CSTK Initial Patient Population<br><br>0 = Not in CSTK Initial Patient Population<br>1 = Ischemic Stroke Without Procedure<br>2 = Ischemic Stroke With IV t-PA, IA t-PA, or MER<br>3 = Hemorrhagic Stroke | descriptive |
| 1448 | report_calculations_only_complete | Section Header: *Form Status*<br>Complete? | dropdown<br>0 Incomplete<br>1 Unverified<br>2 Complete |

Instrument: **Special Initiatives** (special_initiatives) Collapse

| | | | |
|---|---|---|---|
| 1449 | ems_patientarrival_desc<br><br>Show the field ONLY if:<br>[gs_symptomlocation]<>'' or [gs_patientarrival]<>'' and [hide_nonessential_fields(1)]='0' | Section Header: *EMS Data Elements*<br>Patient location when stroke symptoms discovered: [gs_symptomlocation]<br>How patient arrived at your hospital: [gs_patientarrival] | descriptive |
| 1450 | ems_recordlater<br><br>Show the field ONLY if:<br>([site]='SMCA' or [site]='SMCH' or [site]='SMCW' or [site]='UMCB') and ([gs_patientarrival]='1' or [gs_patientarrival]='3' or [gs_patientarrival]='10') and [ems_recordavail]<>'1' and [hide_nonessential_fields(1)]='0' | Patient care record available at a later time during hospitalization? | radio<br>1 Yes<br>2 No/ND<br><br>Custom alignment: RH |

| | | | |
|---|---|---|---|
| 1451 | emsx_recordabstractor<br><br>Show the field ONLY if:<br>[gs_patientarrival]='1' or [gs_patientarrival]='3' or [gs_patientarrival]='10' and [hide_nonessential_fields(1)]='0' | ✪ EMS run sheet obtained by abstractor? | yesno<br>1 Yes<br>0 No<br><br>Custom alignment: RH<br>Field Annotation: @HIDDEN |
| 1452 | ems_agency_unk<br><br>Show the field ONLY if:<br>([site]='SMCA' or [site]='SMCH' or [site]='SMCW' or [site]='UMCB') and ([gs_patientarrival]='1' or [gs_patientarrival]='3' or [gs_patientarrival]='10') and [hide_nonessential_fields(1)]='0' | EMS Agency Unknown | radio<br>1 Unknown |

| | | | |
|---|---|---|---|
| 1453 | ems_agency_smca<br><br>Show the field ONLY if:<br>[site]='SMCA' and ([gs_patientarrival]='1' or [gs_patientarrival]='3' or [gs_patientarrival]='10') and [ems_agency_unk]<>'1' and [hide_nonessential_fields(1)]='0' | EMS agency name or number | dropdown<br>7000 Acadian Ambulance Services<br>7026 Aero Care<br>7027 Air Evac Lifeteam - Brownwood<br>7028 American Medical Response<br>6999 Austin / Travis County EMS<br>7029 B&M Ambulance Service, Inc. (Lee County EMS)<br>7030 Blanco Volunteer Ambulance Corp (Blanco EMS)<br>7031 Buda Fire Dept EMS (Hays County EMS District 8)<br>7032 Burnet Fire & EMS<br>7033 Fayette County EMS<br>7034 Gonzales County EMS<br>7035 Harker Heights Fire Dept EMS<br>7036 Luling EMS (City of Luling)<br>7037 Marble Falls Area EMS<br>7038 Pflugerville Fire Dept EMS (Travis County ESD 2)<br>7039 PHI Air Medical<br>7040 San Marcos-Hays County EMS<br>7475 St Joseph Regional Health Center EMS<br>7041 STAR Flight (Travis County Star Flight)<br>7042 Williamson County EMS (Georgetown, TX)<br>7043 Wimberley EMS |
| 1454 | ems_agency_smch<br><br>Show the field ONLY if:<br>[site]='SMCH' and ([gs_patientarrival]='1' or [gs_patientarrival]='3' or [gs_patientarrival]='10') and [ems_agency_unk]<>'1' and [hide_nonessential_fields(1)]='0' | EMS agency name or number | dropdown<br>7044 Acadian Ambulance Services<br>7045 Austin / Travis County EMS<br>7046 B&M Ambulance Service, Inc. (Lee County EMS)<br>7047 Buda Fire Dept EMS (Hays County EMS District 8)<br>7048 Lockhart EMS (City of Lockhart)<br>7049 Luling EMS (City of Luling)<br>7050 San Marcos-Hays County EMS<br>7051 SouthernCross Ambulance Inc (New Braunfels)<br>7052 Wimberley EMS |

<table>
<tr>
<td>1455</td>
<td>ems_agency_smcw<br><br>Show the field ONLY if:<br>[site]='SMCW' and ([gs_patientarrival]='1' or [gs_patientarrival]='3' or [gs_patientarrival]='10') and [ems_agency_unk]<>'1' and [hide_nonessential_fields(1)]='0'</td>
<td>EMS agency name or number</td>
<td>dropdown
<table>
<tr><td>7053</td><td>Acadian Ambulance Services</td></tr>
<tr><td>7054</td><td>Air Evac Lifeteam - Brownwood</td></tr>
<tr><td>7055</td><td>American Medical Response</td></tr>
<tr><td>7056</td><td>Austin / Travis County EMS</td></tr>
<tr><td>7057</td><td>Burnet Fire & EMS</td></tr>
<tr><td>7058</td><td>Georgetown Fire Department EMS (City of Georgetown)</td></tr>
<tr><td>7059</td><td>Harker Heights Fire Dept EMS</td></tr>
<tr><td>7061</td><td>Llano County EMS (Scott & White)</td></tr>
<tr><td>7063</td><td>Marble Falls Area EMS</td></tr>
<tr><td>7064</td><td>Pflugerville Fire Dept EMS (Travis County ESD 2)</td></tr>
<tr><td>7060</td><td>PHI Air Medical</td></tr>
<tr><td>7062</td><td>Scott & White EMS</td></tr>
<tr><td>7065</td><td>STAR Flight (Travis County Star Flight)</td></tr>
<tr><td>7066</td><td>Williamson County EMS (Georgetown, TX)</td></tr>
</table>
</td>
</tr>
</table>

| | | | |
|---|---|---|---|
| 1456 | ems_agency_umcb<br><br>Show the field ONLY if:<br>[site]='UMCB' and ([gs_patientarrival]='1' or [gs_patientarrival]='3' or [gs_patientarrival]='10') and [ems_agency_unk]<>'1' and [hide_nonessential_fields(1)]='0' | EMS agency name or number | dropdown (autocomplete)<br>6997 Acadian Ambulance Services<br>7004 Air Evac Lifeteam - Brownwood<br>7005 American Medical Response<br>6998 Austin / Travis County EMS<br>7006 B&M Ambulance Service, Inc. (Lee County EMS)<br>7007 Blanco Volunteer Ambulance Corp (Blanco EMS)<br>7008 Buda Fire Dept EMS (Hays County EMS District 8)<br>7010 Burnet Fire & EMS<br>7009 Colorado County EMS<br>7011 Fayette County EMS<br>7012 Harker Heights Fire Dept EMS<br>7013 Lavaca County Rescue Service<br>7014 Llano County EMS (Scott & White)<br>7015 Lockhart EMS (City of Lockhart)<br>7016 Luling EMS (City of Luling)<br>7017 Pflugerville Fire Dept EMS (Travis County ESD 2)<br>7018 PHI Air Medical<br>7019 San Marcos-Hays County EMS<br>7020 Seguin Fire/EMS (City of Seguin)<br>7021 SouthernCross Ambulance Inc (New Braunfels)<br>7022 St. Joseph's Regional EMS (Bryan, TX)<br>7023 STAR Flight (Travis County Star Flight)<br>7024 Williamson County EMS (Georgetown, TX)<br>7025 Wimberley EMS |
| 1457 | ems_runseq_unk<br><br>Show the field ONLY if:<br>([site]='SMCA' or [site]='SMCH' or [site]='SMCW' or [site]='UMCB') and ([gs_patientarrival]='1' or [gs_patientarrival]='3' or [gs_patientarrival]='10') and [hide_nonessential_fields(1)]='0' | Run/Sequence Number Unknown | radio<br>1 Unknown |

| | | | |
|---|---|---|---|
| 1458 | ems_runseq<br><br>Show the field ONLY if:<br>([site]='SMCA' or [site]='SMCH' or [site]='SMCW' or [site]='UMCB') and ([gs_patientarrival]='1' or [gs_patientarrival]='3' or [gs_patientarrival]='10') and [ems_runseq_unk]<>'1' and [hide_nonessential_fields(1)]='0' | Run/Sequence number | text<br>Field Annotation: @CHARLIMIT=25 |
| 1459 | desc_ems_times<br><br>Show the field ONLY if:<br>[hide_nonessential_fields(1)]='0' | | descriptive |
| 1460 | ems_msuct_precision<br><br>Show the field ONLY if:<br>([site]='SMCA' or [site]='SMCH' or [site]='SMCW' or [site]='UMCB') and [gs_patientarrival]='10' and [hide_nonessential_fields(1)]='0' | Date/Time Brain Imaging initiated by MSU Precision | dropdown<br>5 \| Date & Time (MM/DD/YYYY HH24:MM)<br>3 \| Date Only (MM/DD/YYYY)<br>0 \| Unknown<br><br>Field Annotation: @HIDDEN |
| 1461 | ems_msuct<br><br>Show the field ONLY if:<br>([site]='SMCA' or [site]='SMCH' or [site]='SMCW' or [site]='UMCB') and [gs_patientarrival]='10' and [ems_msuct_precision]<>'0' and [hide_nonessential_fields(1)]='0' | Date/Time Brain Imaging initiated by MSU<br>*If Date Only, enter 00:00 for time.* | text (datetime_mdy, Min: 2010-01-01 00:00, Max: 2099-12-31 23:59), Identifier<br>Field Annotation: @HIDDEN |
| 1462 | ems_msutpa_precision<br><br>Show the field ONLY if:<br>([site]='SMCA' or [site]='SMCH' or [site]='SMCW' or [site]='UMCB') and [gs_patientarrival]='10' and [hide_nonessential_fields(1)]='0' | Date/Time IV tPA administered by MSU Precision | dropdown<br>5 \| Date & Time (MM/DD/YYYY HH24:MM)<br>3 \| Date Only (MM/DD/YYYY)<br>0 \| Unknown<br><br>Field Annotation: @HIDDEN |
| 1463 | ems_msutpa<br><br>Show the field ONLY if:<br>([site]='SMCA' or [site]='SMCH' or [site]='SMCW' or [site]='UMCB') and [gs_patientarrival]='10' and [ems_msutpa_precision]<>'0' and [hide_nonessential_fields(1)]='0' | Date/Time IV tPA administered by MSU<br>*If Date Only, enter 00:00 for time.* | text (datetime_mdy, Min: 2010-01-01 00:00, Max: 2099-12-31 23:59), Identifier<br>Field Annotation: @HIDDEN |

| | | | |
|---|---|---|---|
| 1464 | ems_callrecdt_precision<br><br>Show the field ONLY if:<br>([site]='SMCA' or [site]='SMCH' or [site]='SMCW' or [site]='UMCB') and ([gs_patientarrival]='1' or [gs_patientarrival]='3' or [gs_patientarrival]='10') and [hide_nonessential_fields(1)]='0' | EMS Unit Notified by Dispatch Precision | dropdown<br>5 \| Date & Time (MM/DD/YYYY HH24:MM)<br>3 \| Date Only (MM/DD/YYYY)<br>0 \| Unknown |
| 1465 | ems_callrecdt<br><br>Show the field ONLY if:<br>([site]='SMCA' or [site]='SMCH' or [site]='SMCW' or [site]='UMCB') and ([gs_patientarrival]='1' or [gs_patientarrival]='3' or [gs_patientarrival]='10') and [ems_callrecdt_precision]<>'0' and [hide_nonessential_fields(1)]='0' | EMS Unit Notified by Dispatch<br>*If Date Only, enter 00:00 for time.* | text (datetime_mdy, Min: 2010-01-01 00:00, Max: 2099-12-31 23:59), Identifier |
| 1466 | ems_dispatch<br><br>Show the field ONLY if:<br>([site]='SMCA' or [site]='SMCH' or [site]='SMCW' or [site]='UMCB') and ([gs_patientarrival]='1' or [gs_patientarrival]='3' or [gs_patientarrival]='10') and [hide_nonessential_fields(1)]='0' | Dispatched as suspected stroke? | radio<br>1 \| Yes<br>2 \| No<br>3 \| Not Documented<br><br>Custom alignment: RH |
| 1467 | ems_arrivescenedt_precision<br><br>Show the field ONLY if:<br>([site]='SMCA' or [site]='SMCH' or [site]='SMCW' or [site]='UMCB') and ([gs_patientarrival]='1' or [gs_patientarrival]='3' or [gs_patientarrival]='10') | EMS Unit Arrived on Scene Precision | dropdown<br>5 \| Date & Time (MM/DD/YYYY HH24:MM)<br>3 \| Date Only (MM/DD/YYYY)<br>0 \| Unknown |
| 1468 | ems_arrivescenedt<br><br>Show the field ONLY if:<br>([site]='SMCA' or [site]='SMCH' or [site]='SMCW' or [site]='UMCB') and ([gs_patientarrival]='1' or [gs_patientarrival]='3' or [gs_patientarrival]='10') and [ems_arrivescenedt_precision]<>'0' | EMS Unit Arrived on Scene<br>*If Date Only, enter 00:00 for time.* | text (datetime_mdy, Min: 2010-01-01 00:00, Max: 2099-12-31 23:59), Identifier |

| | | | |
|---|---|---|---|
| 1469 | ems_firstmedcont_precision<br><br>Show the field ONLY if:<br>([site]='SMCA' or [site]='SMCH' or [site]='SMCW' or [site]='UMCB') and ([gs_patientarrival]='1' or [gs_patientarrival]='3' or [gs_patientarrival]='10') | EMS Arrived at Patient Precision | dropdown<br>5 \| Date & Time (MM/DD/YYYY HH24:MM)<br>0 \| Unknown |
| 1470 | ems_firstmedcont<br><br>Show the field ONLY if:<br>([site]='SMCA' or [site]='SMCH' or [site]='SMCW' or [site]='UMCB') and ([gs_patientarrival]='1' or [gs_patientarrival]='3' or [gs_patientarrival]='10') and [ems_firstmedcont_precision]<>'0' | EMS Arrived at Patient<br>*If Date Only, enter 00:00 for time.* | text (datetime_mdy, Min: 2010-01-01 00:00, Max: 2099-12-31 23:59), Identifier |
| 1471 | ems_scenedepdt_precision<br><br>Show the field ONLY if:<br>([site]='SMCA' or [site]='SMCH' or [site]='SMCW' or [site]='UMCB') and ([gs_patientarrival]='1' or [gs_patientarrival]='3' or [gs_patientarrival]='10') | EMS Unit Left Scene Precision | dropdown<br>5 \| Date & Time (MM/DD/YYYY HH24:MM)<br>3 \| Date Only (MM/DD/YYYY)<br>0 \| Unknown |
| 1472 | ems_scenedepdt<br><br>Show the field ONLY if:<br>([site]='SMCA' or [site]='SMCH' or [site]='SMCW' or [site]='UMCB') and ([gs_patientarrival]='1' or [gs_patientarrival]='3' or [gs_patientarrival]='10') and [ems_scenedepdt_precision]<>'0' | EMS Unit Left Scene<br>*If Date Only, enter 00:00 for time.* | text (datetime_mdy, Min: 2010-01-01 00:00, Max: 2099-12-31 23:59), Identifier |
| 1473 | ems_lkwdt_precision<br><br>Show the field ONLY if:<br>([site]='SMCA' or [site]='SMCH' or [site]='SMCW' or [site]='UMCB') and ([gs_patientarrival]='1' or [gs_patientarrival]='3' or [gs_patientarrival]='10') and [ems_lkwunk]<>'1' | Last Known Well as Documented by EMS Precision | dropdown<br>5 \| Date & Time (MM/DD/YYYY HH24:MM)<br>3 \| Date Only (MM/DD/YYYY)<br>0 \| Unknown |

| | | | |
|---|---|---|---|
| 1474 | ems_lkwdt<br><br>Show the field ONLY if:<br>([site]='SMCA' or [site]='SMCH' or [site]='SMCW' or [site]='UMCB') and ([gs_patientarrival]='1' or [gs_patientarrival]='3' or [gs_patientarrival]='10') and [ems_lkwdt_precision]<>'0' and [ems_lkwunk]<>'1' | Last Known Well as Documented by EMS<br>*If Date Only, enter 00:00 for time.* | text (datetime_mdy, Min: 2010-01-01 00:00, Max: 2099-12-31 23:59), Identifier |
| 1475 | ems_lkwunk<br><br>Show the field ONLY if:<br>([site]='SMCA' or [site]='SMCH' or [site]='SMCW' or [site]='UMCB') and ([gs_patientarrival]='1' or [gs_patientarrival]='3' or [gs_patientarrival]='10') | EMS LKW Unknowable | radio<br>1 \| Unknowable |
| 1476 | ems_symptomsdt_precision<br><br>Show the field ONLY if:<br>([site]='SMCA' or [site]='SMCH' or [site]='SMCW' or [site]='UMCB') and ([gs_patientarrival]='1' or [gs_patientarrival]='3' or [gs_patientarrival]='10') and [ems_symptomunk]<>'1' and [hide_nonessential_fields(1)]='0' | Discovery of Stroke symptoms by EMS Precision | dropdown<br>5 \| Date & Time (MM/DD/YYYY HH24:MM)<br>3 \| Date Only (MM/DD/YYYY)<br>0 \| Unknown |
| 1477 | ems_symptomsdt<br><br>Show the field ONLY if:<br>([site]='SMCA' or [site]='SMCH' or [site]='SMCW' or [site]='UMCB') and ([gs_patientarrival]='1' or [gs_patientarrival]='3' or [gs_patientarrival]='10') and [ems_symptomsdt_precision]<>'0' and [ems_symptomunk]<>'1' and [hide_nonessential_fields(1)]='0' | Discovery of Stroke symptoms by EMS<br>*If Date Only, enter 00:00 for time.* | text (datetime_mdy, Min: 2010-01-01 00:00, Max: 2099-12-31 23:59), Identifier |
| 1478 | ems_symptomunk<br><br>Show the field ONLY if:<br>([site]='SMCA' or [site]='SMCH' or [site]='SMCW' or [site]='UMCB') and ([gs_patientarrival]='1' or [gs_patientarrival]='3' or [gs_patientarrival]='10') and [hide_nonessential_fields(1)]='0' | EMS Discovery of Stroke symptoms unknowable | radio<br>1 \| Unknowable |

| | | | |
|---|---|---|---|
| 1479 | ems_prehosp_emsdt_precision<br><br>Show the field ONLY if:<br>[gs_prehosp_ems]='1' and [hide_nonessential_fields(1)]='0' | Date/Time Pre-Notification provided to Hospital Precision | dropdown<br>5 \| Date & Time (MM/DD/YYYY HH24:MM)<br>0 \| Unknown |
| 1480 | ems_prehosp_emsdt<br><br>Show the field ONLY if:<br>[gs_prehosp_ems]='1' and [ems_prehosp_emsdt_precision]<>'0' and [hide_nonessential_fields(1)]='0' | Date/Time Pre-Notification provided to Hospital<br>*If Date Only, enter 00:00 for time.* | text (datetime_mdy, Min: 2010-01-01 00:00, Max: 2099-12-31 23:59), Identifier |
| 1481 | ems_alert<br><br>Show the field ONLY if:<br>[gs_prehosp_ems]='1' and [hide_nonessential_fields(1)]='0' | Additional Information provided as part of pre-notification? | checkbox<br>1 \| ems_alert___1 \| Blood Glucose value<br>2 \| ems_alert___2 \| Blood Pressure<br>3 \| ems_alert___3 \| Result of Stroke Screen/Severity Score<br>4 \| ems_alert___4 \| LKW time per EMS<br>5 \| ems_alert___5 \| Seizure Activity |
| 1482 | ems_bgl_nd<br><br>Show the field ONLY if:<br>([site]='SMCA' or [site]='SMCH' or [site]='SMCW' or [site]='UMCB') and ([gs_patientarrival]='1' or [gs_patientarrival]='3' or [gs_patientarrival]='10') | Blood Glucose level: ND | dropdown<br>2 \| Glucometer Not Available<br>3 \| Patient Refused<br>1 \| Not Documented<br><br>Custom alignment: RH |
| 1483 | ems_bgl_val<br><br>Show the field ONLY if:<br>([site]='SMCA' or [site]='SMCH' or [site]='SMCW' or [site]='UMCB') and ([gs_patientarrival]='1' or [gs_patientarrival]='3' or [gs_patientarrival]='10') and [ems_bgl_nd]='' | Blood Glucose value | radio<br>1 \| Too High<br>2 \| Too Low<br><br>Custom alignment: RH |
| 1484 | ems_bgl<br><br>Show the field ONLY if:<br>([site]='SMCA' or [site]='SMCH' or [site]='SMCW' or [site]='UMCB') and ([gs_patientarrival]='1' or [gs_patientarrival]='3' or [gs_patientarrival]='10') and [ems_bgl_nd]='' and [ems_bgl_val]='' | Blood Glucose level (mg/dL) | text (integer) |

| | | | |
|---|---|---|---|
| 1485 | ems_bpnd<br><br>Show the field ONLY if:<br>([site]='SMCA' or [site]='SMCH' or [site]='SMCW' or [site]='UMCB') and ([gs_patientarrival]='1' or [gs_patientarrival]='3' or [gs_patientarrival]='10') and [hide_nonessential_fields(1)]='0' | BP by EMS - ND | radio<br>1 Not Documented |
| 1486 | ems_bps<br><br>Show the field ONLY if:<br>([site]='SMCA' or [site]='SMCH' or [site]='SMCW' or [site]='UMCB') and ([gs_patientarrival]='1' or [gs_patientarrival]='3' or [gs_patientarrival]='10') and [ems_bpnd]='' and [hide_nonessential_fields(1)]='0' | Initial Blood Pressure by EMS - Systolic<br>*mmHg (Systolic)* | text (integer, Min: 0, Max: 999) |
| 1487 | ems_bpd<br><br>Show the field ONLY if:<br>([site]='SMCA' or [site]='SMCH' or [site]='SMCW' or [site]='UMCB') and ([gs_patientarrival]='1' or [gs_patientarrival]='3' or [gs_patientarrival]='10') and [ems_bpnd]='' and [hide_nonessential_fields(1)]='0' | Initial Blood Pressure by EMS - Diastolic<br>*mmHg (Diastolic)* | text (integer, Min: 0, Max: 999) |
| 1488 | ems_suspstroke<br><br>Show the field ONLY if:<br>([site]='SMCA' or [site]='SMCH' or [site]='SMCW' or [site]='UMCB') and ([gs_patientarrival]='1' or [gs_patientarrival]='3' or [gs_patientarrival]='10') | Suspected stroke? | radio<br>1 Yes<br>2 No<br>3 Not documented<br><br>Custom alignment: RH |

| | | | |
|---|---|---|---|
| 1489 | ems_prehospscreen<br><br>Show the field ONLY if:<br>([site]='SMCA' or [site]='SMCH' or [site]='SMCW' or [site]='UMCB') and ([gs_patientarrival]='1' or [gs_patientarrival]='3' or [gs_patientarrival]='10') | Indicate the stroke screen tool used: | dropdown<br>14 BE FAST<br>4 CPSS<br>5 DPSS<br>6 FAST<br>7 LAPSS<br>8 MASS<br>9 Med PACS<br>15 MEND<br>10 mLAPSS<br>11 OPSST<br>12 ROSIER<br>13 Other<br>1 Stroke screen tool used, but tool used is unknown<br>2 No stroke screen tool used<br>3 Not documented |
| 1490 | ems_prehospscreenot<br><br>Show the field ONLY if:<br>[ems_prehospscreen]='13' | Stroke screen tool - If other, specify: | text |
| 1491 | ems_outcome<br><br>Show the field ONLY if:<br>([site]='SMCA' or [site]='SMCH' or [site]='SMCW' or [site]='UMCB') and ([gs_patientarrival]='1' or [gs_patientarrival]='3' or [gs_patientarrival]='10') and [ems_prehospscreen]<>'' and [ems_prehospscreen]<>'2' and [ems_prehospscreen]<>'3' | Stroke Screen Outcome | dropdown<br>1 Positive<br>2 Negative<br>3 Not Documented |

| | | | |
|---|---|---|---|
| 1492 | ems_sevscale<br><br>Show the field ONLY if:<br>[gs_patientarrival]='1' or [gs_patientarrival]='10' | Indicate the severity scale used | dropdown<br>1 CPSSS/CSTAT<br>2 FAST ED<br>3 LAMS<br>10 MPSS<br>5 RACE<br>11 VAN<br>6 Other<br>7 Severity scale used, but tool used is unknown<br>8 No severity scale used<br>9 Not Documented |
| 1493 | ems_sevscaleot<br><br>Show the field ONLY if:<br>[ems_sevscale]='6' | EMS Severity Scale - If other, specify: | text |
| 1494 | ems_poslvo<br><br>Show the field ONLY if:<br>([site]='SMCA' or [site]='SMCH' or [site]='SMCW' or [site]='UMCB') and ([gs_patientarrival]='1' or [gs_patientarrival]='10') and [ems_sevscale]<>'9' and [hide_nonessential_fields(1)]='0' | Positive for LVO? | dropdown<br>1 Yes<br>2 No<br>3 Not Documented |
| 1495 | ems_sevscore<br><br>Show the field ONLY if:<br>([site]='SMCA' or [site]='SMCH' or [site]='SMCW' or [site]='UMCB') and ([gs_patientarrival]='1' or [gs_patientarrival]='10') and [ems_sevscale]<>'' and [ems_sevscale]<>'8' and [ems_sevscale]<>'9' and [ems_sevscale]<>'11' and [ems_sevscorend]<>'1' | If severity scale assessment completed, enter total score: | text (integer, Min: 0, Max: 99) |

| | | | |
|---|---|---|---|
| 1496 | ems_sevscorend<br><br>Show the field ONLY if:<br>([site]='SMCA' or [site]='SMCH' or [site]='SMCW' or [site]='UMCB') and ([gs_patientarrival]='1' or [gs_patientarrival]='3' or [gs_patientarrival]='10') and [ems_sevscale]<>'' and [ems_sevscale]<>'9' and [hide_nonessential_fields(1)]='0' | Severity Score ND | radio<br>1 Not Documented |
| 1497 | ems_destdec<br><br>Show the field ONLY if:<br>([site]='SMCA' or [site]='SMCH' or [site]='SMCW' or [site]='UMCB') and ([gs_patientarrival]='1' or [gs_patientarrival]='3' or [gs_patientarrival]='10') and [hide_nonessential_fields(1)]='0' | How was destination decision made? | radio<br>1 Directed to designated stroke center by protocol<br>2 Directed to nearest facility by protocol<br>3 Patient/Family choice<br>4 Online Medical Direction<br>5 Closest facility<br>6 Other<br>7 Unknown/Not Documented |
| 1498 | ems_destdec_oth<br><br>Show the field ONLY if:<br>[ems_destdec]='6' and [hide_nonessential_fields(1)]='0' | If Other, Specify | text<br>Field Annotation: @CHARLIMIT=200 |
| 1499 | ems_thromcl<br><br>Show the field ONLY if:<br>([site]='SMCA' or [site]='SMCH' or [site]='SMCW' or [site]='UMCB') and ([gs_patientarrival]='1' or [gs_patientarrival]='3' or [gs_patientarrival]='10') and [hide_nonessential_fields(1)]='0' | Was a Thrombolytic Checklist used? | radio<br>1 Yes<br>2 No/ND<br><br>Custom alignment: RH |
| 1500 | ems_sevdest<br><br>Show the field ONLY if:<br>([site]='SMCA' or [site]='SMCH' or [site]='SMCW' or [site]='UMCB') and ([gs_patientarrival]='1' or [gs_patientarrival]='3' or [gs_patientarrival]='10') and [ems_sevscale]<>'' and [ems_sevscale]<>'9' and [hide_nonessential_fields(1)]='0' | If severity scale used, did result alter hospital destination (e.g. CSC vs. PSC)? | dropdown<br>1 Yes<br>2 No<br>3 Not Documented |

| | | | |
|---|---|---|---|
| 1501 | gs_prehdata<br><br>Show the field ONLY if:<br>([site]='SMCA' or [site]='SMCH' or [site]='SMCW' or [site]='UMCB') and ([gs_patientarrival]='1' or [gs_patientarrival]='3' or [gs_patientarrival]='10') | Source used to obtain prehospital care data: | dropdown<br>1 Hospital records on EMS<br>2 EMS records<br>3 Other |
| 1502 | special_initiatives_complete | Section Header: *Form Status*<br><br>Complete? | dropdown<br>0 Incomplete<br>1 Unverified<br>2 Complete |

Instrument: **MaRISS** (mariss) Collapse

| | | | |
|---|---|---|---|
| 1503 | m_patient | MaRISS Patient | radio<br>1 Yes<br>2 No<br><br>Custom alignment: RH |
| 1504 | m_consent<br><br>Show the field ONLY if:<br>[m_patient] = '1' | Patient provided MaRISS informed consent? | radio<br>1 Yes<br>2 No<br><br>Custom alignment: RH |
| 1505 | m_number<br><br>Show the field ONLY if:<br>[m_patient] = '1' | MaRISS Number | text<br>Custom alignment: RH |
| 1506 | m_riss_desc<br><br>Show the field ONLY if:<br>[m_patient] = '1' | Section Header: *Baseline Data*<br><br>Defining Rapidly Improving Stroke | descriptive |

| | | | |
|---|---|---|---|
| 1507 | m_riss<br><br>Show the field ONLY if:<br>[m_patient] = '1' | Main reason/criteria to code Rapidly Improving Stroke Symptoms<br>*Check all that apply* | checkbox<br>1 \| m_riss___1 \| Neurological function or NIHSS improvement in absolute points from baseline<br>2 \| m_riss___2 \| Proportion or percent improvement in neurological function or NIHSS from baseline<br>3 \| m_riss___3 \| Final NIHSS value<br>4 \| m_riss___4 \| Improvement to non-disabling condition<br>5 \| m_riss___5 \| Other<br><br>Custom alignment: LV |
| 1508 | m_rissspec<br><br>Show the field ONLY if:<br>[m_riss(5)] = '1' | If Other, Specify:<br>*60 character limit* | text<br>Field Annotation: @CHARLIMIT=60 |
| 1509 | m_evidence_desc<br><br>Show the field ONLY if:<br>[m_patient] = '1' | Pre-Hospital Arrival Neurologic Fluctuations | descriptive |
| 1510 | m_evidence<br><br>Show the field ONLY if:<br>[m_patient] = '1' | Is there evidence by direct interview or documentation of neurologic status change between onset of symptoms and hospital arrival? | radio<br>1 \| Yes<br>2 \| No<br>3 \| Not Documented/Unable to determine<br><br>Custom alignment: LH |
| 1511 | m_fluc_desc<br><br>Show the field ONLY if:<br>[m_evidence] = '1' | If YES, please note if they improved (I) (↑) or worsened (W) (↓). Note if more than one fluctuation. | descriptive |
| 1512 | m_fluc1<br><br>Show the field ONLY if:<br>[m_evidence] = '1' | Fluctuation 1 | radio<br>1 \| (I)<br>2 \| (W)<br><br>Custom alignment: RH |
| 1513 | m_fluc2<br><br>Show the field ONLY if:<br>[m_fluc1] <> '' | Fluctuation 2 | radio<br>1 \| (I)<br>2 \| (W)<br><br>Custom alignment: RH |

| | | | |
|---|---|---|---|
| 1514 | m_fluc3<br><br>Show the field ONLY if:<br>[m_fluc2] <> '' | Fluctuation 3 | radio<br>1 (I)<br>2 (W)<br><br>Custom alignment: RH |
| 1515 | m_fluc4<br><br>Show the field ONLY if:<br>[m_fluc3] <> '' | Fluctuation 4 | radio<br>1 (I)<br>2 (W)<br><br>Custom alignment: RH |
| 1516 | m_flucoverall<br><br>Show the field ONLY if:<br>[m_evidence] = '1' | Overall, did patient improve or worsen? | radio<br>1 Improve<br>2 Worsen<br><br>Custom alignment: RH |
| 1517 | m_nihss_bl<br><br>Show the field ONLY if:<br>[m_patient] = '1' | Baseline NIH Stroke Scale: | radio<br>1 Yes<br>2 No<br><br>Custom alignment: RH |
| 1518 | m_nihssdt_bl<br><br>Show the field ONLY if:<br>[m_nihss_bl] = '1' | Date/Time of baseline NIH Stroke Scale: | text (datetime_mdy, Min: 2010-01-01 00:00, Max: 2099-12-31 23:59), Identifier |
| 1519 | m_nihss1a_bl<br><br>Show the field ONLY if:<br>[m_nihss_bl] = '1' | 1.a. Level of consciousness (Baseline) | dropdown<br>2 0-Alert<br>3 1-Not alert, but arousable with minimal stimulation<br>4 2-Not alert, requires repeated stimulation to attend<br>5 3-Coma |
| 1520 | m_nihss1b_bl<br><br>Show the field ONLY if:<br>[m_nihss_bl] = '1' | 1.b. Ask patient the month and their age (Baseline) | dropdown<br>2 0-Answers both correctly<br>3 1-Answers one correctly<br>4 2-Both incorrect |

| | | | |
|---|---|---|---|
| 1521 | m_nihss1c_bl<br><br>Show the field ONLY if:<br>[m_nihss_bl] = '1' | 1.c. Ask patient to open and close eyes; make fist and let go (Baseline) | dropdown<br>2 0-Obeys both correctly<br>3 1-Obeys one correctly<br>4 2-Both incorrect |
| 1522 | m_nihss2_bl<br><br>Show the field ONLY if:<br>[m_nihss_bl] = '1' | 2. Best gaze (Baseline) | dropdown<br>2 0-Normal<br>3 1-Partial gaze palsy<br>4 2-Forced deviation |
| 1523 | m_nihss3_bl<br><br>Show the field ONLY if:<br>[m_nihss_bl] = '1' | 3. Visual field testing (Baseline) | dropdown<br>2 0-No visual field loss<br>3 1-Partial hemianopia<br>4 2-Complete hemianopia<br>5 3-Bilateral hemianopia (blind including cortical blindness) |
| 1524 | m_nihss4_bl<br><br>Show the field ONLY if:<br>[m_nihss_bl] = '1' | 4. Facial paresis (Baseline) | dropdown<br>2 0-Normal symmetrical movement<br>3 1-Minor paralysis (flattened nasolabial fold, asymmetry on smiling)<br>4 2-Partial paralysis (total or near paralysis of lower face)<br>5 3-Complete paralysis of one or both sides |
| 1525 | m_nihss5l_bl<br><br>Show the field ONLY if:<br>[m_nihss_bl] = '1' | 5l. Motor function - left arm (Baseline) | dropdown<br>2 0-Normal (extends arms 90 (or 45) degrees for 10 seconds without drift)<br>3 1-Drift<br>4 2-Some effort against gravity<br>5 3-No effort against gravity<br>6 4-No movement<br>7 U-Untestable (Joint fused or limb amputated) |

| | | | |
|---|---|---|---|
| 1526 | m_nihss5r_bl<br><br>Show the field ONLY if:<br>[m_nihss_bl] = '1' | 5r. Motor function - right arm (Baseline) | dropdown<br>2 0-Normal (extends arms 90 (or 45) degrees for 10 seconds without drift)<br>3 1-Drift<br>4 2-Some effort against gravity<br>5 3-No effort against gravity<br>6 4-No movement<br>7 U-Untestable (Joint fused or limb amputated) |
| 1527 | m_nihss6l_bl<br><br>Show the field ONLY if:<br>[m_nihss_bl] = '1' | 6l. Motor function - left leg (Baseline) | dropdown<br>2 0-Normal (hold leg 30 degrees position for 5 seconds)<br>3 1-Drift<br>4 2-Some effort against gravity<br>5 3-No effort against gravity<br>6 4-No movement<br>7 U-Untestable (Joint fused or limb amputated) |
| 1528 | m_nihss6r_bl<br><br>Show the field ONLY if:<br>[m_nihss_bl] = '1' | 6r. Motor function - right leg (Baseline) | dropdown<br>2 0-Normal (hold leg 30 degrees position for 5 seconds)<br>3 1-Drift<br>4 2-Some effort against gravity<br>5 3-No effort against gravity<br>6 4-No movement<br>7 U-Untestable (Joint fused or limb amputated) |
| 1529 | m_nihss7_bl<br><br>Show the field ONLY if:<br>[m_nihss_bl] = '1' | 7. Limb ataxia (Baseline) | dropdown<br>2 0-No ataxia<br>3 1-Present in one limb<br>4 2-Present in two limbs<br>5 U-Untestable (Joint fused or limb amputated) |
| 1530 | m_nihss8_bl<br><br>Show the field ONLY if:<br>[m_nihss_bl] = '1' | 8. Sensory (Baseline) | dropdown<br>2 0-Normal<br>3 1-Mild to moderate decrease in sensation<br>4 2-Severe to total sensory loss |

| | | | |
|---|---|---|---|
| 1531 | m_nihss9_bl<br><br>Show the field ONLY if:<br>[m_nihss_bl] = '1' | 9. Best language (Baseline) | dropdown<br>2 — 0-No aphasia<br>3 — 1-Mild to moderate aphasia<br>4 — 2-Severe aphasia<br>5 — 3-Mute |
| 1532 | m_nihss10_bl<br><br>Show the field ONLY if:<br>[m_nihss_bl] = '1' | 10. Dysarthria (Baseline) | dropdown<br>2 — 0-Normal articulation<br>3 — 1-Mild to moderate slurring of words<br>4 — 2-Near unintelligible or unable to speak<br>5 — U-Intubated or other physical barrier |
| 1533 | m_nihss11_bl<br><br>Show the field ONLY if:<br>[m_nihss_bl] = '1' | 11. Extinction and inattention (Baseline) | dropdown<br>2 — 0-Normal<br>3 — 1-Inattention or extinction to bilateral stimulation in one modality<br>4 — 2-Severe hemi-inattention or hemi-inattention to multiple modalities |
| 1534 | m_nihsstotal_bl | NIHSS Total Score (Baseline) | calc<br>Calculation: if([m_nihss1a_bl]='',0,[m_nihss1a_bl]-2) + if([m_nihss1b_bl]='',0,[m_nihss1b_bl]-2) + if([m_nihss1c_bl]='',0,[m_nihss1c_bl]-2) + if([m_nihss2_bl]='',0,[m_nihss2_bl]-2) + if([m_nihss3_bl]='',0,[m_nihss3_bl]-2) + if([m_nihss4_bl]='',0,[m_nihss4_bl]-2) + if([m_nihss5l_bl]='',0,if([m_nihss5l_bl]=7,0,[m_nihss5l_bl]-2)) + if([m_nihss5r_bl]='',0,if([m_nihss5r_bl]=7,0,[m_nihss5r_bl]-2)) + if([m_nihss6l_bl]='',0,if([m_nihss6l_bl]=7,0,[m_nihss6l_bl]-2)) + if([m_nihss6r_bl]='',0,if([m_nihss6r_bl]=7,0,[m_nihss6r_bl]-2)) + if([m_nihss7_bl]='',0,if([m_nihss7_bl]=5,0,[m_nihss7_bl]-2)) + if([m_nihss8_bl]='',0,[m_nihss8_bl]-2) + if([m_nihss9_bl]='',0,[m_nihss9_bl]-2) + if([m_nihss10_bl]='',0,if([m_nihss10_bl]=5,0,[m_nihss10_bl]-2)) + if([m_nihss11_bl]='',0,[m_nihss11_bl]-2) |
| 1535 | m_nihss_pt<br><br>Show the field ONLY if:<br>[m_patient] = '1' | Section Header: *Treatment Data*<br><br>NIH Stroke Scale immediately prior to treatment: | radio<br>1 — Yes<br>2 — No<br><br>Custom alignment: RH |
| 1536 | m_nihssdt_pt<br><br>Show the field ONLY if:<br>[m_nihss_pt] = '1' | Date/Time of NIH Stroke Scale immediately prior to treatment: | text (datetime_mdy, Min: 2010-01-01 00:00, Max: 2099-12-31 23:59), Identifier |

| | | | |
|---|---|---|---|
| 1537 | m_nochgbas<br><br>Show the field ONLY if:<br>[m_nihss_pt] = '1' | No Change from Baseline | radio<br>1 No Change |
| 1538 | m_nihss1a_pt<br><br>Show the field ONLY if:<br>[m_nihss_pt] = '1' | 1.a. Level of consciousness (Prior to Treatment) | dropdown<br>2 0-Alert<br>3 1-Not alert, but arousable with minimal stimulation<br>4 2-Not alert, requires repeated stimulation to attend<br>5 3-Coma |
| 1539 | m_nihss1b_pt<br><br>Show the field ONLY if:<br>[m_nihss_pt] = '1' | 1.b. Ask patient the month and their age (Prior to Treatment) | dropdown<br>2 0-Answers both correctly<br>3 1-Answers one correctly<br>4 2-Both incorrect |
| 1540 | m_nihss1c_pt<br><br>Show the field ONLY if:<br>[m_nihss_pt] = '1' | 1.c. Ask patient to open and close eyes; make fist and let go (Prior to Treatment) | dropdown<br>2 0-Obeys both correctly<br>3 1-Obeys one correctly<br>4 2-Both incorrect |
| 1541 | m_nihss2_pt<br><br>Show the field ONLY if:<br>[m_nihss_pt] = '1' | 2. Best gaze (Prior to Treatment) | dropdown<br>2 0-Normal<br>3 1-Partial gaze palsy<br>4 2-Forced deviation |
| 1542 | m_nihss3_pt<br><br>Show the field ONLY if:<br>[m_nihss_pt] = '1' | 3. Visual field testing (Prior to Treatment) | dropdown<br>2 0-No visual field loss<br>3 1-Partial hemianopia<br>4 2-Complete hemianopia<br>5 3-Bilateral hemianopia (blind including cortical blindness) |
| 1543 | m_nihss4_pt<br><br>Show the field ONLY if:<br>[m_nihss_pt] = '1' | 4. Facial paresis (Prior to Treatment) | dropdown<br>2 0-Normal symmetrical movement<br>3 1-Minor paralysis (flattened nasolabial fold, asymmetry on smiling)<br>4 2-Partial paralysis (total or near paralysis of lower face)<br>5 3-Complete paralysis of one or both sides |

<table>
<tr><td>1544</td><td>m_nihss5l_pt<br><br>Show the field ONLY if:<br>[m_nihss_pt] = '1'</td><td>5l. Motor function - left arm (Prior to Treatment)</td><td>dropdown<table><tr><td>2</td><td>0-Normal (extends arms 90 (or 45) degrees for 10 seconds without drift)</td></tr><tr><td>3</td><td>1-Drift</td></tr><tr><td>4</td><td>2-Some effort against gravity</td></tr><tr><td>5</td><td>3-No effort against gravity</td></tr><tr><td>6</td><td>4-No movement</td></tr><tr><td>7</td><td>U-Untestable (Joint fused or limb amputated)</td></tr></table></td></tr>
<tr><td>1545</td><td>m_nihss5r_pt<br><br>Show the field ONLY if:<br>[m_nihss_pt] = '1'</td><td>5r. Motor function - right arm (Prior to Treatment)</td><td>dropdown<table><tr><td>2</td><td>0-Normal (extends arms 90 (or 45) degrees for 10 seconds without drift)</td></tr><tr><td>3</td><td>1-Drift</td></tr><tr><td>4</td><td>2-Some effort against gravity</td></tr><tr><td>5</td><td>3-No effort against gravity</td></tr><tr><td>6</td><td>4-No movement</td></tr><tr><td>7</td><td>U-Untestable (Joint fused or limb amputated)</td></tr></table></td></tr>
<tr><td>1546</td><td>m_nihss6l_pt<br><br>Show the field ONLY if:<br>[m_nihss_pt] = '1'</td><td>6l. Motor function - left leg (Prior to Treatment)</td><td>dropdown<table><tr><td>2</td><td>0-Normal (hold leg 30 degrees position for 5 seconds)</td></tr><tr><td>3</td><td>1-Drift</td></tr><tr><td>4</td><td>2-Some effort against gravity</td></tr><tr><td>5</td><td>3-No effort against gravity</td></tr><tr><td>6</td><td>4-No movement</td></tr><tr><td>7</td><td>U-Untestable (Joint fused or limb amputated)</td></tr></table></td></tr>
<tr><td>1547</td><td>m_nihss6r_pt<br><br>Show the field ONLY if:<br>[m_nihss_pt] = '1'</td><td>6r. Motor function - right leg (Prior to Treatment)</td><td>dropdown<table><tr><td>2</td><td>0-Normal (hold leg 30 degrees position for 5 seconds)</td></tr><tr><td>3</td><td>1-Drift</td></tr><tr><td>4</td><td>2-Some effort against gravity</td></tr><tr><td>5</td><td>3-No effort against gravity</td></tr><tr><td>6</td><td>4-No movement</td></tr><tr><td>7</td><td>U-Untestable (Joint fused or limb amputated)</td></tr></table></td></tr>
</table>

| | | | |
|---|---|---|---|
| 1548 | m_nihss7_pt<br><br>Show the field ONLY if:<br>[m_nihss_pt] = '1' | 7. Limb ataxia (Prior to Treatment) | dropdown<br>2 0-No ataxia<br>3 1-Present in one limb<br>4 2-Present in two limbs<br>5 U-Untestable (Joint fused or limb amputated) |
| 1549 | m_nihss8_pt<br><br>Show the field ONLY if:<br>[m_nihss_pt] = '1' | 8. Sensory (Prior to Treatment) | dropdown<br>2 0-Normal<br>3 1-Mild to moderate decrease in sensation<br>4 2-Severe to total sensory loss |
| 1550 | m_nihss9_pt<br><br>Show the field ONLY if:<br>[m_nihss_pt] = '1' | 9. Best language (Prior to Treatment) | dropdown<br>2 0-No aphasia<br>3 1-Mild to moderate aphasia<br>4 2-Severe aphasia<br>5 3-Mute |
| 1551 | m_nihss10_pt<br><br>Show the field ONLY if:<br>[m_nihss_pt] = '1' | 10. Dysarthria (Prior to Treatment) | dropdown<br>2 0-Normal articulation<br>3 1-Mild to moderate slurring of words<br>4 2-Near unintelligible or unable to speak<br>5 U-Intubated or other physical barrier |
| 1552 | m_nihss11_pt<br><br>Show the field ONLY if:<br>[m_nihss_pt] = '1' | 11. Extinction and inattention (Prior to Treatment) | dropdown<br>2 0-Normal<br>3 1-Inattention or extinction to bilateral stimulation in one modality<br>4 2-Severe hemi-inattention or hemi-inattention to multiple modalities |

| | | | |
|---|---|---|---|
| 1553 | m_nihsstotal_pt | NIHSS Total Score (Prior to Treatment) | calc<br>Calculation: if([m_nihss1a_pt]='',0,[m_nihss1a_pt]-2) + if([m_nihss1b_pt]='',0,[m_nihss1b_pt]-2) + if([m_nihss1c_pt]='',0,[m_nihss1c_pt]-2) + if([m_nihss2_pt]='',0,[m_nihss2_pt]-2) + if([m_nihss3_pt]='',0,[m_nihss3_pt]-2) + if([m_nihss4_pt]='',0,[m_nihss4_pt]-2) + if([m_nihss5l_pt]='',0,if([m_nihss5l_pt]=7,0,[m_nihss5l_pt]-2)) + if([m_nihss5r_pt]='',0,if([m_nihss5r_pt]=7,0,[m_nihss5r_pt]-2)) + if([m_nihss6l_pt]='',0,if([m_nihss6l_pt]=7,0,[m_nihss6l_pt]-2)) + if([m_nihss6r_pt]='',0,if([m_nihss6r_pt]=7,0,[m_nihss6r_pt]-2)) + if([m_nihss7_pt]='',0,if([m_nihss7_pt]=5,0,[m_nihss7_pt]-2)) + if([m_nihss8_pt]='',0,[m_nihss8_pt]-2) + if([m_nihss9_pt]='',0,[m_nihss9_pt]-2) + if([m_nihss10_pt]='',0,if([m_nihss10_pt]=5,0,[m_nihss10_pt]-2)) + if([m_nihss11_pt]='',0,[m_nihss11_pt]-2) |
| 1554 | m_nihss_post<br>Show the field ONLY if:<br>[m_patient] = '1' | NIH Stroke Scale post treatment (24 hr or neurological worsening): | radio<br>1 Yes<br>2 No<br>Custom alignment: RH |
| 1555 | m_nihssdt_post<br>Show the field ONLY if:<br>[m_nihss_post] = '1' | Date/Time of post treatment NIH Stroke Scale: | text (datetime_mdy, Min: 2010-01-01 00:00, Max: 2099-12-31 23:59), Identifier |
| 1556 | m_nihss1a_post<br>Show the field ONLY if:<br>[m_nihss_post] = '1' | 1.a. Level of consciousness (Post Treatment) | dropdown<br>2 0-Alert<br>3 1-Not alert, but arousable with minimal stimulation<br>4 2-Not alert, requires repeated stimulation to attend<br>5 3-Coma |
| 1557 | m_nihss1b_post<br>Show the field ONLY if:<br>[m_nihss_post] = '1' | 1.b. Ask patient the month and their age (Post Treatment) | dropdown<br>2 0-Answers both correctly<br>3 1-Answers one correctly<br>4 2-Both incorrect |
| 1558 | m_nihss1c_post<br>Show the field ONLY if:<br>[m_nihss_post] = '1' | 1.c. Ask patient to open and close eyes; make fist and let go (Post Treatment) | dropdown<br>2 0-Obeys both correctly<br>3 1-Obeys one correctly<br>4 2-Both incorrect |

| | | | |
|---|---|---|---|
| 1559 | m_nihss2_post<br><br>Show the field ONLY if:<br>[m_nihss_post] = '1' | 2. Best gaze (Post Treatment) | dropdown<br>2 0-Normal<br>3 1-Partial gaze palsy<br>4 2-Forced deviation |
| 1560 | m_nihss3_post<br><br>Show the field ONLY if:<br>[m_nihss_post] = '1' | 3. Visual field testing (Post Treatment) | dropdown<br>2 0-No visual field loss<br>3 1-Partial hemianopia<br>4 2-Complete hemianopia<br>5 3-Bilateral hemianopia (blind including cortical blindness) |
| 1561 | m_nihss4_post<br><br>Show the field ONLY if:<br>[m_nihss_post] = '1' | 4. Facial paresis (Post Treatment) | dropdown<br>2 0-Normal symmetrical movement<br>3 1-Minor paralysis (flattened nasolabial fold, asymmetry on smiling)<br>4 2-Partial paralysis (total or near paralysis of lower face)<br>5 3-Complete paralysis of one or both sides |
| 1562 | m_nihss5l_post<br><br>Show the field ONLY if:<br>[m_nihss_post] = '1' | 5l. Motor function - left arm (Post Treatment) | dropdown<br>2 0-Normal (extends arms 90 (or 45) degrees for 10 seconds without drift)<br>3 1-Drift<br>4 2-Some effort against gravity<br>5 3-No effort against gravity<br>6 4-No movement<br>7 U-Untestable (Joint fused or limb amputated) |
| 1563 | m_nihss5r_post<br><br>Show the field ONLY if:<br>[m_nihss_post] = '1' | 5r. Motor function - right arm (Post Treatment) | dropdown<br>2 0-Normal (extends arms 90 (or 45) degrees for 10 seconds without drift)<br>3 1-Drift<br>4 2-Some effort against gravity<br>5 3-No effort against gravity<br>6 4-No movement<br>7 U-Untestable (Joint fused or limb amputated) |

| | | | |
|---|---|---|---|
| 1564 | m_nihss6l_post<br><br>Show the field ONLY if:<br>[m_nihss_post] = '1' | 6l. Motor function - left leg (Post Treatment) | dropdown<br>2 \| 0-Normal (hold leg 30 degrees position for 5 seconds)<br>3 \| 1-Drift<br>4 \| 2-Some effort against gravity<br>5 \| 3-No effort against gravity<br>6 \| 4-No movement<br>7 \| U-Untestable (Joint fused or limb amputated) |
| 1565 | m_nihss6r_post<br><br>Show the field ONLY if:<br>[m_nihss_post] = '1' | 6r. Motor function - right leg (Post Treatment) | dropdown<br>2 \| 0-Normal (hold leg 30 degrees position for 5 seconds)<br>3 \| 1-Drift<br>4 \| 2-Some effort against gravity<br>5 \| 3-No effort against gravity<br>6 \| 4-No movement<br>7 \| U-Untestable (Joint fused or limb amputated) |
| 1566 | m_nihss7_post<br><br>Show the field ONLY if:<br>[m_nihss_post] = '1' | 7. Limb ataxia (Post Treatment) | dropdown<br>2 \| 0-No ataxia<br>3 \| 1-Present in one limb<br>4 \| 2-Present in two limbs<br>5 \| U-Untestable (Joint fused or limb amputated) |
| 1567 | m_nihss8_post<br><br>Show the field ONLY if:<br>[m_nihss_post] = '1' | 8. Sensory (Post Treatment) | dropdown<br>2 \| 0-Normal<br>3 \| 1-Mild to moderate decrease in sensation<br>4 \| 2-Severe to total sensory loss |
| 1568 | m_nihss9_post<br><br>Show the field ONLY if:<br>[m_nihss_post] = '1' | 9. Best language (Post Treatment) | dropdown<br>2 \| 0-No aphasia<br>3 \| 1-Mild to moderate aphasia<br>4 \| 2-Severe aphasia<br>5 \| 3-Mute |

| | | | |
|---|---|---|---|
| 1569 | m_nihss10_post<br><br>Show the field ONLY if:<br>[m_nihss_post] = '1' | 10. Dysarthria (Post Treatment) | dropdown<br>2 — 0-Normal articulation<br>3 — 1-Mild to moderate slurring of words<br>4 — 2-Near unintelligible or unable to speak<br>5 — U-Intubated or other physical barrier |
| 1570 | m_nihss11_post<br><br>Show the field ONLY if:<br>[m_nihss_post] = '1' | 11. Extinction and inattention (Post Treatment) | dropdown<br>2 — 0-Normal<br>3 — 1-Inattention or extinction to bilateral stimulation in one modality<br>4 — 2-Severe hemi-inattention or hemi-inattention to multiple modalities |
| 1571 | m_nihsstotal_post | NIHSS Total Score (Post Treatment) | calc<br>Calculation: if([m_nihss1a_post]='',0,[m_nihss1a_post]-2) + if([m_nihss1b_post]='',0,[m_nihss1b_post]-2) + if([m_nihss1c_post]='',0,[m_nihss1c_post]-2) + if([m_nihss2_post]='',0,[m_nihss2_post]-2) + if([m_nihss3_post]='',0,[m_nihss3_post]-2) + if([m_nihss4_post]='',0,[m_nihss4_post]-2) + if([m_nihss5l_post]='',0,if([m_nihss5l_post]=7,0,[m_nihss5l_post]-2)) + if([m_nihss5r_post]='',0,if([m_nihss5r_post]=7,0,[m_nihss5r_post]-2)) + if([m_nihss6l_post]='',0,if([m_nihss6l_post]=7,0,[m_nihss6l_post]-2)) + if([m_nihss6r_post]='',0,if([m_nihss6r_post]=7,0,[m_nihss6r_post]-2)) + if([m_nihss7_post]='',0,if([m_nihss7_post]=5,0,[m_nihss7_post]-2)) + if([m_nihss8_post]='',0,[m_nihss8_post]-2) + if([m_nihss9_post]='',0,[m_nihss9_post]-2) + if([m_nihss10_post]='',0,if([m_nihss10_post]=5,0,[m_nihss10_post]-2)) + if([m_nihss11_post]='',0,[m_nihss11_post]-2) |
| 1572 | m_nihss_dc<br><br>Show the field ONLY if:<br>[m_patient] = '1' | Section Header: *Day 3 or at Discharge (if discharged prior to day 3) Data*<br>NIH Stroke Scale Day 3 or at discharge (if discharged prior to day 3): | radio<br>1 — Yes<br>2 — No<br><br>Custom alignment: RH |

| | | | |
|---|---|---|---|
| 1573 | m_nihssdt_dc<br><br>Show the field ONLY if:<br>[m_nihss_dc] = '1' | Date/Time of Day 3 or discharge NIH Stroke Scale: | text (datetime_mdy, Min: 2010-01-01 00:00, Max: 2099-12-31 23:59), Identifier |
| 1574 | m_nihss1a_dc<br><br>Show the field ONLY if:<br>[m_nihss_dc] = '1' | 1.a. Level of consciousness (Discharge) | dropdown<br>2 0-Alert<br>3 1-Not alert, but arousable with minimal stimulation<br>4 2-Not alert, requires repeated stimulation to attend<br>5 3-Coma |
| 1575 | m_nihss1b_dc<br><br>Show the field ONLY if:<br>[m_nihss_dc] = '1' | 1.b. Ask patient the month and their age (Discharge) | dropdown<br>2 0-Answers both correctly<br>3 1-Answers one correctly<br>4 2-Both incorrect |
| 1576 | m_nihss1c_dc<br><br>Show the field ONLY if:<br>[m_nihss_dc] = '1' | 1.c. Ask patient to open and close eyes; make fist and let go (Discharge) | dropdown<br>2 0-Obeys both correctly<br>3 1-Obeys one correctly<br>4 2-Both incorrect |
| 1577 | m_nihss2_dc<br><br>Show the field ONLY if:<br>[m_nihss_dc] = '1' | 2. Best gaze (Discharge) | dropdown<br>2 0-Normal<br>3 1-Partial gaze palsy<br>4 2-Forced deviation |
| 1578 | m_nihss3_dc<br><br>Show the field ONLY if:<br>[m_nihss_dc] = '1' | 3. Visual field testing (Discharge) | dropdown<br>2 0-No visual field loss<br>3 1-Partial hemianopia<br>4 2-Complete hemianopia<br>5 3-Bilateral hemianopia (blind including cortical blindness) |
| 1579 | m_nihss4_dc<br><br>Show the field ONLY if:<br>[m_nihss_dc] = '1' | 4. Facial paresis (Discharge) | dropdown<br>2 0-Normal symmetrical movement<br>3 1-Minor paralysis (flattened nasolabial fold, asymmetry on smiling)<br>4 2-Partial paralysis (total or near paralysis of lower face)<br>5 3-Complete paralysis of one or both sides |

| | | | |
|---|---|---|---|
| 1580 | m_nihss5l_dc<br><br>Show the field ONLY if:<br>[m_nihss_dc] = '1' | 5l. Motor function - left arm (Discharge) | dropdown<br>2 — 0-Normal (extends arms 90 (or 45) degrees for 10 seconds without drift)<br>3 — 1-Drift<br>4 — 2-Some effort against gravity<br>5 — 3-No effort against gravity<br>6 — 4-No movement<br>7 — U-Untestable (Joint fused or limb amputated) |
| 1581 | m_nihss5r_dc<br><br>Show the field ONLY if:<br>[m_nihss_dc] = '1' | 5r. Motor function - right arm (Discharge) | dropdown<br>2 — 0-Normal (extends arms 90 (or 45) degrees for 10 seconds without drift)<br>3 — 1-Drift<br>4 — 2-Some effort against gravity<br>5 — 3-No effort against gravity<br>6 — 4-No movement<br>7 — U-Untestable (Joint fused or limb amputated) |
| 1582 | m_nihss6l_dc<br><br>Show the field ONLY if:<br>[m_nihss_dc] = '1' | 6l. Motor function - left leg (Discharge) | dropdown<br>2 — 0-Normal (hold leg 30 degrees position for 5 seconds)<br>3 — 1-Drift<br>4 — 2-Some effort against gravity<br>5 — 3-No effort against gravity<br>6 — 4-No movement<br>7 — U-Untestable (Joint fused or limb amputated) |
| 1583 | m_nihss6r_dc<br><br>Show the field ONLY if:<br>[m_nihss_dc] = '1' | 6r. Motor function - right leg (Discharge) | dropdown<br>2 — 0-Normal (hold leg 30 degrees position for 5 seconds)<br>3 — 1-Drift<br>4 — 2-Some effort against gravity<br>5 — 3-No effort against gravity<br>6 — 4-No movement<br>7 — U-Untestable (Joint fused or limb amputated) |

| | | | |
|---|---|---|---|
| 1584 | m_nihss7_dc<br><br>Show the field ONLY if:<br>[m_nihss_dc] = '1' | 7. Limb ataxia (Discharge) | dropdown<br>2 0-No ataxia<br>3 1-Present in one limb<br>4 2-Present in two limbs<br>5 U-Untestable (Joint fused or limb amputated) |
| 1585 | m_nihss8_dc<br><br>Show the field ONLY if:<br>[m_nihss_dc] = '1' | 8. Sensory (Discharge) | dropdown<br>2 0-Normal<br>3 1-Mild to moderate decrease in sensation<br>4 2-Severe to total sensory loss |
| 1586 | m_nihss9_dc<br><br>Show the field ONLY if:<br>[m_nihss_dc] = '1' | 9. Best language (Discharge) | dropdown<br>2 0-No aphasia<br>3 1-Mild to moderate aphasia<br>4 2-Severe aphasia<br>5 3-Mute |
| 1587 | m_nihss10_dc<br><br>Show the field ONLY if:<br>[m_nihss_dc] = '1' | 10. Dysarthria (Discharge) | dropdown<br>2 0-Normal articulation<br>3 1-Mild to moderate slurring of words<br>4 2-Near unintelligible or unable to speak<br>5 U-Intubated or other physical barrier |
| 1588 | m_nihss11_dc<br><br>Show the field ONLY if:<br>[m_nihss_dc] = '1' | 11. Extinction and inattention (Discharge) | dropdown<br>2 0-Normal<br>3 1-Inattention or extinction to bilateral stimulation in one modality<br>4 2-Severe hemi-inattention or hemi-inattention to multiple modalities |

| | | | |
|---|---|---|---|
| 1589 | m_nihsstotal_dc | NIHSS Total Score (Discharge) | calc<br>Calculation: if([m_nihss1a_dc]='',0,[m_nihss1a_dc]-2) + if([m_nihss1b_dc]='',0,[m_nihss1b_dc]-2) + if([m_nihss1c_dc]='',0,[m_nihss1c_dc]-2) + if([m_nihss2_dc]='',0,[m_nihss2_dc]-2) + if([m_nihss3_dc]='',0, [m_nihss3_dc]-2) + if([m_nihss4_dc]='',0,[m_nihss4_dc]-2) + if([m_nihss5l_dc]='',0,if([m_nihss5l_dc]=7,0,[m_nihss5l_dc]-2)) + if([m_nihss5r_dc]='',0,if([m_nihss5r_dc]=7,0, [m_nihss5r_dc]-2)) + if([m_nihss6l_dc]='',0,if([m_nihss6l_dc]=7,0,[m_nihss6l_dc]-2)) + if([m_nihss6r_dc]='',0,if([m_nihss6r_dc]=7,0, [m_nihss6r_dc]-2)) + if([m_nihss7_dc]='',0,if([m_nihss7_dc]=5,0,[m_nihss7_dc]-2)) + if([m_nihss8_dc]='',0,[m_nihss8_dc]-2) + if([m_nihss9_dc]='',0, [m_nihss9_dc]-2) + if([m_nihss10_dc]='',0,if([m_nihss10_dc]=5,0, [m_nihss10_dc]-2)) + if([m_nihss11_dc]='',0,[m_nihss11_dc]-2) |
| 1590 | m_toast_nd<br><br>Show the field ONLY if:<br>[m_patient] = '1' | TOAST Classification Not Documented | radio<br>1 ND |
| 1591 | m_toast_day3<br><br>Show the field ONLY if:<br>[m_patient] = '1' and [m_toast_nd] <> '1' | TOAST Classification at Day 3 or discharge (if discharged prior to day 3): | radio<br>1 Large-artery atherosclerosis<br>2 Cardioembolism (high-risk/medium-risk)<br>3 Small-vessel occlusion (lacune)<br>5 Stroke of undetermined etiology<br>4 Stroke of other determined etiology<br><br>Custom alignment: LV |
| 1592 | m_largeartery<br><br>Show the field ONLY if:<br>[m_toast_day3] = '1' | Large-artery Atherosclerosis Type<br>*Check all that apply* | checkbox<br>1 m_largeartery___1 Extracranial atherosclerotic disease (Carotid or Vertebral artery plaque)<br>2 m_largeartery___2 Intracranial atherosclerotic disease<br>3 m_largeartery___3 Aortic Arch atherosclerotic disease<br><br>Custom alignment: LV |

<table>
<tr><td>1593</td><td>m_cardioem<br><br>Show the field ONLY if:<br>[m_toast_day3] = '2'</td><td>Cardioembolism Type<br>Check all that apply</td><td>checkbox<br>
<table>
<tr><td>1</td><td>m_cardioem___1</td><td>Left atrial thrombus</td></tr>
<tr><td>3</td><td>m_cardioem___3</td><td>Left ventricular thrombus</td></tr>
<tr><td>5</td><td>m_cardioem___5</td><td>Atrial Fibrillation</td></tr>
<tr><td>7</td><td>m_cardioem___7</td><td>Paroxysmal Atrial Fibrillation</td></tr>
<tr><td>9</td><td>m_cardioem___9</td><td>Sick sinus Syndrome</td></tr>
<tr><td>11</td><td>m_cardioem___11</td><td>Sustained atrial flutter</td></tr>
<tr><td>13</td><td>m_cardioem___13</td><td>Recent Myocardial infarction (within 1 month)</td></tr>
<tr><td>15</td><td>m_cardioem___15</td><td>Rheumatoid mitral or aortic valve disease</td></tr>
<tr><td>17</td><td>m_cardioem___17</td><td>Bioprosthetic and mechanical heart valve</td></tr>
<tr><td>2</td><td>m_cardioem___2</td><td>Chronic myocardial infarction together with low ejection fraction < 28%</td></tr>
<tr><td>4</td><td>m_cardioem___4</td><td>Symptomatic congestive heart failure with ejection fraction < 30%</td></tr>
<tr><td>6</td><td>m_cardioem___6</td><td>Dilated cardiomyopathy</td></tr>
<tr><td>8</td><td>m_cardioem___8</td><td>Nonbacterial thrombotic endocarditis</td></tr>
<tr><td>10</td><td>m_cardioem___10</td><td>Infective endocarditis</td></tr>
<tr><td>12</td><td>m_cardioem___12</td><td>Papillary Fibroelastoma</td></tr>
<tr><td>14</td><td>m_cardioem___14</td><td>Left atrial myoma</td></tr>
<tr><td>16</td><td>m_cardioem___16</td><td>Other cardioembolic source</td></tr>
</table>
<br>Custom alignment: LV</td></tr>
<tr><td>1594</td><td>m_othercard<br><br>Show the field ONLY if:<br>[m_cardioem(16)] = '1'</td><td>Other cardioembolic source, specify:<br>60 character limit</td><td>text<br>Field Annotation: @CHARLIMIT=60</td></tr>
</table>

<table>
<tr><td>1595</td><td>m_undeterminedstroke<br><br>Show the field ONLY if:<br>[m_toast_day3] = '5'</td><td>Stroke of undetermined etiology<br>*Check all that apply*</td><td>checkbox<table><tr><td>1</td><td>m_undeterminedstroke___1</td><td>Multiple etiologies seem likely</td></tr><tr><td>2</td><td>m_undeterminedstroke___2</td><td>Don't know despite thorough evaluation</td></tr><tr><td>3</td><td>m_undeterminedstroke___3</td><td>Don't know, evaluation incomplete</td></tr></table><br>Custom alignment: LV</td></tr>
<tr><td>1596</td><td>m_multspec<br><br>Show the field ONLY if:<br>[m_undeterminedstroke(1)] = '1'</td><td>Multiple etiologies seem likely, specify:<br>*60 character limit*</td><td>text<br>Field Annotation: @CHARLIMIT=60</td></tr>
<tr><td>1597</td><td>m_otherstroke<br><br>Show the field ONLY if:<br>[m_toast_day3] = '4'</td><td>Stroke of other determined etiology<br>*Check all that apply*</td><td>checkbox<table><tr><td>1</td><td>m_otherstroke___1</td><td>Hypercoagulable states*</td></tr><tr><td>2</td><td>m_otherstroke___2</td><td>Iatrogenic</td></tr><tr><td>3</td><td>m_otherstroke___3</td><td>Arterial dissection (carotid or vertebral)</td></tr><tr><td>4</td><td>m_otherstroke___4</td><td>Other</td></tr></table><br>Custom alignment: LV</td></tr>
<tr><td>1598</td><td>m_otherstkspec<br><br>Show the field ONLY if:<br>[m_otherstroke(4)] = '1'</td><td>Specify stroke of other determined etiology<br>*60 character limit*</td><td>text<br>Field Annotation: @CHARLIMIT=60</td></tr>
<tr><td>1599</td><td>m_hyperco<br><br>Show the field ONLY if:<br>[m_otherstroke(1)] = '1'</td><td>*If hypercoagulable states:<br>*Check all that apply*</td><td>checkbox<table><tr><td>1</td><td>m_hyperco___1</td><td>Malignancy</td></tr><tr><td>2</td><td>m_hyperco___2</td><td>Antiphospholipid antibody syndrome</td></tr><tr><td>3</td><td>m_hyperco___3</td><td>Inherited thrombophilia (including factor V Leiden deficiency, prothrombin gene mutation, protein C deficiency, protein S deficiency, antithrombin deficiency)</td></tr><tr><td>4</td><td>m_hyperco___4</td><td>Other specified hypercoagulable state.</td></tr></table><br>Custom alignment: LV</td></tr>
<tr><td>1600</td><td>m_spechyp<br><br>Show the field ONLY if:<br>[m_hyperco(4)] = '1'</td><td>Other specified hypercoagulable state. Specify:<br>*60 character limit*</td><td>text<br>Field Annotation: @CHARLIMIT=60</td></tr>
</table>

| | | | |
|---|---|---|---|
| 1601 | m_neurochange_desc<br><br>Show the field ONLY if:<br>[m_patient] = '1' | Post-Hospital Arrival Neurologic Fluctuations | descriptive |
| 1602 | m_neurochange<br><br>Show the field ONLY if:<br>[m_patient] = '1' | Is there evidence by direct interview or documentation of neurologic status change between hospital arrival and day 3 or hospital discharge, whichever occurred first? | radio<br>1 Yes<br>2 No<br>3 Not Documented/Unable to determine<br><br>Custom alignment: LH |
| 1603 | m_fluc_day3_desc<br><br>Show the field ONLY if:<br>[m_neurochange] = '1' | If YES, please note if they improved (I) (↑) or worsened (W) (↓). Note if more than one fluctuation. | descriptive |
| 1604 | m_fluc1_day3<br><br>Show the field ONLY if:<br>[m_neurochange] = '1' | Fluctuation 1 | radio<br>1 (I)<br>2 (W)<br><br>Custom alignment: RH |
| 1605 | m_fluc2_day3<br><br>Show the field ONLY if:<br>[m_fluc1_day3] <> '' | Fluctuation 2 | radio<br>1 (I)<br>2 (W)<br><br>Custom alignment: RH |
| 1606 | m_fluc3_day3<br><br>Show the field ONLY if:<br>[m_fluc2_day3] <> '' | Fluctuation 3 | radio<br>1 (I)<br>2 (W)<br><br>Custom alignment: RH |
| 1607 | m_fluc4_day3<br><br>Show the field ONLY if:<br>[m_fluc3_day3] <> '' | Fluctuation 4 | radio<br>1 (I)<br>2 (W)<br><br>Custom alignment: RH |

| | | | |
|---|---|---|---|
| 1608 | m_flucoverall_day3<br><br>Show the field ONLY if:<br>[m_neurochange] = '1' | Overall, did patient improve or worsen? | radio<br>1 Improve<br>2 Worsen<br><br>Custom alignment: RH |
| 1609 | m_selfreport<br><br>Show the field ONLY if:<br>[m_patient] = '1' | Was this patient selected as a web based patient self-reported outcomes patient? | radio<br>1 Yes<br>2 No<br>3 Not Documented<br><br>Custom alignment: LH |
| 1610 | m_infcons<br><br>Show the field ONLY if:<br>[m_patient] = '1' | MaRISS informed consent provided? | radio<br>1 Yes<br>2 No<br><br>Custom alignment: RH |
| 1611 | mariss_complete | Section Header: *Form Status*<br><br>Complete? | dropdown<br>0 Incomplete<br>1 Unverified<br>2 Complete |
| Instrument: **Historic** (historic) | | | Collapse |
| 1612 | transfer_facility<br><br>Show the field ONLY if:<br>[gs_patientarrival] = '3' | ✪ Name of transferring facility | text<br>Field Annotation: @HIDDEN |
| 1613 | diagnostic_tests<br><br>Show the field ONLY if:<br>([site]='SMCA' or [site]='SMCH' or [site]='SMCW' or [site]='UMCB') and ([treatment_recvd]='1' or [gs_notadmit]<>'1') | Section Header: *Diagnostic Tests*<br><br>Diagnostic Tests used for Acute Stroke Patients (Diagnostic Only)<br>*Select all that apply.* | checkbox<br>CT \| diagnostic_tests___ct \| CT<br>CTA \| diagnostic_tests___cta \| CTA<br>MRI \| diagnostic_tests___mri \| MRI<br>MRA \| diagnostic_tests___mra \| MRA<br>NeuroAngio \| diagnostic_tests___neuroangio \| Neuro Angiography<br><br>Custom alignment: LH<br>Field Annotation: @HIDDEN |

| | | | |
|---|---|---|---|
| 1614 | dnv_23<br><br>Show the field ONLY if:<br>[diagnostic_tests(NeuroAngio)] = '1' | In-Hospital Stroke or Death within 24 hours of diagnostic Neuroangiography.<br>(Exclude patients if they have undergone a therapeutic angiographic intervention as part of the same procedure or within the first 24 hours after the diagnostic procedure unless the complication is identified before the therapeutic intervention.) | yesno<br>1 Yes<br>0 No<br><br>Custom alignment: LH<br>Field Annotation: @HIDDEN |
| 1615 | nimodipine_ontime_num<br><br>Show the field ONLY if:<br>([site] = 'SMCA' or [site] = 'UMCB') and [gs_notadmit]<>'1' | Section Header: *Nimodipine On Time*<br><br>Number of Nimodipine doses given on time per orders/Rx. | text (integer)<br>Field Annotation: @HIDDEN |
| 1616 | nimodipine_ontime_den<br><br>Show the field ONLY if:<br>([site] = 'SMCA' or [site] = 'UMCB') and [gs_notadmit]<>'1' | Number of Nimodipine doses required per orders/Rx. | text (integer)<br>Field Annotation: @HIDDEN |
| 1617 | calc_nimodipine_ontime<br><br>Show the field ONLY if:<br>([site] = 'SMCA' or [site] = 'UMCB') and [gs_notadmit]<>'1' | Nimodipine On-Time Compliance<br>*Percent* | calc<br>Calculation:<br>round(([nimodipine_ontime_num]/[nimodipine_ontime_den])*10[illegible]<br>0)<br>Field Annotation: @HIDDEN |
| 1618 | neuro_check_x48<br><br>Show the field ONLY if:<br>[site] = 'SMCH' and [gs_notadmit]<>'1' | Section Header: *SMCH Neuro Checks (All patients in first 48 hours)*<br><br>Were Neuro Checks completed as ordered in the first 48 Hours? | yesno<br>1 Yes<br>0 No<br><br>Custom alignment: RH<br>Field Annotation: @HIDDEN |
| 1619 | neuro_check_x48_num<br><br>Show the field ONLY if:<br>[site] = 'SMCH' and [gs_notadmit]<>'1' | Number of Neuro Checks completed as ordered in the first 48 Hours:<br>*Neuro Checks Performed (numerator)* | text (integer)<br>Custom alignment: RH<br>Field Annotation: @HIDDEN |
| 1620 | neuro_check_x48_den<br><br>Show the field ONLY if:<br>[site] = 'SMCH' and [gs_notadmit]<>'1' | Number of Neuro Checks ordered in the first 48 Hours:<br>*Neuro Checks Ordered (denominator)* | text (integer)<br>Custom alignment: RH<br>Field Annotation: @HIDDEN |
| 1621 | calc_neuro_check_x48<br><br>Show the field ONLY if:<br>[site] = 'SMCH' and [gs_notadmit]<>'1' | Neuro Checks Compliance (first 48 Hours):<br>*Percent* | calc<br>Calculation:<br>round(([neuro_check_x48_num]/[neuro_check_x48_den])*100,1)<br>Custom alignment: RH<br>Field Annotation: @HIDDEN |

| | | | |
|---|---|---|---|
| 1622 | cva_poc_na | Section Header: *CVA Plan of Care*<br>CVA POC Not Applicable<br>*Patient was comfort measures only, expired, or was discharged within 24 hours.* | checkbox<br>1 \| cva_poc_na___1 \| N/A<br><br>Field Annotation: @HIDDEN |
| 1623 | cva_poc_sample<br><br>Show the field ONLY if:<br>([site]='SMCA' or [site]='SMCH' or [site]='SMCW' or [site]='UMCB') and [gs_notadmit]='2' and [cva_poc_na(1)]=0 | Part of the CVA POC Sample?<br>*Program Manager will select sample patients.* | radio<br>1 \| Add this patient to the CVA POC sample.<br><br>Field Annotation: @HIDDEN |
| 1624 | cva_poc_init<br><br>Show the field ONLY if:<br>([site]='SMCA' or [site]='SMCH' or [site]='SMCW' or [site]='UMCB') and [gs_notadmit]='2' and [cva_poc_na(1)]=0 | Was the CVA Plan of Care Initiated within 24 hours of admission? | yesno<br>1 \| Yes<br>0 \| No<br><br>Custom alignment: RH<br>Field Annotation: @HIDDEN |
| 1625 | cva_poc_signed<br><br>Show the field ONLY if:<br>([site]='SMCA' or [site]='SMCH' or [site]='SMCW' or [site]='UMCB') and [gs_notadmit]='2' and [cva_poc_na(1)]=0 | Was the CVA Plan of Care signed and dated by all RNs? | yesno<br>1 \| Yes<br>0 \| No<br><br>Custom alignment: RH<br>Field Annotation: @HIDDEN |
| 1626 | cva_poc_num<br><br>Show the field ONLY if:<br>([site]='SMCA' or [site]='SMCH' or [site]='SMCW' or [site]='UMCB') and [gs_notadmit]='2' and [cva_poc_na(1)]=0 | How many days was the CVA Plan of Care reviewed and updated?<br>*Numerator* | text (integer)<br>Custom alignment: RH<br>Field Annotation: @HIDDEN |
| 1627 | cva_poc_den<br><br>Show the field ONLY if:<br>([site]='SMCA' or [site]='SMCH' or [site]='SMCW' or [site]='UMCB') and [gs_notadmit]='2' and [cva_poc_na(1)]=0 | Total number of days the CVA Plan of Care should have been reviewed and updated?<br>*Denominator* | text (integer)<br>Custom alignment: RH<br>Field Annotation: @HIDDEN |
| 1628 | calc_cva_poc<br><br>Show the field ONLY if:<br>([site]='SMCA' or [site]='SMCH' or [site]='SMCW' or [site]='UMCB') and [gs_notadmit]='2' and [cva_poc_na(1)]=0 | CVA Plan of Care Compliance | calc<br>Calculation: round(([cva_poc_num]/[cva_poc_den])*100,1)<br>Custom alignment: RH<br>Field Annotation: @HIDDEN |

| | | | |
|---|---|---|---|
| 1629 | cva_poc_risk_fact_init<br><br>Show the field ONLY if:<br>([site]='SMCA' or [site]='SMCH' or [site]='SMCW' or [site]='UMCB') an d [gs_notadmit]='2' and [cva_poc_ na(1)]=0 | Was Stroke Education on Risk Factors initiated on the CVA POC? | yesno<br>1 Yes<br>0 No<br><br>Custom alignment: RH<br>Field Annotation: @HIDDEN |
| 1630 | cva_poc_risk_fact_ident<br><br>Show the field ONLY if:<br>([site]='SMCA' or [site]='SMCH' or [site]='SMCW' or [site]='UMCB') an d [gs_notadmit]='2' and [cva_poc_ na(1)]=0 | Were all appropriate patient specific risk factors selected for stroke education on the CVA POC? | yesno<br>1 Yes<br>0 No<br><br>Custom alignment: RH<br>Field Annotation: @HIDDEN |
| 1631 | cva_poc_indiv_needs_ident<br><br>Show the field ONLY if:<br>[cva_poc_sample]='1' | Were individual needs identified for the patient based on their condition and co-morbidities on any POC? | yesno<br>1 Yes<br>0 No<br><br>Custom alignment: RH<br>Field Annotation: @HIDDEN |
| 1632 | cva_poc_notes<br><br>Show the field ONLY if:<br>([site]='SMCA' or [site]='SMCH' or [site]='SMCW' or [site]='UMCB') an d [gs_notadmit]='2' and [cva_poc_ na(1)]=0 | Identify specific areas of CVA POC non-compliance | notes<br>Field Annotation: @HIDDEN |
| 1633 | telemetry_sample<br><br>Show the field ONLY if:<br>[telemetry_ordered] = '1' | Section Header: *Telemetry Compliance*<br><br>Part of the Telemetry Sample?<br>*Program Manager will select sample patients.* | radio<br>1 Add this patient to the telemetry sample<br><br>Field Annotation: @HIDDEN |
| 1634 | telemetry_ordered<br><br>Show the field ONLY if:<br>([site] = 'SMCA' or [site] = 'SMCH' o r [site] = 'SMCW' or [site] = 'UMC B') and [gs_notadmit] = '2' | Did this patient have telemetry orders for this hospital stay? | yesno<br>1 Yes<br>0 No<br><br>Custom alignment: RH<br>Field Annotation: @HIDDEN |
| 1635 | telemetry_in_chart<br><br>Show the field ONLY if:<br>[telemetry_sample] = '1' | Telemetry strips in chart per shift per policy (all tiers).<br>Tiers 1 & 2 also require interpretation, signature and date. | text (integer)<br>Field Annotation: @HIDDEN |

| | | | |
|---|---|---|---|
| 1636 | telemetry_required<br><br>Show the field ONLY if:<br>[telemetry_sample] = '1' | Number of Telemetry strips required to be in chart per policy. | text (integer)<br>Field Annotation: @HIDDEN |
| 1637 | calc_telemetry_compliance<br><br>Show the field ONLY if:<br>[telemetry_sample] = '1' | Telemetry Compliance<br>*Percent* | calc<br>Calculation:<br>round(([telemetry_in_chart]/[telemetry_required])*100,1)<br>Field Annotation: @HIDDEN |
| 1638 | telemetry_notes<br><br>Show the field ONLY if:<br>[telemetry_sample] = '1' | Identify specific areas of Telemetry non-compliance | notes<br>Field Annotation: @HIDDEN |
| 1639 | dnv_13_proc_start<br><br>Show the field ONLY if:<br>([site]='SMCA' or [site]='SMCH' or [site]='SMCW' or [site]='UMCB') and ([procedures_interventions(1)]='1' or [procedures_interventions(2)]='1') and [dnv_18_rupt]<>'2' and [jc_princicd10diagnosis]<>'I671' | Section Header: *DNV 13: Clipping or Coiling for Aneurysm*<br><br>Surgical Clipping or Endovascular Coiling procedure start time. | text (datetime_mdy, Min: 2010-01-01 00:00, Max: 2099-12-31 23:59), Identifier<br>Custom alignment: RH<br>Field Annotation: @HIDDEN |
| 1640 | dnv_13_excl_reason<br><br>Show the field ONLY if:<br>[aneurysmal_sah]='1' and (([procedures_interventions(1)]='1' or [procedures_interventions(2)]='1') or datediff([jc_arrdatetime], [dnv_13_proc_start], "h", "mdy", true)>36) | Was a reason documented for not treating the ruptured aneurysm with a clipping or coiling procedure?<br>(Or not treating within 36 hours of arrival?) | yesno<br>1 Yes<br>0 No<br><br>Custom alignment: RH<br>Field Annotation: @HIDDEN |
| 1641 | tpa_tool_md_num<br><br>Show the field ONLY if:<br>([site]='SMCA' or [site]='SMCH' or [site]='SMCW' or [site]='UMCB') and ([treatment_type(IAtPA)]='1' or [treatment_type(IVtPA)]='1' or [treatment_type(MER)]='1' or [gs_ivthroinit]='1' or [gs_ivtpaoutside]='1') | ✪ Clinician Notification Numerator<br>*Number Done Correctly.Enter 1 if there were no opportunities for this section.* | text (integer), Required<br>Custom alignment: RH<br>Field Annotation: @HIDDEN |
| 1642 | tpa_tool_md_den<br><br>Show the field ONLY if:<br>([site]='SMCA' or [site]='SMCH' or [site]='SMCW' or [site]='UMCB') and ([treatment_type(IAtPA)]='1' or [treatment_type(IVtPA)]='1' or [treatment_type(MER)]='1' or [gs_ivthroinit]='1' or [gs_ivtpaoutside]='1') | ✪ Clinician Notification Denominator<br>*Number that should have been done.Enter 1 if there were no opportunities for this section.* | text (integer), Required<br>Custom alignment: RH<br>Field Annotation: @HIDDEN |

| | | | |
|---|---|---|---|
| 1643 | tpa_tool_handoff_num<br><br>Show the field ONLY if:<br>([site]='SMCA' or [site]='SMCH' or [site]='SMCW' or [site]='UMCB') and ([treatment_type(IAtPA)]='1' or [treatment_type(IVtPA)]='1' or [treatment_type(MER)]='1' or [gs_ivthroinit]='1' or [gs_ivtpaoutside]='1') | ✪ Post Thrombolytic Hand Off Numerator<br>*Number Done Correctly.Enter 1 if there were no opportunities for this section.* | text (integer), Required<br>Custom alignment: RH<br>Field Annotation: @HIDDEN |
| 1644 | tpa_tool_handoff_den<br><br>Show the field ONLY if:<br>([site]='SMCA' or [site]='SMCH' or [site]='SMCW' or [site]='UMCB') and ([treatment_type(IAtPA)]='1' or [treatment_type(IVtPA)]='1' or [treatment_type(MER)]='1' or [gs_ivthroinit]='1' or [gs_ivtpaoutside]='1') | ✪ Post Thrombolytic Hand Off Denominator<br>*Number that should have been done.Enter 1 if there were no opportunities for this section.* | text (integer), Required<br>Custom alignment: RH<br>Field Annotation: @HIDDEN |
| 1645 | problem_list<br><br>Show the field ONLY if:<br>[site] = 'SMCW' and ([treatment_recvd]='1' or [gs_notadmit]<>'1') | Section Header: *Problem List & Individualized Risk Factors*<br><br>Problem List populated? | yesno<br>1 Yes<br>0 No<br><br>Custom alignment: RH<br>Field Annotation: @HIDDEN |
| 1646 | risk_factors<br><br>Show the field ONLY if:<br>[site] = 'SMCW' and ([treatment_recvd]='1' or [gs_notadmit]<>'1') | Is there documentation that the patient received education on individualized specific risk factors? | yesno<br>1 Yes<br>0 No<br><br>Custom alignment: RH<br>Field Annotation: @HIDDEN |
| 1647 | gs_insurance<br><br>Show the field ONLY if:<br>([treatment_recvd]='1' or [gs_notadmit]<>'1') | Health Insurance Status<br>*Select all that apply.* | checkbox<br>1 gs_insurance___1 Medicare<br>5 gs_insurance___5 Medicaid<br>2 gs_insurance___2 Private/VA/Champus/Other Insurance<br>3 gs_insurance___3 Self Pay/No Insurance<br>4 gs_insurance___4 ND<br><br>Field Annotation: @HIDDEN @NONEOFTHEABOVE='4' |

| | | | |
|---|---|---|---|
| 1648 | historic_complete | Section Header: *Form Status*<br>Complete? | dropdown<br>0 Incomplete<br>1 Unverified<br>2 Complete |
